# The Impact of SARS-CoV-2 Lineages (Variants) on the COVID-19 Epidemic in South Africa

**DOI:** 10.1101/2021.10.22.21265316

**Authors:** Thabo Mabuka, Natalie Naidoo, Nesisa Ncube, Thabo Yiga, Michael Ross, Kuzivakwashe Kurehwa, Mothabisi Nare, Andrea Silaji, Tinashe Ndemera, Tlaleng Lemeke, Taiwo Ridwan Ademola, Willie Macharia, Mthokozisi Sithole

## Abstract

Emerging SARS-CoV-2 variants have been attributed to the occurrence of secondary and tertiary COVID-19 epidemic waves and also threatening vaccine efforts due to their immune invasiveness. Since the importation of SARS-CoV-2 in South Africa, with the first reported COVID-19 case on the 5th of March 2020, South Africa has observed 3 consecutive COVID-19 epidemic waves. The evolution of SARS-CoV-2 has played a significant role in the resurgence of COVID-19 epidemic waves in South Africa and across the globe. South Africa has a unique observation of the evolution of SARS-CoV-2, with distinct SARS-CoV-2 lineages dominating certain epidemic periods. This unique observation allows for an investigation of the detected SARS-CoV-2 lineages’ impact on COVID-19 transmissibility and severity through analysis of epidemiological data. In this study, inferential statistical analysis was conducted on South African COVID-19 epidemiological data to investigate the impact of SARS-CoV-2 lineages in the South African COVID-19 epidemiology. The general methodology in this study involved the collation of South African COVID-19 epidemiological data, the regression and normalisation of the epidemiological data, and inferential statistical analysis. This study shows that the evolution of SARS-CoV-2 resulted in an increase in COVID-19 transmissibility and severity in South Africa. The Delta SARS-CoV-2 VOC resulted in increased COVID-19 transmissibility in the South African population by 53.9 to 54.8 % more than the Beta SARS-CoV-2 VOC and the predominantly B.1.1.54, B.1.1.56 C.1 SA SARS-CoV-2 lineage cluster. The Beta SARS-CoV-2 VOC resulted in more severe COVID-19 in South Africa than the Delta SARS-CoV-2 VOC. While, both the Beta and Delta SARS-CoV-2 VOC resulted in more severe COVID-19 than the initial SARS-CoV-2 lineages detected in South Africa’s first epidemic wave period. The Delta, Beta SARS-CoV-2 VOCs and the predominantly B.1.1.54, B.1.1.56 C.1 SA SARS-CoV-2 lineage cluster were observed to cause similar COVID-19 hospital case fatality and discharge rates in South African hospitals.

## 1. Introduction

On the 11^th^ of March 2020, the World Health Organisation (WHO) declared the Coronavirus Disease 2019 (COVID-19) a global pandemic [1]. The COVID-19 pandemic has resulted in more than 4 809 907 deaths in the reporting period up to the 1^st^ of October 2021 [2]. Public health measures such as nationwide lockdowns aimed at reducing the transmission of COVID-19 have come at a great cost to the world economy [3]. While, there is global consensus on the health risk posed by COVID-19, ground-breaking vaccine developments, and a great drive towards the vaccination of the world population against COVID-19. There are however challenges that persist in controlling the Global COVID-19 transmission and severity. One challenge is the large disparity in access to vaccines between the developing and developed worlds [4]. Another challenge is the emergent of SARS-CoV-2 lineages and sub-lineages (variants) with increased transmissibility [5]. Lineages and sub-lineages are a series of entities (in this case genetic) forming a single line of direct ancestry and descent [6]. Emerging SARS-CoV-2 variants have been attributed to the occurrence of secondary and tertiary COVID-19 epidemic waves and also threatening vaccine efforts due to their immune invasiveness [7].

SARS-CoV-2 is the virus that causes COVID-19 upon infecting a human host. SARS-CoV-2’s early characterisation was reported as a positive pan-Betacoronavirus from bronchoalveolar lavage samples and its whole genome sequence acquired through Illumina and nanopore sequencing. It was identified to have features typical of Coronaviruses belonging to the Betacoronavirus 2B lineage. Betacoronaviruses belong to the subfamily *Othrocoronavirinae* of the *Coronaviridae* family, order *Nidovirales*. A close relationship was found between SARS-CoV-2 with the BatCoV Rat G13 virus (from the bat (*Rhinolophus affinis*)) at 96 % identity [8]. The genetic sequence of SARS-CoV-2 was shared on the 12^th^ of January 2020 [9,10]. Coronaviruses are positive-stranded Ribonucleic Acid (RNA) viruses. The presence of spike glycoproteins in their envelope gives them a crown-like appearance under an electron microscope [11]. The genome of SARS-CoV-2 is a single positive-stranded RNA approximately 29 903 bases (nucleotides) pairs in length [9,10,12,13]. There are at least 50 different open reading frames (ORFs) in the SARS-CoV-2 genome which include start codon (AUG) and stop codons (UAG, UAA, UGA). Origin of transcription sequences allow for SARS-CoV-2 to encode for 50 protiens with structural, non-structural and accessory functions [10,14,15]. Two-thirds of the SARS-CoV-2 genome encodes for the two main transcription units, ORF1a and ORF1b which are attributed in the encoding of polyproteins, PP1a and PP1ab respectively. PP1ab contains ORFs for 16 nonstructural proteins (Nsp1-Nsp16). Non-structural proteins have functions in replication, proofreading, translation, suppressing host proteins, blocking immune responses, and stabilisation [10,15]. One-third of the SARS-CoV-2 genome encodes the structure and accessory proteins. Accessory genes are positioned along with the structural genes. There are at least nine ORFs for accessory proteins in the SARS-CoV-2 genome. Accessory proteins do not play a significant role in viral replication but play a role in the interaction between the viral genome and the host particularly blocking the production of cytokines [10,14,15]. With regards to structure, there are four main structural proteins in SARS-CoV-2. These are the Spike (S), Membrane (M), Envelope (E), and the Nucleocapsid (N) proteins. The Nucleocapsid proteins coat the SARS-COV-2 RNA for genetic protection [10,13–15].

During infection, the Spike glycoprotein undergoes cleavage (unique furin-like cleavage site (FCS)) into amino (N) terminal S1 and carboxyl (C)-terminal S2 subunits [10,16]. The SI subunit facilitates the incorporation of SAR-CoV-2 into the host cell while the S2 subunit is responsible for membrane fusion. The S1 subunit has a receptor binding (RBD) and N-terminal (NTD) domain. SARS-CoV-2 binds to the human angiotensin-converting enzyme 2 (ACE2) receptors which are abundant in the respiratory epithelium but are also expressed by other cells such as the upper oesophagus, enterocytes (ileum), myocardial cells, proximal tubular cells (kidney), and urothelial cells (bladder). The binding is achieved through the SARS-CoV-2 Spike Protein (S1). Once the attachment has been achieved priming of the SARS-CoV-2 Spike Protein (S2) by the host transmembrane serine protease 2 (TMPRSS2) allows for viral cell entry and replication endocytosis [10,13].

One of the first reported SARS-CoV-2 genomes was the Wuhan-Hu-1 (MN908947.3) (sampled on the 26^th^ of December 2019) [9] and the Wuhan/WH04/2020 [17] strains. In July 2020, a dynamic nomenclature for SARS-CoV-2 lineage assignment was designed by Rambaut et al, 2020 [12,18] and an algorithm to outline links in viral genomes developed known as the *Phylogenetic Assignment of Named Global Outbreak LINeages* (PANGOLIN) [18,19]. Based on this classification, 2 major lineages can be identified in the main phylogeny of SARS-CoV-2, lineages A and B. Lineage A can be defined based on the Wuhan/WH04/2020 genome sharing 2 nucleotides (positions 8 782 in ORF1ab and 28 144 in ORF8) while lineage B has different nucleotides present at those sites such as the Wuhan-Hu-1 genome [12,18]. By the 20^th^ of December 2020, there were 261 487 whole or partial genome sequences publicly available, shared through the Global Initiative on Sharing Avian Influenza Data (GISAID) [12]. From this genomic data, sub-lineages of lineages A and B were identified. Two main lineages were also identified, lineage C and D, and these were reassigned to the classification of lineage B. More than 266 lineages/sub-lineages were identified globally [12,18]. In the year 2020, lineage B and sub-lineage B.1 were the most reported in genetic samples worldwide [12].

Whole-genome sequencing of 104 strains of SAR-CoV-2 from patients with COVID-19 symptom onsets in the period of December 2019 to Mid-February 2020 showed 99.9 % homology, without significant mutations [8]. However, the rapid spread of SARS-CoV-2 has allowed the virus opportune replications to evolve into different lineages and sub-lineages. RNA viruses are prone to genetic evolution with resultant genomes that have different characteristics than the ancestral strain [13]. Coronaviruses including SARS-CoV-2 can proofread during replication however this has not limited the mutation of the SARS-CoV-2 genome in forming new lineages and sub-lineages [20]. To prioritise global monitoring and research and to inform the ongoing response to the COVID-19 pandemic, SARS-CoV-2 variants have been characterised as either ‘Variants of Concern’ (VOCs)’ or ‘Variants of Interest’ (VOIs)’. The main characteristics of VOCs are that they have evidence of an increase in transmissibility, more severe disease that leads to increased hospitalisation or deaths, therefore reducing the effectiveness of public health and social measures [21]. Additionally, VOCs significantly reduce the neutralisation of antibodies generated during previous infection or vaccination which ultimately reduces the effectiveness of treatments or vaccines, or diagnostic detection [22]. Variants of Interest (VOIs) are variants whose changes have predicted genetic markers that are known to affect virus characteristics such as transmissibility, disease severity, immune escape, diagnostic or therapeutic escape [21]. They are also identified to have a predictable increase in transmissibility or disease severity thus having an apparent epidemiological impact to suggest an emerging risk to global public health [21,22]. The SARS-CoV-2 variants that have been characterised as VOCs by WHO are the Alpha (B.1.1.7), Beta (B.1.351, B1.351.2, B.1.351.3), Gamma (P.1, P.1.1, P.1.2, P.1.4, P1.6, P.1.7), and Delta (B.1.617.2, AY.1, AY.2, AY.3, AY.3.1) SARS-CoV-2 lineages (WHO, 2021; CDC, 2021). While, the variants that have been characterised as VOIs are the Eta (B.1.525), Iota (B.1.526), Kappa (B.1.617.1), and Lambda (C.37) SARS-CoV-2 lineages [21].

The Alpha SARS-CoV-2 VOC (B.1.1.7 lineage) (formerly GR/501Y.V1) was reported in December 2020 based on United Kingdom whole-genome sequence samples from patients with positive laboratory tests of SARS-CoV-2 [23,24]. This lineage was also detected in commercial assays due to the absence of the S gene in PCR samples. The B.1.1.7 lineage had 17 mutations in its genome. Eight of these mutations were in the Spike protein at Δ69-70 deletion, Δ144 deletion, N501Y, A570D, P681H, T716I, S982A, and D1118H [13,23,25]. The mutation at N501Y showed increased affinity by the Spike protein in SARS-CoV-2 to the ACE 2 receptors. The B.1.1.7 lineage was reported to be 43 % to 82 % more transmissible and became the dominant SAR-CoV-2 lineage in the UK [26]. The Gamma SARS-CoV-2 variant (P.1, P.1.1, P.1.2, P.1.4, P1.6, P.1.7 lineages) (formerly GR/501Y.V3.) was identified in December 2020 in Brazil [27] and the United States(US) in January 2021 [13]. The P.1 lineage had 10 mutations in the Spike protein at 18F, T20N, P26S, D138Y, R190S, H655Y, T1027I V1176, K417T, E484K, and N501Y. Three of those mutations (L18F, K417N, E484K) were located in the RBD [27]. The Gamma SARS-CoV-2 variant showed reduced neutralization by monoclonal antibody therapies, post-vaccination, and convalescent sera [28]. The Delta (B.1.617.2, AY.1, AY.2, AY.3, AY.3.1 lineage) and the Kappa (B.1.617.1 lineage) SARS-CoV-2 variants were first identified in December 2020 in India [29]. The Delta SARS-CoV-2 variant was initially considered a VOI by the WHO but was subsequently classed as a VOC during the deadly second COVID-19 epidemic wave in India in April 2021 [30,31]. The Delta SARS-CoV-2 VOC had 10 mutations in the Spike protein at T19R, (G142D*), 156del, 157del, R158G, L452R, T478K, D614G, P681R, and D950N [29]. While the Kappa SARS-CoV-2 VOI had Spike protein mutations of interest at T95I, G142D, E154K, L452R, E484Q, D614G, P681R, and Q1071H [32]. The Delta SARS-CoV-2 variant was also detected in March 2021 in the US and became the dominant SARS-CoV-2 lineage in the US. Prior, the Eta (B.1.525 lineage), and Iota (B.1.526 lineage) SARS-CoV-2 VOIs were detected in the US in November 2020 [13]. They had Spike protein mutations of interest at B.1.525: A67V, Δ69/70, Δ144, E484K, D614G, Q677H, F888L and B.1.526: L5F*, T95I, D253G, S477N*, E484K*, D614G, A701V* respectively [33,34]. While in South America, the Lambda (C.37 lineage) SARS-CoV-2 variant was detected in Peru and classed as a VOI due to its increased presence in SARS-CoV-2 genetic samples from the region [35–37].

Of interest in this study is the impact of the SARS-CoV lineages and sub-lineages in the COVID-19 epidemic waves in South Africa. Since the importation of SARS-CoV-2 in South Africa, with the first reported COVID-19 case on the 5^th^ of March 2020, South Africa has observed 3 consecutive COVID-19 epidemic waves [2,38]. The first COVID-19 epidemic wave in South Africa was observed between the reporting periods of 5 March to 30 September 2020 with a peak of 173 587 Active COVID-19 cases and a peak date of 26 July 2020 [39,40]. The second COVID-19 epidemic wave in South Africa resurged on the 1^st^ of December 2020 with 212 529 peak Active COVID-19 cases and a peak date of 15 January 2021 [41]. On the 27th of April 2021, the third COVID-19 epidemic wave in South Africa resurged. The third COVID-19 epidemic wave in South Africa had two observed peaks on the 10^th^ of July and on the 26^th^ of August 2021 with 211 052 and 169 039 Active COVID-19 cases respectively lasting up to the end of September 2021 [2]. The response by the Government of South Africa to the COVID-19 epidemic was the establishment of a National Coronavirus Command Council to oversee the epidemic, the use of health policy measures including NPIs to try to mitigate the transmission of COVID-19 and the implementation of COVID-19 vaccination programmes to try to vaccinate the South African population against COVID-19 [42–47]. As of 19 September 2021, 15.8 million people have been vaccinated against COVID-19 in South Africa mainly with the Pfizer-BioNTech (Comirnaty) and the Johnson & Johnson/Janssen COVID-19 vaccines [48].

“Globally, systems have been established and are being strengthened to detect ‘signals’ of potential VOIs or VOCs and assess these based on the risk posed to global public health” [21]. In South Africa, the Network for Genomics Surveillance in South Africa (NGS-SA) was formed to understand the spread of SARS-CoV-2 [49]. The NGS-SA was launched in June 2020 and this consortium comprised of the National Health Laboratory Services (NHLS) and associated academic institutions, the National Institute for Communicable Diseases (NICD), KZN Research Innovation & Sequencing Platform (KRISP), Stellenbosch University (SU), University of Cape Town (UCT). University of Free State (UFS), University of Pretoria (UP), University of KwaZulu-Natal (UKZN), and the South African National Bioinformatics Institute (SANBI) [50]. In the first COVID-19 epidemic wave in South Africa, 16 SARS-CoV-2 lineages specific to South Africa were identified from 1 365 high-quality whole genomes [49]. From these 16 lineages, three main clusters (B.1.1.54, B.1.1.56, and C.1 SARS-CoV-2 lineages) were identified to have caused approximately 42% of the SARS-CoV-2 infections in South Africa [49]. Another sub lineage specific to South Africa was the B.1.106 lineage that emerged in the Kwa-Zulu Natal province in a nosocomial outbreak during the first COVID-19 epidemic wave [49]. The prevalence of this sub-lineage decreased as a result of control measures [49,51]. The C.I lineage (first identified C lineage of SARS-CoV-2) was the most geographically spread lineage in South Africa’s first COVID-19 epidemic wave [49]. Before the resurgence of the second COVID-19 epidemic wave in South Africa, the Beta (B.1.351, B1.351.2, B.1.351.3 lineages) SARS-CoV-2 VOC (formerly GR/501Y.V2) was identified in an analysis of 2 704 South African SARS-CoV-2 genotypes (samples up to 14^th^ of December 2020) from the GISAID database. The Beta (B.1.351 lineage) SARS-CoV-2 VOC was detected in samples collected in the months of October 2020 [52]. The Beta (B.1.351 lineage) SARS-CoV-2 VOC had 9 mutations in the Spike protein at L18F, D80A, D215G, R246I, K417N, E484K, N501Y, D614G, and A701V. Three of those mutations (K417N, E484K, N501Y)) were located in the RBD. The Beta SARS-CoV-2 lineage became the dominant lineage in South Africa’s second COVID-19 epidemic wave rapidly replacing the three main clusters (B.1.1.54, B.1.1.56 and C.1 SARS-CoV-2 lineages) identified during the first COVID-19 epidemic wave [52]. A study by [53] showed that the Beta SARS-CoV-2 lineage required a half-maximal inhibitory concentration (*IC5*0) 6 to 200 fold higher than the lineages identified in the first wave. Indicating that the Beta SARS-CoV-2 variant may escape neutralising antibody response developed from prior infections [53]. In the resurgence of the third COVID-19 epidemic wave in South Africa, four SARS-CoV variants were identified which were the Alpha, Beta, Eta, and Delta SARS-CoV variants. Genomic data for South African samples, identified 65 % of 1147 whole genomes from May 2021 to be the Beta SARS-CoV-2 variant. The Alpha, Delta, and Eta SARS-CoV-2 variants accounted for 6 %, 16 %, and 1 % of those samples respectively. In June 2021, with 2931 genetic sequences in that period, the Delta SARS-CoV-2 variant had become the dominant variant in samples collected in South Africa at 66 % while the Beta and Alpha SARS-CoV-2 variants accounted for 16 % and 4 % respectively [54]. By the end of South Africa’s third COVID-19 epidemic wave in September 2021, the Delta SARS-CoV-2 variant accounted for 96 % of the 186 whole-genome sampled in that period while the C1.2 SARS-CoV-2 lineage account for 1 % of those samples [54]. The C1.2 SARS-CoV-2 lineage was a new South Africa specific SARS-CoV-2 lineage identified in South African samples in the month of May 2021 and evolved from the C.1 SARS-CoV-2 lineage. The C.1.2 SARS-CoV-2 lineage had several mutations with multiple substitutions (R190S, D215G, N484K, N501Y, H655Y and T859N) and deletions (Y144del, L242-A243del) in the Spike protein. There was also an accumulation of additional mutation (C136F, Y449H and N679K) in the C.1.2 SARS-CoV-2 thought to likely impact neutralisation sensitivity. The C.1.2 has been detected across the majority of South African provinces and in seven other countries [55].

The evolution of SARS-CoV-2 has played a significant role in the resurgence of COVID-19 epidemic waves in South Africa and across the globe. South Africa has a unique observation of the evolution of SARS-CoV-2, with distinct SARS-CoV-2 lineages dominating certain epidemic periods. This unique observation allows for an investigation of the detected SARS-CoV-2 lineages’ impact on COVID-19 transmissibility and severity through analysis of epidemiological data. In this study, inferential statistical analysis was conducted on South African COVID-19 epidemiological data to investigate the impact of SARS-CoV-2 lineages in the South African COVID-19 epidemiology.

## 2. Methodology

The general methodology in this study involved the collation of South African COVID-19 epidemiological data, the regression and normalisation of the epidemiological data, and inferential statistical analysis.

### 2.1 Collation of South African COVID-19 Epidemiological Data

#### 2.1.1 South African COVID-19 Testing and Reported Case Data

South African COVID-19 reported case data (Cumulative and Daily COVID-19 Cases, Recovered and Deaths) for the reporting period of 22 January 2020 to 19 September 2021 were obtained from the Johns Hopkins University (JHU) Center for Systems Science and Engineering (CSSE) COVID-19 Database [2]. While the South African COVID-19 testing data (Cumulative and Daily COVID-19 tests) were obtained from the Our World In Data project [56] for the reporting period of 14 February 2020 to 16 September 2021. COVID-19 Active cases were then calculated using Equation 1 per reported date:

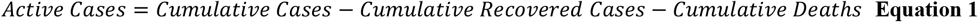

#### 2.1.2 South African COVID-19 Hospitalisation and Excess (Natural) Deaths Data

South African COVID-19 hospitalisation data were obtained from the National Institute for Communicable Diseases (NICD) DATCOV surveillance system for the period of 24 May 2020 to 19 September 2021. The NICD sentinel hospital surveillance system was designed to monitor and describe trends of COVID-19 hospitalizations and the epidemiology of hospitalized patients in South Africa [57]. The South African COVID-19 hospitalisation data was composed of the Number of Facilities Reporting, Admission Status Data (Daily COVID-19 Hospital Admitted Cases, Hospitalised in High Care, Intensive Care Unit, Isolation Ward, On Oxygen and Ventilator), Cumulative COVID-19 Admission Age Profile, Cumulative COVID-19 Hospital Deaths Age Profile and Cumulative COVID-19 Patients Discharged Alive. The COVID-19 Hospital Daily Discharge Rate (DR) and Case Fatality Rate (CFR) were then calculated based on the methodology in [58]. First, the COVID-19 daily patient discharge and deaths were calculated using Equation 2 and Equation 3 respectively:

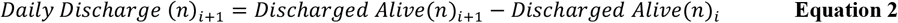

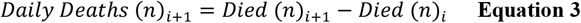

Where n is the number of patients and i is the reported case date. Then the COVID-19 Hospital Daily Discharge Rate (DR) and Case Fatality Rate (CFR) were then calculated using Equation 4 and Equation 5 respectively.

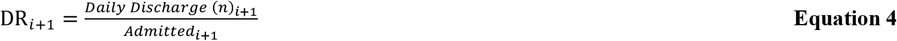

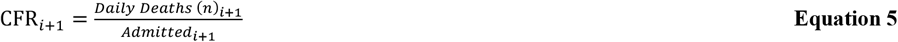

Where n is the number of patients and i is the reported case date. Excess mortality is an account of deaths from all causes relative to expected deaths based on previous trends/observations. Excess mortality/deaths allow for accounting for miscounted or underreported COVID-19 Deaths and indirect Deaths related to the COVID-19 pandemic [59]. South African Weekly Natural and Excess (Natural) Deaths were obtained from the South African Medical Research Council (SAMRC) [60] for the reporting period of 29 December 2019 to 18 September 2021. The Weekly Unreported Excess Deaths (Natural) to COVID-19 Death Ratio (ECDR) was then calculated based on the methodology in [58] using Equation 6:

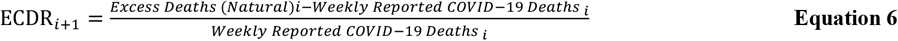

Where i is the Weekly Reported Date.

#### 2.1.3 Stratification of South African COVID-19 Epidemiological Data

To draw inferential comparisons regarding the impact of the evolution of SARS-CoV-2 in the South African COVID-19 epidemiology, the collated South African COVID-19 epidemiological data in Section 2.1.1 and 2.1.2 was stratified based on the observed COVID-19 epidemic wave period 1, 2 and 3 in South Africa. Based on a review of [49,52,54], Table 1 was generated summarising the Sotuh African (SA) SARS-CoV-2 lineage clusters observed in the genomic data in the observed COVID-19 epidemic wave periods. The South African COVID-19 epidemic wave period 1, 2 and 3 were classed as reported case data in the period of 5 March 2020 to 30 September 2020, 01 October 2020 to 26 April 2021 and 27 April 2021 to 19 September 2021 respectively. The labels of stratified variables were given a reference of “_1”, “_2”, “_3” for the three respectively epidemic periods. By this stratification, the cluster of lineages identified in Table 1 was assumed to be the SARS-COV-2 lineages resulting in the respective COVID-19 epidemic waves in South Africa. For Cumulative Epidemiological Data (South African Cumulative COVID-19 Admission Age Profile, Cumulative COVIID-19 Hospital Deaths Age Profile and Cumulative COVID-19 Patients Discharged Alive) the data was adjusted using Equation 7 to remove the cumulative data from the previous COVID-19 epidemic period.

**Table 1:** SARS-CoV-2 lineages identified in genome samples obtained from patients with laboratory-confirmed COVID-19 for the three observed South African COVID-19 epidemic wave periods. Data extracted from [49,52,54]

| COVID-19 Epidemic Wave | Epidemic Period | Genome Samples | Genome Sampling Period | SARS-CoV-2 Lineages in Samples | SARS-CoV-2 Lineages in Epidemic Period | SA Specific SARS-CoV-2 Lineages | Emerging Dominant SARS-CoV-2 Lineages Spatiotemporal Phylogeographic | Emerging Dominant SARS-CoV-2 Lineages in Samples (%) | SARS-CoV-2 Lineage Cluster |
| --- | --- | --- | --- | --- | --- | --- | --- | --- | --- |
| 1 | 2020/03/05-2020/09/30 | 1365 | 2020/03/06-2020/08/26 | A, B, B.1, B.1.1, B.1.1.1, B.1.1.10, B.1.1.26, B.1.1.34, B.1.1.35, B.1.1.40, B.1.1.52, B.1.1.53, B.1.1.54, B.1.1.56, B.1.1.57, B.1.1.62, B.1.1.64, B.1.1.66, B.1.1.67, B.1.1.75, B.1.1.106, B.1.1.132, B.1.1.133, B.1.1.140, B.1.1.144, B.1.2.3, B.1.35, B.1.5, B.1.5.16, B.1.5.19, B.1.5.22, B.1.79, B.1.8, B.1.98, B.2, B.2.1, B.2.2, B.3, B.6, B.9, C.1, C.2 | A, B, B.1, B.1.1, B.1.1.1, B.1.1.10, B.1.1.26, B.1.1.34, B.1.1.35, B.1.1.40, B.1.1.52, B.1.1.53, B.1.1.54, B.1.1.56, B.1.1.57, B.1.1.62, B.1.1.64, B.1.1.66, B.1.1.67, B.1.1.75, B.1.1.106, B.1.1.132, B.1.1.133, B.1.1.140, B.1.1.144, B.1.2.3, B.1.35, B.1.5, B.1.5.16, B.1.5.19, B.1.5.22, B.1.79, B.1.8, B.1.98, B.2, B.2.1, B.2.2, B.3, B.6, B.9, C.1, C.2 | B.1.1.34, B.1.1.40, B.1.1.52, B.1.1.53, B.1.1.54, B.1.1.56, B.1.1.57, B.1.1.62, B.1.1.66, B.1.1.106, B.1.1.133, B.1.1.144, B.1.5.16, B.1.5.19, C.1, C.2 | **B.1.106** (April 2020), **B.1.1.54** (poor temporal signaling), **B.1.1.56** (mid-March 2020 (95% highest posterior density (HPD) 2020-02-15 to 2020-03-30)), **C.1** (early May 2020 (95% HPD 2020-04-24 to 2020-05-24)) | B.1.1.54, B.1.1.56, C.1 (42.1 % of total genomes) | 1 |
| 2 | 2020/10/01-2021/04/26 | 2 573 | 2020/03/05-2020/12/10 | A, B, B.1, B.1.1, B.1.1.1, B.1.1.10, B.1.1.109, B.1.1.117, B.1.1.119, B.1.1.156, B.1.1.159, B.1.1.163, B.1.1.2, B.1.1.220, B.1.1.252, B.1.1.254, B.1.1.316, B.1.1.34, B.1.1.4, B.1.1.40, B.1.1.52, B.1.1.53, B.1.1.54, B.1.1.56, B.1.1.57, B.1.1.62, B.1.1.64, B.1.1.67, B.1.1.75, B.1.1.84, B.1.1.91, B.1.1.99, B.1.1.106, B.1.1.124, B.1.1.131, B.1.1.140, B.1.1.141, B.1.1.144, B.1.1.153, B.1.1.178, B.1.1.83, B.1.222, B.1.225, B.1.237, B.1.242, B.1.275, B.1.315, B.1.36, B.1.5, B.1.5.16, B.1.5.19, B.1.5.31, B.1.8, B.1.98, B.3, B.39, B.40, B.52, B.6, C.1, C.2, C.6, C.9, H.1 | B.1.1.1, B.1.1.119, B.1.1.54, B.1.1.56, C.1, B.1.351, B.1.1.156, B.1.1.252, B.1.1.254, B.1.1.34, B.1.1.57, B.1.1.220, B.1.1.44, B.1.237, B.1.351, B.1.5, B.1.5.19, B.1.5.31, C.2, C.9, B.1.106 | B.1.1.34, B.1.1.54, B.1.1.56, C.1, B.1.351, B.1.1.57, B.1.1.106, B.1.1.44, B.1.5.19, C.2 | **B.1.351** (early August (95% highest posterior density ranging from the middle of July to the end of August 2020)) | B.1.351 (Approximately 90 % of Samples by End of November 2020) | 2 |
| 3 | 2021/04/27-2021/09/19 | 9462 | 2021/05-2021/09 | B.1.351, B.1.617.1, B.1.617.2, B.1.525, B.1.1.7, C.1.2, Other (1.74 %), unassigned (5.20 %) | B.1.351, B.1.617.1, B.1.617.2, B.1.525, B.1.1.7, C.1.2 | B.1.351, C.1.2 |  | B.1.617.2 (Approximately Over 65 % in June, 85 % in July and over 90 % of samples in August and September 2021) | 3 |

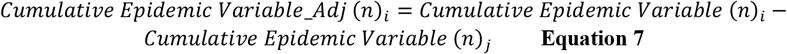

Where n is the number of patients, i is the reported date and j is the last reported date of the previous COVID-19 epidemic period. The stratification of data in this study was done by splitting the data using the epidemic period variable in the IBM SPSS STATISTICS 27 Software.

### 2.2 Regression of South African COVID-19 Epidemiological Data

In an epidemic, several factors influence the outcome of the observed epidemic. This includes testing, reporting capacity and population movement for a human to human infectious diseases such as COVID-19 (disease daily effective contact rate) [61]. For inferential statistical comparative analysis to be conducted between epidemiological data from different epidemic periods, the covariance of the epidemiological data needs to be accounted for in the analysis. Covariance is the measure of the combined variability of two random variables [62]. The independent variable in the covariance is a predictor variable influencing the outcome of a dependent variable.

#### 2.2.1 Linear Regression of COVID-19 Daily Tests and Cases

[58] showed that there was a positive correlation between South African COVID-19 Daily Tests and Cases in the first COVID-19 epidemic period. The South African COVID-19 daily testing was also shown not to be consistent throughout the period. To understand the covariance between South African COVID-19 daily tests and cases, Descriptive Statistical Analysis, Bivariate Analysis using the Two-tailed Pearson and Spearman tests and Analysis of Variance using the Univariate General Linear Model was conducted on the COVID-19 Daily Tests and Cases (Dependent Variable) using the IBM SPSS STATISTICS 27 Software for the three COVID-19 epidemic periods. Table 2 shows that the mean COVID-19 daily tests in the first, second and third South African COVID-19 epidemic wave period were 20 575±14 062, 31 046±14 115 and 46 822±18 460 respectively. The value of these means indicates the difference in testing capacity in the three epidemic periods. The Skewness values shown in Table 2 indicate that the normal distribution of the South African COVID-19 Daily Tests and Cases were positively skewed with the COVID-19 Daily Cases being more skewed than the Daily Tests. The skewness of COVID-19 Daily Cases was expected considering that the distribution is lognormal (Galton distribution) with differential (calculus) relations to normally distributed random variables such as the COVID-19 effective daily contact rate in this probability space. The application of differentials to model COVID-19 using random variables in stochastic COVID-19 epidemiological modelling was shown to be effective in modelling the COVID-19 epidemiology in South Africa [58,63,64].

**Table 2:** Statistical Sample Number (N), Range, Minimum, Maximum, Mean, Standard Deviation (Std Deviation), Variance, Coefficient of Skewness, Kurtosis Coefficient of COVID-19 Daily Tests, New Cases in the First, Second and Third COVID-19 Epidemic Wave in South Africa **(Descriptive Statistics**)

| Descriptive Statistics |  |  |  |  |  |  |  |  |  |  |  |  |  |
| --- | --- | --- | --- | --- | --- | --- | --- | --- | --- | --- | --- | --- | --- |
| COVID-19_Epidemic_Wave |  | N | Range | Minimum | Maximum | Mean |  | Std. Deviation | Variance | Skewness |  | Kurtosis |  |
|  |  | Statistic | Statistic | Statistic | Statistic | Statistic | Std. Error | Statistic | Statistic | Statistic | Std. Error | Statistic | Std. Error |
| 1 | COVID-19_Daily_Tests | 203 | 56659 | 4 | 56663 | 20575 | 987 | 14062 | 197739959 | 0.474 | 0.171 | -0.590 | 0.340 |
|  | COVID-19_Daily_New_Cases | 253 | 13944 | 0 | 13944 | 2665 | 235 | 3736 | 13958924 | 1.620 | 0.153 | 1.564 | 0.305 |
|  | Valid N (listwise) | 203 |  |  |  |  |  |  |  |  |  |  |  |
| 2 | COVID-19_Daily_Tests | 201 | 66765 | 10402 | 77167 | 31046 | 996 | 14115 | 199227395 | 1.164 | 0.172 | 1.179 | 0.341 |
|  | COVID-19_Daily_New_Cases | 208 | 21543 | 437 | 21980 | 4336 | 349 | 5034 | 25340444 | 1.793 | 0.169 | 2.386 | 0.336 |
|  | Valid N (listwise) | 201 |  |  |  |  |  |  |  |  |  |  |  |
| 3 | COVID-19_Daily_Tests | 119 | 116166 | 16194 | 132360 | 46822 | 1692 | 18460 | 340760831 | 1.060 | 0.222 | 2.884 | 0.440 |
|  | COVID-19_Daily_New_Cases | 127 | 26485 | 0 | 26485 | 9459 | 537 | 6047 | 36560682 | 0.418 | 0.215 | -0.417 | 0.427 |
|  | Valid N (listwise) | 119 |  |  |  |  |  |  |  |  |  |  |  |

Table 3 shows that the Pearson (Spearman) Correlation Coefficients between COVID-19 Daily Tests (Independent Variable) and COVID-19 Daily Cases (Dependent Variable) in the First, Second and Third COVID-19 Epidemic Wave in South Africa were 0.910 (0.955), 0.877 (0.751) and 0.821 (0.867) respectively. The values of the Pearson (Spearman) Correlation Coefficients observed indicate a strong positive correlation between COVID-19 Daily Tests and Cases with p-values more than 95 % confidence. The F values are shown in Table 4 for the Mean Square Regression and Residual between COVID-19 Daily Tests (Independent Variable) and COVID-19 Daily Cases (Dependent Variable) indicate that the residual error between the linear predicted value and the actual values are relatively small. These results in Section 2.2.1 show that COVID-19 Daily Cases in South Africa were dependent on the COVID-19 Daily test with a positive linear relationship based on the Standardized Coefficients shown in Table 5.

**Table 3:**
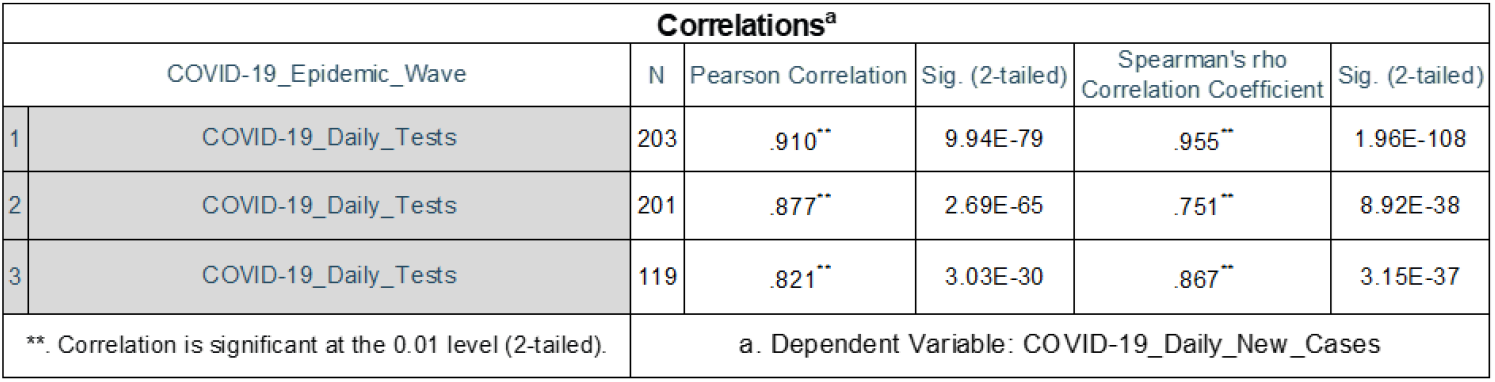
Pearson and Spearman’s Correlation Coefficients and P-Values (Sig. (2-tailed)) between COVID-19 Daily Tests (Independent Variable) and COVID-19 Daily New Cases (Dependent Variable) in the First, Second and Third COVID-19 Epidemic Wave in South Africa **(Correlations**)

**Table 4:** Sum of Squares, Degrees of Freedom (Df_, Mean Square, F values between Mean Square Regression and Residual (F), between COVID-19 Daily Tests (Independent Variable) and COVID-19 Daily New Cases (Dependent Variable) in the First, Second and Third COVID-19 Epidemic Wave in South Africa **(Analysis of Variance (ANOVA)**)

| ANOVA<sup>a</sup> |  |  |  |  |  |  |  |
| --- | --- | --- | --- | --- | --- | --- | --- |
| COVID-19_Epidemic_Wave |  |  | Sum of Squares | df | Mean Square | F | Sig. |
| 1 | 1 | Regression | 2548543000 | 1 | 2548543000 | 967 | .000 |
|  |  | Residual | 529940020 | 201 | 2636518 |  |  |
|  |  | Total | 3078483020 | 202 |  |  |  |
| 2 | 1 | Regression | 4024599374 | 1 | 4024599374 | 664 | .000 |
|  |  | Residual | 1206837402 | 199 | 6064510 |  |  |
|  |  | Total | 5231436776 | 200 |  |  |  |
| 3 | 1 | Regression | 2999260614 | 1 | 2999260614 | 242 | .000 |
|  |  | Residual | 1451354776 | 117 | 12404742 |  |  |
|  |  | Total | 4450615390 | 118 |  |  |  |
| a. Dependent Variable: COVID-19_Daily_New_Cases |  |  |  |  |  |  |  |
| b. Predictors: (Constant), COVID-19_Daily_Tests |  |  |  |  |  |  |  |

**Table 5:**
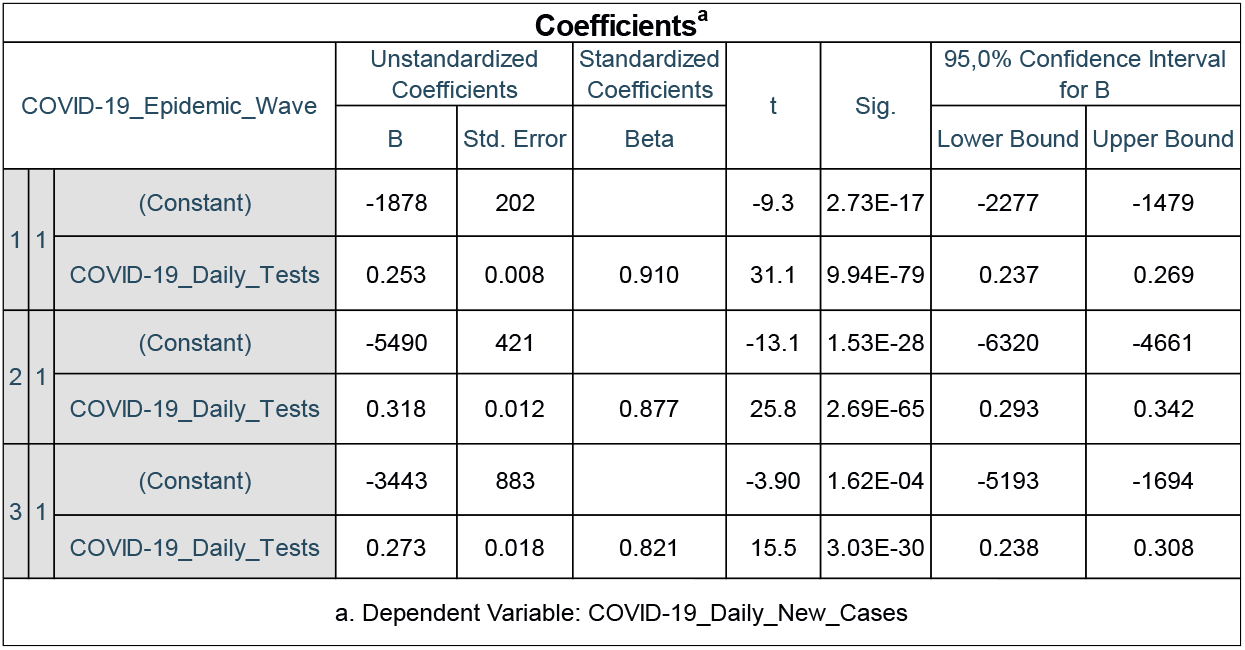
Unstandardized Coefficients (B, Std. Error), Standardized Coefficients (Beta), t-calculated (t), p-value calculated (Sig.) and 95.0 % Confidence Interval for B between COVID-19 Daily Tests (Independent Variable) and COVID-19 Daily New Cases (Dependent Variable) in the First, Second and Third COVID-19 Epidemic Wave in South Africa **(Coefficients**)

#### 2.2.2 Linear Regression of Number of Reporting Hospitals, COVID-19 Active and Hospitalised Cases

The NICD DATCOV surveillance system in South Africa only started publishing reports on the reporting date of 24 May 2020 thus data from 5 March 2020 to 23 May 2020 in the first COVID-19 epidemic wave period is missing. Data from 9 October 2020 to 26 October 2020 in the second COVID-19 epidemic wave period was also missing. The number of hospitals reporting to the NICD DATCOV surveillance system in South Africa’ first COVID-19 epidemic wave period was initially 204 facilities and the facilities increased to 551 by the end of this period. In South Africa’s second and third COVID-19 epidemic wave periods, the facilities reporting by the end of these periods were 646 and 664 respectively. To understand the covariance between South African COVID-19 Active Cases, Number of Facilities Reporting to the NICD DATCoV surveillance system and COVID-19 Hospital Admitted Cases (Dependent Variable), Bivariate Analysis using the Two-tailed Pearson and Spearman tests and Analysis of Variance using the Univariate General Linear Model was conducted on these variables using the IBM SPSS STATISTICS 27 Software for the three COVID-19 epidemic periods. Table 6 and Table 7 show that the Pearson (Spearman) Correlation and Standardized Coefficients between Facilities Reporting to NICD DATCoV (Independent Variable) and COVID-19 Hospital Admitted Cases (Dependent Variable) in the First, Second and Third COVID-19 Epidemic Wave in South Africa were 0.336 (0.391), 0.136 (−0.260), 0.948 (0.881) and 0.271, 0.193, 0.623 respectively. The values of the Standardized Coefficients, Pearson and Spearman’s Correlation Coefficients observed indicate a moderate positive correlation between the Number of Facilities Reporting to NICD DATCoV and COVID-19 Hospital Admitted Cases in the first and second epidemic period. A strong positive correlation for the third epidemic period was observed. Table 6 and Table 7 also show that the Pearson (Spearman) Correlation and Standardized Coefficients between COVID-19 Active Cases (Independent Variable) and COVID-19 Hospital Admitted Cases (Dependent Variable) in the First, Second and Third COVID-19 Epidemic Wave in South Africa were 0.932 (0.959), 0.873 (0.875), 0.904 (0.850) and 0.913, 0.885, 0.404 respectively.

**Table 6:**
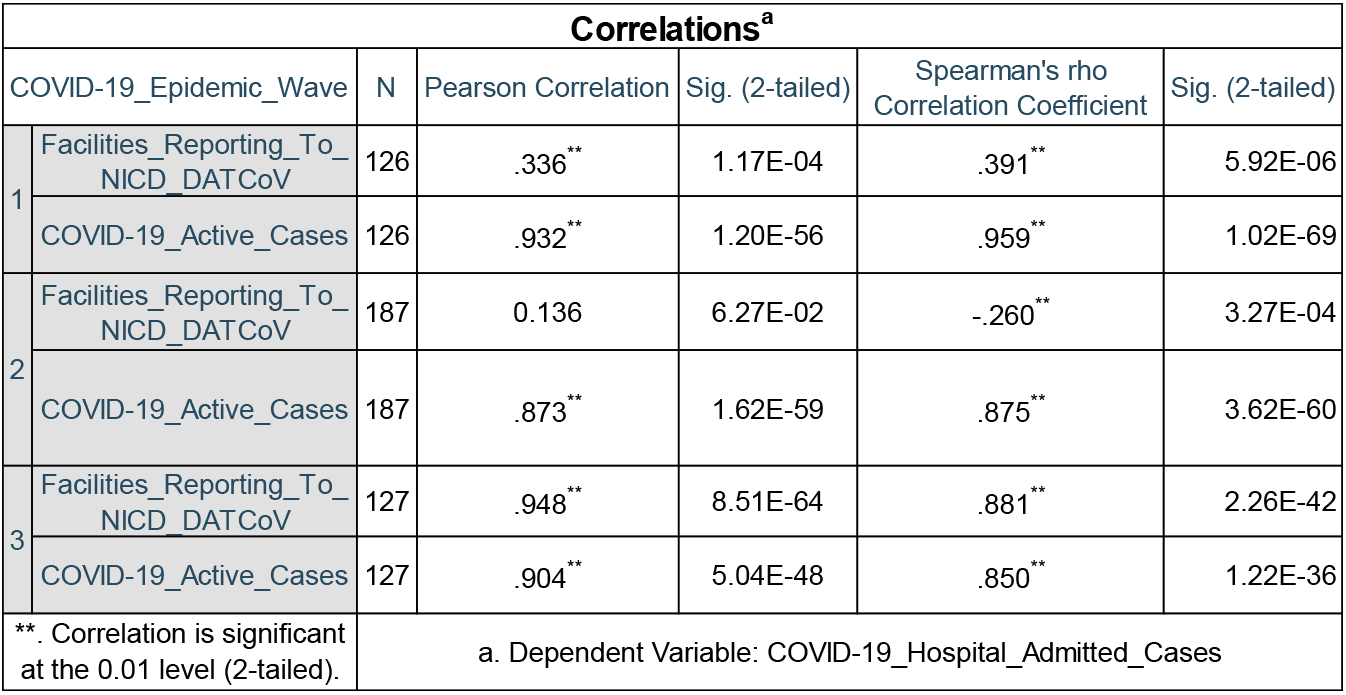
Pearson and Spearman’s Correlation Coefficients and P-Values (Sig. (2-tailed)) between Number of Facilities Reporting to the NICD DATCoV System (Independent Variable), COVID-19 Active Cases (Covariant Variable) and COVID-19 Hospital Admitted Cases (Dependent Variable) in the First, Second and Third COVID-19 Epidemic Wave in South Africa **(Correlations**)

| Correlations<sup>a</sup> |  |  |  |  |  |  |
| --- | --- | --- | --- | --- | --- | --- |
| COVID-19_Epidemic_Wave |  | N | Pearson Correlation | Sig. (2-tailed) | Spearman's rho<br>Correlation Coefficient | Sig. (2-tailed) |
| 1 | Facilities_Reporting_To_NICD_DATCoV | 126 | .336** | 1.17E-04 | .391** | 5.92E-06 |
|  | COVID-19_Active_Cases | 126 | .932** | 1.20E-56 | .959** | 1.02E-69 |
| 2 | Facilities_Reporting_To_NICD_DATCoV | 187 | 0.136 | 6.27E-02 | -.260** | 3.27E-04 |
|  | COVID-19_Active_Cases | 187 | .873** | 1.62E-59 | .875** | 3.62E-60 |
| 3 | Facilities_Reporting_To_NICD_DATCoV | 127 | .948** | 8.51E-64 | .881** | 2.26E-42 |
|  | COVID-19_Active_Cases | 127 | .904** | 5.04E-48 | .850** | 1.22E-36 |
| **. Correlation is significant at the 0.01 level (2-tailed). |  | a. Dependent Variable: COVID-19_Hospital_Admitted_Cases |  |  |  |  |

**Table 7:** Unstandardized Coefficients (B, Std. Error), Standardized Coefficients (Beta), t-calculated (t), p-value calculated (Sig.) and 95.0 % Confidence Interval for B between Number of Facilities Reporting to the NICD DATCoV System (Independent Variable), COVID-19 Active Cases (Covariant Variable) and COVID-19 Hospital Admitted Cases (Dependent Variable) in the First, Second and Third COVID-19 Epidemic Wave in South Africa **(Coefficients**)

| Coefficients<sup>a</sup> |  |  |  |  |  |  |  |  |  |
| --- | --- | --- | --- | --- | --- | --- | --- | --- | --- |
| COVID-19_Epidemic_Wave |  |  | Unstandardized Coefficients |  | Standardized Coefficients | t | Sig. | 95,0% Confidence Interval for B |  |
|  |  |  | B | Std. Error | Beta |  |  | Lower Bound | Upper Bound |
| 1 | 1 | (Constant) | -69.4 | 171 |  | -0.407 | 0.685 | -407 | 268 |
|  |  | Facilities_Reporting_To_NICD_DATCoV | 5.55 | 0.444 | 0.271 | 12.5 | 1.16E-23 | 4.68 | 6.43 |
|  |  | COVID-19_Active_Cases | 0.036 | 0.001 | 0.913 | 42.1 | 6.30E-75 | 0.034 | 0.037 |
| 2 | 1 | (Constant) | -24246 | 4729 |  | -5.13 | 7.42E-07 | -33577 | -14916 |
|  |  | Facilities_Reporting_To_NICD_DATCoV | 43.9 | 7.52 | 0.193 | 5.83 | 2.39E-08 | 29.0 | 58.7 |
|  |  | COVID-19_Active_Cases | 0.069 | 0.003 | 0.885 | 26.7 | 3.02E-65 | 0.064 | 0.074 |
| 3 | 1 | (Constant) | -219210 | 11232 |  | -19.5 | 1.28E-39 | -241442 | -196978 |
|  |  | Facilities_Reporting_To_NICD_DATCoV | 345 | 17.4 | 0.623 | 19.8 | 3.49E-40 | 310 | 379 |
|  |  | COVID-19_Active_Cases | 0.031 | 0.002 | 0.404 | 12.8 | 1.70E-24 | 0.026 | 0.035 |

a. Dependent Variable: COVID-19\_Hospital\_Admitted\_Cases

The values of the Standardized, Pearson and Spearman’s Correlation Coefficients observed indicate a strong positive correlation between COVID-19 Active and Hospital Admitted Cases. This result was expected based on probability theory. An increase in COVID-19 active cases increases the probability of stochastic proportions of COVID-19 severity including severe and critical COVID-19 thus higher hospitalisations. This correlation was also well demonstrated by stochastic COVID-19 epidemiological models such as in [58,63,64].

#### 2.2.3 Regression of the COVID-19 Daily Effective Contact Rate and Movement Restrictions (NPIs)

South Africa’s COVID-19 health policy response to the COVID-19 epidemic waves in South Africa was implemented in the form of National Lockdown Alert Level policies. The National Lockdown Alert Level policies were largely entry and exit screening at borders, limitations of movements and gatherings, closure/limitations of institutions and business activities, ban/limiting of alcohol and tobacco industries, isolation, quarantine of potentially infected persons, contact tracing protocols, use of personal protective equipment (PPE) and hygienic protocols [58]. In South Africa’s first COVID-19 epidemic wave period the National Lockdown Alert Levels **5** *(26 March to 30 April 2020)*, **4** *(1 May to 31 May 2020)*,**3** *(I June to 17 August 2020)*, and **2** *(18 August to 20 September 2020)* were implemented [65]. In South Africa’s second COVID-19 epidemic wave period the National Lockdown Adjusted Alert Levels **1** *(21 September to 28 December 2020; 1 March to 30 May 2021)* and **3** *(29 December 2020 to 28 February 2021)* were implemented. Whilst for South Africa’s third COVID-19 epidemic wave period the National Lockdown Adjusted Alert Levels **2** *(31 May to 15 June 2021; 13 September 2021)*, **3** *(16 June to 27 June 2021; 26 July to 12 September 2021)* and **4** *(28 June to 25 July 2021)* were implemented.

The daily effective contact rate is the average number of adequate contacts per infective per day. It is directly proportional to the reproductive number [61]. [58] showed the impact of NPIs particularly movement restriction on the COVID-19 daily effective contact rate in South Africa. In this study, a relationship between the community mobility of the South African population and the COVID-19 Daily Effective Contact Rate was established. [58] showed that an increase in 1 Alert Level in the South African National Lockdown Alert Levels was 30 % more effective in reducing population movement and resulted in a reduction in the COVID-19 Daily Effective Contact Rate by 4.13 % to 14.6 % compared to the preceding Alert Level. The study also showed that the National Lockdown Alert Levels 1 and 2 had a similar impact on the population movements in the South African communities in the first COVID-19 epidemic wave period [58]. Figure 1 shows the South African community mobility in the Retail and Recreation, Grocery and Pharmacy, Parks, Transit Stations, Workplaces and Residences locations in the three COVID-19 epidemic periods. The modulus mean movement change from baseline in South African locations in the period of 26 March to 25 May 2020 for the first COVID-19 epidemic wave period were higher than those observed in the second and third COVID-19 epidemic wave periods. This period corresponds to the implementation of the National Lockdown Alert Levels 5 and 4 in the first COVID-19 epidemic wave. In the second and third COVID-19 epidemic wave periods the highest alert levels implemented were the adjusted Alert Level 3 and 4 respectively. The adjustment in these Alert Levels resulted in more eased movement restrictions than its predecessors while the National Alert level 5 lockdown implemented in the first COVID-19 epidemic wave period had the hardest population movement restrictions experienced by the South African population. These differences resulted in the lower modulus mean movement change from baselines in South African locations in the second and third COVID-19 epidemic wave periods. Thus, caution must be taken when concluding the impact of SARS-CoV-2 lineages on the observed COVID-19 epidemic in South Africa as the different movement restrictions might have also had an impact.

**Figure 1:**
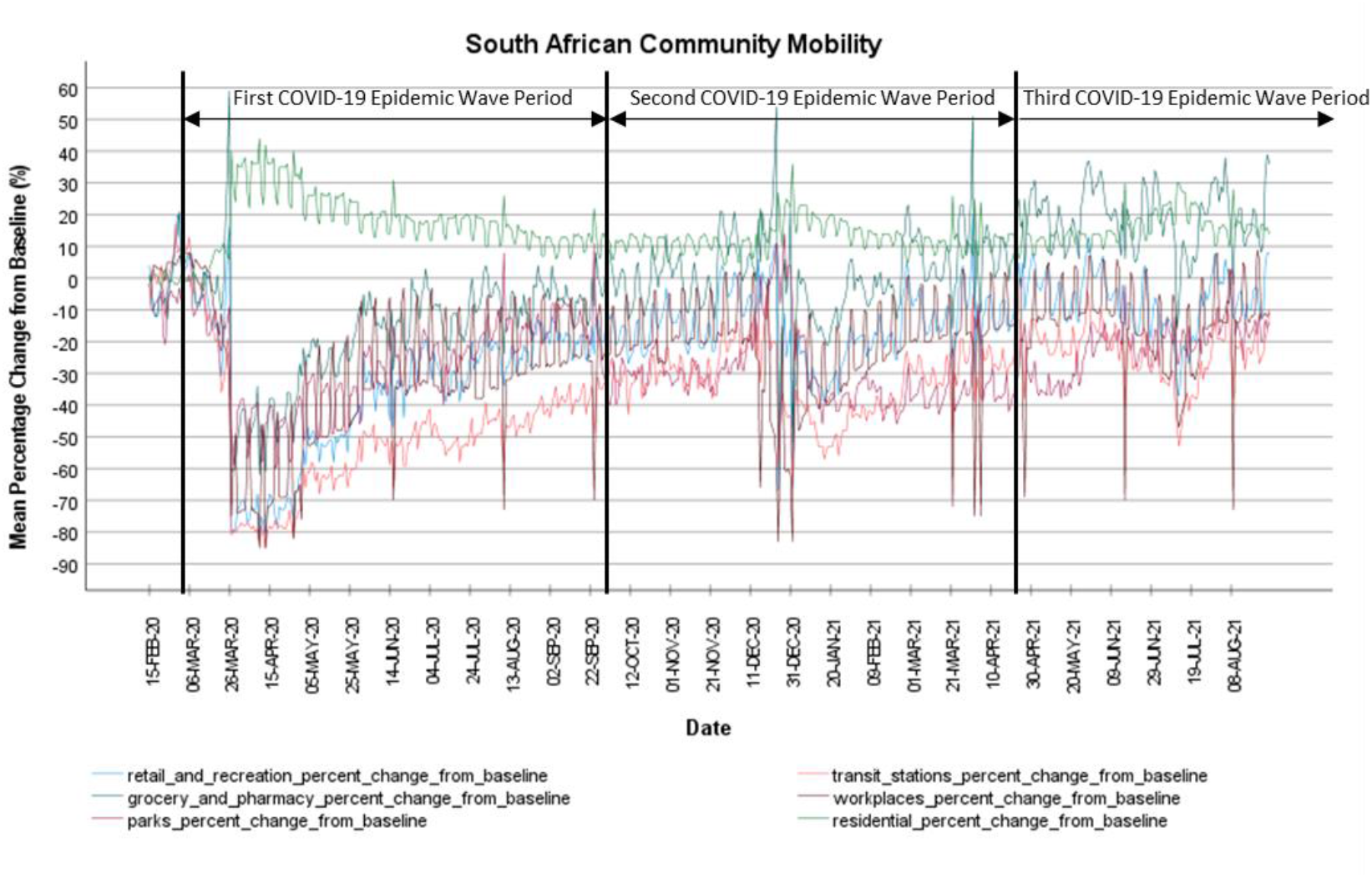
South African Community Google Mobility in Retail and Recreation, Grocery and Pharmacy, Parks, Transit Stations, Workplaces and Residences during the period of 2020/02/15 to 2021/08/27 [66]

### 2.3 Normalisation and Inferential Statistical Analysis of South African COVID-19 Epidemiological Data

#### 2.3.1 Comparative Analysis on the COVID-19 Transmissibility in the South African COVID-19 Epidemic Wave Periods

Disease transmissibility is the ability for a disease to be transmitted from one individual to another through infection. The basic reproductive number (R0) is a parameter that is largely used to describe the transmissibility of a disease. The basic reproductive number is the number of secondary infections that a primary infected person would produce in a completely susceptible population [61]. If R0 is greater than one then the disease is in an endemic equilibrium which results in positive exponential growth in cases. If the R0 is less than 1 then the disease is in the disease-free equilibrium which results in negative exponential growth in cases. The peak of the COVID-19 epidemic waves represents the turning point of the effective reproductive number. [58] showed through stochastic COVID-19 epidemiological modelling that the effective reproductive number in the first COVID-19 epidemic wave in South Africa was between 1.98 to 0.40. The NICD conducted a modelling analysis to estimate the initial basic reproductive number in South Africa using the method in [67] and estimated that it was 1.29 (95%CI: 1.9601.58)) during the National Lockdown Alert Level 5 rising to 1.5 by end of April 2020, 1.05 (95%CI:1.01-1.09) during the National Lockdown Alert Level 3 between 1 June and 1 August 2020 [68].

In this study, the COVID-19 transmissibility was measured through the magnitude of mean and variance of the COVID-19 Daily and Active Cases. Considering the linear positive correlation between the COVID-19 Daily Tests and Cases, for the comparative inferential analysis of COVID-19 Daily and Active Cases between the COVID-19 epidemic wave periods a normalised parameter (COVID-19 Daily Positive Tests) was developed based on Equation 8 to normalise the variance of testing in reported cases.

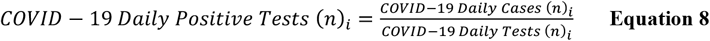

Where n is the number of cases/tests and i is the reported case date. For the comparative inferential statistical analysis conducted to understand the impact of the SARS-CoV-2 lineage clusters in Table 1 on the COVID-19 transmissibility during the three COVID-19 epidemic wave periods in South Africa the following analysis was conducted: 1) Descriptive Statistical Analysis on the COVID-19 Daily, Active Cases, Daily Positive Tests. 2) Paired Samples T-test at 95 % Confidence Intervals in the COVID-19 Daily Positive Tests between the three COVID-19 epidemic wave periods using the IBM SPSS STATISTICS 27 Software with the following test pairings:

- COVID-19 Daily Positive Tests_1 with COVID-19 Daily Positive Tests_2.
- COVID-19 Daily Positive Tests_1 with COVID-19 Daily Positive Tests_3.
- COVID-19 Daily Positive Tests_2 with COVID-19 Daily Positive Tests_3.

#### 2.3.2 Comparative Analysis on the COVID-19 Severity in the South African COVID-19 Epidemic Wave Periods

Disease severity is the measure of the relative proportions of symptoms and deaths due to the disease. SARS-COV-2 infections cause various symptoms such as coughing/sore throat, fever, myalgia or fatigue, respiratory symptoms, pneumonia. In moderate disease, pneumonia is reported and becomes severe in severe cases. In critical cases, it can cause acute respiratory distress syndrome (ARDS), dyspnoea, respiratory failure, sepsis, septic shock, acute thrombosis and multiple organ failure [58,69–71]. Patients with severe and critical COVID-19 are most likely to be hospitalised while those with moderate COVID-19 may present to an emergency unit or primary care/outpatient department [71]. Thus, the COVID-19 hospital admission status (indicative of care provided for COVID-19 inpatients), Admission and Deaths Age Profile (indicative of the severity by age), CFR (indicative of COVID-19 deaths in hospital) and the ECDR parameter (indicative of COVID-19 deaths outside of hospitals) can help one understand the severity of COVID-19. In this study, the COVID-19 severity was therefore measured through the magnitude of the mean and variance of the COVID-19 Hospital Admitted Cases, Admission Status, Admission Age Profile, Deaths Age Profile, CFR, DR and ECDR. Considering the linear positive correlation between the COVID-19 Active and Hospital Admitted Cases, for the comparative inferential analysis of COVID-19 Hospital Admitted Cases between the COVID-19 epidemic wave periods a normalised parameter (COVID-19 Hospital to Active Cases) was developed based on Equation 9 to normalise the variance of active cases in hospital admitted cases.

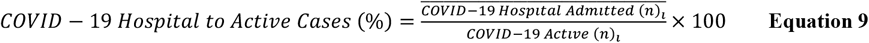

Where n is the number of patients and i is the reported case date. Considering the before mentioned variance as well as the positive correlation between the Number of Facilities Reporting to NICD DATCoV and COVID-19 Hospital Admitted Cases, for the comparative inferential analysis of COVID-19 Admission Status, Admission and Deaths Age Profile between the COVID-19 epidemic wave periods, normalised parameters were developed based on Equation 10, 11 and 12 to normalise the variance of active cases and Number of Facilities Reporting to the NICD DATCoV.

Where the COVID-19 Admitted Status (n)i is the number of COVID-19 patients in the General Ward, High Care, Intensive Care Unit, Isolation Ward, On Oxygen and Ventilators respectively.

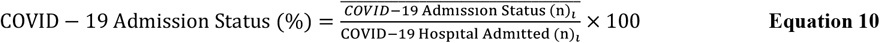

Where the COVID-19 Admitted Age Profile (n)i is the number of COVID-19 admitted patients in the ages of 0 to 9, 10 to 19, 20 to 29, 30 to 39, 40 to 49, 50 to 59, 60 to 69, 70 to 79 and 80 to 89 years respectively.

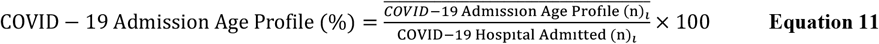

Where the COVID-19 Hospital Death Age Profile (n)i is the number of COVID-19 hospitalised patient deaths in the ages of 0 to 9, 10 to 19, 20 to 29, 30 to 39, 40 to 49, 50 to 59, 60 to 69, 70 to 79 and 80 to 89 years respectively.

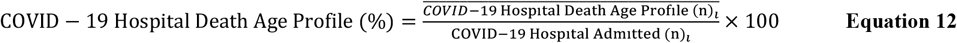

Cumulative COVID-19 Age Death Risk Ratios or Case Fatality Age Risk Ratio (CFARR) in the three COVID-19 epidemic wave periods for COVID-19 hospitalised patients deaths were then developed using Equation 13 referenced to the 0 to 9 age group.

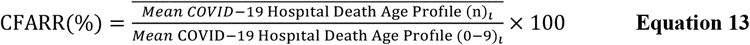

For the comparative inferential statistical analysis conducted to understand the impact of the SARS-CoV-2 lineage clusters in Table 1 on the COVID-19 severity during the three COVID-19 epidemic wave periods in South Africa the following analysis was conducted: 1) Descriptive Statistical Analysis 2) Paired Samples T-test at 95 % Confidence Intervals in the COVID-19 Hospital to Active Cases, Admission Status, Admission Age Profile, Hospital Death Age Profile (%), CFR, DR and ECDR between the three COVID-19 epidemic wave periods using the IBM SPSS STATISTICS 27 Software. The test was conducted with the same test pairings regarding the COVID-19 epidemic wave periods as described in Section 2.3.1.

## 3. Results and Discussions

### 3.1 Impact of SARS-CoV-2 Lineages on the COVID-19 Transmissibility in South Africa

Table 8 shows the descriptive statistics for the COVID-19 active, daily cases and daily positive tests for the first, second and third COVID-19 epidemic wave periods in South Africa. Table 8 shows that the mean COVID-19 daily active cases in South Africa’s first, second and third COVID-19 epidemic wave period were 51 747±54 618, 66 178±53 878 and 112 236±60 814 respectively. The maximum COVID-19 daily active cases were 173 590, 239 799 and 211 052 respectively while the COVID-19 daily cases were 3 211±3 882, 4 336±5 034 and 8 947±5 845 respectively. The mean COVID-19 active cases and daily cases in the third COVID-19 epidemic period in South Africa were observed to be higher than those of the first and second COVID-19 epidemic wave period.

**Table 8:** Statistical Sample Number (N), Minimum, Maximum, Mean, Standard Deviation (Std. Deviation) of COVID-19 Active Cases, Daily Cases and Positive Tests in the First, Second and Third COVID-19 Epidemic Wave Period in South Africa **(Descriptive Statistics**)

| Descriptive Statistics |  |  |  |  |  |  |  |  |  |
| --- | --- | --- | --- | --- | --- | --- | --- | --- | --- |
|  | COVID-19 Active Cases |  |  | COVID-19 Daily Cases |  |  | COVID-19 Daily Positive Tests |  |  |
| COVID-19 Epidemic Wave Period | 1 | 2 | 3 | 1 | 2 | 3 | 1 | 2 | 3 |
| N | 210 | 208 | 146 | 210 | 208 | 146 | 200 | 201 | 138 |
| Minimum | 1 | 19809 | -76869 | 0 | 437 | 0 | 0.00 | 2.71 | 0.00 |
| Maximum | 173590 | 239799 | 211052 | 13944 | 21980 | 26485 | 34.0 | 33.7 | 63.8 |
| Mean | 51747 | 66178 | 112236 | 3211 | 4336 | 8947 | 11.6 | 11.5 | 17.8 |
| Std. Deviation | 54618 | 53878 | 60814 | 3882 | 5034 | 5845 | 8.52 | 8.45 | 8.91 |

The COVID-19 daily positive tests indicate the transmissibility of COVID-19 based on the detection rate of COVID-19 and account for covariance in the testing rate for statistical comparison between epidemic wave periods. The mean COVID-19 daily positive tests in South Africa’s first, second and third COVID-19 epidemic wave period were 11.6 ±8.52 %, 11.5±8.45 % and 17.8±8.91 % respectively. The mean COVID-19 daily positive tests in South Africa’s first and second COVID-19 epidemic wave had a difference of 0.86 %. This difference was relatively small compared to the difference between the mean COVID-19 daily active and cases in the same periods. The relative difference of the mean COVID-19 daily positive tests, active and cases shows the need to account for the covariance of COVID-19 testing in COVID-19 daily active and cases to improve the accuracy of the respective parameters. The relatively small difference between the mean COVID-19 daily positive tests in South Africa’s first and second COVID-19 epidemic wave suggests that the SARS-CoV-2 lineage cluster 1 (predominantly B.1.1.54, B.1.1.56 C.1 SARS-CoV-2 lineages) detected in the first COVID-19 epidemic wave period resulted in similar COVID-19 transmissibility in the South Africa population when compared with the SARS-CoV-2 lineage cluster 2 (predominantly Beta (B.1.351) SARS-CoV-2 VOC) detected in the second COVID-19 epidemic wave. The mean COVID-19 daily positive tests in South Africa’s third COVID-19 epidemic wave were 53.9 and 54.8 % more than those of the first and second COVID-19 epidemic wave period respectively. This result suggests that the SARS-CoV-2 lineage cluster 3 (predominantly Delta (B.1.617.2) SARS-CoV-2 VOC) detected in the third COVID-19 epidemic wave resulted in higher COVID-19 transmissibility in the South African population than the SARS-CoV-2 lineage clusters 1 and 2. The difference between the mean COVID-19 daily positive tests in South Africa’s first, second and third COVID-19 epidemic wave period can also be observed in Figure 2. A paired T-test of the COVID-19 daily positive tests between the first and second COVID-19 epidemic wave period showed no significant difference at 95 % confidence interval between these COVID-19 epidemic periods with a p-value of 0.709 (shown in Table 9). While the paired T-test of the COVID-19 daily positive tests between the Pair 2 (first and third COVID-19 epidemic wave period) and Pair 3 (second and third COVID-19 epidemic wave period) showed significant difference at 95 % confidence interval between the respective COVID-19 epidemic periods with p-values of 1.82×10^−11^ and 5.87×10^−05^ respectively (shown in Table 9).

**Table 9:** Mean Paired Differences, Standard Deviation (Std.) Paired Differences, Standard Error of Mean (Std. Error Mean), 95 % Confidence Interval (CI) if the Upper and Lower Difference, t-value, degrees of freedom (df) and P-value (Sig. (2-tailed) for the COVID-19 Daily Positive Tests between Pair 1 (1^st^ and 2^nd^ COVID-19 Epidemic Wave Period), Pair 2 (1^st^ and 3^rd^ COVID-19 Epidemic Wave Period) and Pair 3 (2^nd^ and 3^rd^ COVID-19 Epidemic Wave Period) in South Africa. **(Paired Samples T-Test**)

| Paired Samples Test |  |  |  |
| --- | --- | --- | --- |
|  | Pair 1 | Pair 2 | Pair 3 |
|  | COVID-19 Daily Positive Tests_1-COVID-19 Daily Positive Tests_2 | COVID-19 Daily Positive Tests_1-COVID-19 Daily Positive Tests_3 | COVID-19 Daily Positive Tests_2-COVID-19 Daily Positive Tests_3 |
| Mean Paired Differences | 0.36 | -8.68 | -2.54 |
| Std. Deviation Paired Differences | 13.2 | 13.3 | 6.99 |
| Std. Error Mean | 0.96 | 1.18 | 0.61 |
| 95% CI of the Difference Lower | -1.53 | -11.00 | -3.75 |
| 95% CI of the Difference Upper | 2.25 | -6.35 | -1.33 |
| t | 0.37 | -7.38 | -4.15 |
| df | 190 | 127 | 130 |
| Sig. (2-tailed) | 0.709 | 1.82E-11 | 5.87E-05 |

**Figure 2:**
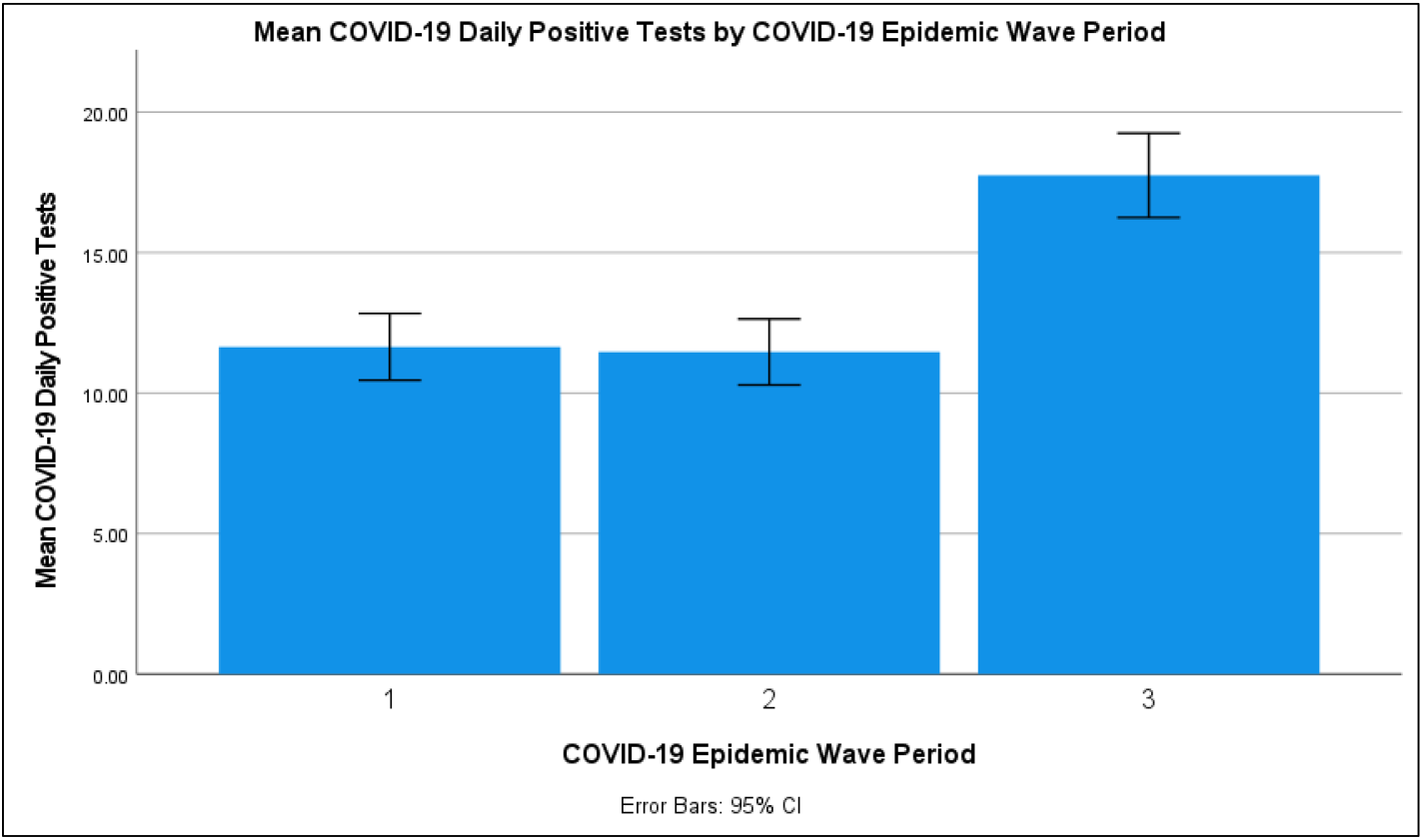
The mean COVID-19 Daily Positive Tests (with 95 % Confidence Interval (CI) Error Bars) in the First, Second and Third COVID-19 Epidemic Wave Period in South Africa

### 3.2 Impact of SARS-CoV-2 Linages on the COVID-19 Severity in South Africa

#### 3.2.1 Impact of SARS-CoV-2 Lineages on the COVID-19 Hospital Admissions

The COVID-19 hospital to active cases indicates the severity of COVID-19 based on the relative amount of COVID-19 hospitalised cases to the active cases and accounts for the covariance of active cases in hospitalised cases. According to WHO, patients with severe and critical COVID-19 are most likely to be hospitalised while those with moderate COVID-19 may present to an emergency unit or primary care/outpatient department [71]. Table 10 shows the descriptive statistics for the COVID-19 hospital to active cases for the first, second and third COVID-19 epidemic wave periods in South Africa. Table 10 shows that the mean COVID-19 daily hospital to active cases in South Africa’s first, second and third COVID-19 epidemic wave period were 6.8±1.82 %, 14.5±4.68 % and 10.6±2.81 % respectively. The second COVID-19 epidemic wave period in South Africa had the highest COVID-19 hospital to active cases followed by the third COVID-19 epidemic wave period. The difference of the mean COVID-19 hospital to active cases between Pair 1 (COVID-19 epidemic wave period 1 and 2), Pair 2 (COVID-19 epidemic wave period 1 and 3), Pair 3 (COVID-19 epidemic wave period 2 and 3) was 113 %, 55.8 % and 36.8 % respectively. The difference between the mean COVID-19 hospital to active cases in South Africa’s first, second and third COVID-19 epidemic wave period can also be observed in Figure 3. Paired T-tests of the COVID-19 hospital to active cases between the Pair 1, Pair 2 and Pair 3 showed significant difference at 95 % confidence interval between the respective COVID-19 epidemic periods with p-values of 4.25×10^−21^, 9.10×10^−44^ and 3.90×10^−06^ respectively (shown in Table 11).

**Table 10:** Statistical Sample Number (N), Minimum, Maximum, Mean, Standard Deviation (Std. Deviation) of COVID-19 Hospital to Active Cases in the First, Second and Third COVID-19 Epidemic Wave Period in South Africa **(Descriptive Statistics**)

| Descriptive Statistics |  |  |  |
| --- | --- | --- | --- |
|  | COVID-19 Hospital To Active Cases (%) |  |  |
| COVID-19 Epidemic Wave Period | 1 | 2 | 3 |
| N | 126 | 187 | 145 |
| Minimum | 4.0 | 6.5 | 7.2 |
| Maximum | 12.7 | 23.5 | 18.3 |
| Mean | 6.8 | 14.5 | 10.6 |
| Std. Deviation | 1.82 | 4.68 | 2.81 |

**Table 11:** Mean Paired Differences, Standard Deviation (Std.) Paired Differences, Standard Error of Mean (Std. Error Mean), 95 % Confidence Interval (CI) if the Upper and Lower Difference, t-value, degrees of freedom (df) and P-value (Sig. (2-tailed) for the COVID-19 Hospital to Active Cases between Pair 1 (1^st^ and 2^nd^ COVID-19 Epidemic Wave Period), Pair 2 (1^st^ and 3^rd^ COVID-19 Epidemic Wave Period) and Pair 3 (2^nd^ and 3^rd^ COVID-19 Epidemic Wave Period) in South Africa. **(Paired Samples T-Test**)

| Paired Samples Test |  |  |  |
| --- | --- | --- | --- |
|  | COVID-19 Hospital To Active Cases(%) |  |  |
| Test Pairing | Pair 1 | Pair 2 | Pair 3 |
| Mean Paired Differences | -4.608 | -3.853 | 2.173 |
| Std. Deviation Paired Differences | 4.000 | 1.998 | 5.005 |
| Std. Error Mean | 0.389 | 0.179 | 0.449 |
| 95% CI of the Difference Lower | -5.379 | -4.207 | 1.283 |
| 95% CI of the Difference Upper | -3.838 | -3.499 | 3.063 |
| t | -11.86 | -21.56 | 4.83 |
| df | 105 | 124 | 123 |
| Sig. (2-tailed) | 4.25E-21 | 9.10E-44 | 3.90E-06 |

**Figure 3:**
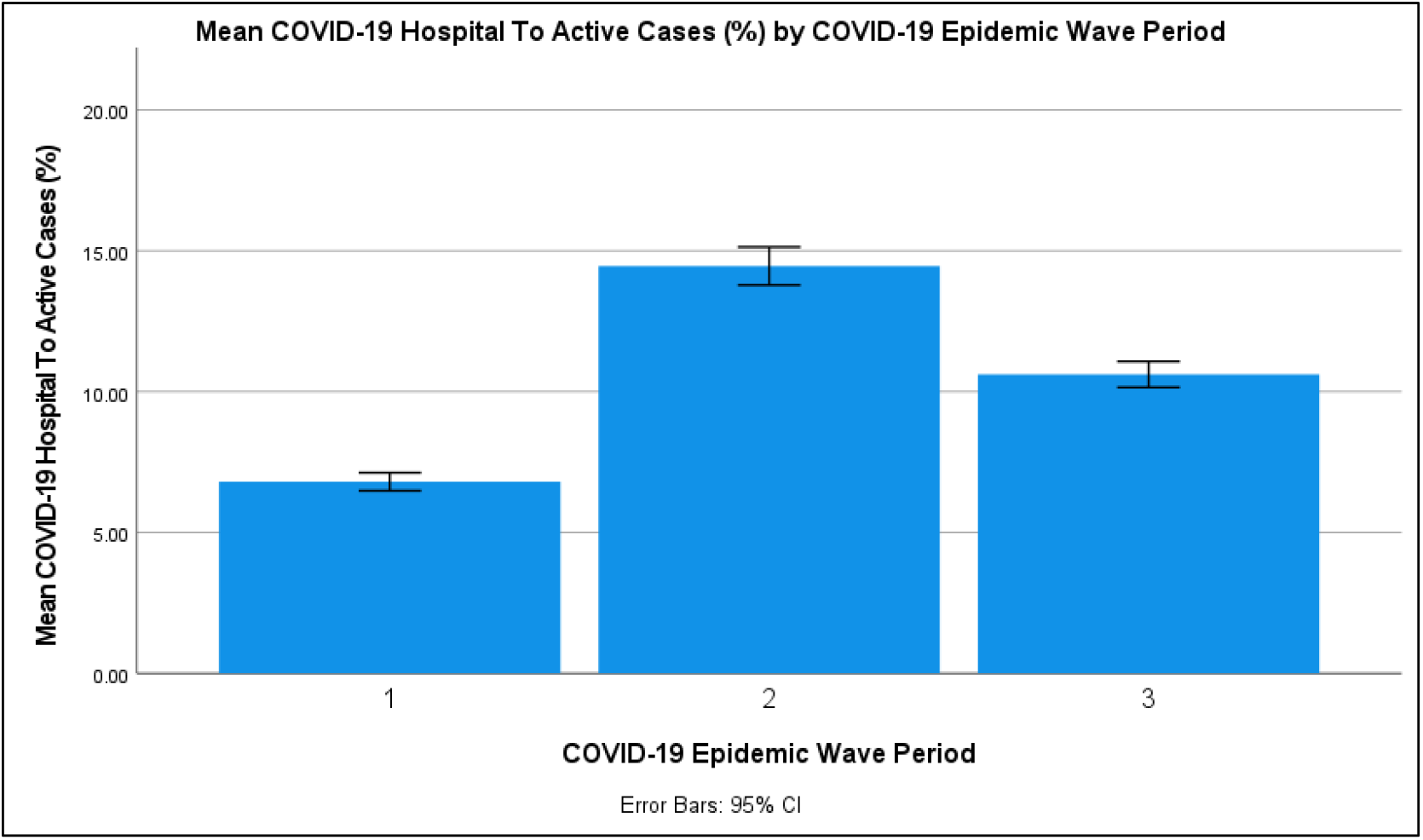
The mean COVID-19 Hospital to Active Cases (with 95 % Confidence Interval (CI) Error Bars) in the First, Second and Third COVID-19 Epidemic Wave Period in South Africa

The results in Section 3.2.1 suggests that the SARS-CoV lineage cluster 2 (predominantly Beta (B.1.351) SARS-CoV-2 VOC) resulted in an increase in severity of COVID-19 in the second COVID-19 epidemic wave period resulting in higher COVID-19 daily hospitalisation per active cases compared to the SARS-CoV-2 lineage clusters 1 and 3 in the first and third COVID-19 epidemic wave period in South Africa respectively. While the SARS-CoV lineage cluster 3 (predominantly Delta (B.1.617.2) SARS-CoV-2 VOC) resulted in an increase in severity of COVID-19 resulting in higher daily COVID-19 hospitalisation per active cases compared to the SARS-CoV-2 lineage cluster 1.

#### 3.2.2 Impact of SARS-CoV-2 Lineages on the COVID-19 Hospital Admission Status

Table 12 shows the descriptive statistics for the COVID-19 hospital admission status for the first, second and third COVID-19 epidemic wave periods in South Africa. Table 12 shows that the mean COVID-19 hospitalised cases in the first COVID-19 epidemic wave period in the general ward, high care, intensive care unit, on oxygen and ventilator were 72.8±2.6 %, 7.9±1.0 %, 16.7±2.2 %, 16.8±5.8 % and 7.8±1.8 % respectively. The mean COVID-19 hospitalised cases in the second COVID-19 epidemic wave period in the general ward, high care, intensive care unit, on oxygen and ventilator were 78.7±3.2 %, 7.3±1.0 %, 14.1±2.3 %, 27.6±4.5 % and 6.9±1.8 % respectively. While the mean COVID-19 hospitalised cases in the third COVID-19 epidemic wave period in the general ward, high care, intensive care unit, on oxygen and ventilator were 76.0±2.5 %, 7.6±1.1 %, 16.3±1.7 %, 21.9±4.8 % and 8.7±0.6 % respectively. Figure 4 shows the COVID-19 hospital admission status profile in the first, second and third COVID-19 epidemic wave periods in South Africa.

**Table 12:** Statistical Sample Number (N), Minimum, Maximum, Mean, Standard Deviation (Std. Deviation) of COVID-19 Hospitalised Cases in the General Ward, High Care, Intensive Care Unit, On Oxygen, On Ventilator in the First, Second and Third COVID-19 Epidemic Wave Period in South Africa **(Descriptive Statistics**)

| Descriptive Statistics |  |  |  |  |  |  |  |  |  |  |  |  |  |  |  |
| --- | --- | --- | --- | --- | --- | --- | --- | --- | --- | --- | --- | --- | --- | --- | --- |
|  | COVID-19 Hospitalised General Ward (%) |  |  | COVID-19 Hospitalised High Care (%) |  |  | COVID-19 Hospitalised Intensive Care Unit (%) |  |  | COVID-19 Hospitalised On Oxygen (%) |  |  | COVID-19 Hospitalised On Ventilator (%) |  |  |
| COVID-19 Epidemic Wave Period | 1 | 2 | 3 | 1 | 2 | 3 | 1 | 2 | 3 | 1 | 2 | 3 | 1 | 2 | 3 |
| N | 125 | 187 | 146 | 125 | 187 | 146 | 125 | 187 | 146 | 125 | 187 | 146 | 125 | 187 | 146 |
| Minimum | 68.7 | 73.4 | 71.5 | 5.1 | 5.3 | 6.1 | 11.9 | 8.5 | 12.9 | 10.6 | 18.7 | 17.2 | 4.9 | 4.3 | 7.2 |
| Maximum | 78.1 | 86.2 | 80.7 | 10.5 | 9.4 | 10.1 | 20.6 | 17.6 | 19.5 | 30.8 | 38.6 | 34.2 | 10.8 | 9.4 | 10.4 |
| Mean | 72.8 | 78.7 | 76.0 | 7.9 | 7.3 | 7.6 | 16.7 | 14.1 | 16.3 | 16.8 | 27.6 | 21.9 | 7.8 | 6.9 | 8.7 |
| Std. Deviation | 2.6 | 3.2 | 2.5 | 1.0 | 1.0 | 1.1 | 2.2 | 2.3 | 1.7 | 5.8 | 4.5 | 4.8 | 1.8 | 1.8 | 0.6 |

**Figure 4:**
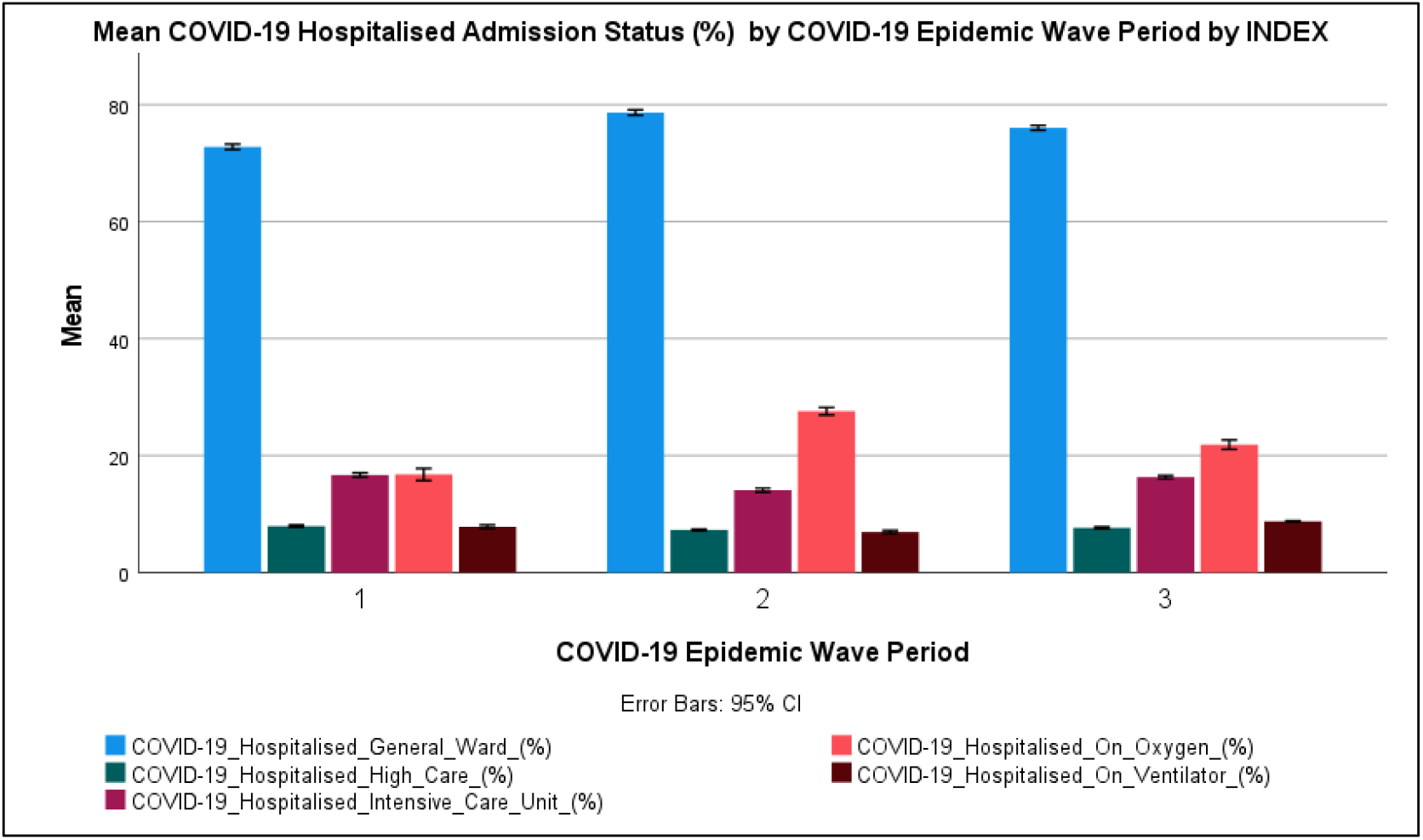
The mean COVID-19 Hospitalised Admission Status (with 95 % Confidence Interval (CI) Error Bars) in the First, Second and Third COVID-19 Epidemic Wave Period in South Africa

Figure 4 shows that most COVID-19 hospitalised cases in South Africa were hospitalised in the general ward (72.8-78.7 %). Figure 4 also shows that the COVID-19 patients on Oxygen were the second-largest admission status (16.8-27.6 %) followed by the COVID-19 patients in the intensive care unit (14.1-16.7 %). The COVID-19 patients on oxygen in the second and third COVID-19 epidemic wave period were higher than those observed in the first COVID-19 epidemic wave period with those in the second COVID-19 epidemic wave period being the highest. Paired T-tests of the mean COVID-19 hospital admission status in the Pair 1, Pair 2 and Pair 3 showed significant difference at 95 % confidence interval between the respective COVID-19 epidemic periods with p-values in the range of 2.86×10^−41^ to 0.00219 respectively (shown in Table 13). Table 13 shows that the COVID-19 patients admitted in the intensive care unit in the first and third COVID-19 epidemic wave period were not significantly different at a 95 % confidence interval with a p-value of 0.514.

**Table 13:**
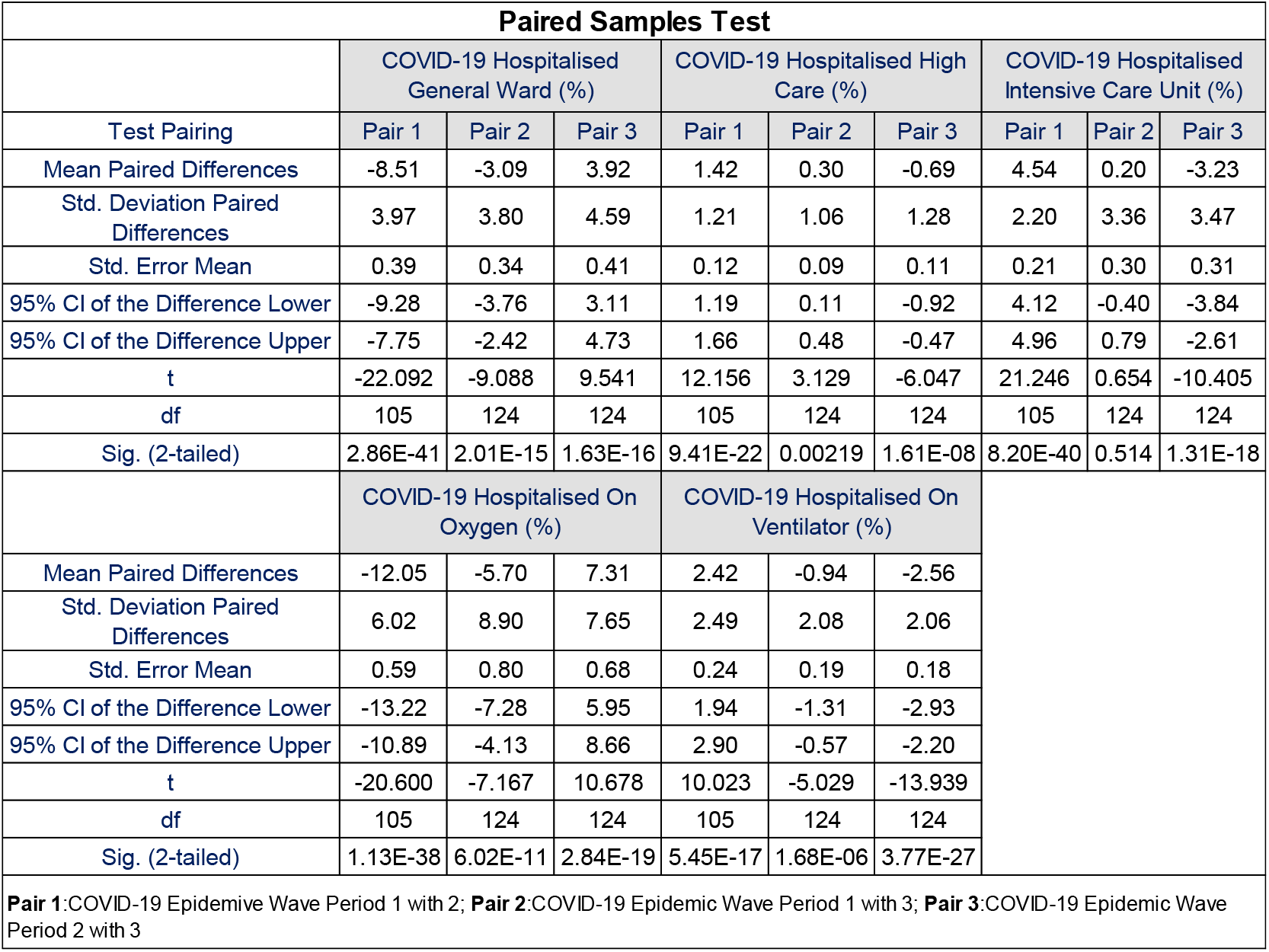
Mean Paired Differences, Standard Deviation (Std.) Paired Differences, Standard Error of Mean (Std. Error Mean), 95 % Confidence Interval (CI) if the Upper and Lower Difference, t-value, degrees of freedom (df) and P-value (Sig. (2-tailed) for the COVID-19 Hospitalised Cases in the General Ward, High Care, Intensive Care Unit, On Oxygen, On Ventilator between Pair 1 (1^st^ and 2^nd^ COVID-19 Epidemic Wave Period), Pair 2 (1^st^ and 3^rd^ COVID-19 Epidemic Wave Period) and Pair 3 (2^nd^ and 3^rd^ COVID-19 Epidemic Wave Period) in South Africa. **(Paired Samples T-Test**)

The results in Section 3.2.2 suggest that the SARS-CoV-2 lineage clusters observed in the first, second and third COVID-19 epidemic wave periods in South Africa resulted in a similar distribution of the COVID-19 hospital admission status profile however the mean COVID-19 admission status was significantly different at 95 % confidence interval. The results also show that the SARS-CoV-2 lineage cluster 1 (predominantly B.1.1.54, B.1.1.56 C.1 SARS-CoV-2 lineages) and SARS-CoV-2 lineage cluster 3 (predominantly Delta (B.1.617.2) SARS-CoV-2 VOC) resulted in the same COVID-19 patients admitted in the intensive care unit relative to the total COVID-19 hospital admitted cases at 95 % confidence interval. The SARS-CoV-2 lineage cluster 2 (predominantly Beta (B.1.351) SARS-CoV-2 VOC) was observed to have resulted in higher COVID-19 patients on oxygen than the SARS-CoV-2 lineage clusters 1 and 3.

#### 3.2.3 Impact of SARS-CoV-2 Lineages on the COVID-19 Hospital Admission Age Profile

Table 14 shows the descriptive statistics for the COVID-19 hospital admission age profile for the first, second and third COVID-19 epidemic wave periods in South Africa. Table 14Table 12 shows that the mean COVID-19 hospitalised cases in the first COVID-19 epidemic wave period in the ages of 0 to 9, 10 to 19, 20 to 29, 30 to 39, 40 to 49, 50 to 59, 60 to 69, 70 to 79 and 80 to 89 years were 2.4±0.6 %, 1.8±0.3 %, 7.5±2.6 %, 17.4±4.4 %, 19.5±4.9 %, 24.0±6.1 %, 16.2±5.4 %, 7.1±4.4 % and 5.6±1.6 % respectively. The mean COVID-19 hospitalised cases in the second COVID-19 epidemic wave period in the ages of 0 to 9, 10 to 19, 20 to 29, 30 to 39, 40 to 49, 50 to 59, 60 to 69, 70 to 79 and 80 to 89 years were 1.9±0.3 %, 4.4±2.2 %, 7.2±2.0 %, 14.2±1.7 %, 17.0±1.2 %, 19.6±4.8 %, 18.2±1.8 %, 9.8±3.2 % and 5.0±0.6 % respectively. While, the mean COVID-19 hospitalised cases in the third COVID-19 epidemic wave period in the ages of 0 to 9, 10 to 19, 20 to 29, 30 to 39, 40 to 49, 50 to 59, 60 to 69, 70 to 79 and 80 to 89 years were 2.3±0.5 %, 2.3±0.5 %, 6.4±7.4 %, 9.4±3.8 %, 13.5±3.3 %, 12.9±9.1 %, 15.5±7.4 %, 12.8±3.2 % and 6.5±1.7 % respectively. Figure 5 shows the COVID-19 hospital admission age profile in the first, second and third COVID-19 epidemic wave periods in South Africa.

**Table 14:**
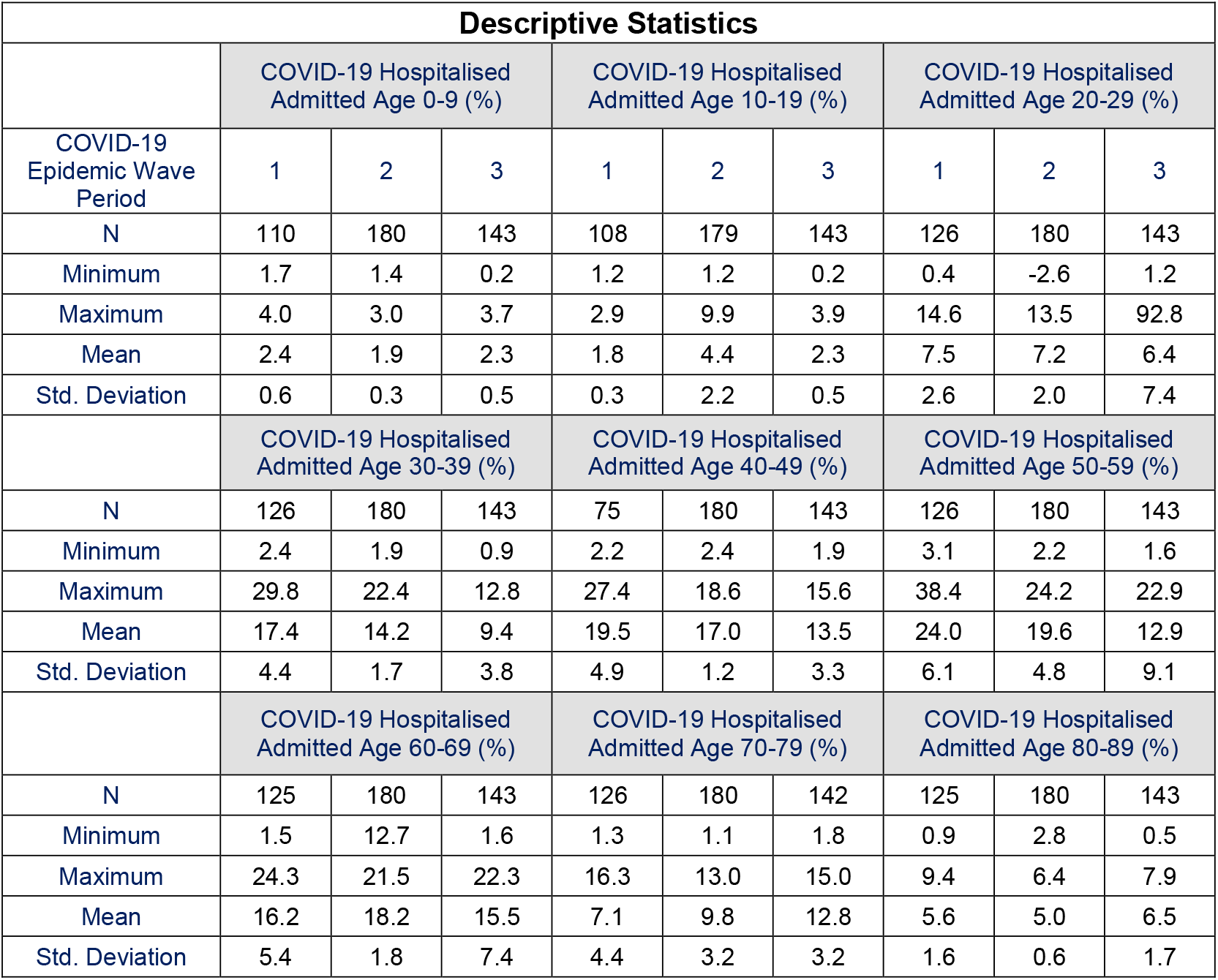
Statistical Sample Number (N), Minimum, Maximum, Mean, Standard Deviation (Std. Deviation) of COVID-19 Hospitalised Cases in the Ages of 0 to 9, 10 to 19, 20 to 29, 30 to 39, 40 to 49, 50 to 59, 60 to 69, 70 to 79, 80 to 89 years in the First, Second and Third COVID-19 Epidemic Wave Period in South Africa **(Descriptive Statistics**)

| Descriptive Statistics |  |  |  |  |  |  |  |  |  |
| --- | --- | --- | --- | --- | --- | --- | --- | --- | --- |
|  | COVID-19 Hospitalised Admitted Age 0-9 (%) |  |  | COVID-19 Hospitalised Admitted Age 10-19 (%) |  |  | COVID-19 Hospitalised Admitted Age 20-29 (%) |  |  |
| COVID-19 Epidemic Wave Period | 1 | 2 | 3 | 1 | 2 | 3 | 1 | 2 | 3 |
| N | 110 | 180 | 143 | 108 | 179 | 143 | 126 | 180 | 143 |
| Minimum | 1.7 | 1.4 | 0.2 | 1.2 | 1.2 | 0.2 | 0.4 | -2.6 | 1.2 |
| Maximum | 4.0 | 3.0 | 3.7 | 2.9 | 9.9 | 3.9 | 14.6 | 13.5 | 92.8 |
| Mean | 2.4 | 1.9 | 2.3 | 1.8 | 4.4 | 2.3 | 7.5 | 7.2 | 6.4 |
| Std. Deviation | 0.6 | 0.3 | 0.5 | 0.3 | 2.2 | 0.5 | 2.6 | 2.0 | 7.4 |
|  | COVID-19 Hospitalised Admitted Age 30-39 (%) |  |  | COVID-19 Hospitalised Admitted Age 40-49 (%) |  |  | COVID-19 Hospitalised Admitted Age 50-59 (%) |  |  |
| N | 126 | 180 | 143 | 75 | 180 | 143 | 126 | 180 | 143 |
| Minimum | 2.4 | 1.9 | 0.9 | 2.2 | 2.4 | 1.9 | 3.1 | 2.2 | 1.6 |
| Maximum | 29.8 | 22.4 | 12.8 | 27.4 | 18.6 | 15.6 | 38.4 | 24.2 | 22.9 |
| Mean | 17.4 | 14.2 | 9.4 | 19.5 | 17.0 | 13.5 | 24.0 | 19.6 | 12.9 |
| Std. Deviation | 4.4 | 1.7 | 3.8 | 4.9 | 1.2 | 3.3 | 6.1 | 4.8 | 9.1 |
|  | COVID-19 Hospitalised Admitted Age 60-69 (%) |  |  | COVID-19 Hospitalised Admitted Age 70-79 (%) |  |  | COVID-19 Hospitalised Admitted Age 80-89 (%) |  |  |
| N | 125 | 180 | 143 | 126 | 180 | 142 | 125 | 180 | 143 |
| Minimum | 1.5 | 12.7 | 1.6 | 1.3 | 1.1 | 1.8 | 0.9 | 2.8 | 0.5 |
| Maximum | 24.3 | 21.5 | 22.3 | 16.3 | 13.0 | 15.0 | 9.4 | 6.4 | 7.9 |
| Mean | 16.2 | 18.2 | 15.5 | 7.1 | 9.8 | 12.8 | 5.6 | 5.0 | 6.5 |
| Std. Deviation | 5.4 | 1.8 | 7.4 | 4.4 | 3.2 | 3.2 | 1.6 | 0.6 | 1.7 |

**Figure 5:**
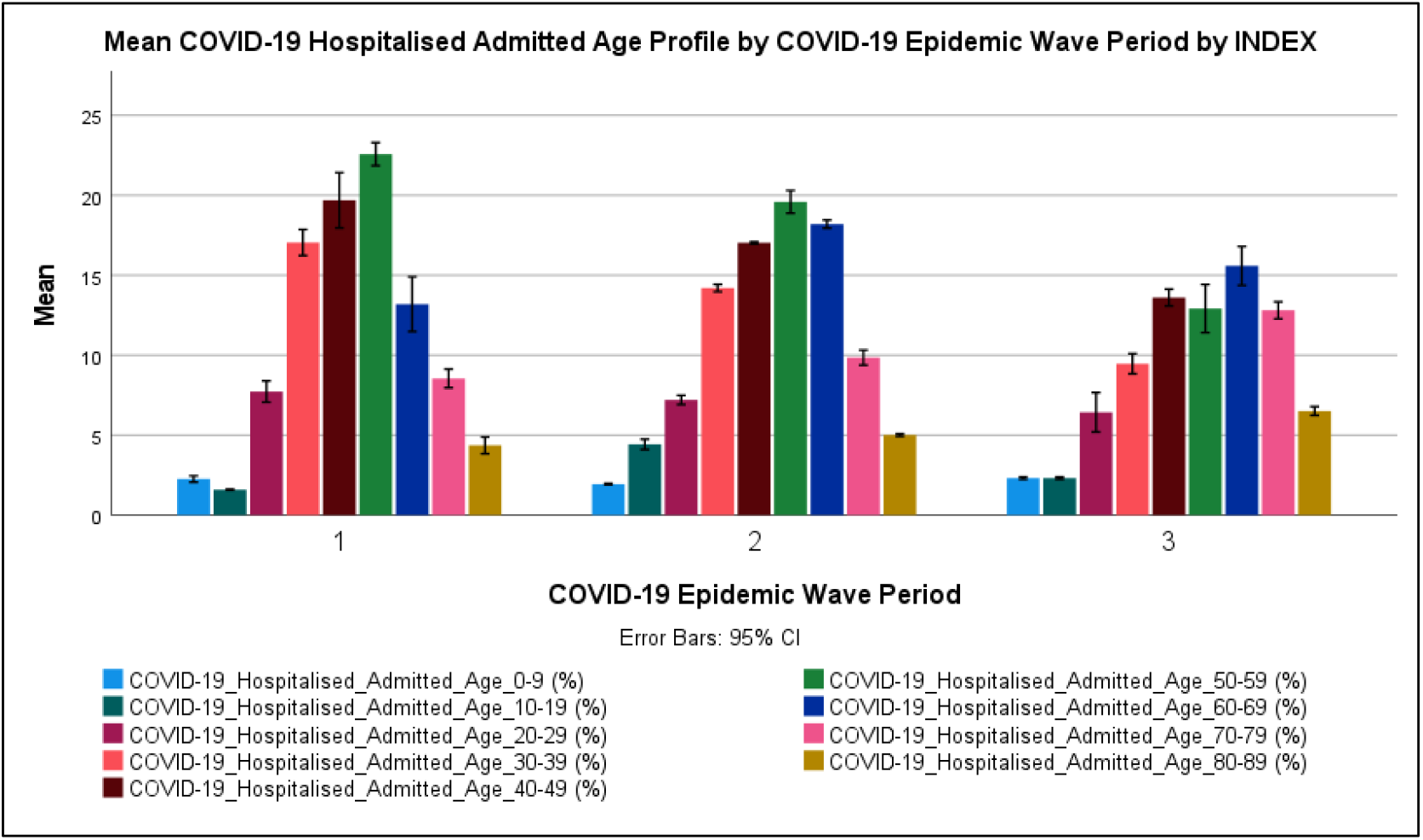
The mean COVID-19 Hospitalised Admission Age Profile (with 95 % Confidence Interval (CI) Error Bars) in the First, Second and Third COVID-19 Epidemic Wave Period in South Africa

Figure 5 shows that most COVID-19 hospitalised cases in South Africa’s first, second and third COVID-19 epidemic wave period were in the ages of 50 to59 (19.6 %), 50 to 59 (24.0 %) and 60 to 69 (15.5 %) years respectively. COVID-19 patients in the age groups of 40 to 49 were the second-largest admitted age groups in the first and third COVID-19 epidemic wave period (14.1-16.7 %) while for the second COVID-19 epidemic wave period it was the age group of 60 to 69 years (18.2 %). COVID-19 admitted patients in the age groups of 0 to 19 years were relatively low (1.8 to 4.4 %) and highest in the second COVID-19 epidemic wave period. The incidence of COVID-19 in children has been reported to be low around the world particularly severe and critical COVID-19 requiring hospitalisation [8]. Figure 5 shows that the COVID-19 hospital admitted age profile for the first and second COVID-19 epidemic wave period was relative normally distributed, while the distribution in the third COVID epidemic wave period was left-skewed with increased incidence in the age groups over 70 years.

In general, paired T-tests of the COVID-19 hospital admission age profile in Pair 1, Pair 2 and Pair 3 showed significant differences at 95 % confidence interval between the respective COVID-19 epidemic periods with p-values in the range of 7.66×10^−44^ to 0.040 respectively (shown in Table 15). However, Table 15 also shows that the COVID-19 patients admitted in age group 0 to 9 years in first and third COVID-19 epidemic wave periods, age group 20 to 29 years in second and third COVID-19 epidemic wave periods, age group 60 to 69 years in first and, second and third COVID-19 epidemic wave periods, age group 70 to 79 years in first and second COVID-19 epidemic wave periods were significant similar at 95 confidence interval with p-values of 0.111 to 0.530.

**Table 15:**
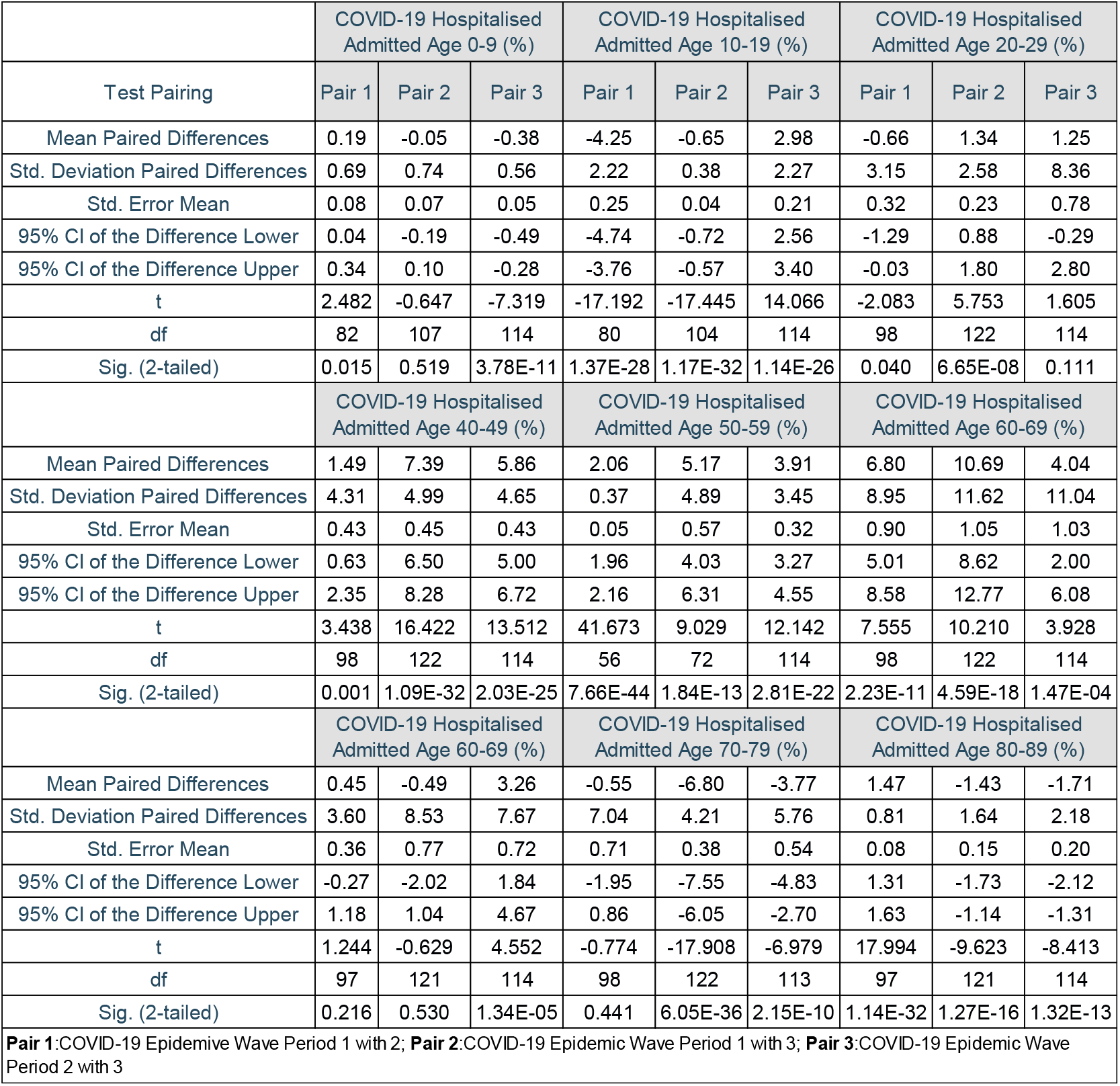
Mean Paired Differences, Standard Deviation (Std.) Paired Differences, Standard Error of Mean (Std. Error Mean), 95 % Confidence Interval (CI) if the Upper and Lower Difference, t-value, degrees of freedom (df) and P-value (Sig. (2-tailed) for the COVID-19 Hospitalised Cases in the Ages of 0 to 9, 10 to 19, 20 to 29, 30 to 39, 40 to 49, 50 to 59, 60 to 69, 70 to 79, 80 to 89 years between Pair 1 (1^st^ and 2^nd^ COVID-19 Epidemic Wave Period), Pair 2 (1^st^ and 3^rd^ COVID-19 Epidemic Wave Period) and Pair 3 (2^nd^ and 3^rd^ COVID-19 Epidemic Wave Period) in South Africa. **(Paired Samples T-Test**)

The results in Section 3.2.3 suggest that the SARS-CoV-2 lineage clusters observed in the first, second and third COVID-19 epidemic wave periods in South Africa resulted in different distributions of the COVID-19 hospital admission age profile. The results also show that the SARS-CoV-2 lineage cluster 3 (predominantly Delta (B.1.617.2) resulted in an increased incidence in COVID-19 patients in the age groups over 70 years compared to the SARS SARS-CoV-2 lineage cluster 1 (predominantly B.1.1.54, B.1.1.56 C.1 SARS-CoV-2 lineages) and the SARS-CoV-2 lineage cluster 2 (predominantly Beta (B.1.351) SARS-CoV-2 VOC).

#### 3.2.4 Impact of SARS-CoV-2 Lineages on the COVID-19 Hospital Deaths Age Profile

Table 16 shows the descriptive statistics for the COVID-19 hospital death age profile for the first, second and third COVID-19 epidemic wave periods in South Africa. Table 16Table 12 shows that the mean COVID-19 hospitalised deaths in the first COVID-19 epidemic wave period in the ages of 0 to 9, 10 to 19, 20 to 29, 30 to 39, 40 to 49, 50 to 59, 60 to 69, 70 to 79 and 80 to 89 years were 0.194±0.111 %, 0.254±0.089 %, 1.34±0.312 %, 5.20±0.62 %, 11.7±0.9 %, 14.8±12.0 %, 25.8±2.74 %, 18.1±2.18 % and 11.7±2.28 % respectively. The mean COVID-19 hospitalised deaths in the second COVID-19 epidemic wave period in the ages of 0 to 9, 10 to 19, 20 to 29, 30 to 39, 40 to 49, 50 to 59, 60 to 69, 70 to 79 and 80 to 89 years were 0.250±0.095 %, 0.398±0.132 %, 1.97±0.500 %, 5.67±0.5 %, 10.8±0.3 %, 41.2±20.7 %, 28.5±4.27 %, 20.0±3.12 % and 10.6±1.94 % respectively.

**Table 16:** Statistical Sample Number (N), Minimum, Maximum, Mean, Standard Deviation (Std. Deviation) of COVID-19 Hospitalised Deaths in the Ages of 0 to 9, 10 to 19, 20 to 29, 30 to 39, 40 to 49, 50 to 59, 60 to 69, 70 to 79, 80 to 89 years in the First, Second and Third COVID-19 Epidemic Wave Period in South Africa **(Descriptive Statistics**)

| Descriptive Statistics |  |  |  |  |  |  |  |  |  |
| --- | --- | --- | --- | --- | --- | --- | --- | --- | --- |
|  | COVID-19 Hospital Deaths Age 0-9 (%) |  |  | COVID-19 Hospital Deaths Age 10-19 (%) |  |  | COVID-19 Hospital Deaths Age 20-29 (%) |  |  |
| COVID-19 Epidemic Wave Period | 1 | 2 | 3 | 1 | 2 | 3 | 1 | 2 | 3 |
| N | 126 | 187 | 146 | 126 | 187 | 146 | 126 | 187 | 146 |
| Minimum | 0.00 | 0.00 | 0.00 | 0.00 | 0.00 | 0.00 | 0.00 | 0.00 | 0.00 |
| Maximum | 0.64 | 0.44 | 75.00 | 0.34 | 1.25 | 0.31 | 2.26 | 3.81 | 1.61 |
| Mean | 0.194 | 0.250 | 0.944 | 0.254 | 0.398 | 0.217 | 1.341 | 1.965 | 0.990 |
| Std. Deviation | 0.111 | 0.095 | 6.176 | 0.089 | 0.132 | 0.074 | 0.312 | 0.500 | 0.297 |
|  | COVID-19 Hospital Deaths Age 30-39 (%) |  |  | COVID-19 Hospital Deaths Age 40-49 (%) |  |  | COVID-19 Hospital Deaths Age 50-59 (%) |  |  |
| N | 126 | 187 | 146 | 126 | 187 | 146 | 125 | 162 | 146 |
| Minimum | 0.73 | 5.10 | 1.96 | 5.13 | 9.81 | 0.00 | 0.00 | 0.00 | 8.39 |
| Maximum | 6.33 | 7.27 | 25.00 | 13.32 | 11.63 | 10.38 | 27.91 | 98.20 | 25.00 |
| Mean | 5.20 | 5.67 | 4.60 | 11.7 | 10.8 | 7.8 | 14.8 | 41.2 | 18.1 |
| Std. Deviation | 0.62 | 0.5 | 2.0 | 0.9 | 0.3 | 2.0 | 12.0 | 20.7 | 4.0 |
|  | COVID-19 Hospital Deaths Age 60-69 (%) |  |  | COVID-19 Hospital Deaths Age 70-79 (%) |  |  | COVID-19 Hospital Deaths Age 80-89(%) |  |  |
| N | 126 | 187 | 146 | 126 | 187 | 146 | 126 | 187 | 146 |
| Minimum | 0.00 | 0.00 | 0.00 | 0.00 | 0.00 | 8.54 | 0.00 | 0.00 | 5.41 |
| Maximum | 28.86 | 31.19 | 36.97 | 19.83 | 21.31 | 50.00 | 13.96 | 23.75 | 25.00 |
| Mean | 25.82 | 28.54 | 25.58 | 18.11 | 20.01 | 21.28 | 11.74 | 10.61 | 13.40 |
| Std. Deviation | 2.74 | 4.27 | 6.93 | 2.18 | 3.12 | 5.43 | 2.28 | 1.94 | 3.24 |

While, the mean COVID-19 hospitalised deaths in the third COVID-19 epidemic wave period in the ages of 0 to 9, 10 to 19, 20 to 29, 30 to 39, 40 to 49, 50 to 59, 60 to 69, 70 to 79 and 80 to 89 years were 0.944±6.18 %, 0.217±0.074 %, 0.990±0.297 %, 4.60±2.0 %, 7.8±2.0 %, 18.1±4.0 %, 25.6±6.93 %, 21.3±5.43 % and 13.4±3.24 % respectively. Figure 5 Figure 6 shows the COVID-19 hospital death age profile in the first, second and third COVID-19 epidemic wave periods in South Africa.

**Figure 6:**
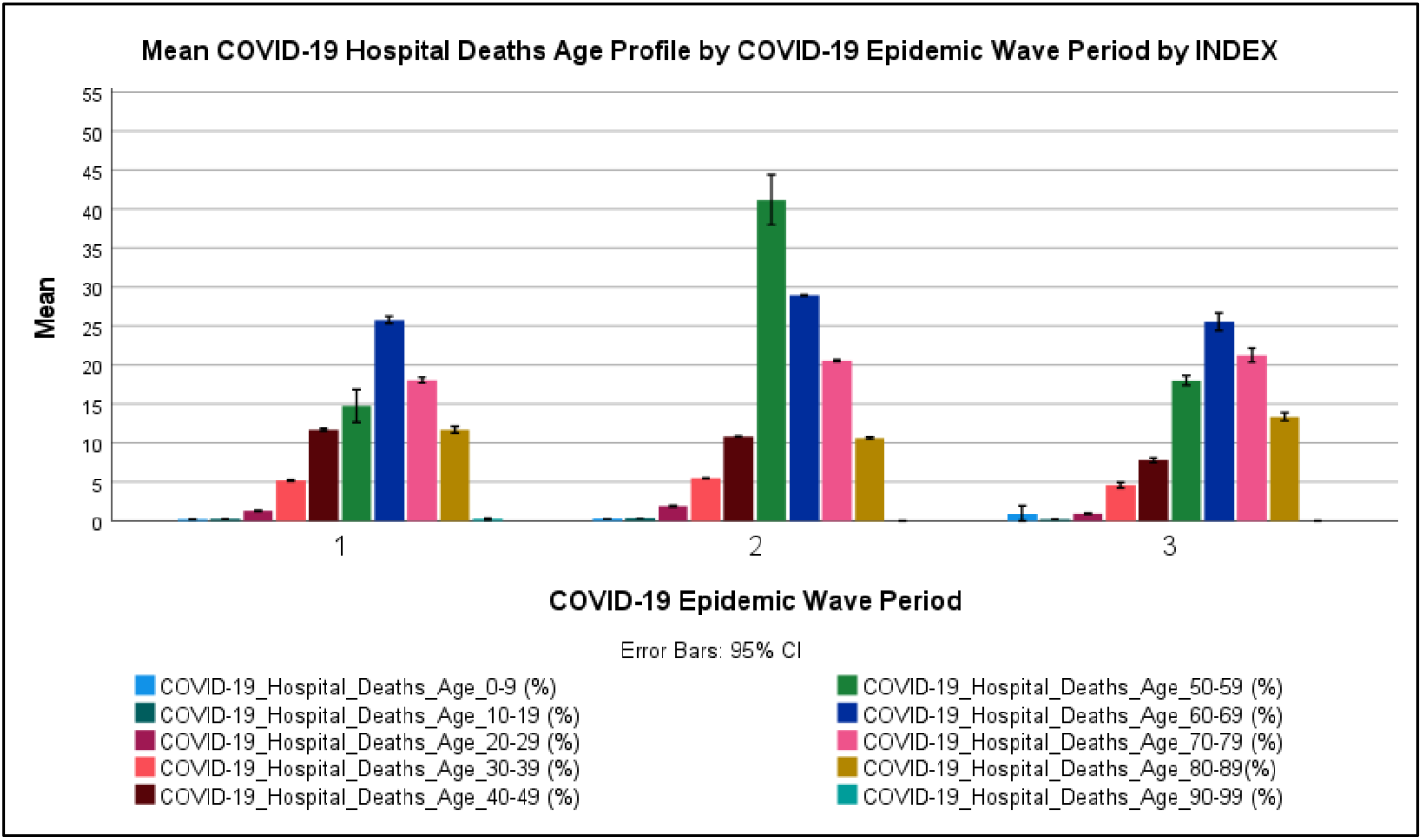
The mean COVID-19 Hospitalised Death Age Profile (with 95 % Confidence Interval (CI) Error Bars) in the First, Second and Third COVID-19 Epidemic Wave Period in South Africa

Figure 6 shows that most COVID-19 hospitalised deaths in South Africa in the first, second and third COVID-19 epidemic wave period were in the ages of 60 to 69 (25.8 %), 50 to 59 (41.2 %) and 60 to 69 (25.6 %) years respectively. COVID-19 hospitalised deaths in the age groups of 70 to 79 were the second-largest death age groups in the first and third COVID-19 epidemic wave period (18.1-21.3 %) while for the second COVID-19 epidemic wave period it was the age group of 60 to 69 years (28.5 %). COVID-19 hospitalised deaths in the age groups of 0 to 29 years were relatively low (0.194 to 1.97 %). The incidence of COVID-19 deaths in children has been reported to be low around the world [8]. Figure 6 shows that the COVID-19 hospitalised death age profile for the first, second and third COVID-19 epidemic wave periods were similar in distribution except for the high incidence of COVID-19 hospitalised deaths in the age group of 50 to 59 in the second COVID-19 epidemic wave period. In general, paired T-tests of the mean COVID-19 hospitalised deaths age groups between the Pair 1, Pair 2 and Pair 3 showed significant difference at 95 % confidence interval between the respective COVID-19 epidemic periods with p-values in the range of 5.04×10^−40^ to 0.00238 respectively (shown in Table 17). Table 17Table 13 shows that the COVID-19 hospitalised deaths in age groups of 0 to 9 years in the first, second and third COVID-19 epidemic wave period were not significantly different at a 95 % confidence interval with p-values of 0.144 to 0.177.

**Table 17:**
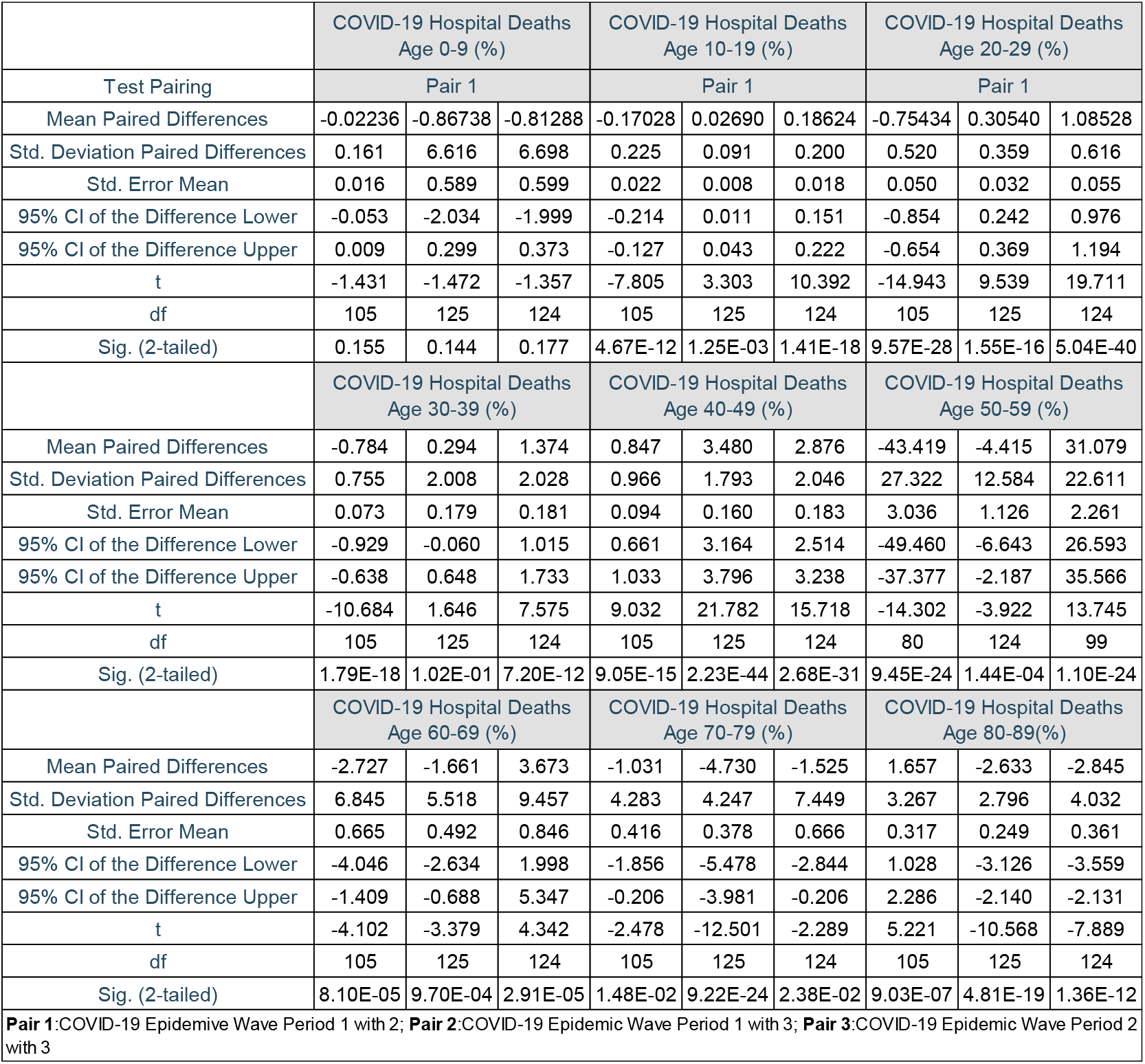
Mean Paired Differences, Standard Deviation (Std.) Paired Differences, Standard Error of Mean (Std. Error Mean), 95 % Confidence Interval (CI) if the Upper and Lower Difference, t-value, degrees of freedom (df) and P-value (Sig. (2-tailed) for the COVID-19 Hospitalised Deaths in the Ages of 0 to 9, 10 to 19, 20 to 29, 30 to 39, 40 to 49, 50 to 59, 60 to 69, 70 to 79, 80 to 89 years between Pair 1 (1^st^ and 2^nd^ COVID-19 Epidemic Wave Period), Pair 2 (1^st^ and 3^rd^ COVID-19 Epidemic Wave Period) and Pair 3 (2^nd^ and 3^rd^ COVID-19 Epidemic Wave Period) in South Africa. **(Paired Samples T-Test**)

|  | COVID-19 Hospital Deaths<br>Age 0-9 (%) |  |  | COVID-19 Hospital Deaths<br>Age 10-19 (%) |  |  | COVID-19 Hospital Deaths<br>Age 20-29 (%) |  |  |
| --- | --- | --- | --- | --- | --- | --- | --- | --- | --- |
| Test Pairing | Pair 1 |  |  | Pair 1 |  |  | Pair 1 |  |  |
| Mean Paired Differences | -0.02236 | -0.86738 | -0.81288 | -0.17028 | 0.02690 | 0.18624 | -0.75434 | 0.30540 | 1.08528 |
| Std. Deviation Paired Differences | 0.161 | 6.616 | 6.698 | 0.225 | 0.091 | 0.200 | 0.520 | 0.359 | 0.616 |
| Std. Error Mean | 0.016 | 0.589 | 0.599 | 0.022 | 0.008 | 0.018 | 0.050 | 0.032 | 0.055 |
| 95% CI of the Difference Lower | -0.053 | -2.034 | -1.999 | -0.214 | 0.011 | 0.151 | -0.854 | 0.242 | 0.976 |
| 95% CI of the Difference Upper | 0.009 | 0.299 | 0.373 | -0.127 | 0.043 | 0.222 | -0.654 | 0.369 | 1.194 |
| t | -1.431 | -1.472 | -1.357 | -7.805 | 3.303 | 10.392 | -14.943 | 9.539 | 19.711 |
| df | 105 | 125 | 124 | 105 | 125 | 124 | 105 | 125 | 124 |
| Sig. (2-tailed) | 0.155 | 0.144 | 0.177 | 4.67E-12 | 1.25E-03 | 1.41E-18 | 9.57E-28 | 1.55E-16 | 5.04E-40 |
|  | COVID-19 Hospital Deaths<br>Age 30-39 (%) |  |  | COVID-19 Hospital Deaths<br>Age 40-49 (%) |  |  | COVID-19 Hospital Deaths<br>Age 50-59 (%) |  |  |
| Mean Paired Differences | -0.784 | 0.294 | 1.374 | 0.847 | 3.480 | 2.876 | -43.419 | -4.415 | 31.079 |
| Std. Deviation Paired Differences | 0.755 | 2.008 | 2.028 | 0.966 | 1.793 | 2.046 | 27.322 | 12.584 | 22.611 |
| Std. Error Mean | 0.073 | 0.179 | 0.181 | 0.094 | 0.160 | 0.183 | 3.036 | 1.126 | 2.261 |
| 95% CI of the Difference Lower | -0.929 | -0.060 | 1.015 | 0.661 | 3.164 | 2.514 | -49.460 | -6.643 | 26.593 |
| 95% CI of the Difference Upper | -0.638 | 0.648 | 1.733 | 1.033 | 3.796 | 3.238 | -37.377 | -2.187 | 35.566 |
| t | -10.684 | 1.646 | 7.575 | 9.032 | 21.782 | 15.718 | -14.302 | -3.922 | 13.745 |
| df | 105 | 125 | 124 | 105 | 125 | 124 | 80 | 124 | 99 |
| Sig. (2-tailed) | 1.79E-18 | 1.02E-01 | 7.20E-12 | 9.05E-15 | 2.23E-44 | 2.68E-31 | 9.45E-24 | 1.44E-04 | 1.10E-24 |
|  | COVID-19 Hospital Deaths<br>Age 60-69 (%) |  |  | COVID-19 Hospital Deaths<br>Age 70-79 (%) |  |  | COVID-19 Hospital Deaths<br>Age 80-89(%) |  |  |
| Mean Paired Differences | -2.727 | -1.661 | 3.673 | -1.031 | -4.730 | -1.525 | 1.657 | -2.633 | -2.845 |
| Std. Deviation Paired Differences | 6.845 | 5.518 | 9.457 | 4.283 | 4.247 | 7.449 | 3.267 | 2.796 | 4.032 |
| Std. Error Mean | 0.665 | 0.492 | 0.846 | 0.416 | 0.378 | 0.666 | 0.317 | 0.249 | 0.361 |
| 95% CI of the Difference Lower | -4.046 | -2.634 | 1.998 | -1.856 | -5.478 | -2.844 | 1.028 | -3.126 | -3.559 |
| 95% CI of the Difference Upper | -1.409 | -0.688 | 5.347 | -0.206 | -3.981 | -0.206 | 2.286 | -2.140 | -2.131 |
| t | -4.102 | -3.379 | 4.342 | -2.478 | -12.501 | -2.289 | 5.221 | -10.568 | -7.889 |
| df | 105 | 125 | 124 | 105 | 125 | 124 | 105 | 125 | 124 |
| Sig. (2-tailed) | 8.10E-05 | 9.70E-04 | 2.91E-05 | 1.48E-02 | 9.22E-24 | 2.38E-02 | 9.03E-07 | 4.81E-19 | 1.36E-12 |
| **Pair 1:** COVID-19 Epidemic Wave Period 1 with 2; **Pair 2:** COVID-19 Epidemic Wave Period 1 with 3; **Pair 3:** COVID-19 Epidemic Wave Period 2 with 3 |  |  |  |  |  |  |  |  |  |

Table 18 shows the cumulative COVID-19 death risk ratio for South African COVID-19 hospital death age groups for the first, second and third COVID-19 epidemic wave period in South Africa regarding the age group of 0 to 9 years. The cumulative risk of death in COVID-19 hospitalised deaths increased with increasing age group groups with age groups of 50 to 69 years having the highest risk.

**Table 18:** Cumulative COVID-19 Death Risk Ratio (CFARR) for South African hospital Death Age Groups (Age Group 0-9 as reference (Ref)) for the First, Second and Third COVID-19 Epidemic Wave Period in South Africa **(Cumulative Risk**)

| Cumulative Risk Ratios |  |  |  |
| --- | --- | --- | --- |
|  | CFARR |  |  |
| COVID-19 Epidemic Wave Period | 1 | 2 | 3 |
| COVID-19 Hospital Deaths Age 0-9 (%) | 1.00 | 1.00 | 1.00 |
| COVID-19 Hospital Deaths Age 10-19 (%) | 1.31 | 1.59 | 0.23 |
| COVID-19 Hospital Deaths Age 20-29 (%) | 6.91 | 7.85 | 1.05 |
| COVID-19 Hospital Deaths Age 30-39 (%) | 26.78 | 22.66 | 4.88 |
| COVID-19 Hospital Deaths Age 40-49 (%) | 60.38 | 43.18 | 8.29 |
| COVID-19 Hospital Deaths Age 50-59 (%) | 76.01 | 164.86 | 19.13 |
| COVID-19 Hospital Deaths Age 60-69 (%) | 132.96 | 114.10 | 27.11 |
| COVID-19 Hospital Deaths Age 70-79 (%) | 93.24 | 80.00 | 22.55 |
| COVID-19 Hospital Deaths Age 80-89(%) | 60.47 | 42.42 | 14.20 |

The results in Section 3.2.4 suggest that the SARS-CoV-2 lineage clusters observed in the first, second and third COVID-19 epidemic wave periods in South Africa resulted in similar distributions of the COVID-19 hospitalised death age profile. The results also show that the SARS-CoV-2 lineage cluster 2 (predominantly Beta (B.1.351) SARS-CoV-2 VOC) resulted in an increase in the incident of hospitalised deaths in age groups of 50 to 59 compared to SARS-CoV-2 lineage cluster 1 (predominantly B.1.1.54, B.1.1.56 C.1 SARS-CoV-2 lineages) and SARS-CoV-2 lineage cluster 3 (predominantly Delta (B.1.617.2) SARS-CoV-2 VOC).

#### 3.2.5 Impact of SARS-CoV-2 Lineages on the CFR, DR and ECDR

Table 19 shows the descriptive statistics for the COVID-19 hospital case fatality rate (COVID-19 CRF), hospital discharge rate (COVID-19 DR), natural deaths, excess natural deaths, weekly reported COVID-19 deaths and the weekly unreported excess deaths (natural) to COVID-19 death ratio (ECDR) for the first, second and third COVID-19 epidemic wave periods in South Africa. Table 19 shows that the mean COVID-19 hospital case fatality rate in South Africa’s first, second and third COVID-19 epidemic wave periods were 2.06±1.10 %, 2.29±1.60 % and 2.08±1.16 % respectively. The mean COVID-19 hospital discharge rate in South Africa’s first, second and third COVID-19 epidemic wave periods were 8.40±4.89 %, 7.73±8.89 % and 6.84±2.72 % respectively. The mean weekly natural deaths in South Africa’s first, second and third COVID-19 epidemic wave period were 10 398±2 420, 12 006±4 301 and 14 506±2 826 respectively. The mean weekly excess natural deaths in South Africa’s first, second and third COVID-19 epidemic wave period were 1 619±2 110, 3 864±4 243 and 5 271±2 636 respectively. The mean weekly reported COVID-19 deaths in South Africa’s first, second and third COVID-19 epidemic wave period were 614±556, 1 226±1 153 and 1 719±808 respectively. The Weekly Unreported Excess Deaths (Natural) to COVID-19 Death Ratio (ECDR) allows for the accounting of potential COVID-19 pandemic related natural deaths that are not reported. Table 19 shows the mean ECDR in South Africa’s first, second and third COVID-19 epidemic wave period were 1.06±1.68, 2.17±1.07 and 2.27±0.99 respectively. These results suggest that the number of excess natural deaths not accounted for in the COVID-19 reported deaths were the same as the COVID-19 reported deaths in the first COVID-19 epidemic wave period and twice the COVID-19 reported deaths in the second and third COVID-19 epidemic wave period.

**Table 19:** Statistical Sample Number (N), Minimum, Maximum, Mean, Standard Deviation (Std. Deviation) of COVID-19 Hospital Case Fatality Rate (COVID-19 CRF), Hospital Discharge Rate (COVID-19 DR), Weekly Natural Deaths, Excess Natural Deaths, Reported COVID-19 Deaths and the Weekly Unreported Excess Deaths (Natural) to COVID-19 Death Ratio (ECDR) in the First, Second and Third COVID-19 Epidemic Wave Period in South Africa **(Descriptive Statistics**)

| Descriptive Statistics |  |  |  |  |  |  |  |  |  |  |  |  |  |  |  |  |  |  |
| --- | --- | --- | --- | --- | --- | --- | --- | --- | --- | --- | --- | --- | --- | --- | --- | --- | --- | --- |
|  | COVID-19 CFR |  |  | COVID-19 DR |  |  | Natural Deaths |  |  | Excess Natural Deaths |  |  | Weekly Reported COVID-19 Deaths |  |  | ECDR |  |  |
| COVID-19 Epidemic Wave Period | 1 | 2 | 3 | 1 | 2 | 3 | 1 | 2 | 3 | 1 | 2 | 3 | 1 | 2 | 3 | 1 | 2 | 3 |
| N | 121 | 183 | 146 | 121 | 183 | 146 | 30 | 30 | 18 | 29 | 30 | 18 | 30 | 30 | 18 | 30 | 30 | 18 |
| Minimum | 0.00% | 0.00% | 0.09% | 0.00% | -25.62% | -3.91% | 7932 | 9027 | 10690 | 871 | 831 | 1899 | 1 | 281 | 566 | -0.49 | 0.41 | 0.93 |
| Maximum | 6.87% | 12.89% | 10.11% | 42.52% | 77.62% | 15.96% | 15864 | 24198 | 19873 | 6673 | 16106 | 10253 | 2057 | 4027 | 2812 | 5.69 | 4.67 | 4.64 |
| Mean | 2.06% | 2.29% | 2.08% | 8.40% | 7.73% | 6.84% | 10398 | 12006 | 14506 | 1619 | 3864 | 5271 | 614 | 1226 | 1719 | 1.06 | 2.17 | 2.27 |
| Std. Deviation | 1.10% | 1.60% | 1.16% | 4.89% | 8.98% | 2.72% | 2420 | 4301 | 2826 | 2110 | 4243 | 2636 | 556 | 1153 | 808 | 1.68 | 1.07 | 0.99 |

Paired T-tests of the COVID-19 hospital case fatality rate in Pair 1, Pair 2 and Pair 3 showed no significant difference at 99 % confidence interval with p-values of 0.028, 0.431 and 0.146 respectively (shown in Table 20). This result suggests that the mean COVID-19 hospital case fatality rates in South Africa’s three COVID-19 epidemic wave periods were statistically similar. Paired T-tests of the COVID-19 hospital discharge rate in Pair 1 and Pair 3 showed no significant difference at 95 % confidence interval with p-values of 0.703 and 0.179 respectively (shown in Table 20). While the paired T-test in Pair 2 showed a significant difference at a 95 % confidence interval with a p-value of 0.005. This result suggests that the mean COVID-19 hospital discharge rate in the first and second, second and third COVID-19 epidemic wave periods were statistically similar while that between the first and third COVID-19 wave periods were different. Paired T-tests of the excess natural deaths, weekly reported COVID-19 deaths and ECDR in Pair 3 showed no significant difference at 95 % confidence interval with p-values of 0.908, 0.918 and 0.760 respectively (shown in Table 20). While the paired T-test in Pair 1 and Pair 2 showed a significant difference at a 95 % confidence interval with p-values in the range of 0.001 and 3.64×10^−06^ to 0.015 (shown in Table 20). These results suggest that the mean excess natural deaths weekly reported COVID-19 deaths and ECDR in the second and third COVID-19 epidemic wave periods were statistically similar while that between the first and third, first and second COVID-19 wave periods were statistically different.

**Table 20:** Mean Paired Differences, Standard Deviation (Std.) Paired Differences, Standard Error of Mean (Std. Error Mean), 95 % Confidence Interval (CI) if the Upper and Lower Difference, t-value, degrees of freedom (df) and P-value (Sig. (2-tailed) for the COVID-19 Hospital Case Fatality Rate (COVID-19 CRF), Hospital Discharge Rate (COVID-19 DR), Natural Deaths, Excess Natural Deaths, Weekly Reported COVID-19 Deaths and the Weekly Unreported Excess Deaths (Natural) to COVID-19 Death Ratio (ECDR)between Pair 1 (1^st^ and 2^nd^ COVID-19 Epidemic Wave Period), Pair 2 (1^st^ and 3^rd^ COVID-19 Epidemic Wave Period) and Pair 3 (2^nd^ and 3^rd^ COVID-19 Epidemic Wave Period) in South Africa. **(Paired Samples T-Test**)

|  | COVID-19 CFR |  |  | COVID-19 Discharge Rate |  |  | Excess Natural Deaths |  |  | Weekly Reported COVID-19 Deaths |  |  | Excess (Natural) to Natural Deaths (%) |  |  |
| --- | --- | --- | --- | --- | --- | --- | --- | --- | --- | --- | --- | --- | --- | --- | --- |
| Test Pairing | Pair 1 | Pair 2 | Pair 3 | Pair 1 | Pair 2 | Pair 3 | Pair 1 | Pair 2 | Pair 3 | Pair 1 | Pair 2 | Pair 3 | Pair 1 | Pair 2 | Pair 3 |
| Mean Paired Differences | -0.005 | -0.001 | 0.003 | 0.004 | 0.015 | 0.014 | -2341 | -3945 | 127 | -612 | -1349 | -22 | -14.57 | -24.53 | -1.13 |
| Std. Deviation Paired Differences | 0.020 | 0.016 | 0.020 | 0.111 | 0.057 | 0.112 | 4660 | 4088 | 4553 | 1296 | 852 | 905 | 20.06 | 20.53 | 15.44 |
| Std. Error Mean | 0.002 | 0.001 | 0.002 | 0.011 | 0.005 | 0.010 | 865 | 992 | 1073 | 237 | 201 | 213 | 3.66 | 4.84 | 3.64 |
| 95% CI of the Difference Lower | -0.009 | -0.004 | -0.001 | -0.018 | 0.004 | -0.006 | -4113 | -6047 | -2137 | -1096 | -1773 | -472 | -22.06 | -34.74 | -8.81 |
| 95% CI of the Difference Upper | 0.000 | 0.002 | 0.006 | 0.026 | 0.025 | 0.034 | -568 | -1843 | 2390 | -128 | -925 | 428 | -7.08 | -14.32 | 6.55 |
| t | -2.227 | -0.790 | 1.463 | 0.383 | 2.834 | 1.350 | -3 | -4 | 0 | -3 | -7 | 0 | -3.98 | -5.07 | -0.31 |
| df | 98 | 120 | 120 | 98 | 120 | 120 | 28 | 16 | 17 | 29 | 17 | 17 | 29.00 | 17.00 | 17.00 |
| Sig. (2-tailed) | 0.028 | 0.431 | 0.146 | 0.703 | 0.005 | 0.179 | 0.011 | 0.001 | 0.908 | 0.015 | 3.64E-06 | 0.918 | 4.25E-04 | 9.47E-05 | 0.760 |
| Pair 1: COVID-19 Epidemic Wave Period 1 with 2; Pair 2: COVID-19 Epidemic Wave Period 1 with 3; Pair 3: COVID-19 Epidemic Wave Period 2 with 3 |  |  |  |  |  |  |  |  |  |  |  |  |  |  |  |

The results in Section 3.2.5 suggest that the SARS-CoV-2 lineage clusters identified in the first, second and third COVID-19 epidemic wave periods in South Africa resulted in similar COVID-19 hospital case-fatality rates. The SARS-CoV-2 lineage cluster 2 (predominantly Beta (B.1.351) SARS-CoV-2 VOC) and the SARS-CoV-2 lineage cluster 3 (predominantly Delta (B.1.617.2) SARS-CoV-2 VOC) resulted in similar COVID-19 discharge rates, excess natural deaths, weekly reported COVID-19 deaths and ECDR. While, the SARS-CoV-2 lineage cluster 1 (predominantly B.1.1.54, B.1.1.56 C.1 SARS-CoV-2 lineages) resulted in statistically different excess natural deaths, weekly reported COVID-19 deaths and ECDR relative to the aforementioned SARS-CoV-2 clusters.

## 4. Conclusions

Based on the COVID-19 detection rate in South Africa, the SA SARS-CoV-2 lineage cluster 3 which was predominantly the Delta (B.1.617.2) SARS-CoV-2 VOC resulted in an increase in the COVID-19 transmissibility in the South African population by 53.9 to 54.8 %. This is relative to the SA SARS-CoV-2 lineage cluster 1 (predominantly B.1.1.54, B.1.1.56 C.1 SARS-CoV-2 lineages) and the SA SARS-CoV-2 lineage cluster 2 (predominantly Beta (B.1.351) SARS-CoV-2 VOC) respectively. The SA SARS-CoV-2 lineage cluster 1 and 2 resulted in similar COVID-19 transmissibility in the South African population.

Based on the daily COVID-19 hospitalisations per active case, the SA SARS-CoV-2 lineage cluster 2 (predominantly the Beta (B.1.351) SARS-CoV-2 VOC) was observed to result in increased hospitalisation relative to the SA SARS-CoV-2 lineage clusters 1 and 3. COVID-19 hospitalisations per active case were observed to have increased by 113 % and 36.8 % respectively. While the SA SARS-CoV-2 lineage cluster 3 (predominantly the Delta (B.1.617.2) SARS-CoV-2 VOC) was also observed to have resulted in an increased COVID-19 hospitalisation relative to the SA SARS-CoV-2 lineage cluster 1. The COVID-19 hospitalisations per active case increased by 55.8 %. SA SARS-CoV-2 lineage clusters 1, 2 and 3 resulted in a similar distribution of the COVID-19 hospital admission status profile in South African hospitals. Most COVID-19 hospitalised cases in South Africa were hospitalised in the general ward (72.8-78.7 %). COVID-19 patients on oxygen were the second-largest hospital admission status (16.8-27.6 %) in South Africa followed by the COVID-19 patients in the intensive care unit (14.1-16.7 %). SA SARS-CoV-2 lineage clusters 1 and 3 resulted in the same COVID-19 patients admitted to the intensive care unit. While the SA SARS-CoV-2 lineage cluster 2 resulted in higher COVID-19 patients on oxygen than the SA SARS-CoV-2 lineage clusters 1 and 3. This result highlights the increased COVID-19 severity due to the Beta SARS-CoV-2 VOC. The SA SARS-CoV-2 lineage clusters 1, 2 and 3 resulted in different distributions of the COVID-19 hospital admission age profile. Most COVID-19 hospitalised cases in South Africa were in the ages of 50 to 69 years. COVID-19 patients in the age groups of 40 to 49 years were the second-largest hospital admitted age group in South Africa. COVID-19 admitted patients in the age groups of 0 to 19 years were relatively low (1.8 to 4.4 %) and highest in the second COVID-19 epidemic wave period. The SA SARS-CoV-2 lineage cluster 3 (predominantly the Delta (B.1.617.2) SARS-COV-2 VOC) resulted in an increased incidence in COVID-19 patients in the age groups over 70 years compared to the SA SARS-CoV-2 lineage clusters 1 and 2. The SA SARS-CoV-2 lineage clusters 1, 2 and 3 resulted in a similar distribution of the COVID-19 hospitalised death age profile. Most COVID-19 hospitalised deaths in South Africa in the first, second and third COVID-19 epidemic wave period were in the ages of 50 to 69 years. COVID-19 hospitalised deaths in the age groups of 0 to 29 years in South Africa were relatively low (0.194 to 1.97 %). The cumulative risk of death in COVID-19 hospitalised deaths increased with increasing age group groups with age groups of 50 to 69 years having the highest risk. The SA SARS-CoV-2 lineage cluster 2 (predominantly the Beta (B.1.351) SARS-CoV-2 VOC) resulted in an increase in the incidence of hospitalised deaths in age groups of 50 to 59 compared to the SARS-CoV-2 lineage clusters 1 and 3.

The mean COVID-19 hospital case fatality rate in South Africa’s first, second and third COVID-19 epidemic wave periods were 2.06±1.10 %, 2.29±1.60 % and 2.08±1.16 % respectively. The number of excess natural deaths not accounted for in the COVID-19 reported deaths were the same as the COVID-19 reported deaths in the first COVID-19 epidemic wave period and twice the COVID-19 reported deaths in the second and third COVID-19 epidemic wave period. The SA SARS-CoV-2 lineage clusters 1, 2 and 3 resulted in similar COVID-19 hospital case-fatality rates in South Africa. While the SA SARS-CoV-2 lineages clusters 2 and 3 resulted in similar COVID-19 discharge rates, excess natural deaths, weekly reported COVID-19 deaths and ECDR in South Africa. The SA SARS-CoV-2 lineage cluster 1 (predominantly the B.1.1.54, B.1.1.56 C.1 SARS-CoV-2 lineages) resulted in statistically different excess natural deaths, weekly reported COVID-19 deaths and ECDR in South Africa relative to the aforementioned SARS-CoV-2 clusters.

This study shows that the evolution of SARS-CoV-2 resulted in an increase in COVID-19 transmissibility and severity in South Africa. The Delta SARS-CoV-2 VOC resulted in increased COVID-19 transmissibility in the South African population while both the Beta SARS-CoV-2 VOC and Delta SARS-CoV-2 VOC resulted in more severe COVID-19 than the initial SARS-CoV-2 lineages detected in South Africa’s first epidemic wave period.

## Supporting information

Analysis_Dataset_(COVID-19 Hospitalised Cases Admission Age Profile)

SPSS_Program_(COVID-19 Hospitalised Cases Admission Age)

Analysis_Dataset_(COVID-19 Hospitalised Cases Admission Status)

SPSS_Program_(COVID-19 Hospitalised Cases Admission Status)

Analysis_Dataset_(COVID-19 Hospitalised Cases Discharge,Death)

SPSS_Program_(COVID-19 Hospitalised Cases Discharge, Deaths)

Analysis_Dataset_(COVID-19 Hospitalised Deaths Age Profile)

SPSS_Program_(COVID-19 Hospitalised Deaths Age Profile)

Analysis_Dataset_(COVID-19 Reported Cases)

SPSS_Program_(COVID-19 Reported Cases)

Analysis_Dataset_(Weekly COVID-19-Excess Deaths)

SPSS_Program_(Weekly COVID-19-Excess Deaths)

## Data Availability

All data produced in the present study are available upon reasonable request to the authors

https://www.afrikanresearchinitiative.com/

## 5 Acknowledgements

ARI would like to thank the members of the ARI African Disease Demographic Research Group (ADDRG) and African COVID-19 Modelling Research Group (ACMRG) for their voluntary commitment to working in the ARI COVID-19 Research Project. We acknowledge the work by the National Institute for Communicable Diseases (NICD), Western Cape Department of Health Provincial Health Data Centre (PHDC), South African Medical Research Council (SAMRC) and the Network for Genomics Surveillance in South Africa (NGS-SA) in which the ARI COVID-19 Project draws a lot of its data from. Lastly, we want to salute the scientific community, governments, health care workers, essential personnel in their response to the pandemic and we pay homage to those who have lost their lives due to the COVID-19 pandemic.

