## Supplementary material for "The Impact of SARS-CoV-2 Lineages (Variants) on the COVID-19 Epidemic in South Africa": SPSS_Program_(COVID-19 Hospitalised Cases Admission Age)

```
 GET DATA
  /TYPE=XLSX
  /FILE='C:\Users\ThaboMabuka\Google Drive\ARI Projects\Research Projects\COVID-19 in Afr
ica\Papers\ACMRG\The Impact of SARS-CoV-2 Variants on the COVID-19 Epidemic in South Afri
ca\Data\P2_Analysis_Dataset_(COVID-19 Hospitalised Cases Admission Age Profile).xlsx'
  /SHEET=name 'Hospitalised_Case_Data_Age_Prof'
  /CELLRANGE=FULL
  /READNAMES=ON
  /LEADINGSPACES IGNORE=YES
  /TRAILINGSPACES IGNORE=YES
  /DATATYPEMIN PERCENTAGE=95.0
  /HIDDEN IGNORE=YES.
EXECUTE.
DATASET NAME DataSet1 WINDOW=FRONT.

SAVE OUTFILE='C:\Users\ThaboMabuka\Google Drive\ARI Projects\Research Projects\COVID-19 i
n '+
    'Africa\Papers\ACMRG\The Impact of SARS-CoV-2 Variants on the COVID-19 Epidemic in So
uth '+
    'Africa\Data\P2_Analysis_Dataset_(COVID-19 Hospitalised Cases Admission Age Profile).
sav'
  /COMPRESSED.
DATASET ACTIVATE DataSet1.

SAVE OUTFILE='C:\Users\ThaboMabuka\Google Drive\ARI Projects\Research Projects\COVID-19 i
n '+
    'Africa\Papers\ACMRG\The Impact of SARS-CoV-2 Variants on the COVID-19 Epidemic in So
uth '+
    'Africa\Data\P2_Analysis_Dataset_(COVID-19 Hospitalised Cases Admission Age Profile).
sav'
  /COMPRESSED.
* Define Variable Properties.
*EpidemicWave.
VARIABLE LEVEL  EpidemicWave(SCALE).
EXECUTE.
SORT CASES  BY EpidemicWave.
SPLIT FILE LAYERED BY EpidemicWave.
DATASET ACTIVATE DataSet1.

SAVE OUTFILE='C:\Users\ThaboMabuka\Google Drive\ARI Projects\Research Projects\COVID-19 i
n '+
    'Africa\Papers\ACMRG\The Impact of SARS-CoV-2 Variants on the COVID-19 Epidemic in So
uth '+
    'Africa\Data\P2_Analysis_Dataset_(COVID-19 Hospitalised Cases Admission Age Profile).
sav'
  /COMPRESSED.
```

```
DESCRIPTIVES VARIABLES=COVID19_Hospital_Admitted_CasesAdj COVID19_Hospitalised_Admitted_A
ge_09nAdj
    COVID19_Hospitalised_Admitted_Age_1019nAdj COVID19_Hospitalised_Admitted_Age_2029nAdj
    COVID19_Hospitalised_Admitted_Age_3039nAdj COVID19_Hospitalised_Admitted_Age_4049nAdj
    COVID19_Hospitalised_Admitted_Age_5059nAdj COVID19_Hospitalised_Admitted_Age_6069nAdj
    COVID19_Hospitalised_Admitted_Age_7079nAdj COVID19_Hospitalised_Admitted_Age_8089nAdj
    COVID19_Hospitalised_Admitted_Age_9099nAdj
  /STATISTICS=MEAN STDDEV MIN MAX.
```

### Descriptives

#### Notes

| | | |
|---|---|---|
| Output Created | | 24-SEP-2021 13:42:16 |
| Comments | | |
| Input | Data | C:\Users\ThaboMabuka\Google Drive\ARI Projects\Research Projects\COVID-19 in Africa\Papers\ACMRG\The Impact of SARS-CoV-2 Variants on the COVID-19 Epidemic in South Africa\Data\P2_Analysis_Dataset_(COVID-19 Hospitalised Cases Admission Age Profile).sav |
| | Active Dataset | DataSet1 |
| | Filter | <none> |
| | Weight | <none> |
| | Split File | Epidemic Wave |
| | N of Rows in Working Data File | 484 |
| Missing Value Handling | Definition of Missing | User defined missing values are treated as missing. |
| | Cases Used | All non-missing data are used. |

**Notes**

| | | |
|---|---|---|
| Syntax | | DESCRIPTIVES VARIABLES=COVID19_Hospital_Admitted_CasesAdj COVID19_Hospitalised_Admitted_Age_09nAdj<br><br>COVID19_Hospitalised_Admitted_Age_1019nAdj COVID19_Hospitalised_Admitted_Age_2029nAdj<br><br>COVID19_Hospitalised_Admitted_Age_3039nAdj COVID19_Hospitalised_Admitted_Age_4049nAdj<br><br>COVID19_Hospitalised_Admitted_Age_5059nAdj COVID19_Hospitalised_Admitted_Age_6069nAdj<br><br>COVID19_Hospitalised_Admitted_Age_7079nAdj COVID19_Hospitalised_Admitted_Age_8089nAdj<br><br>COVID19_Hospitalised_Admitted_Age_9099nAdj<br>/STATISTICS=MEAN STDDEV MIN MAX. |
| Resources | Processor Time | 00:00:00,02 |
| | Elapsed Time | 00:00:00,02 |

[DataSet1] C:\Users\ThaboMabuka\Google Drive\ARI Projects\Research Projects\COVID-19 in Africa\Papers\ACMRG\The Impact of SARS-CoV-2 Variants on the COVID-19 Epidemic in South Africa\Data\P2_Analysis_Dataset_(COVID-19 Hospitalised Cases Admission Age Profile).sav

**Descriptive Statistics**

| Epidemic Wave | | N | Minimum | Maximum | Mean | Std. Deviation |
|---|---|---|---|---|---|---|
| 1 | COVID-19_Hospital_Admitted_Cases Adj | 126 | 2487 | 72675 | 33886.15 | 23604.312 |
| | COVID-19_Hospitalised_Admitted_Age_0-9 (n) Adj | 110 | 78 | 1259 | 588.03 | 369.991 |
| | COVID-19_Hospitalised_Admitted_Age_10-19 (n) Adj | 108 | 36 | 1160 | 498.26 | 358.950 |
| | COVID-19_Hospitalised_Admitted_Age_20-29 (n) Adj | 126 | 1 | 4637 | 2336.69 | 1370.268 |
| | COVID-19_Hospitalised_Admitted_Age_30-39 (n) Adj | 126 | 283 | 10785 | 5542.13 | 3415.844 |
| | COVID-19_Hospitalised_Admitted_Age_40-49 (n) Adj | 75 | 535 | 13711 | 8091.09 | 4599.106 |
| | COVID-19_Hospitalised_Admitted_Age_50-59 (n) Adj | 126 | 177 | 17288 | 8629.18 | 5869.226 |
| | COVID-19_Hospitalised_Admitted_Age_60-69 (n) Adj | 125 | 373 | 12913 | 6082.80 | 4309.646 |
| | COVID-19_Hospitalised_Admitted_Age_70-79 (n) Adj | 126 | 164 | 7674 | 3577.62 | 2549.440 |
| | COVID-19_Hospitalised_Admitted_Age_80-89 (n) Adj | 126 | 0 | 4408 | 2009.44 | 1500.876 |
| | COVID-19_Hospitalised_Admitted_Age_90-99 (n) Adj | 16 | 23 | 113 | 64.19 | 33.628 |
| | Valid N (listwise) | 16 | | | | |
| 2 | COVID-19_Hospital_Admitted_Cases Adj | 180 | 544 | 185807 | 108276.23 | 62616.392 |
| | COVID-19_Hospitalised_Admitted_Age_0-9 (n) Adj | 187 | 9 | 3704 | 2005.53 | 1269.632 |
| | COVID-19_Hospitalised_Admitted_Age_10-19 (n) Adj | 179 | 1944 | 5500 | 3762.46 | 1192.323 |

### Descriptive Statistics

| Epidemic Wave | | N | Minimum | Maximum | Mean | Std. Deviation |
|---|---|---|---|---|---|---|
| | COVID-19_Hospitalised_Admitted_Age_20-29 (n) Adj | 187 | -3282 | 12710 | 7266.44 | 4080.157 |
| | COVID-19_Hospitalised_Admitted_Age_30-39 (n) Adj | 187 | 122 | 24544 | 14117.93 | 8172.464 |
| | COVID-19_Hospitalised_Admitted_Age_40-49 (n) Adj | 181 | 111 | 31809 | 18089.20 | 10345.167 |
| | COVID-19_Hospitalised_Admitted_Age_50-59 (n) Adj | 186 | 92 | 39964 | 22524.18 | 14238.960 |
| | COVID-19_Hospitalised_Admitted_Age_60-69 (n) Adj | 186 | 69 | 36362 | 20039.11 | 13053.260 |
| | COVID-19_Hospitalised_Admitted_Age_70-79 (n) Adj | 187 | -7674 | 22252 | 11861.08 | 8395.599 |
| | COVID-19_Hospitalised_Admitted_Age_80-89 (n) Adj | 187 | -4408 | 10274 | 5413.60 | 3896.362 |
| | COVID-19_Hospitalised_Admitted_Age_90-99 (n) Adj | 0 | | | | |
| | Valid N (listwise) | 0 | | | | |
| 3 | COVID-19_Hospital_Admitted_Cases Adj | 145 | 27 | 872061 | 108692.69 | 142059.125 |
| | COVID-19_Hospitalised_Admitted_Age_0-9 (n) Adj | 145 | 6 | 4754 | 1883.52 | 1585.966 |
| | COVID-19_Hospitalised_Admitted_Age_10-19 (n) Adj | 145 | 3 | 4717 | 1784.54 | 1514.629 |
| | COVID-19_Hospitalised_Admitted_Age_20-29 (n) Adj | 145 | 3 | 163798 | 5539.10 | 13734.789 |
| | COVID-19_Hospitalised_Admitted_Age_30-39 (n) Adj | 145 | 2 | 20364 | 8407.53 | 6951.186 |
| | COVID-19_Hospitalised_Admitted_Age_40-49 (n) Adj | 143 | 88 | 25423 | 10983.09 | 8869.081 |

### Descriptive Statistics

| Epidemic Wave | | N | Minimum | Maximum | Mean | Std. Deviation |
|---|---|---|---|---|---|---|
| | COVID-19_Hospitalised_Admitted_Age_50-59 (n) Adj | 145 | 18 | 34935 | 15386.77 | 12338.755 |
| | COVID-19_Hospitalised_Admitted_Age_60-69 (n) Adj | 144 | 137 | 29432 | 13462.90 | 10165.269 |
| | COVID-19_Hospitalised_Admitted_Age_70-79 (n) Adj | 144 | -2 | 21256 | 9552.92 | 7391.585 |
| | COVID-19_Hospitalised_Admitted_Age_80-89 (n) Adj | 145 | 1 | 11544 | 5117.92 | 4043.080 |
| | COVID-19_Hospitalised_Admitted_Age_90-99 (n) Adj | 0 | | | | |
| | Valid N (listwise) | 0 | | | | |

```
 DATASET ACTIVATE DataSet1.

SAVE OUTFILE='C:\Users\ThaboMabuka\Google Drive\ARI Projects\Research Projects\COVID-19 i
n '+
    'Africa\Papers\ACMRG\The Impact of SARS-CoV-2 Variants on the COVID-19 Epidemic in So
uth '+
    'Africa\Data\P2_Analysis_Dataset_(COVID-19 Hospitalised Cases Admission Age Profile).
sav'
  /COMPRESSED.
DESCRIPTIVES VARIABLES=COVID19_Hospitalised_Admitted_Age_09p
    COVID19_Hospitalised_Admitted_Age_1019p COVID19_Hospitalised_Admitted_Age_2029p
    COVID19_Hospitalised_Admitted_Age_3039p COVID19_Hospitalised_Admitted_Age_4049p
    COVID19_Hospitalised_Admitted_Age_5059p COVID19_Hospitalised_Admitted_Age_6069p
    COVID19_Hospitalised_Admitted_Age_7079p COVID19_Hospitalised_Admitted_Age_8089p
  /STATISTICS=MEAN STDDEV MIN MAX.
```

### Descriptives

### Notes

| | | |
|---|---|---|
| Output Created | | 24-SEP-2021 13:46:02 |
| Comments | | |
| Input | Data | C:\Users\ThaboMabuka\Google Drive\ARI Projects\Research Projects\COVID-19 in Africa\Papers\ACMRG\The Impact of SARS-CoV-2 Variants on the COVID-19 Epidemic in South Africa\Data\P2_Analysis_Dataset_(COVID-19 Hospitalised Cases Admission Age Profile).sav |
| | Active Dataset | DataSet1 |
| | Filter | <none> |
| | Weight | <none> |
| | Split File | Epidemic Wave |
| | N of Rows in Working Data File | 484 |
| Missing Value Handling | Definition of Missing | User defined missing values are treated as missing. |
| | Cases Used | All non-missing data are used. |

### Notes

| | | |
|---|---|---|
| Syntax | | DESCRIPTIVES VARIABLES=COVID19_Hospitalised_Admitted_Age_09p<br><br>COVID19_Hospitalised_Admitted_Age_1019p COVID19_Hospitalised_Admitted_Age_2029p<br><br>COVID19_Hospitalised_Admitted_Age_3039p COVID19_Hospitalised_Admitted_Age_4049p<br><br>COVID19_Hospitalised_Admitted_Age_5059p COVID19_Hospitalised_Admitted_Age_6069p<br><br>COVID19_Hospitalised_Admitted_Age_7079p COVID19_Hospitalised_Admitted_Age_8089p<br>/STATISTICS=MEAN STDDEV MIN MAX. |
| Resources | Processor Time | 00:00:00,00 |
| | Elapsed Time | 00:00:00,01 |

### Descriptive Statistics

| Epidemic Wave | | N | Minimum | Maximum | Mean |
|---|---|---|---|---|---|
| 1 | COVID-19_Hospitalised_Admitted_Age_-9 (%) | 110 | 1.675313228 | 3.963175364 | 2.361431461 |
| | COVID-19_Hospitalised_Admitted_Age_1-19 (%) | 108 | 1.227412259 | 2.941354387 | 1.808271501 |
| | COVID-19_Hospitalised_Admitted_Age_2-29 (%) | 126 | .4298725372 | 14.59167334 | 7.543636452 |
| | COVID-19_Hospitalised_Admitted_Age_3-39 (%) | 126 | 2.418925579 | 29.82385987 | 17.44257124 |
| | COVID-19_Hospitalised_Admitted_Age_4-49 (%) | 75 | 2.194599855 | 27.35821334 | 19.50917121 |
| | COVID-19_Hospitalised_Admitted_Age_5-59 (%) | 126 | 3.132586675 | 38.42449842 | 24.03139816 |
| | COVID-19_Hospitalised_Admitted_Age_6-69 (%) | 125 | 1.475785224 | 24.33947158 | 16.20351919 |
| | COVID-19_Hospitalised_Admitted_Age_7-79 (%) | 126 | 1.332855357 | 16.33918278 | 7.122307752 |
| | COVID-19_Hospitalised_Admitted_Age_8> (%) | 125 | .8763619138 | 9.432565227 | 5.585726516 |
| | Valid N (listwise) | 49 | | | |
| 2 | COVID-19_Hospitalised_Admitted_Age_-9 (%) | 180 | 1.427124927 | 2.951471892 | 1.944617795 |
| | COVID-19_Hospitalised_Admitted_Age_1-19 (%) | 179 | 1.234176229 | 9.875259875 | 4.428142511 |
| | COVID-19_Hospitalised_Admitted_Age_2-29 (%) | 180 | -2.57787832 | 13.51475882 | 7.241102817 |
| | COVID-19_Hospitalised_Admitted_Age_3-39 (%) | 180 | 1.937462245 | 22.42647588 | 14.24980486 |
| | COVID-19_Hospitalised_Admitted_Age_4-49 (%) | 180 | 2.444117648 | 18.55883878 | 16.95072448 |

### Descriptive Statistics

| Epidemic Wave | | Std. Deviation |
|---|---|---|
| 1 | COVID-19_Hospitalised_Admitted_Age_-9 (%) | .5905730788 |
| | COVID-19_Hospitalised_Admitted_Age_1-19 (%) | .3258803823 |
| | COVID-19_Hospitalised_Admitted_Age_2-29 (%) | 2.558389437 |
| | COVID-19_Hospitalised_Admitted_Age_3-39 (%) | 4.409429234 |
| | COVID-19_Hospitalised_Admitted_Age_4-49 (%) | 4.863219140 |
| | COVID-19_Hospitalised_Admitted_Age_5-59 (%) | 6.063214497 |
| | COVID-19_Hospitalised_Admitted_Age_6-69 (%) | 5.418769393 |
| | COVID-19_Hospitalised_Admitted_Age_7-79 (%) | 4.431948324 |
| | COVID-19_Hospitalised_Admitted_Age_8> (%) | 1.586242624 |
| | Valid N (listwise) | |
| 2 | COVID-19_Hospitalised_Admitted_Age_-9 (%) | .2529089611 |
| | COVID-19_Hospitalised_Admitted_Age_1-19 (%) | 2.215244950 |
| | COVID-19_Hospitalised_Admitted_Age_2-29 (%) | 2.007615424 |
| | COVID-19_Hospitalised_Admitted_Age_3-39 (%) | 1.672246078 |
| | COVID-19_Hospitalised_Admitted_Age_4-49 (%) | 1.174046116 |

### Descriptive Statistics

| Epidemic Wave | | N | Minimum | Maximum | Mean |
|---|---|---|---|---|---|
| | COVID-19_Hospitalised_Admitted_Age_5-59 (%) | 180 | 2.196229312 | 24.21257678 | 19.58326440 |
| | COVID-19_Hospitalised_Admitted_Age_6-69 (%) | 180 | 12.68382353 | 21.54515678 | 18.17093018 |
| | COVID-19_Hospitalised_Admitted_Age_7-79 (%) | 180 | 1.142151956 | 12.96479572 | 9.836544613 |
| | COVID-19_Hospitalised_Admitted_Age_8> (%) | 180 | 2.755867795 | 6.433823529 | 5.021271894 |
| | Valid N (listwise) | 179 | | | |
| 3 | COVID-19_Hospitalised_Admitted_Age_-9 (%) | 143 | .1796624386 | 3.715172786 | 2.303641983 |
| | COVID-19_Hospitalised_Admitted_Age_1-19 (%) | 143 | .1943981592 | 3.869969425 | 2.301898275 |
| | COVID-19_Hospitalised_Admitted_Age_2-29 (%) | 143 | 1.212529857 | 92.83395759 | 6.400934605 |
| | COVID-19_Hospitalised_Admitted_Age_3-39 (%) | 143 | .8626648950 | 12.78527979 | 9.422052246 |
| | COVID-19_Hospitalised_Admitted_Age_4-49 (%) | 143 | 1.949773863 | 15.59535392 | 13.53301749 |
| | COVID-19_Hospitalised_Admitted_Age_5-59 (%) | 143 | 1.588572653 | 22.88873169 | 12.85774624 |
| | COVID-19_Hospitalised_Admitted_Age_6-69 (%) | 143 | 1.631139987 | 22.26947862 | 15.50391475 |
| | COVID-19_Hospitalised_Admitted_Age_7-79 (%) | 142 | 1.813751828 | 14.95641285 | 12.81489046 |
| | COVID-19_Hospitalised_Admitted_Age_8> (%) | 143 | .5338864896 | 7.944923685 | 6.479383415 |
| | Valid N (listwise) | 142 | | | |

### Descriptive Statistics

| Epidemic Wave | | Std. Deviation |
|---|---|---|
| | COVID-19_Hospitalised_Admitted_Age_5-59 (%) | 4.806732896 |
| | COVID-19_Hospitalised_Admitted_Age_6-69 (%) | 1.759374536 |
| | COVID-19_Hospitalised_Admitted_Age_7-79 (%) | 3.186588159 |
| | COVID-19_Hospitalised_Admitted_Age_8> (%) | .5584807721 |
| | Valid N (listwise) | |
| 3 | COVID-19_Hospitalised_Admitted_Age_-9 (%) | .4942584211 |
| | COVID-19_Hospitalised_Admitted_Age_1-19 (%) | .4794853978 |
| | COVID-19_Hospitalised_Admitted_Age_2-29 (%) | 7.433585834 |
| | COVID-19_Hospitalised_Admitted_Age_3-39 (%) | 3.846468675 |
| | COVID-19_Hospitalised_Admitted_Age_4-49 (%) | 3.293144367 |
| | COVID-19_Hospitalised_Admitted_Age_5-59 (%) | 9.076652193 |
| | COVID-19_Hospitalised_Admitted_Age_6-69 (%) | 7.354439364 |
| | COVID-19_Hospitalised_Admitted_Age_7-79 (%) | 3.193311434 |
| | COVID-19_Hospitalised_Admitted_Age_8> (%) | 1.676387658 |
| | Valid N (listwise) | |

```
* Define Variable Properties.
```

```
*EpidemicWave.
VARIABLE LEVEL  EpidemicWave(NOMINAL).
EXECUTE.
SPSSINC SPLIT DATASET SPLITVAR=EpidemicWave
/OUTPUT DIRECTORY= "C:\Users\ThaboMabuka\Google Drive\ARI Projects\Research Projects\COVI
D-19 in "+
    "Africa\Papers\ACMRG\The Impact of SARS-CoV-2 Variants on the COVID-19 Epidemic in So
uth "+
    "Africa\Data" DELETECONTENTS=NO
/OPTIONS NAMES=VALUES.

>Warning # 10903.  Command name: AGGREGATE
>A SPLIT FILE command is in effect but will be ignored for AGGREGATE
>processing.
```

### SPSSINC SPLIT DATASET

#### Notes

| | | |
|---|---|---|
| Output Created | | 24-SEP-2021 13:49:53 |
| Comments | | |
| Input | Data | C:\Users\ThaboMabuka\Google Drive\ARI Projects\Research Projects\COVID-19 in Africa\Papers\ACMRG\The Impact of SARS-CoV-2 Variants on the COVID-19 Epidemic in South Africa\Data\P2_Analysis_Dataset_(COVID-19 Hospitalised Cases Admission Age Profile).sav |
| | Active Dataset | DataSet1 |
| | Filter | <none> |
| | Weight | <none> |
| | Split File | Epidemic Wave |
| Syntax | | BEGIN PROGRAM '# '. |
| Resources | Processor Time | 00:00:00,00 |
| | Elapsed Time | 00:00:00,03 |

```
[DataSet1] C:\Users\ThaboMabuka\Google Drive\ARI Projects\Research Projects\COVID-19 in A
frica\Papers\ACMRG\The Impact of SARS-CoV-2 Variants on the COVID-19 Epidemic in South Af
rica\Data\P2_Analysis_Dataset_(COVID-19 Hospitalised Cases Admission Age Profile).sav
```

**Split File Information**

| | Settings and Statistics |
| --- | --- |
| Split Variable Names | EpidemicWave |
| Output Directory | C:\Users\Thabo Mabuka\Google Drive\ARI Projects\Research Projects\COVID-19 in Africa\Papers\ACMRG\The Impact of SARS-CoV-2 Variants on the COVID-19 Epidemic in South Africa\Data |
| Files Deleted | 0 |
| Files Written | 3 |
| File List | None |
| Directories Cleared | No |

### Values and File Names for Split Files Written

| | Values or Labels | Directory | Data File |
|---|---|---|---|
| 1 | 1 | C:\Users\Thabo Mabuka\Google Drive\ARI Projects\Research Projects\COVID-19 in Africa\Papers\ACMRG\The Impact of SARS-CoV-2 Variants on the COVID-19 Epidemic in South Africa\Data | 1.sav |

#### Values and File Names for Split Files Written

| | Values or Labels | Directory | Data File |
|---|---|---|---|
| 2 | 2 | C: \Users\Thabo Mabuka\Google Drive\ARI Projects\Research Projects\COVID-19 in Africa\Papers\ACMRG\The Impact of SARS-CoV-2 Variants on the COVID-19 Epidemic in South Africa\Data | 2.sav |
| 3 | 3 | C: \Users\Thabo Mabuka\Google Drive\ARI Projects\Research Projects\COVID-19 in Africa\Papers\ACMRG\The Impact of SARS-CoV-2 Variants on the COVID-19 Epidemic in South Africa\Data | 3.sav |

Based on Variables: EpidemicWave

```
DATASET ACTIVATE DataSet1.

SAVE OUTFILE='C:\Users\ThaboMabuka\Google Drive\ARI Projects\Research Projects\COVID-19 i
n '+
    'Africa\Papers\ACMRG\The Impact of SARS-CoV-2 Variants on the COVID-19 Epidemic in So
uth '+
    'Africa\Data\P2_Analysis_Dataset_(COVID-19 Hospitalised Cases Admission Age Profile).
sav'
```

```
  /COMPRESSED.



 DATASET ACTIVATE DataSet2.

SAVE OUTFILE='C:\Users\ThaboMabuka\Google Drive\ARI Projects\Research Projects\COVID-19 i
n '+
    'Africa\Papers\ACMRG\The Impact of SARS-CoV-2 Variants on the COVID-19 Epidemic in So
uth '+
    'Africa\Data\P2_Analysis_(COVID-19 Hospitalised Cases Admission Age Profile_2.sav'
  /COMPRESSED.
DATASET ACTIVATE DataSet1.
GET
  FILE='C:\Users\ThaboMabuka\Google Drive\ARI Projects\Research Projects\COVID-19 in Afri
ca\Papers\ACMRG\The Impact of SARS-CoV-2 Variants on the COVID-19 Epidemic in South Afric
a\Data\P2_Analysis_(COVID-19 Hospitalised Cases Admission Age Profile_3.sav'.
DATASET NAME DataSet3 WINDOW=FRONT.
DATASET ACTIVATE DataSet3.

SAVE OUTFILE='C:\Users\ThaboMabuka\Google Drive\ARI Projects\Research Projects\COVID-19 i
n '+
    'Africa\Papers\ACMRG\The Impact of SARS-CoV-2 Variants on the COVID-19 Epidemic in So
uth '+
    'Africa\Data\P2_Analysis_(COVID-19 Hospitalised Cases Admission Age Profile_3.sav'
  /COMPRESSED.
DATASET ACTIVATE DataSet3.

SAVE OUTFILE='C:\Users\ThaboMabuka\Google Drive\ARI Projects\Research Projects\COVID-19 i
n '+
    'Africa\Papers\ACMRG\The Impact of SARS-CoV-2 Variants on the COVID-19 Epidemic in So
uth '+
    'Africa\Data\P2_Analysis_(COVID-19 Hospitalised Cases Admission Age Profile_3.sav'
  /COMPRESSED.
DATASET ACTIVATE DataSet1.
SORT CASES BY Data_Point.
DATASET ACTIVATE DataSet2.
SORT CASES BY Data_Point.
DATASET ACTIVATE DataSet1.
MATCH FILES /FILE=*
  /FILE='DataSet2'
  /BY Data_Point.
EXECUTE.
SORT CASES BY Data_Point.
DATASET ACTIVATE DataSet3.
SORT CASES BY Data_Point.
DATASET ACTIVATE DataSet1.
MATCH FILES /FILE=*
  /FILE='DataSet3'
```

```
  /BY Data_Point.
EXECUTE.
File #2
     KEY:        .

>Warning # 5132
>Duplicate key in a file.  The BY variables do not uniquely identify each case
>on the indicated file.  Please check the results carefully.
DATASET CLOSE DataSet1.
DATASET ACTIVATE DataSet3.
GET
  FILE='C:\Users\ThaboMabuka\Google Drive\ARI Projects\Research Projects\COVID-19 in Afri
ca\Papers\ACMRG\The Impact of SARS-CoV-2 Variants on the COVID-19 Epidemic in South Afric
a\Data\P2_Analysis_(COVID-19 Hospitalised Cases Admission Age Profile_1.sav'.
DATASET NAME DataSet4 WINDOW=FRONT.
DATASET ACTIVATE DataSet3.
DATASET ACTIVATE DataSet3.

SAVE OUTFILE='C:\Users\ThaboMabuka\Google Drive\ARI Projects\Research Projects\COVID-19 i
n '+
    'Africa\Papers\ACMRG\The Impact of SARS-CoV-2 Variants on the COVID-19 Epidemic in So
uth '+
    'Africa\Data\P2_Analysis_(COVID-19 Hospitalised Cases Admission Age Profile_3.sav'
  /COMPRESSED.
DATASET ACTIVATE DataSet4.
SORT CASES BY Data_Point.
DATASET ACTIVATE DataSet2.
SORT CASES BY Data_Point.
DATASET ACTIVATE DataSet4.
MATCH FILES /FILE=*
  /FILE='DataSet2'
  /BY Data_Point.
EXECUTE.
SORT CASES BY Data_Point.
DATASET ACTIVATE DataSet3.
SORT CASES BY Data_Point.
DATASET ACTIVATE DataSet4.
MATCH FILES /FILE=*
  /FILE='DataSet3'
  /BY Data_Point.
EXECUTE.
DATASET ACTIVATE DataSet4.

SAVE OUTFILE='C:\Users\ThaboMabuka\Google Drive\ARI Projects\Research Projects\COVID-19 i
n '+
    'Africa\Papers\ACMRG\The Impact of SARS-CoV-2 Variants on the COVID-19 Epidemic in So
uth '+
    'Africa\Data\P2_Analysis_(COVID-19 Hospitalised Cases Admission Age Profile_1.sav'
  /COMPRESSED.
```

```
SAVE OUTFILE='C:\Users\ThaboMabuka\Google Drive\ARI Projects\Research Projects\COVID-19 i
n '+
    'Africa\Papers\ACMRG\The Impact of SARS-CoV-2 Variants on the COVID-19 Epidemic in So
uth '+
    'Africa\Data\P2_Analysis_(COVID-19 Hospitalised Cases Admission Age Profile_M.sav'
  /COMPRESSED.
DATASET ACTIVATE DataSet3.
DATASET ACTIVATE DataSet3.

SAVE OUTFILE='C:\Users\ThaboMabuka\Google Drive\ARI Projects\Research Projects\COVID-19 i
n '+
    'Africa\Papers\ACMRG\The Impact of SARS-CoV-2 Variants on the COVID-19 Epidemic in So
uth '+
    'Africa\Data\P2_Analysis_(COVID-19 Hospitalised Cases Admission Age Profile_3.sav'
  /COMPRESSED.
DATASET ACTIVATE DataSet2.
DATASET CLOSE DataSet3.
DATASET ACTIVATE DataSet2.

SAVE OUTFILE='C:\Users\ThaboMabuka\Google Drive\ARI Projects\Research Projects\COVID-19 i
n '+
    'Africa\Papers\ACMRG\The Impact of SARS-CoV-2 Variants on the COVID-19 Epidemic in So
uth '+
    'Africa\Data\P2_Analysis_(COVID-19 Hospitalised Cases Admission Age Profile_2.sav'
  /COMPRESSED.
DATASET ACTIVATE DataSet4.
DATASET CLOSE DataSet2.
GET
  FILE='C:\Users\ThaboMabuka\Google Drive\ARI Projects\Research Projects\COVID-19 in Afri
ca\Papers\ACMRG\The Impact of SARS-CoV-2 Variants on the COVID-19 Epidemic in South Afric
a\Data\P2_Analysis_(COVID-19 Hospitalised Cases Admission Age Profile_1.sav'.
DATASET NAME DataSet5 WINDOW=FRONT.
DATASET ACTIVATE DataSet4.
DATASET CLOSE DataSet5.
GET
  FILE='C:\Users\ThaboMabuka\Google Drive\ARI Projects\Research Projects\COVID-19 in Afri
ca\Papers\ACMRG\The Impact of SARS-CoV-2 Variants on the COVID-19 Epidemic in South Afric
a\Data\P2_Analysis_(COVID-19 Hospitalised Cases Admission Age Profile_1.sav'.
DATASET NAME DataSet6 WINDOW=FRONT.
DATASET ACTIVATE DataSet6.

SAVE OUTFILE='C:\Users\ThaboMabuka\Google Drive\ARI Projects\Research Projects\COVID-19 i
n '+
    'Africa\Papers\ACMRG\The Impact of SARS-CoV-2 Variants on the COVID-19 Epidemic in So
uth '+
    'Africa\Data\P2_Analysis_(COVID-19 Hospitalised Cases Admission Age Profile_1.sav'
  /COMPRESSED.
DATASET ACTIVATE DataSet6.
```

```
SAVE OUTFILE='C:\Users\ThaboMabuka\Google Drive\ARI Projects\Research Projects\COVID-19 i
n '+
    'Africa\Papers\ACMRG\The Impact of SARS-CoV-2 Variants on the COVID-19 Epidemic in So
uth '+
    'Africa\Data\P2_Analysis_(COVID-19 Hospitalised Cases Admission Age Profile_1.sav'
  /COMPRESSED.
DATASET ACTIVATE DataSet4.
DATASET CLOSE DataSet6.
T-TEST PAIRS=COVID19_Hospital_Admitted_CasesAdj_1 COVID19_Hospital_Admitted_CasesAdj_1
    COVID19_Hospital_Admitted_CasesAdj_2 COVID19_Hospitalised_Admitted_Age_09nAdj_1
    COVID19_Hospitalised_Admitted_Age_09nAdj_1 COVID19_Hospitalised_Admitted_Age_09nAdj_2
    COVID19_Hospitalised_Admitted_Age_1019nAdj_1 COVID19_Hospitalised_Admitted_Age_1019nA
dj_1
    COVID19_Hospitalised_Admitted_Age_1019nAdj_2 COVID19_Hospitalised_Admitted_Age_2029nA
dj_1
    COVID19_Hospitalised_Admitted_Age_2029nAdj_1 COVID19_Hospitalised_Admitted_Age_2029nA
dj_2
    COVID19_Hospitalised_Admitted_Age_3039nAdj_1 COVID19_Hospitalised_Admitted_Age_3039nA
dj_1
    COVID19_Hospitalised_Admitted_Age_3039nAdj_2 COVID19_Hospitalised_Admitted_Age_4049nA
dj_1
    COVID19_Hospitalised_Admitted_Age_4049nAdj_1 COVID19_Hospitalised_Admitted_Age_4049nA
dj_2
    COVID19_Hospitalised_Admitted_Age_5059nAdj_1 COVID19_Hospitalised_Admitted_Age_5059nA
dj_1
    COVID19_Hospitalised_Admitted_Age_5059nAdj_2 COVID19_Hospitalised_Admitted_Age_6069nA
dj_1
    COVID19_Hospitalised_Admitted_Age_6069nAdj_1 COVID19_Hospitalised_Admitted_Age_6069nA
dj_2
    COVID19_Hospitalised_Admitted_Age_7079nAdj_1 COVID19_Hospitalised_Admitted_Age_7079nA
dj_1
    COVID19_Hospitalised_Admitted_Age_7079nAdj_2 COVID19_Hospitalised_Admitted_Age_8089nA
dj_1
    COVID19_Hospitalised_Admitted_Age_8089nAdj_1 COVID19_Hospitalised_Admitted_Age_8089nA
dj_2 WITH
    COVID19_Hospital_Admitted_CasesAdj_2 COVID19_Hospital_Admitted_CasesAdj_3
    COVID19_Hospital_Admitted_CasesAdj_3 COVID19_Hospitalised_Admitted_Age_09nAdj_2
    COVID19_Hospitalised_Admitted_Age_09nAdj_3 COVID19_Hospitalised_Admitted_Age_09nAdj_3
    COVID19_Hospitalised_Admitted_Age_1019nAdj_2 COVID19_Hospitalised_Admitted_Age_1019nA
dj_3
    COVID19_Hospitalised_Admitted_Age_1019nAdj_3 COVID19_Hospitalised_Admitted_Age_2029nA
dj_2
    COVID19_Hospitalised_Admitted_Age_2029nAdj_3 COVID19_Hospitalised_Admitted_Age_2029nA
dj_3
    COVID19_Hospitalised_Admitted_Age_3039nAdj_2 COVID19_Hospitalised_Admitted_Age_3039nA
dj_3
    COVID19_Hospitalised_Admitted_Age_3039nAdj_3 COVID19_Hospitalised_Admitted_Age_4049nA
dj_2
```

```
    COVID19_Hospitalised_Admitted_Age_4049nAdj_3 COVID19_Hospitalised_Admitted_Age_4049nA
dj_3
    COVID19_Hospitalised_Admitted_Age_5059nAdj_2 COVID19_Hospitalised_Admitted_Age_5059nA
dj_3
    COVID19_Hospitalised_Admitted_Age_5059nAdj_3 COVID19_Hospitalised_Admitted_Age_6069nA
dj_2
    COVID19_Hospitalised_Admitted_Age_6069nAdj_3 COVID19_Hospitalised_Admitted_Age_6069nA
dj_3
    COVID19_Hospitalised_Admitted_Age_7079nAdj_2 COVID19_Hospitalised_Admitted_Age_7079nA
dj_3
    COVID19_Hospitalised_Admitted_Age_7079nAdj_3 COVID19_Hospitalised_Admitted_Age_8089nA
dj_2
    COVID19_Hospitalised_Admitted_Age_8089nAdj_3 COVID19_Hospitalised_Admitted_Age_8089nA
dj_3 (PAIRED)
  /ES DISPLAY(TRUE) STANDARDIZER(SD)
  /CRITERIA=CI(.9500)
  /MISSING=ANALYSIS.
```

### T-Test

#### Notes

| | | |
|---|---|---|
| Output Created | | 24-SEP-2021 15:04:31 |
| Comments | | |
| Input | Data | C:\Users\ThaboMabuka\Google Drive\ARI Projects\Research Projects\COVID-19 in Africa\Papers\ACMRG\The Impact of SARS-CoV-2 Variants on the COVID-19 Epidemic in South Africa\Data\P2_Analysis_(COVID-19 Hospitalised Cases Admission Age Profile_M.sav |
| | Active Dataset | DataSet4 |
| | Filter | <none> |
| | Weight | <none> |
| | Split File | <none> |
| | N of Rows in Working Data File | 208 |

**Notes**

| | | |
|---|---|---|
| Missing Value Handling | Definition of Missing | User defined missing values are treated as missing. |
| | Cases Used | Statistics for each analysis are based on the cases with no missing or out-of-range data for any variable in the analysis. |

### Notes

| | |
|---|---|
| Syntax | T-TEST<br>PAIRS=COVID19_Hospital_Admitted_CasesAdj_1 COVID19_Hospital_Admitted_CasesAdj_1<br><br>COVID19_Hospital_Admitted_CasesAdj_2 COVID19_Hospitalised_Admitted_Age_09nAdj_1<br><br>COVID19_Hospitalised_Admitted_Age_09nAdj_1 COVID19_Hospitalised_Admitted_Age_09nAdj_2<br><br>COVID19_Hospitalised_Admitted_Age_1019nAdj_1 COVID19_Hospitalised_Admitted_Age_1019nAdj_1<br><br>COVID19_Hospitalised_Admitted_Age_1019nAdj_2 COVID19_Hospitalised_Admitted_Age_2029nAdj_1<br><br>COVID19_Hospitalised_Admitted_Age_2029nAdj_1 COVID19_Hospitalised_Admitted_Age_2029nAdj_2<br><br>COVID19_Hospitalised_Admitted_Age_3039nAdj_1 COVID19_Hospitalised_Admitted_Age_3039nAdj_1<br><br>COVID19_Hospitalised_Admitted_Age_3039nAdj_2 COVID19_Hospitalised_Admitted_Age_4049nAdj_1<br><br>COVID19_Hospitalised_Admitted_Age_4049nAdj_1 COVID19_Hospitalised_Admitted_Age_4049nAdj_2<br><br>COVID19_Hospitalised_Admitted_Age_5059nAdj_1 COVID19_Hospitalised_Admitted_Age_5059nAdj_1<br><br>COVID19_Hospitalised_Admitted_Age_5059nAdj_2 COVID19_Hospitalised_Admitted_Age_6069nAdj_1<br><br>COVID19_Hospitalised_Admitted_Age_6069nAdj_1 COVID19_Hospitalised_Admitted_Age_6069nAdj_2<br><br>COVID19_Hospitalised_A [illegible] |

### Notes

| | | |
|---|---|---|
| Resources | Processor Time | 00:00:00,03 |
| | Elapsed Time | 00:00:00,03 |

### Paired Samples Statistics

| | | Mean | N | Std. Deviation | Std. Error Mean |
|---|---|---|---|---|---|
| Pair 1 | COVID-19_Hospital_Admitted_Cases Adj_1 | 40596.93 | 99 | 20848.397 | 2095.343 |
| | COVID-19_Hospital_Admitted_Cases Adj_2 | 59307.74 | 99 | 37929.607 | 3812.069 |
| Pair 2 | COVID-19_Hospital_Admitted_Cases Adj_1 | 33761.15 | 125 | 23657.392 | 2115.981 |
| | COVID-19_Hospital_Admitted_Cases Adj_3 | 65264.50 | 125 | 71981.017 | 6438.178 |
| Pair 3 | COVID-19_Hospital_Admitted_Cases Adj_2 | 73039.97 | 117 | 48745.286 | 4506.503 |
| | COVID-19_Hospital_Admitted_Cases Adj_3 | 131563.02 | 117 | 148683.349 | 13745.780 |
| Pair 4 | COVID-19_Hospitalised_Admitted_Age_0-9 (n) Adj_1 | 660.38 | 90 | 348.698 | 36.756 |
| | COVID-19_Hospitalised_Admitted_Age_0-9 (n) Adj_2 | 908.47 | 90 | 693.942 | 73.148 |
| Pair 5 | COVID-19_Hospitalised_Admitted_Age_0-9 (n) Adj_1 | 588.03 | 110 | 369.991 | 35.277 |
| | COVID-19_Hospitalised_Admitted_Age_0-9 (n) Adj_3 | 1313.99 | 110 | 1260.253 | 120.160 |
| Pair 6 | COVID-19_Hospitalised_Admitted_Age_0-9 (n) Adj_2 | 1287.38 | 124 | 922.981 | 82.886 |
| | COVID-19_Hospitalised_Admitted_Age_0-9 (n) Adj_3 | 2130.32 | 124 | 1542.127 | 138.487 |

### Paired Samples Statistics

| | | Mean | N | Std. Deviation | Std. Error Mean |
|---|---|---|---|---|---|
| Pair 7 | COVID-19_Hospitalised_Admitted_Age_10-19 (n) Adj_1 | 627.63 | 81 | 312.366 | 34.707 |
| | COVID-19_Hospitalised_Admitted_Age_10-19 (n) Adj_2 | 2626.91 | 81 | 477.773 | 53.086 |
| Pair 8 | COVID-19_Hospitalised_Admitted_Age_10-19 (n) Adj_1 | 493.44 | 107 | 357.111 | 34.523 |
| | COVID-19_Hospitalised_Admitted_Age_10-19 (n) Adj_3 | 1096.04 | 107 | 940.970 | 90.967 |
| Pair 9 | COVID-19_Hospitalised_Admitted_Age_10-19 (n) Adj_2 | 3040.83 | 116 | 802.242 | 74.486 |
| | COVID-19_Hospitalised_Admitted_Age_10-19 (n) Adj_3 | 2150.65 | 116 | 1432.995 | 133.050 |
| Pair 10 | COVID-19_Hospitalised_Admitted_Age_20-29 (n) Adj_1 | 2598.82 | 106 | 1269.946 | 123.348 |
| | COVID-19_Hospitalised_Admitted_Age_20-29 (n) Adj_2 | 4297.83 | 106 | 2322.287 | 225.560 |
| Pair 11 | COVID-19_Hospitalised_Admitted_Age_20-29 (n) Adj_1 | 2325.52 | 125 | 1370.010 | 122.537 |
| | COVID-19_Hospitalised_Admitted_Age_20-29 (n) Adj_3 | 3603.31 | 125 | 3083.773 | 275.821 |
| Pair 12 | COVID-19_Hospitalised_Admitted_Age_20-29 (n) Adj_2 | 4977.89 | 124 | 3009.524 | 270.263 |
| | COVID-19_Hospitalised_Admitted_Age_20-29 (n) Adj_3 | 6293.16 | 124 | 14701.781 | 1320.259 |
| Pair 13 | COVID-19_Hospitalised_Admitted_Age_30-39 (n) Adj_1 | 6198.66 | 106 | 3167.291 | 307.635 |
| | COVID-19_Hospitalised_Admitted_Age_30-39 (n) Adj_2 | 7947.19 | 106 | 4922.029 | 478.070 |

### Paired Samples Statistics

| | | Mean | N | Std. Deviation | Std. Error Mean |
|---|---|---|---|---|---|
| Pair 14 | COVID-19_Hospitalised_Admitted_Age_30-39 (n) Adj_1 | 5513.37 | 125 | 3414.240 | 305.379 |
| | COVID-19_Hospitalised_Admitted_Age_30-39 (n) Adj_3 | 6938.48 | 125 | 5943.525 | 531.605 |
| Pair 15 | COVID-19_Hospitalised_Admitted_Age_30-39 (n) Adj_2 | 9626.76 | 124 | 6257.557 | 561.945 |
| | COVID-19_Hospitalised_Admitted_Age_30-39 (n) Adj_3 | 9491.40 | 124 | 6754.653 | 606.586 |
| Pair 16 | COVID-19_Hospitalised_Admitted_Age_40-49 (n) Adj_1 | 9783.55 | 58 | 3303.428 | 433.761 |
| | COVID-19_Hospitalised_Admitted_Age_40-49 (n) Adj_2 | 12430.09 | 58 | 7012.502 | 920.787 |
| Pair 17 | COVID-19_Hospitalised_Admitted_Age_40-49 (n) Adj_1 | 8212.30 | 73 | 4573.835 | 535.327 |
| | COVID-19_Hospitalised_Admitted_Age_40-49 (n) Adj_3 | 11337.19 | 73 | 8133.965 | 952.009 |
| Pair 18 | COVID-19_Hospitalised_Admitted_Age_40-49 (n) Adj_2 | 12543.63 | 116 | 8213.831 | 762.635 |
| | COVID-19_Hospitalised_Admitted_Age_40-49 (n) Adj_3 | 13067.74 | 116 | 8290.605 | 769.763 |
| Pair 19 | COVID-19_Hospitalised_Admitted_Age_50-59 (n) Adj_1 | 9845.68 | 105 | 5388.291 | 525.843 |
| | COVID-19_Hospitalised_Admitted_Age_50-59 (n) Adj_2 | 11786.31 | 105 | 8893.485 | 867.915 |
| Pair 20 | COVID-19_Hospitalised_Admitted_Age_50-59 (n) Adj_1 | 8579.01 | 125 | 5865.651 | 524.640 |
| | COVID-19_Hospitalised_Admitted_Age_50-59 (n) Adj_3 | 12952.74 | 125 | 10887.368 | 973.796 |

### Paired Samples Statistics

| | | Mean | N | Std. Deviation | Std. Error Mean |
|---|---|---|---|---|---|
| Pair 21 | COVID-19_Hospitalised_Admitted_Age_50-59 (n) Adj_2 | 14841.82 | 123 | 11301.594 | 1019.030 |
| | COVID-19_Hospitalised_Admitted_Age_50-59 (n) Adj_3 | 17516.48 | 123 | 11841.502 | 1067.712 |
| Pair 22 | COVID-19_Hospitalised_Admitted_Age_60-69 (n) Adj_1 | 6946.02 | 104 | 4005.368 | 392.759 |
| | COVID-19_Hospitalised_Admitted_Age_60-69 (n) Adj_2 | 10051.51 | 104 | 7893.019 | 773.974 |
| Pair 23 | COVID-19_Hospitalised_Admitted_Age_60-69 (n) Adj_1 | 6090.81 | 123 | 4293.773 | 387.156 |
| | COVID-19_Hospitalised_Admitted_Age_60-69 (n) Adj_3 | 11394.87 | 123 | 8994.504 | 811.007 |
| Pair 24 | COVID-19_Hospitalised_Admitted_Age_60-69 (n) Adj_2 | 13028.93 | 122 | 10169.251 | 920.681 |
| | COVID-19_Hospitalised_Admitted_Age_60-69 (n) Adj_3 | 15313.48 | 122 | 9637.790 | 872.565 |
| Pair 25 | COVID-19_Hospitalised_Admitted_Age_70-79 (n) Adj_1 | 4049.56 | 106 | 2385.881 | 231.737 |
| | COVID-19_Hospitalised_Admitted_Age_70-79 (n) Adj_2 | 5589.16 | 106 | 5338.725 | 518.543 |
| Pair 26 | COVID-19_Hospitalised_Admitted_Age_70-79 (n) Adj_1 | 3555.66 | 125 | 2547.703 | 227.874 |
| | COVID-19_Hospitalised_Admitted_Age_70-79 (n) Adj_3 | 8187.90 | 125 | 6582.445 | 588.752 |
| Pair 27 | COVID-19_Hospitalised_Admitted_Age_70-79 (n) Adj_2 | 7274.24 | 123 | 6629.325 | 597.746 |
| | COVID-19_Hospitalised_Admitted_Age_70-79 (n) Adj_3 | 10790.93 | 123 | 7071.744 | 637.638 |

### Paired Samples Statistics

| | | Mean | N | Std. Deviation | Std. Error Mean |
|---|---|---|---|---|---|
| Pair 28 | COVID-19_Hospitalised_Admitted_Age_80-89 (n) Adj_1 | 2281.41 | 106 | 1411.997 | 137.145 |
| | COVID-19_Hospitalised_Admitted_Age_80-89 (n) Adj_2 | 2501.15 | 106 | 2473.325 | 240.231 |
| Pair 29 | COVID-19_Hospitalised_Admitted_Age_80-89 (n) Adj_1 | 1996.32 | 125 | 1499.638 | 134.132 |
| | COVID-19_Hospitalised_Admitted_Age_80-89 (n) Adj_3 | 4320.37 | 125 | 3563.849 | 318.760 |
| Pair 30 | COVID-19_Hospitalised_Admitted_Age_80-89 (n) Adj_2 | 3320.91 | 124 | 3083.981 | 276.950 |
| | COVID-19_Hospitalised_Admitted_Age_80-89 (n) Adj_3 | 5782.35 | 124 | 3882.187 | 348.631 |

### Paired Samples Correlations

| | | N | Correlation | Sig. |
|---|---|---|---|---|
| Pair 1 | COVID-19_Hospital_Admitted_Cases Adj_1 & COVID-19_Hospital_Admitted_Cases Adj_2 | 99 | .906 | .000 |
| Pair 2 | COVID-19_Hospital_Admitted_Cases Adj_1 & COVID-19_Hospital_Admitted_Cases Adj_3 | 125 | .808 | .000 |
| Pair 3 | COVID-19_Hospital_Admitted_Cases Adj_2 & COVID-19_Hospital_Admitted_Cases Adj_3 | 117 | .842 | .000 |
| Pair 4 | COVID-19_Hospitalised_Admitted_Age_0-9 (n) Adj_1 & COVID-19_Hospitalised_Admitted_Age_0-9 (n) Adj_2 | 90 | .937 | .000 |

### Paired Samples Correlations

| | | N | Correlation | Sig. |
|---|---|---|---|---|
| Pair 5 | COVID-19_Hospitalised_Admitted_Age_0-9 (n) Adj_1 & COVID-19_Hospitalised_Admitted_Age_0-9 (n) Adj_3 | 110 | .970 | .000 |
| Pair 6 | COVID-19_Hospitalised_Admitted_Age_0-9 (n) Adj_2 & COVID-19_Hospitalised_Admitted_Age_0-9 (n) Adj_3 | 124 | .990 | .000 |
| Pair 7 | COVID-19_Hospitalised_Admitted_Age_10-19 (n) Adj_1 & COVID-19_Hospitalised_Admitted_Age_10-19 (n) Adj_2 | 81 | .994 | .000 |
| Pair 8 | COVID-19_Hospitalised_Admitted_Age_10-19 (n) Adj_1 & COVID-19_Hospitalised_Admitted_Age_10-19 (n) Adj_3 | 107 | .985 | .000 |
| Pair 9 | COVID-19_Hospitalised_Admitted_Age_10-19 (n) Adj_2 & COVID-19_Hospitalised_Admitted_Age_10-19 (n) Adj_3 | 116 | .997 | .000 |
| Pair 10 | COVID-19_Hospitalised_Admitted_Age_20-29 (n) Adj_1 & COVID-19_Hospitalised_Admitted_Age_20-29 (n) Adj_2 | 106 | .965 | .000 |
| Pair 11 | COVID-19_Hospitalised_Admitted_Age_20-29 (n) Adj_1 & COVID-19_Hospitalised_Admitted_Age_20-29 (n) Adj_3 | 125 | .973 | .000 |
| Pair 12 | COVID-19_Hospitalised_Admitted_Age_20-29 (n) Adj_2 & COVID-19_Hospitalised_Admitted_Age_20-29 (n) Adj_3 | 124 | .136 | .133 |

### Paired Samples Correlations

| | | N | Correlation | Sig. |
|---|---|---|---|---|
| Pair 13 | COVID-19_Hospitalised_Admitted_Age_30-39 (n) Adj_1 & COVID-19_Hospitalised_Admitted_Age_30-39 (n) Adj_2 | 106 | .927 | .000 |
| Pair 14 | COVID-19_Hospitalised_Admitted_Age_30-39 (n) Adj_1 & COVID-19_Hospitalised_Admitted_Age_30-39 (n) Adj_3 | 125 | .972 | .000 |
| Pair 15 | COVID-19_Hospitalised_Admitted_Age_30-39 (n) Adj_2 & COVID-19_Hospitalised_Admitted_Age_30-39 (n) Adj_3 | 124 | .983 | .000 |
| Pair 16 | COVID-19_Hospitalised_Admitted_Age_40-49 (n) Adj_1 & COVID-19_Hospitalised_Admitted_Age_40-49 (n) Adj_2 | 58 | .904 | .000 |
| Pair 17 | COVID-19_Hospitalised_Admitted_Age_40-49 (n) Adj_1 & COVID-19_Hospitalised_Admitted_Age_40-49 (n) Adj_3 | 73 | .971 | .000 |
| Pair 18 | COVID-19_Hospitalised_Admitted_Age_40-49 (n) Adj_2 & COVID-19_Hospitalised_Admitted_Age_40-49 (n) Adj_3 | 116 | .975 | .000 |
| Pair 19 | COVID-19_Hospitalised_Admitted_Age_50-59 (n) Adj_1 & COVID-19_Hospitalised_Admitted_Age_50-59 (n) Adj_2 | 105 | .899 | .000 |
| Pair 20 | COVID-19_Hospitalised_Admitted_Age_50-59 (n) Adj_1 & COVID-19_Hospitalised_Admitted_Age_50-59 (n) Adj_3 | 125 | .981 | .000 |

#### Paired Samples Correlations

| | | N | Correlation | Sig. |
|---|---|---|---|---|
| Pair 21 | COVID-19_Hospitalised_Admitted_Age_50-59 (n) Adj_2 & COVID-19_Hospitalised_Admitted_Age_50-59 (n) Adj_3 | 123 | .966 | .000 |
| Pair 22 | COVID-19_Hospitalised_Admitted_Age_60-69 (n) Adj_1 & COVID-19_Hospitalised_Admitted_Age_60-69 (n) Adj_2 | 104 | .907 | .000 |
| Pair 23 | COVID-19_Hospitalised_Admitted_Age_60-69 (n) Adj_1 & COVID-19_Hospitalised_Admitted_Age_60-69 (n) Adj_3 | 123 | .992 | .000 |
| Pair 24 | COVID-19_Hospitalised_Admitted_Age_60-69 (n) Adj_2 & COVID-19_Hospitalised_Admitted_Age_60-69 (n) Adj_3 | 122 | .955 | .000 |
| Pair 25 | COVID-19_Hospitalised_Admitted_Age_70-79 (n) Adj_1 & COVID-19_Hospitalised_Admitted_Age_70-79 (n) Adj_2 | 106 | .901 | .000 |
| Pair 26 | COVID-19_Hospitalised_Admitted_Age_70-79 (n) Adj_1 & COVID-19_Hospitalised_Admitted_Age_70-79 (n) Adj_3 | 125 | .995 | .000 |
| Pair 27 | COVID-19_Hospitalised_Admitted_Age_70-79 (n) Adj_2 & COVID-19_Hospitalised_Admitted_Age_70-79 (n) Adj_3 | 123 | .944 | .000 |
| Pair 28 | COVID-19_Hospitalised_Admitted_Age_80-89 (n) Adj_1 & COVID-19_Hospitalised_Admitted_Age_80-89 (n) Adj_2 | 106 | .857 | .000 |

### Paired Samples Correlations

| | | N | Correlation | Sig. |
|---|---|---|---|---|
| Pair 29 | COVID-19_Hospitalised_Admitted_Age_80-89 (n) Adj_1 & COVID-19_Hospitalised_Admitted_Age_80-89 (n) Adj_3 | 125 | .971 | .000 |
| Pair 30 | COVID-19_Hospitalised_Admitted_Age_80-89 (n) Adj_2 & COVID-19_Hospitalised_Admitted_Age_80-89 (n) Adj_3 | 124 | .932 | .000 |

### Paired Samples Test

| | | Paired Differences | | | |
|---|---|---|---|---|---|
| | | | | | 95% Confidence ... |
| | | Mean | Std. Deviation | Std. Error Mean | Lower |
| Pair 1 | COVID-19_Hospital_Admitted_Cases Adj_1 - COVID-19_Hospital_Admitted_Cases Adj_2 | -18710.808 | 20978.136 | 2108.382 | -22894.824 |
| Pair 2 | COVID-19_Hospital_Admitted_Cases Adj_1 - COVID-19_Hospital_Admitted_Cases Adj_3 | -31503.344 | 54663.105 | 4889.217 | -41180.473 |
| Pair 3 | COVID-19_Hospital_Admitted_Cases Adj_2 - COVID-19_Hospital_Admitted_Cases Adj_3 | -58523.051 | 110805.871 | 10244.006 | -78812.597 |
| Pair 4 | COVID-19_Hospitalised_Admitted_Age_0-9 (n) Adj_1 - COVID-19_Hospitalised_Admitted_Age_0-9 (n) Adj_2 | -248.089 | 387.040 | 40.798 | -329.153 |
| Pair 5 | COVID-19_Hospitalised_Admitted_Age_0-9 (n) Adj_1 - COVID-19_Hospitalised_Admitted_Age_0-9 (n) Adj_3 | -725.964 | 906.027 | 86.386 | -897.178 |

### Paired Samples Test

| | | Paired ... 95% Confidence Interval of the ... Upper | t | df | Sig. (2-tailed) |
|---|---|---|---|---|---|
| Pair 1 | COVID-19_Hospital_Admitted_Cases Adj_1 - COVID-19_Hospital_Admitted_Cases Adj_2 | -14526.793 | -8.874 | 98 | .000 |
| Pair 2 | COVID-19_Hospital_Admitted_Cases Adj_1 - COVID-19_Hospital_Admitted_Cases Adj_3 | -21826.215 | -6.443 | 124 | .000 |
| Pair 3 | COVID-19_Hospital_Admitted_Cases Adj_2 - COVID-19_Hospital_Admitted_Cases Adj_3 | -38233.506 | -5.713 | 116 | .000 |
| Pair 4 | COVID-19_Hospitalised_Admitted_Age_0-9 (n) Adj_1 - COVID-19_Hospitalised_Admitted_Age_0-9 (n) Adj_2 | -167.025 | -6.081 | 89 | .000 |
| Pair 5 | COVID-19_Hospitalised_Admitted_Age_0-9 (n) Adj_1 - COVID-19_Hospitalised_Admitted_Age_0-9 (n) Adj_3 | -554.749 | -8.404 | 109 | .000 |

### Paired Samples Test

| | | Paired Differences | | | |
|---|---|---|---|---|---|
| | | | | | 95% Confidence ... |
| | | Mean | Std. Deviation | Std. Error Mean | Lower |
| Pair 6 | COVID-19_Hospitalised_Admitted_Age_0-9 (n) Adj_2 - COVID-19_Hospitalised_Admitted_Age_0-9 (n) Adj_3 | -842.944 | 641.288 | 57.589 | -956.938 |
| Pair 7 | COVID-19_Hospitalised_Admitted_Age_10-19 (n) Adj_1 - COVID-19_Hospitalised_Admitted_Age_10-19 (n) Adj_2 | -1999.284 | 170.841 | 18.982 | -2037.060 |
| Pair 8 | COVID-19_Hospitalised_Admitted_Age_10-19 (n) Adj_1 - COVID-19_Hospitalised_Admitted_Age_10-19 (n) Adj_3 | -602.598 | 592.414 | 57.271 | -716.143 |
| Pair 9 | COVID-19_Hospitalised_Admitted_Age_10-19 (n) Adj_2 - COVID-19_Hospitalised_Admitted_Age_10-19 (n) Adj_3 | 890.181 | 636.480 | 59.096 | 773.124 |
| Pair 10 | COVID-19_Hospitalised_Admitted_Age_20-29 (n) Adj_1 - COVID-19_Hospitalised_Admitted_Age_20-29 (n) Adj_2 | -1699.009 | 1147.038 | 111.410 | -1919.915 |
| Pair 11 | COVID-19_Hospitalised_Admitted_Age_20-29 (n) Adj_1 - COVID-19_Hospitalised_Admitted_Age_20-29 (n) Adj_3 | -1277.792 | 1779.711 | 159.182 | -1592.858 |
| Pair 12 | COVID-19_Hospitalised_Admitted_Age_20-29 (n) Adj_2 - COVID-19_Hospitalised_Admitted_Age_20-29 (n) Adj_3 | -1315.274 | 14601.290 | 1311.235 | -3910.783 |

### Paired Samples Test

| | | Paired ... 95% Confidence Interval of the ... Upper | t | df | Sig. (2-tailed) |
|---|---|---|---|---|---|
| Pair 6 | COVID-19_Hospitalised_Admitted_Age_0-9 (n) Adj_2 - COVID-19_Hospitalised_Admitted_Age_0-9 (n) Adj_3 | -728.949 | -14.637 | 123 | .000 |
| Pair 7 | COVID-19_Hospitalised_Admitted_Age_10-19 (n) Adj_1 - COVID-19_Hospitalised_Admitted_Age_10-19 (n) Adj_2 | -1961.508 | -105.323 | 80 | .000 |
| Pair 8 | COVID-19_Hospitalised_Admitted_Age_10-19 (n) Adj_1 - COVID-19_Hospitalised_Admitted_Age_10-19 (n) Adj_3 | -489.053 | -10.522 | 106 | .000 |
| Pair 9 | COVID-19_Hospitalised_Admitted_Age_10-19 (n) Adj_2 - COVID-19_Hospitalised_Admitted_Age_10-19 (n) Adj_3 | 1007.238 | 15.063 | 115 | .000 |
| Pair 10 | COVID-19_Hospitalised_Admitted_Age_20-29 (n) Adj_1 - COVID-19_Hospitalised_Admitted_Age_20-29 (n) Adj_2 | -1478.104 | -15.250 | 105 | .000 |
| Pair 11 | COVID-19_Hospitalised_Admitted_Age_20-29 (n) Adj_1 - COVID-19_Hospitalised_Admitted_Age_20-29 (n) Adj_3 | -962.726 | -8.027 | 124 | .000 |
| Pair 12 | COVID-19_Hospitalised_Admitted_Age_20-29 (n) Adj_2 - COVID-19_Hospitalised_Admitted_Age_20-29 (n) Adj_3 | 1280.234 | -1.003 | 123 | .318 |

**Paired Samples Test**

| | | Paired Differences | | | |
|---|---|---|---|---|---|
| | | | | | 95% Confidence ... |
| | | Mean | Std. Deviation | Std. Error Mean | Lower |
| Pair 13 | COVID-19_Hospitalised_Admitted_Age_30-39 (n) Adj_1 - COVID-19_Hospitalised_Admitted_Age_30-39 (n) Adj_2 | -1748.528 | 2316.210 | 224.970 | -2194.603 |
| Pair 14 | COVID-19_Hospitalised_Admitted_Age_30-39 (n) Adj_1 - COVID-19_Hospitalised_Admitted_Age_30-39 (n) Adj_3 | -1425.112 | 2742.923 | 245.334 | -1910.698 |
| Pair 15 | COVID-19_Hospitalised_Admitted_Age_30-39 (n) Adj_2 - COVID-19_Hospitalised_Admitted_Age_30-39 (n) Adj_3 | 135.355 | 1309.024 | 117.554 | -97.336 |
| Pair 16 | COVID-19_Hospitalised_Admitted_Age_40-49 (n) Adj_1 - COVID-19_Hospitalised_Admitted_Age_40-49 (n) Adj_2 | -2646.534 | 4269.136 | 560.565 | -3769.047 |
| Pair 17 | COVID-19_Hospitalised_Admitted_Age_40-49 (n) Adj_1 - COVID-19_Hospitalised_Admitted_Age_40-49 (n) Adj_3 | -3124.890 | 3853.375 | 451.003 | -4023.949 |
| Pair 18 | COVID-19_Hospitalised_Admitted_Age_40-49 (n) Adj_2 - COVID-19_Hospitalised_Admitted_Age_40-49 (n) Adj_3 | -524.112 | 1859.375 | 172.639 | -866.076 |
| Pair 19 | COVID-19_Hospitalised_Admitted_Age_50-59 (n) Adj_1 - COVID-19_Hospitalised_Admitted_Age_50-59 (n) Adj_2 | -1940.638 | 4682.619 | 456.977 | -2846.840 |

### Paired Samples Test

| | | Paired ... 95% Confidence Interval of the ... Upper | t | df | Sig. (2-tailed) |
|---|---|---|---|---|---|
| Pair 13 | COVID-19_Hospitalised_Admitted_Age_30-39 (n) Adj_1 - COVID-19_Hospitalised_Admitted_Age_30-39 (n) Adj_2 | -1302.454 | -7.772 | 105 | .000 |
| Pair 14 | COVID-19_Hospitalised_Admitted_Age_30-39 (n) Adj_1 - COVID-19_Hospitalised_Admitted_Age_30-39 (n) Adj_3 | -939.526 | -5.809 | 124 | .000 |
| Pair 15 | COVID-19_Hospitalised_Admitted_Age_30-39 (n) Adj_2 - COVID-19_Hospitalised_Admitted_Age_30-39 (n) Adj_3 | 368.045 | 1.151 | 123 | .252 |
| Pair 16 | COVID-19_Hospitalised_Admitted_Age_40-49 (n) Adj_1 - COVID-19_Hospitalised_Admitted_Age_40-49 (n) Adj_2 | -1524.022 | -4.721 | 57 | .000 |
| Pair 17 | COVID-19_Hospitalised_Admitted_Age_40-49 (n) Adj_1 - COVID-19_Hospitalised_Admitted_Age_40-49 (n) Adj_3 | -2225.831 | -6.929 | 72 | .000 |
| Pair 18 | COVID-19_Hospitalised_Admitted_Age_40-49 (n) Adj_2 - COVID-19_Hospitalised_Admitted_Age_40-49 (n) Adj_3 | -182.148 | -3.036 | 115 | .003 |
| Pair 19 | COVID-19_Hospitalised_Admitted_Age_50-59 (n) Adj_1 - COVID-19_Hospitalised_Admitted_Age_50-59 (n) Adj_2 | -1034.436 | -4.247 | 104 | .000 |

### Paired Samples Test

| | | Paired Differences | | | |
|---|---|---|---|---|---|
| | | | | | 95% Confidence ... |
| | | Mean | Std. Deviation | Std. Error Mean | Lower |
| Pair 20 | COVID-19_Hospitalised_Admitted_Age_50-59 (n) Adj_1 - COVID-19_Hospitalised_Admitted_Age_50-59 (n) Adj_3 | -4373.728 | 5262.072 | 470.654 | -5305.284 |
| Pair 21 | COVID-19_Hospitalised_Admitted_Age_50-59 (n) Adj_2 - COVID-19_Hospitalised_Admitted_Age_50-59 (n) Adj_3 | -2674.659 | 3061.745 | 276.068 | -3221.163 |
| Pair 22 | COVID-19_Hospitalised_Admitted_Age_60-69 (n) Adj_1 - COVID-19_Hospitalised_Admitted_Age_60-69 (n) Adj_2 | -3105.490 | 4584.106 | 449.509 | -3996.985 |
| Pair 23 | COVID-19_Hospitalised_Admitted_Age_60-69 (n) Adj_1 - COVID-19_Hospitalised_Admitted_Age_60-69 (n) Adj_3 | -5304.057 | 4762.118 | 429.386 | -6154.069 |
| Pair 24 | COVID-19_Hospitalised_Admitted_Age_60-69 (n) Adj_2 - COVID-19_Hospitalised_Admitted_Age_60-69 (n) Adj_3 | -2284.541 | 3018.451 | 273.278 | -2825.566 |
| Pair 25 | COVID-19_Hospitalised_Admitted_Age_70-79 (n) Adj_1 - COVID-19_Hospitalised_Admitted_Age_70-79 (n) Adj_2 | -1539.604 | 3352.954 | 325.668 | -2185.343 |
| Pair 26 | COVID-19_Hospitalised_Admitted_Age_70-79 (n) Adj_1 - COVID-19_Hospitalised_Admitted_Age_70-79 (n) Adj_3 | -4632.248 | 4057.093 | 362.877 | -5350.484 |

### Paired Samples Test

| | | Paired ... 95% Confidence Interval of the ... Upper | t | df | Sig. (2-tailed) |
|---|---|---|---|---|---|
| Pair 20 | COVID-19_Hospitalised_Admitted_Age_50-59 (n) Adj_1 - COVID-19_Hospitalised_Admitted_Age_50-59 (n) Adj_3 | -3442.172 | -9.293 | 124 | .000 |
| Pair 21 | COVID-19_Hospitalised_Admitted_Age_50-59 (n) Adj_2 - COVID-19_Hospitalised_Admitted_Age_50-59 (n) Adj_3 | -2128.154 | -9.688 | 122 | .000 |
| Pair 22 | COVID-19_Hospitalised_Admitted_Age_60-69 (n) Adj_1 - COVID-19_Hospitalised_Admitted_Age_60-69 (n) Adj_2 | -2213.996 | -6.909 | 103 | .000 |
| Pair 23 | COVID-19_Hospitalised_Admitted_Age_60-69 (n) Adj_1 - COVID-19_Hospitalised_Admitted_Age_60-69 (n) Adj_3 | -4454.045 | -12.353 | 122 | .000 |
| Pair 24 | COVID-19_Hospitalised_Admitted_Age_60-69 (n) Adj_2 - COVID-19_Hospitalised_Admitted_Age_60-69 (n) Adj_3 | -1743.516 | -8.360 | 121 | .000 |
| Pair 25 | COVID-19_Hospitalised_Admitted_Age_70-79 (n) Adj_1 - COVID-19_Hospitalised_Admitted_Age_70-79 (n) Adj_2 | -893.865 | -4.728 | 105 | .000 |
| Pair 26 | COVID-19_Hospitalised_Admitted_Age_70-79 (n) Adj_1 - COVID-19_Hospitalised_Admitted_Age_70-79 (n) Adj_3 | -3914.012 | -12.765 | 124 | .000 |

### Paired Samples Test

| | | Paired Differences | | | |
|---|---|---|---|---|---|
| | | | | | 95% Confidence ... |
| | | Mean | Std. Deviation | Std. Error Mean | Lower |
| Pair 27 | COVID-19_Hospitalised_Admitted_Age_70-79 (n) Adj_2 - COVID-19_Hospitalised_Admitted_Age_70-79 (n) Adj_3 | -3516.691 | 2338.175 | 210.826 | -3934.043 |
| Pair 28 | COVID-19_Hospitalised_Admitted_Age_80-89 (n) Adj_1 - COVID-19_Hospitalised_Admitted_Age_80-89 (n) Adj_2 | -219.745 | 1457.033 | 141.520 | -500.352 |
| Pair 29 | COVID-19_Hospitalised_Admitted_Age_80-89 (n) Adj_1 - COVID-19_Hospitalised_Admitted_Age_80-89 (n) Adj_3 | -2324.048 | 2138.343 | 191.259 | -2702.604 |
| Pair 30 | COVID-19_Hospitalised_Admitted_Age_80-89 (n) Adj_2 - COVID-19_Hospitalised_Admitted_Age_80-89 (n) Adj_3 | -2461.435 | 1508.015 | 135.424 | -2729.498 |

### Paired Samples Test

| | | Paired ... 95% Confidence Interval of the ... Upper | t | df | Sig. (2-tailed) |
|---|---|---|---|---|---|
| Pair 27 | COVID-19_Hospitalised_Admitted_Age_70-79 (n) Adj_2 - COVID-19_Hospitalised_Admitted_Age_70-79 (n) Adj_3 | -3099.340 | -16.681 | 122 | .000 |
| Pair 28 | COVID-19_Hospitalised_Admitted_Age_80-89 (n) Adj_1 - COVID-19_Hospitalised_Admitted_Age_80-89 (n) Adj_2 | 60.862 | -1.553 | 105 | .123 |
| Pair 29 | COVID-19_Hospitalised_Admitted_Age_80-89 (n) Adj_1 - COVID-19_Hospitalised_Admitted_Age_80-89 (n) Adj_3 | -1945.492 | -12.151 | 124 | .000 |
| Pair 30 | COVID-19_Hospitalised_Admitted_Age_80-89 (n) Adj_2 - COVID-19_Hospitalised_Admitted_Age_80-89 (n) Adj_3 | -2193.373 | -18.176 | 123 | .000 |

```
T-TEST PAIRS=COVID19_Hospitalised_Admitted_Age_09p_1 COVID19_Hospitalised_Admitted_Age_09
p_1
    COVID19_Hospitalised_Admitted_Age_09p_2 COVID19_Hospitalised_Admitted_Age_1019p_1
    COVID19_Hospitalised_Admitted_Age_1019p_1 COVID19_Hospitalised_Admitted_Age_1019p_2
    COVID19_Hospitalised_Admitted_Age_2029p_1 COVID19_Hospitalised_Admitted_Age_2029p_1
    COVID19_Hospitalised_Admitted_Age_2029p_2 COVID19_Hospitalised_Admitted_Age_3039p_1
    COVID19_Hospitalised_Admitted_Age_3039p_1 COVID19_Hospitalised_Admitted_Age_3039p_2
    COVID19_Hospitalised_Admitted_Age_4049p_1 COVID19_Hospitalised_Admitted_Age_4049p_1
    COVID19_Hospitalised_Admitted_Age_4049p_2 COVID19_Hospitalised_Admitted_Age_5059p_1
    COVID19_Hospitalised_Admitted_Age_5059p_1 COVID19_Hospitalised_Admitted_Age_5059p_2
    COVID19_Hospitalised_Admitted_Age_6069p_1 COVID19_Hospitalised_Admitted_Age_6069p_1
    COVID19_Hospitalised_Admitted_Age_6069p_2 COVID19_Hospitalised_Admitted_Age_7079p_1
    COVID19_Hospitalised_Admitted_Age_7079p_1 COVID19_Hospitalised_Admitted_Age_7079p_2
    COVID19_Hospitalised_Admitted_Age_8089p_1 COVID19_Hospitalised_Admitted_Age_8089p_1
    COVID19_Hospitalised_Admitted_Age_8089p_2 WITH COVID19_Hospitalised_Admitted_Age_09p_
2
    COVID19_Hospitalised_Admitted_Age_09p_3 COVID19_Hospitalised_Admitted_Age_09p_3
    COVID19_Hospitalised_Admitted_Age_1019p_2 COVID19_Hospitalised_Admitted_Age_1019p_3
    COVID19_Hospitalised_Admitted_Age_1019p_3 COVID19_Hospitalised_Admitted_Age_2029p_2
    COVID19_Hospitalised_Admitted_Age_2029p_3 COVID19_Hospitalised_Admitted_Age_2029p_3
    COVID19_Hospitalised_Admitted_Age_3039p_2 COVID19_Hospitalised_Admitted_Age_3039p_3
    COVID19_Hospitalised_Admitted_Age_3039p_3 COVID19_Hospitalised_Admitted_Age_4049p_2
    COVID19_Hospitalised_Admitted_Age_4049p_3 COVID19_Hospitalised_Admitted_Age_4049p_3
    COVID19_Hospitalised_Admitted_Age_5059p_2 COVID19_Hospitalised_Admitted_Age_5059p_3
    COVID19_Hospitalised_Admitted_Age_5059p_3 COVID19_Hospitalised_Admitted_Age_6069p_2
    COVID19_Hospitalised_Admitted_Age_6069p_3 COVID19_Hospitalised_Admitted_Age_6069p_3
    COVID19_Hospitalised_Admitted_Age_7079p_2 COVID19_Hospitalised_Admitted_Age_7079p_3
    COVID19_Hospitalised_Admitted_Age_7079p_3 COVID19_Hospitalised_Admitted_Age_8089p_2
    COVID19_Hospitalised_Admitted_Age_8089p_3 COVID19_Hospitalised_Admitted_Age_8089p_3 (
PAIRED)
  /ES DISPLAY(TRUE) STANDARDIZER(SD)
  /CRITERIA=CI(.9500)
  /MISSING=ANALYSIS.
```

### T-Test

### Notes

| | | |
|---|---|---|
| Output Created | | 24-SEP-2021 15:12:18 |
| Comments | | |
| Input | Data | C:\Users\ThaboMabuka\Google Drive\ARI Projects\Research Projects\COVID-19 in Africa\Papers\ACMRG\The Impact of SARS-CoV-2 Variants on the COVID-19 Epidemic in South Africa\Data\P2_Analysis_(COVID-19 Hospitalised Cases Admission Age Profile_M.sav |
| | Active Dataset | DataSet4 |
| | Filter | <none> |
| | Weight | <none> |
| | Split File | <none> |
| | N of Rows in Working Data File | 208 |
| Missing Value Handling | Definition of Missing | User defined missing values are treated as missing. |
| | Cases Used | Statistics for each analysis are based on the cases with no missing or out-of-range data for any variable in the analysis. |

### Notes

| | |
|---|---|
| Syntax | T-TEST PAIRS=COVID19_Hospitalised_Admitted_Age_09p_1 COVID19_Hospitalised_Admitted_Age_09p_1<br><br>COVID19_Hospitalised_Admitted_Age_09p_2 COVID19_Hospitalised_Admitted_Age_1019p_1<br><br>COVID19_Hospitalised_Admitted_Age_1019p_1 COVID19_Hospitalised_Admitted_Age_1019p_2<br><br>COVID19_Hospitalised_Admitted_Age_2029p_1 COVID19_Hospitalised_Admitted_Age_2029p_1<br><br>COVID19_Hospitalised_Admitted_Age_2029p_2 COVID19_Hospitalised_Admitted_Age_3039p_1<br><br>COVID19_Hospitalised_Admitted_Age_3039p_1 COVID19_Hospitalised_Admitted_Age_3039p_2<br><br>COVID19_Hospitalised_Admitted_Age_4049p_1 COVID19_Hospitalised_Admitted_Age_4049p_1<br><br>COVID19_Hospitalised_Admitted_Age_4049p_2 COVID19_Hospitalised_Admitted_Age_5059p_1<br><br>COVID19_Hospitalised_Admitted_Age_5059p_1 COVID19_Hospitalised_Admitted_Age_5059p_2<br><br>COVID19_Hospitalised_Admitted_Age_6069p_1 COVID19_Hospitalised_Admitted_Age_6069p_1<br><br>COVID19_Hospitalised_Admitted_Age_6069p_2 COVID19_Hospitalised_Admitted_Age_7079p_1<br><br>COVID19_Hospitalised_Admitted_Age_7079p_1 COVID19_Hospitalised_Admitted_Age_7079p_2 |

### Notes

| | | |
|---|---|---|
| Resources | Processor Time | 00:00:00,03 |
| | Elapsed Time | 00:00:00,03 |

### Paired Samples Statistics

| | | Mean | N | Std. Deviation | Std. Error Mean |
|---|---|---|---|---|---|
| Pair 1 | COVID-19_Hospitalised_Admitted_Age_0-9 (%)_1 | 2.148174496 | 83 | .4374207896 | .0480131693 |
| | COVID-19_Hospitalised_Admitted_Age_0-9 (%)_2 | 1.959495883 | 83 | .3656546364 | .0401358106 |
| Pair 2 | COVID-19_Hospitalised_Admitted_Age_0-9 (%)_1 | 2.363013936 | 108 | .5936404041 | .0571230745 |
| | COVID-19_Hospitalised_Admitted_Age_0-9 (%)_3 | 2.409192141 | 108 | .3234399558 | .0311230243 |
| Pair 3 | COVID-19_Hospitalised_Admitted_Age_0-9 (%)_2 | 1.931891453 | 115 | .3153724130 | .0294086291 |
| | COVID-19_Hospitalised_Admitted_Age_0-9 (%)_3 | 2.314371117 | 115 | .5064383772 | .0472256222 |
| Pair 4 | COVID-19_Hospitalised_Admitted_Age_10-19 (%)_1 | 1.857755765 | 81 | .3448494191 | .0383166021 |
| | COVID-19_Hospitalised_Admitted_Age_10-19 (%)_2 | 6.105045370 | 81 | 2.254924672 | .2505471858 |
| Pair 5 | COVID-19_Hospitalised_Admitted_Age_10-19 (%)_1 | 1.808745609 | 105 | .3184988281 | .0310823030 |
| | COVID-19_Hospitalised_Admitted_Age_10-19 (%)_3 | 2.456250150 | 105 | .2174350608 | .0212194892 |
| Pair 6 | COVID-19_Hospitalised_Admitted_Age_10-19 (%)_2 | 5.210069526 | 115 | 2.389709548 | .2228415644 |
| | COVID-19_Hospitalised_Admitted_Age_10-19 (%)_3 | 2.230738288 | 115 | .4891797006 | .0456162423 |

**Paired Samples Statistics**

| | | Mean | N | Std. Deviation | Std. Error Mean |
|---|---|---|---|---|---|
| Pair 7 | COVID-19_Hospitalised_Admitted_Age_20-29 (%)_1 | 7.080098996 | 99 | 1.902081192 | .1911663526 |
| | COVID-19_Hospitalised_Admitted_Age_20-29 (%)_2 | 7.740538448 | 99 | 2.415861446 | .2428032109 |
| Pair 8 | COVID-19_Hospitalised_Admitted_Age_20-29 (%)_1 | 7.541993727 | 123 | 2.586582149 | .2332242582 |
| | COVID-19_Hospitalised_Admitted_Age_20-29 (%)_3 | 6.202728392 | 123 | .9925656836 | .0894966337 |
| Pair 9 | COVID-19_Hospitalised_Admitted_Age_20-29 (%)_2 | 7.416773305 | 115 | 2.403783583 | .2241539749 |
| | COVID-19_Hospitalised_Admitted_Age_20-29 (%)_3 | 6.165141332 | 115 | 8.264686614 | .7706860006 |
| Pair 10 | COVID-19_Hospitalised_Admitted_Age_30-39 (%)_1 | 16.52576028 | 99 | 4.163113077 | .4184086071 |
| | COVID-19_Hospitalised_Admitted_Age_30-39 (%)_2 | 15.03535348 | 99 | 1.893311263 | .1902849416 |
| Pair 11 | COVID-19_Hospitalised_Admitted_Age_30-39 (%)_1 | 17.45281344 | 123 | 4.459372126 | .4020880436 |
| | COVID-19_Hospitalised_Admitted_Age_30-39 (%)_3 | 10.06234171 | 123 | 3.591312484 | .3238177416 |
| Pair 12 | COVID-19_Hospitalised_Admitted_Age_30-39 (%)_2 | 14.71956694 | 115 | 1.725085047 | .1608650101 |
| | COVID-19_Hospitalised_Admitted_Age_30-39 (%)_3 | 8.859011529 | 115 | 4.096667001 | .3820161677 |
| Pair 13 | COVID-19_Hospitalised_Admitted_Age_40-49 (%)_1 | 19.06691268 | 57 | 2.244356142 | .2972722331 |
| | COVID-19_Hospitalised_Admitted_Age_40-49 (%)_2 | 17.01002942 | 57 | 1.976341577 | .2617728367 |

### Paired Samples Statistics

| | | Mean | N | Std. Deviation | Std. Error Mean |
|---|---|---|---|---|---|
| Pair 14 | COVID-19_Hospitalised_Admitted_Age_40-49 (%)_1 | 19.74259895 | 73 | 4.501354368 | .5268436792 |
| | COVID-19_Hospitalised_Admitted_Age_40-49 (%)_3 | 14.57302221 | 73 | 1.934644627 | .2264330265 |
| Pair 15 | COVID-19_Hospitalised_Admitted_Age_40-49 (%)_2 | 17.27315926 | 115 | .3376878912 | .0314895582 |
| | COVID-19_Hospitalised_Admitted_Age_40-49 (%)_3 | 13.36513731 | 115 | 3.647526773 | .3401336254 |
| Pair 16 | COVID-19_Hospitalised_Admitted_Age_50-59 (%)_1 | 24.67383284 | 99 | 6.110993756 | .6141779813 |
| | COVID-19_Hospitalised_Admitted_Age_50-59 (%)_2 | 17.87814388 | 99 | 5.952140484 | .5982126268 |
| Pair 17 | COVID-19_Hospitalised_Admitted_Age_50-59 (%)_1 | 24.21581661 | 123 | 5.854960984 | .5279240532 |
| | COVID-19_Hospitalised_Admitted_Age_50-59 (%)_3 | 13.52102348 | 123 | 9.435685687 | .8507871266 |
| Pair 18 | COVID-19_Hospitalised_Admitted_Age_50-59 (%)_2 | 18.49705609 | 115 | 5.722339361 | .5336108968 |
| | COVID-19_Hospitalised_Admitted_Age_50-59 (%)_3 | 14.45272002 | 115 | 8.738352784 | .8148555987 |
| Pair 19 | COVID-19_Hospitalised_Admitted_Age_60-69 (%)_1 | 17.48291279 | 98 | 3.577509696 | .3613830523 |
| | COVID-19_Hospitalised_Admitted_Age_60-69 (%)_2 | 17.02993076 | 98 | 1.602129522 | .1618395214 |
| Pair 20 | COVID-19_Hospitalised_Admitted_Age_60-69 (%)_1 | 16.17147864 | 122 | 5.460430246 | .4943641260 |
| | COVID-19_Hospitalised_Admitted_Age_60-69 (%)_3 | 16.65762994 | 122 | 7.038399789 | .6372267759 |

### Paired Samples Statistics

| | | Mean | N | Std. Deviation | Std. Error Mean |
|---|---|---|---|---|---|
| Pair 21 | COVID-19_Hospitalised_Admitted_Age_60-69 (%)_2 | 17.49433220 | 115 | 1.749007680 | .1630958071 |
| | COVID-19_Hospitalised_Admitted_Age_60-69 (%)_3 | 14.23820114 | 115 | 7.481654463 | .6976678760 |
| Pair 22 | COVID-19_Hospitalised_Admitted_Age_70-79 (%)_1 | 7.598999311 | 99 | 4.529824105 | .4552644522 |
| | COVID-19_Hospitalised_Admitted_Age_70-79 (%)_2 | 8.146817804 | 99 | 3.463871585 | .3481321931 |
| Pair 23 | COVID-19_Hospitalised_Admitted_Age_70-79 (%)_1 | 7.036258766 | 123 | 4.444631632 | .4007589380 |
| | COVID-19_Hospitalised_Admitted_Age_70-79 (%)_3 | 13.83508316 | 123 | 1.529504436 | .1379107706 |
| Pair 24 | COVID-19_Hospitalised_Admitted_Age_70-79 (%)_2 | 8.786294934 | 114 | 3.438847160 | .3220775458 |
| | COVID-19_Hospitalised_Admitted_Age_70-79 (%)_3 | 12.55213760 | 114 | 3.502641280 | .3280524126 |
| Pair 25 | COVID-19_Hospitalised_Admitted_Age_80-89 (%)_1 | 6.111455526 | 98 | .7446876480 | .0752248123 |
| | COVID-19_Hospitalised_Admitted_Age_80-89 (%)_2 | 4.642589607 | 98 | .4826877916 | .0487588301 |
| Pair 26 | COVID-19_Hospitalised_Admitted_Age_80-89 (%)_1 | 5.586174860 | 122 | 1.591826676 | .1441172156 |
| | COVID-19_Hospitalised_Admitted_Age_80-89 (%)_3 | 7.017821384 | 122 | .8412355754 | .0761618904 |
| Pair 27 | COVID-19_Hospitalised_Admitted_Age_80-89 (%)_2 | 4.758372319 | 115 | .5166847924 | .0481811053 |
| | COVID-19_Hospitalised_Admitted_Age_80-89 (%)_3 | 6.472211535 | 115 | 1.850118600 | .1725244490 |

### Paired Samples Correlations

| | | N | Correlation | Sig. |
|---|---|---|---|---|
| Pair 1 | COVID-19_Hospitalised_Admitted_Age_0-9 (%)_1 & COVID-19_Hospitalised_Admitted_Age_0-9 (%)_2 | 83 | -.483 | .000 |
| Pair 2 | COVID-19_Hospitalised_Admitted_Age_0-9 (%)_1 & COVID-19_Hospitalised_Admitted_Age_0-9 (%)_3 | 108 | -.241 | .012 |
| Pair 3 | COVID-19_Hospitalised_Admitted_Age_0-9 (%)_2 & COVID-19_Hospitalised_Admitted_Age_0-9 (%)_3 | 115 | .131 | .163 |
| Pair 4 | COVID-19_Hospitalised_Admitted_Age_10-19 (%)_1 & COVID-19_Hospitalised_Admitted_Age_10-19 (%)_2 | 81 | .167 | .136 |
| Pair 5 | COVID-19_Hospitalised_Admitted_Age_10-19 (%)_1 & COVID-19_Hospitalised_Admitted_Age_10-19 (%)_3 | 105 | .029 | .766 |
| Pair 6 | COVID-19_Hospitalised_Admitted_Age_10-19 (%)_2 & COVID-19_Hospitalised_Admitted_Age_10-19 (%)_3 | 115 | .338 | .000 |
| Pair 7 | COVID-19_Hospitalised_Admitted_Age_20-29 (%)_1 & COVID-19_Hospitalised_Admitted_Age_20-29 (%)_2 | 99 | -.054 | .594 |
| Pair 8 | COVID-19_Hospitalised_Admitted_Age_20-29 (%)_1 & COVID-19_Hospitalised_Admitted_Age_20-29 (%)_3 | 123 | .196 | .029 |

**Paired Samples Correlations**

| | | N | Correlation | Sig. |
|---|---|---|---|---|
| Pair 9 | COVID-19_Hospitalised_Admitted_Age_20-29 (%)_2 & COVID-19_Hospitalised_Admitted_Age_20-29 (%)_3 | 115 | .104 | .271 |
| Pair 10 | COVID-19_Hospitalised_Admitted_Age_30-39 (%)_1 & COVID-19_Hospitalised_Admitted_Age_30-39 (%)_2 | 99 | .146 | .149 |
| Pair 11 | COVID-19_Hospitalised_Admitted_Age_30-39 (%)_1 & COVID-19_Hospitalised_Admitted_Age_30-39 (%)_3 | 123 | .246 | .006 |
| Pair 12 | COVID-19_Hospitalised_Admitted_Age_30-39 (%)_2 & COVID-19_Hospitalised_Admitted_Age_30-39 (%)_3 | 115 | -.133 | .157 |
| Pair 13 | COVID-19_Hospitalised_Admitted_Age_40-49 (%)_1 & COVID-19_Hospitalised_Admitted_Age_40-49 (%)_2 | 57 | .992 | .000 |
| Pair 14 | COVID-19_Hospitalised_Admitted_Age_40-49 (%)_1 & COVID-19_Hospitalised_Admitted_Age_40-49 (%)_3 | 73 | .004 | .971 |
| Pair 15 | COVID-19_Hospitalised_Admitted_Age_40-49 (%)_2 & COVID-19_Hospitalised_Admitted_Age_40-49 (%)_3 | 115 | .611 | .000 |
| Pair 16 | COVID-19_Hospitalised_Admitted_Age_50-59 (%)_1 & COVID-19_Hospitalised_Admitted_Age_50-59 (%)_2 | 99 | -.101 | .322 |

### Paired Samples Correlations

| | | N | Correlation | Sig. |
|---|---|---|---|---|
| Pair 17 | COVID-19_Hospitalised_Admitted_Age_50-59 (%)_1 & COVID-19_Hospitalised_Admitted_Age_50-59 (%)_3 | 123 | -.105 | .246 |
| Pair 18 | COVID-19_Hospitalised_Admitted_Age_50-59 (%)_2 & COVID-19_Hospitalised_Admitted_Age_50-59 (%)_3 | 115 | -.128 | .172 |
| Pair 19 | COVID-19_Hospitalised_Admitted_Age_60-69 (%)_1 & COVID-19_Hospitalised_Admitted_Age_60-69 (%)_2 | 98 | .208 | .040 |
| Pair 20 | COVID-19_Hospitalised_Admitted_Age_60-69 (%)_1 & COVID-19_Hospitalised_Admitted_Age_60-69 (%)_3 | 122 | .086 | .348 |
| Pair 21 | COVID-19_Hospitalised_Admitted_Age_60-69 (%)_2 & COVID-19_Hospitalised_Admitted_Age_60-69 (%)_3 | 115 | .008 | .935 |
| Pair 22 | COVID-19_Hospitalised_Admitted_Age_70-79 (%)_1 & COVID-19_Hospitalised_Admitted_Age_70-79 (%)_2 | 99 | -.544 | .000 |
| Pair 23 | COVID-19_Hospitalised_Admitted_Age_70-79 (%)_1 & COVID-19_Hospitalised_Admitted_Age_70-79 (%)_3 | 123 | .321 | .000 |
| Pair 24 | COVID-19_Hospitalised_Admitted_Age_70-79 (%)_2 & COVID-19_Hospitalised_Admitted_Age_70-79 (%)_3 | 114 | -.378 | .000 |

### Paired Samples Correlations

| | | N | Correlation | Sig. |
|---|---|---|---|---|
| Pair 25 | COVID-19_Hospitalised_Admitted_Age_80-89 (%)_1 & COVID-19_Hospitalised_Admitted_Age_80-89 (%)_2 | 98 | .187 | .065 |
| Pair 26 | COVID-19_Hospitalised_Admitted_Age_80-89 (%)_1 & COVID-19_Hospitalised_Admitted_Age_80-89 (%)_3 | 122 | .202 | .026 |
| Pair 27 | COVID-19_Hospitalised_Admitted_Age_80-89 (%)_2 & COVID-19_Hospitalised_Admitted_Age_80-89 (%)_3 | 115 | -.566 | .000 |

### Paired Samples Test

| | | Paired Differences | | | |
|---|---|---|---|---|---|
| | | | | | 95% Confidence ... |
| | | Mean | Std. Deviation | Std. Error Mean | Lower |
| Pair 1 | COVID-19_Hospitalised_Admitted_Age_0-9 (%)_1 - COVID-19_Hospitalised_Admitted_Age_0-9 (%)_2 | .1886786126 | .6924748016 | .0760089842 | .0374725299 |
| Pair 2 | COVID-19_Hospitalised_Admitted_Age_0-9 (%)_1 - COVID-19_Hospitalised_Admitted_Age_0-9 (%)_3 | -.046178205 | .7414341448 | .0713445339 | -.187610423 |
| Pair 3 | COVID-19_Hospitalised_Admitted_Age_0-9 (%)_2 - COVID-19_Hospitalised_Admitted_Age_0-9 (%)_3 | -.382479664 | .5604355519 | .0522608847 | -.486008074 |

### Paired Samples Test

| | | Paired ... 95% Confidence Interval of the ... | | | |
|---|---|---|---|---|---|
| | | Upper | t | df | Sig. (2-tailed) |
| Pair 1 | COVID-19_Hospitalised_Admitted_Age_0-9 (%)_1 - COVID-19_Hospitalised_Admitted_Age_0-9 (%)_2 | .3398846953 | 2.482 | 82 | .015 |
| Pair 2 | COVID-19_Hospitalised_Admitted_Age_0-9 (%)_1 - COVID-19_Hospitalised_Admitted_Age_0-9 (%)_3 | .0952540121 | -.647 | 107 | .519 |
| Pair 3 | COVID-19_Hospitalised_Admitted_Age_0-9 (%)_2 - COVID-19_Hospitalised_Admitted_Age_0-9 (%)_3 | -.278951254 | -7.319 | 114 | .000 |

**Paired Samples Test**

| | | Paired Differences | | | |
|---|---|---|---|---|---|
| | | | | | 95% Confidence ... |
| | | Mean | Std. Deviation | Std. Error Mean | Lower |
| Pair 4 | COVID-19_Hospitalised_Admitted_Age_10-19 (%)_1 - COVID-19_Hospitalised_Admitted_Age_10-19 (%)_2 | -4.24728961 | 2.223428477 | .2470476085 | -4.73893001 |
| Pair 5 | COVID-19_Hospitalised_Admitted_Age_10-19 (%)_1 - COVID-19_Hospitalised_Admitted_Age_10-19 (%)_3 | -.647504541 | .3803292110 | .0371163305 | -.721107617 |
| Pair 6 | COVID-19_Hospitalised_Admitted_Age_10-19 (%)_2 - COVID-19_Hospitalised_Admitted_Age_10-19 (%)_3 | 2.979331238 | 2.271412743 | .2118103305 | 2.559736605 |
| Pair 7 | COVID-19_Hospitalised_Admitted_Age_20-29 (%)_1 - COVID-19_Hospitalised_Admitted_Age_20-29 (%)_2 | -.660439452 | 3.154844842 | .3170738368 | -1.28966216 |
| Pair 8 | COVID-19_Hospitalised_Admitted_Age_20-29 (%)_1 - COVID-19_Hospitalised_Admitted_Age_20-29 (%)_3 | 1.339265335 | 2.581986019 | .2328098390 | .8783950059 |
| Pair 9 | COVID-19_Hospitalised_Admitted_Age_20-29 (%)_2 - COVID-19_Hospitalised_Admitted_Age_20-29 (%)_3 | 1.251631973 | 8.364752883 | .7800172283 | -.293576148 |
| Pair 10 | COVID-19_Hospitalised_Admitted_Age_30-39 (%)_1 - COVID-19_Hospitalised_Admitted_Age_30-39 (%)_2 | 1.490406797 | 4.313960505 | .4335693441 | .6300025448 |

### Paired Samples Test

| | | Paired ... 95% Confidence Interval of the ... Upper | t | df | Sig. (2-tailed) |
|---|---|---|---|---|---|
| Pair 4 | COVID-19_Hospitalised_Admitted_Age_10-19 (%)_1 - COVID-19_Hospitalised_Admitted_Age_10-19 (%)_2 | -3.75564920 | -17.192 | 80 | .000 |
| Pair 5 | COVID-19_Hospitalised_Admitted_Age_10-19 (%)_1 - COVID-19_Hospitalised_Admitted_Age_10-19 (%)_3 | -.573901465 | -17.445 | 104 | .000 |
| Pair 6 | COVID-19_Hospitalised_Admitted_Age_10-19 (%)_2 - COVID-19_Hospitalised_Admitted_Age_10-19 (%)_3 | 3.398925871 | 14.066 | 114 | .000 |
| Pair 7 | COVID-19_Hospitalised_Admitted_Age_20-29 (%)_1 - COVID-19_Hospitalised_Admitted_Age_20-29 (%)_2 | -.031216742 | -2.083 | 98 | .040 |
| Pair 8 | COVID-19_Hospitalised_Admitted_Age_20-29 (%)_1 - COVID-19_Hospitalised_Admitted_Age_20-29 (%)_3 | 1.800135664 | 5.753 | 122 | .000 |
| Pair 9 | COVID-19_Hospitalised_Admitted_Age_20-29 (%)_2 - COVID-19_Hospitalised_Admitted_Age_20-29 (%)_3 | 2.796840095 | 1.605 | 114 | .111 |
| Pair 10 | COVID-19_Hospitalised_Admitted_Age_30-39 (%)_1 - COVID-19_Hospitalised_Admitted_Age_30-39 (%)_2 | 2.350811050 | 3.438 | 98 | .001 |

**Paired Samples Test**

| | | Paired Differences | | | |
|---|---|---|---|---|---|
| | | | | | 95% Confidence ... |
| | | Mean | Std. Deviation | Std. Error Mean | Lower |
| Pair 11 | COVID-19_Hospitalised_Admitted_Age_30-39 (%)_1 - COVID-19_Hospitalised_Admitted_Age_30-39 (%)_3 | 7.390471727 | 4.991242298 | .4500451619 | 6.499562379 |
| Pair 12 | COVID-19_Hospitalised_Admitted_Age_30-39 (%)_2 - COVID-19_Hospitalised_Admitted_Age_30-39 (%)_3 | 5.860555410 | 4.651321655 | .4337379808 | 5.001323811 |
| Pair 13 | COVID-19_Hospitalised_Admitted_Age_40-49 (%)_1 - COVID-19_Hospitalised_Admitted_Age_40-49 (%)_2 | 2.056883260 | .3726433074 | .0493578118 | 1.958007682 |
| Pair 14 | COVID-19_Hospitalised_Admitted_Age_40-49 (%)_1 - COVID-19_Hospitalised_Admitted_Age_40-49 (%)_3 | 5.169576741 | 4.891832749 | .5725457168 | 4.028227714 |
| Pair 15 | COVID-19_Hospitalised_Admitted_Age_40-49 (%)_2 - COVID-19_Hospitalised_Admitted_Age_40-49 (%)_3 | 3.908021951 | 3.451630899 | .3218662409 | 3.270407407 |
| Pair 16 | COVID-19_Hospitalised_Admitted_Age_50-59 (%)_1 - COVID-19_Hospitalised_Admitted_Age_50-59 (%)_2 | 6.795688969 | 8.949479622 | .8994565447 | 5.010746729 |
| Pair 17 | COVID-19_Hospitalised_Admitted_Age_50-59 (%)_1 - COVID-19_Hospitalised_Admitted_Age_50-59 (%)_3 | 10.69479313 | 11.61754479 | 1.047518737 | 8.621125168 |

### Paired Samples Test

| | | Paired ... 95% Confidence Interval of the ... Upper | t | df | Sig. (2-tailed) |
|---|---|---|---|---|---|
| Pair 11 | COVID-19_Hospitalised_Admitted_Age_30-39 (%)_1 - COVID-19_Hospitalised_Admitted_Age_30-39 (%)_3 | 8.281381074 | 16.422 | 122 | .000 |
| Pair 12 | COVID-19_Hospitalised_Admitted_Age_30-39 (%)_2 - COVID-19_Hospitalised_Admitted_Age_30-39 (%)_3 | 6.719787010 | 13.512 | 114 | .000 |
| Pair 13 | COVID-19_Hospitalised_Admitted_Age_40-49 (%)_1 - COVID-19_Hospitalised_Admitted_Age_40-49 (%)_2 | 2.155758839 | 41.673 | 56 | .000 |
| Pair 14 | COVID-19_Hospitalised_Admitted_Age_40-49 (%)_1 - COVID-19_Hospitalised_Admitted_Age_40-49 (%)_3 | 6.310925768 | 9.029 | 72 | .000 |
| Pair 15 | COVID-19_Hospitalised_Admitted_Age_40-49 (%)_2 - COVID-19_Hospitalised_Admitted_Age_40-49 (%)_3 | 4.545636496 | 12.142 | 114 | .000 |
| Pair 16 | COVID-19_Hospitalised_Admitted_Age_50-59 (%)_1 - COVID-19_Hospitalised_Admitted_Age_50-59 (%)_2 | 8.580631209 | 7.555 | 98 | .000 |
| Pair 17 | COVID-19_Hospitalised_Admitted_Age_50-59 (%)_1 - COVID-19_Hospitalised_Admitted_Age_50-59 (%)_3 | 12.76846110 | 10.210 | 122 | .000 |

### Paired Samples Test

| | | Paired Differences | | | |
|---|---|---|---|---|---|
| | | | | | 95% Confidence ... |
| | | Mean | Std. Deviation | Std. Error Mean | Lower |
| Pair 18 | COVID-19_Hospitalised_Admitted_Age_50-59 (%)_2 - COVID-19_Hospitalised_Admitted_Age_50-59 (%)_3 | 4.044336071 | 11.04211042 | 1.029682106 | 2.004543749 |
| Pair 19 | COVID-19_Hospitalised_Admitted_Age_60-69 (%)_1 - COVID-19_Hospitalised_Admitted_Age_60-69 (%)_2 | .4529820289 | 3.603301740 | .3639884421 | -.269434272 |
| Pair 20 | COVID-19_Hospitalised_Admitted_Age_60-69 (%)_1 - COVID-19_Hospitalised_Admitted_Age_60-69 (%)_3 | -.486151302 | 8.530239001 | .7722915519 | -2.01510613 |
| Pair 21 | COVID-19_Hospitalised_Admitted_Age_60-69 (%)_2 - COVID-19_Hospitalised_Admitted_Age_60-69 (%)_3 | 3.256131053 | 7.670218246 | .7152515395 | 1.839223263 |
| Pair 22 | COVID-19_Hospitalised_Admitted_Age_70-79 (%)_1 - COVID-19_Hospitalised_Admitted_Age_70-79 (%)_2 | -.547818493 | 7.042491243 | .7077970013 | -1.95241861 |
| Pair 23 | COVID-19_Hospitalised_Admitted_Age_70-79 (%)_1 - COVID-19_Hospitalised_Admitted_Age_70-79 (%)_3 | -6.79882439 | 4.210493323 | .3796473977 | -7.55037434 |
| Pair 24 | COVID-19_Hospitalised_Admitted_Age_70-79 (%)_2 - COVID-19_Hospitalised_Admitted_Age_70-79 (%)_3 | -3.76584267 | 5.761257795 | .5395912308 | -4.83487021 |

**Paired Samples Test**

| | | Paired ... 95% Confidence Interval of the ... Upper | t | df | Sig. (2-tailed) |
|---|---|---|---|---|---|
| Pair 18 | COVID-19_Hospitalised_Admitted_Age_50-59 (%)_2 - COVID-19_Hospitalised_Admitted_Age_50-59 (%)_3 | 6.084128392 | 3.928 | 114 | .000 |
| Pair 19 | COVID-19_Hospitalised_Admitted_Age_60-69 (%)_1 - COVID-19_Hospitalised_Admitted_Age_60-69 (%)_2 | 1.175398329 | 1.244 | 97 | .216 |
| Pair 20 | COVID-19_Hospitalised_Admitted_Age_60-69 (%)_1 - COVID-19_Hospitalised_Admitted_Age_60-69 (%)_3 | 1.042803526 | -.629 | 121 | .530 |
| Pair 21 | COVID-19_Hospitalised_Admitted_Age_60-69 (%)_2 - COVID-19_Hospitalised_Admitted_Age_60-69 (%)_3 | 4.673038844 | 4.552 | 114 | .000 |
| Pair 22 | COVID-19_Hospitalised_Admitted_Age_70-79 (%)_1 - COVID-19_Hospitalised_Admitted_Age_70-79 (%)_2 | .8567816203 | -.774 | 98 | .441 |
| Pair 23 | COVID-19_Hospitalised_Admitted_Age_70-79 (%)_1 - COVID-19_Hospitalised_Admitted_Age_70-79 (%)_3 | -6.04727445 | -17.908 | 122 | .000 |
| Pair 24 | COVID-19_Hospitalised_Admitted_Age_70-79 (%)_2 - COVID-19_Hospitalised_Admitted_Age_70-79 (%)_3 | -2.69681512 | -6.979 | 113 | .000 |

### Paired Samples Test

| | | Paired Differences | | | |
|---|---|---|---|---|---|
| | | | | | 95% Confidence ... |
| | | Mean | Std. Deviation | Std. Error Mean | Lower |
| Pair 25 | COVID-19_Hospitalised_Admitted_Age_80-89 (%)_1 - COVID-19_Hospitalised_Admitted_Age_80-89 (%)_2 | 1.468865919 | .8081260369 | .0816330572 | 1.306846897 |
| Pair 26 | COVID-19_Hospitalised_Admitted_Age_80-89 (%)_1 - COVID-19_Hospitalised_Admitted_Age_80-89 (%)_3 | -1.43164652 | 1.643330173 | .1487801232 | -1.72619602 |
| Pair 27 | COVID-19_Hospitalised_Admitted_Age_80-89 (%)_2 - COVID-19_Hospitalised_Admitted_Age_80-89 (%)_3 | -1.71383922 | 2.184695726 | .2037239269 | -2.11741475 |

### Paired Samples Test

| | | Paired ... | | | |
|---|---|---|---|---|---|
| | | 95% Confidence Interval of the ... | | | |
| | | Upper | t | df | Sig. (2-tailed) |
| Pair 25 | COVID-19_Hospitalised_Admitted_Age_80-89 (%)_1 - COVID-19_Hospitalised_Admitted_Age_80-89 (%)_2 | 1.630884940 | 17.994 | 97 | .000 |
| Pair 26 | COVID-19_Hospitalised_Admitted_Age_80-89 (%)_1 - COVID-19_Hospitalised_Admitted_Age_80-89 (%)_3 | -1.13709703 | -9.623 | 121 | .000 |
| Pair 27 | COVID-19_Hospitalised_Admitted_Age_80-89 (%)_2 - COVID-19_Hospitalised_Admitted_Age_80-89 (%)_3 | -1.31026369 | -8.413 | 114 | .000 |

### Paired Samples Effect Sizes

| | | | Standardizer[a] | Point Estimate | 95% ... Lower |
|---|---|---|---|---|---|
| Pair 1 | COVID-19_Hospitalised_Admitted_Age_0-9 (%)_1 - COVID-19_Hospitalised_Admitted_Age_0-9 (%)_2 | Cohen's d | .6924748016 | .272 | .053 |
| | | Hedges' correction | .6956618510 | .271 | .052 |
| Pair 2 | COVID-19_Hospitalised_Admitted_Age_0-9 (%)_1 - COVID-19_Hospitalised_Admitted_Age_0-9 (%)_3 | Cohen's d | .7414341448 | -.062 | -.251 |
| | | Hedges' correction | .7440453397 | -.062 | -.250 |
| Pair 3 | COVID-19_Hospitalised_Admitted_Age_0-9 (%)_2 - COVID-19_Hospitalised_Admitted_Age_0-9 (%)_3 | Cohen's d | .5604355519 | -.682 | -.884 |
| | | Hedges' correction | .5622875515 | -.680 | -.881 |
| Pair 4 | COVID-19_Hospitalised_Admitted_Age_10-19 (%)_1 - COVID-19_Hospitalised_Admitted_Age_10-19 (%)_2 | Cohen's d | 2.223428477 | -1.910 | -2.275 |
| | | Hedges' correction | 2.233919100 | -1.901 | -2.264 |
| Pair 5 | COVID-19_Hospitalised_Admitted_Age_10-19 (%)_1 - COVID-19_Hospitalised_Admitted_Age_10-19 (%)_3 | Cohen's d | .3803292110 | -1.702 | -2.001 |
| | | Hedges' correction | .3817074931 | -1.696 | -1.993 |
| Pair 6 | COVID-19_Hospitalised_Admitted_Age_10-19 (%)_2 - COVID-19_Hospitalised_Admitted_Age_10-19 (%)_3 | Cohen's d | 2.271412743 | 1.312 | 1.060 |
| | | Hedges' correction | 2.278918790 | 1.307 | 1.057 |
| Pair 7 | COVID-19_Hospitalised_Admitted_Age_20-29 (%)_1 - COVID-19_Hospitalised_Admitted_Age_20-29 (%)_2 | Cohen's d | 3.154844842 | -.209 | -.408 |
| | | Hedges' correction | 3.166981457 | -.209 | -.406 |

### Paired Samples Effect Sizes

| | | | 95% ... |
|---|---|---|---|
| | | | Upper |
| Pair 1 | COVID-19_Hospitalised_Admitted_Age_0-9 (%)_1 - COVID-19_Hospitalised_Admitted_Age_0-9 (%)_2 | Cohen's d | .491 |
| | | Hedges' correction | .489 |
| Pair 2 | COVID-19_Hospitalised_Admitted_Age_0-9 (%)_1 - COVID-19_Hospitalised_Admitted_Age_0-9 (%)_3 | Cohen's d | .127 |
| | | Hedges' correction | .126 |
| Pair 3 | COVID-19_Hospitalised_Admitted_Age_0-9 (%)_2 - COVID-19_Hospitalised_Admitted_Age_0-9 (%)_3 | Cohen's d | -.478 |
| | | Hedges' correction | -.477 |
| Pair 4 | COVID-19_Hospitalised_Admitted_Age_10-19 (%)_1 - COVID-19_Hospitalised_Admitted_Age_10-19 (%)_2 | Cohen's d | -1.541 |
| | | Hedges' correction | -1.534 |
| Pair 5 | COVID-19_Hospitalised_Admitted_Age_10-19 (%)_1 - COVID-19_Hospitalised_Admitted_Age_10-19 (%)_3 | Cohen's d | -1.401 |
| | | Hedges' correction | -1.396 |
| Pair 6 | COVID-19_Hospitalised_Admitted_Age_10-19 (%)_2 - COVID-19_Hospitalised_Admitted_Age_10-19 (%)_3 | Cohen's d | 1.560 |
| | | Hedges' correction | 1.555 |
| Pair 7 | COVID-19_Hospitalised_Admitted_Age_20-29 (%)_1 - COVID-19_Hospitalised_Admitted_Age_20-29 (%)_2 | Cohen's d | -.010 |
| | | Hedges' correction | -.010 |

**Paired Samples Effect Sizes**

| | | | Standardizer[a] | Point Estimate | 95% ... |
|---|---|---|---|---|---|
| | | | | | Lower |
| Pair 8 | COVID-19_Hospitalised_Admitted_Age_20-29 (%)_1 - COVID-19_Hospitalised_Admitted_Age_20-29 (%)_3 | Cohen's d | 2.581986019 | .519 | .329 |
| | | Hedges' correction | 2.589956479 | .517 | .328 |
| Pair 9 | COVID-19_Hospitalised_Admitted_Age_20-29 (%)_2 - COVID-19_Hospitalised_Admitted_Age_20-29 (%)_3 | Cohen's d | 8.364752883 | .150 | -.034 |
| | | Hedges' correction | 8.392394810 | .149 | -.034 |
| Pair 10 | COVID-19_Hospitalised_Admitted_Age_30-39 (%)_1 - COVID-19_Hospitalised_Admitted_Age_30-39 (%)_2 | Cohen's d | 4.313960505 | .345 | .142 |
| | | Hedges' correction | 4.330556210 | .344 | .141 |
| Pair 11 | COVID-19_Hospitalised_Admitted_Age_30-39 (%)_1 - COVID-19_Hospitalised_Admitted_Age_30-39 (%)_3 | Cohen's d | 4.991242298 | 1.481 | 1.223 |
| | | Hedges' correction | 5.006650011 | 1.476 | 1.219 |
| Pair 12 | COVID-19_Hospitalised_Admitted_Age_30-39 (%)_2 - COVID-19_Hospitalised_Admitted_Age_30-39 (%)_3 | Cohen's d | 4.651321655 | 1.260 | 1.013 |
| | | Hedges' correction | 4.666692282 | 1.256 | 1.010 |
| Pair 13 | COVID-19_Hospitalised_Admitted_Age_40-49 (%)_1 - COVID-19_Hospitalised_Admitted_Age_40-49 (%)_2 | Cohen's d | .3726433074 | 5.520 | 4.465 |
| | | Hedges' correction | .3751621147 | 5.483 | 4.435 |
| Pair 14 | COVID-19_Hospitalised_Admitted_Age_40-49 (%)_1 - COVID-19_Hospitalised_Admitted_Age_40-49 (%)_3 | Cohen's d | 4.891832749 | 1.057 | .767 |
| | | Hedges' correction | 4.917496702 | 1.051 | .763 |

### Paired Samples Effect Sizes

| | | | 95% ... |
|---|---|---|---|
| | | | Upper |
| Pair 8 | COVID-19_Hospitalised_Admitted_Age_20-29 (%)_1 - COVID-19_Hospitalised_Admitted_Age_20-29 (%)_3 | Cohen's d | .706 |
| | | Hedges' correction | .704 |
| Pair 9 | COVID-19_Hospitalised_Admitted_Age_20-29 (%)_2 - COVID-19_Hospitalised_Admitted_Age_20-29 (%)_3 | Cohen's d | .333 |
| | | Hedges' correction | .332 |
| Pair 10 | COVID-19_Hospitalised_Admitted_Age_30-39 (%)_1 - COVID-19_Hospitalised_Admitted_Age_30-39 (%)_2 | Cohen's d | .547 |
| | | Hedges' correction | .545 |
| Pair 11 | COVID-19_Hospitalised_Admitted_Age_30-39 (%)_1 - COVID-19_Hospitalised_Admitted_Age_30-39 (%)_3 | Cohen's d | 1.735 |
| | | Hedges' correction | 1.730 |
| Pair 12 | COVID-19_Hospitalised_Admitted_Age_30-39 (%)_2 - COVID-19_Hospitalised_Admitted_Age_30-39 (%)_3 | Cohen's d | 1.504 |
| | | Hedges' correction | 1.499 |
| Pair 13 | COVID-19_Hospitalised_Admitted_Age_40-49 (%)_1 - COVID-19_Hospitalised_Admitted_Age_40-49 (%)_2 | Cohen's d | 6.569 |
| | | Hedges' correction | 6.525 |
| Pair 14 | COVID-19_Hospitalised_Admitted_Age_40-49 (%)_1 - COVID-19_Hospitalised_Admitted_Age_40-49 (%)_3 | Cohen's d | 1.341 |
| | | Hedges' correction | 1.334 |

### Paired Samples Effect Sizes

| | | | Standardizer[a] | Point Estimate | 95% ... |
| --- | --- | --- | --- | --- | --- |
| | | | | | Lower |
| Pair 15 | COVID-19_Hospitalised_Admitted_Age_40-49 (%)_2 - COVID-19_Hospitalised_Admitted_Age_40-49 (%)_3 | Cohen's d | 3.451630899 | 1.132 | .896 |
| | | Hedges' correction | 3.463037061 | 1.128 | .893 |
| Pair 16 | COVID-19_Hospitalised_Admitted_Age_50-59 (%)_1 - COVID-19_Hospitalised_Admitted_Age_50-59 (%)_2 | Cohen's d | 8.949479622 | .759 | .534 |
| | | Hedges' correction | 8.983908061 | .756 | .532 |
| Pair 17 | COVID-19_Hospitalised_Admitted_Age_50-59 (%)_1 - COVID-19_Hospitalised_Admitted_Age_50-59 (%)_3 | Cohen's d | 11.61754479 | .921 | .708 |
| | | Hedges' correction | 11.65340756 | .918 | .706 |
| Pair 18 | COVID-19_Hospitalised_Admitted_Age_50-59 (%)_2 - COVID-19_Hospitalised_Admitted_Age_50-59 (%)_3 | Cohen's d | 11.04211042 | .366 | .177 |
| | | Hedges' correction | 11.07859987 | .365 | .176 |
| Pair 19 | COVID-19_Hospitalised_Admitted_Age_60-69 (%)_1 - COVID-19_Hospitalised_Admitted_Age_60-69 (%)_2 | Cohen's d | 3.603301740 | .126 | -.073 |
| | | Hedges' correction | 3.617307235 | .125 | -.073 |
| Pair 20 | COVID-19_Hospitalised_Admitted_Age_60-69 (%)_1 - COVID-19_Hospitalised_Admitted_Age_60-69 (%)_3 | Cohen's d | 8.530239001 | -.057 | -.234 |
| | | Hedges' correction | 8.556789983 | -.057 | -.234 |
| Pair 21 | COVID-19_Hospitalised_Admitted_Age_60-69 (%)_2 - COVID-19_Hospitalised_Admitted_Age_60-69 (%)_3 | Cohen's d | 7.670218246 | .425 | .233 |
| | | Hedges' correction | 7.695565033 | .423 | .232 |

### Paired Samples Effect Sizes

| | | | 95% ... |
|---|---|---|---|
| | | | Upper |
| Pair 15 | COVID-19_Hospitalised_Admitted_Age_40-49 (%)_2 - COVID-19_Hospitalised_Admitted_Age_40-49 (%)_3 | Cohen's d | 1.365 |
| | | Hedges' correction | 1.361 |
| Pair 16 | COVID-19_Hospitalised_Admitted_Age_50-59 (%)_1 - COVID-19_Hospitalised_Admitted_Age_50-59 (%)_2 | Cohen's d | .982 |
| | | Hedges' correction | .978 |
| Pair 17 | COVID-19_Hospitalised_Admitted_Age_50-59 (%)_1 - COVID-19_Hospitalised_Admitted_Age_50-59 (%)_3 | Cohen's d | 1.130 |
| | | Hedges' correction | 1.127 |
| Pair 18 | COVID-19_Hospitalised_Admitted_Age_50-59 (%)_2 - COVID-19_Hospitalised_Admitted_Age_50-59 (%)_3 | Cohen's d | .554 |
| | | Hedges' correction | .553 |
| Pair 19 | COVID-19_Hospitalised_Admitted_Age_60-69 (%)_1 - COVID-19_Hospitalised_Admitted_Age_60-69 (%)_2 | Cohen's d | .324 |
| | | Hedges' correction | .323 |
| Pair 20 | COVID-19_Hospitalised_Admitted_Age_60-69 (%)_1 - COVID-19_Hospitalised_Admitted_Age_60-69 (%)_3 | Cohen's d | .121 |
| | | Hedges' correction | .120 |
| Pair 21 | COVID-19_Hospitalised_Admitted_Age_60-69 (%)_2 - COVID-19_Hospitalised_Admitted_Age_60-69 (%)_3 | Cohen's d | .615 |
| | | Hedges' correction | .613 |

### Paired Samples Effect Sizes

| | | | Standardizer[a] | Point Estimate | 95% ... Lower |
|---|---|---|---|---|---|
| Pair 22 | COVID-19_Hospitalised_Admitted_Age_70-79 (%)_1 - COVID-19_Hospitalised_Admitted_Age_70-79 (%)_2 | Cohen's d | 7.042491243 | -.078 | -.275 |
| | | Hedges' correction | 7.069583542 | -.077 | -.274 |
| Pair 23 | COVID-19_Hospitalised_Admitted_Age_70-79 (%)_1 - COVID-19_Hospitalised_Admitted_Age_70-79 (%)_3 | Cohen's d | 4.210493323 | -1.615 | -1.882 |
| | | Hedges' correction | 4.223490904 | -1.610 | -1.876 |
| Pair 24 | COVID-19_Hospitalised_Admitted_Age_70-79 (%)_2 - COVID-19_Hospitalised_Admitted_Age_70-79 (%)_3 | Cohen's d | 5.761257795 | -.654 | -.855 |
| | | Hedges' correction | 5.780465548 | -.651 | -.852 |
| Pair 25 | COVID-19_Hospitalised_Admitted_Age_80-89 (%)_1 - COVID-19_Hospitalised_Admitted_Age_80-89 (%)_2 | Cohen's d | .8081260369 | 1.818 | 1.493 |
| | | Hedges' correction | .8112671019 | 1.811 | 1.487 |
| Pair 26 | COVID-19_Hospitalised_Admitted_Age_80-89 (%)_1 - COVID-19_Hospitalised_Admitted_Age_80-89 (%)_3 | Cohen's d | 1.643330173 | -.871 | -1.078 |
| | | Hedges' correction | 1.648445156 | -.868 | -1.075 |
| Pair 27 | COVID-19_Hospitalised_Admitted_Age_80-89 (%)_2 - COVID-19_Hospitalised_Admitted_Age_80-89 (%)_3 | Cohen's d | 2.184695726 | -.784 | -.992 |
| | | Hedges' correction | 2.191915210 | -.782 | -.989 |

### Paired Samples Effect Sizes

| | | | 95% ... |
|---|---|---|---|
| | | | Upper |
| Pair 22 | COVID-19_Hospitalised_Admitted_Age_70-79 (%)_1 - COVID-19_Hospitalised_Admitted_Age_70-79 (%)_2 | Cohen's d | .120 |
| | | Hedges' correction | .119 |
| Pair 23 | COVID-19_Hospitalised_Admitted_Age_70-79 (%)_1 - COVID-19_Hospitalised_Admitted_Age_70-79 (%)_3 | Cohen's d | -1.345 |
| | | Hedges' correction | -1.340 |
| Pair 24 | COVID-19_Hospitalised_Admitted_Age_70-79 (%)_2 - COVID-19_Hospitalised_Admitted_Age_70-79 (%)_3 | Cohen's d | -.450 |
| | | Hedges' correction | -.449 |
| Pair 25 | COVID-19_Hospitalised_Admitted_Age_80-89 (%)_1 - COVID-19_Hospitalised_Admitted_Age_80-89 (%)_2 | Cohen's d | 2.139 |
| | | Hedges' correction | 2.131 |
| Pair 26 | COVID-19_Hospitalised_Admitted_Age_80-89 (%)_1 - COVID-19_Hospitalised_Admitted_Age_80-89 (%)_3 | Cohen's d | -.661 |
| | | Hedges' correction | -.659 |
| Pair 27 | COVID-19_Hospitalised_Admitted_Age_80-89 (%)_2 - COVID-19_Hospitalised_Admitted_Age_80-89 (%)_3 | Cohen's d | -.574 |
| | | Hedges' correction | -.572 |

a. The denominator used in estimating the effect sizes.
Cohen's d uses the sample standard deviation of the mean difference.
Hedges' correction uses the sample standard deviation of the mean difference, plus a correction factor.

```
 * Chart Builder.
GGRAPH
```

```
  /GRAPHDATASET NAME="graphdataset" VARIABLES=EpidemicWave
    MEANCI(COVID19_Hospitalised_Admitted_Age_09p, 95) MEANCI(COVID19_Hospitalised_Admitte
d_Age_1019p,
    95) MEANCI(COVID19_Hospitalised_Admitted_Age_2029p, 95)
    MEANCI(COVID19_Hospitalised_Admitted_Age_3039p, 95) MEANCI(COVID19_Hospitalised_Admit
ted_Age_4049p,
    95) MEANCI(COVID19_Hospitalised_Admitted_Age_5059p, 95)
    MEANCI(COVID19_Hospitalised_Admitted_Age_6069p, 95) MEANCI(COVID19_Hospitalised_Admit
ted_Age_7079p,
    95) MEANCI(COVID19_Hospitalised_Admitted_Age_8089p, 95) MISSING=LISTWISE REPORTMISSIN
G=NO
    TRANSFORM=VARSTOCASES(SUMMARY="#SUMMARY" INDEX="#INDEX" LOW="#LOW" HIGH="#HIGH")
  /GRAPHSPEC SOURCE=INLINE.
BEGIN GPL
  SOURCE: s=userSource(id("graphdataset"))
  DATA: EpidemicWave=col(source(s), name("EpidemicWave"), unit.category())
  DATA: SUMMARY=col(source(s), name("#SUMMARY"))
  DATA: INDEX=col(source(s), name("#INDEX"), unit.category())
  DATA: LOW=col(source(s), name("#LOW"))
  DATA: HIGH=col(source(s), name("#HIGH"))
  COORD: rect(dim(1,2), cluster(3,0))
  GUIDE: axis(dim(3), label("Epidemic Wave"))
  GUIDE: axis(dim(2), label("Mean"))
  GUIDE: legend(aesthetic(aesthetic.color.interior), label(""))
  GUIDE: text.title(label("Simple Bar Mean of COVID-19_Hospitalised_Admitted_Age_0-9 (%),
 Mean ",
    "of COVID-19_Hospitalised_Admitted_Age_10-19 (%), Mean of ",
    "COVID-19_Hospitalised_Admitted_Age_20-29 (%), Mean of ",
    "COVID-19_Hospitalised_Admitted_Age_30-39 (%), Mean of ",
    "COVID-19_Hospitalised_Admitted_Age_40-49 (%), Mean of ",
    "COVID-19_Hospitalised_Admitted_Age_50-59 (%), Mean of ",
    "COVID-19_Hospitalised_Admitted_Age_60-69 (%), Mean of ",
    "COVID-19_Hospitalised_Admitted_Age_70-79 (%), Mean of ",
    "COVID-19_Hospitalised_Admitted_Age_80-89 (%) by Epidemic Wave by INDEX"))
  GUIDE: text.footnote(label("Error Bars: 95% CI"))
  SCALE: linear(dim(2), include(0))
  SCALE: cat(aesthetic(aesthetic.color.interior), include("0", "1", "2", "3", "4", "5", "
6",
    "7", "8"))
  SCALE: cat(dim(1), include("0", "1", "2", "3", "4", "5", "6", "7", "8"))
  ELEMENT: interval(position(INDEX*SUMMARY*EpidemicWave), color.interior(INDEX),
    shape.interior(shape.square))
  ELEMENT: interval(position(region.spread.range(INDEX*(LOW+HIGH)*EpidemicWave)),
    shape.interior(shape.ibeam))
END GPL.
```

### GGraph

### Notes

| | | |
|---|---|---|
| Output Created | | 10-OCT-2021 21:54:00 |
| Comments | | |
| Input | Data | C: \Users\ThaboMabuka\GOOGLE~1\ARIPRO~1\RESEAR~1\COVID-~1\Papers\ACMRG\THEIMP~1\Data\ADMISS~2\P2_ANA~3.SAV |
| | Active Dataset | DataSet1 |
| | Filter | <none> |
| | Weight | <none> |
| | Split File | <none> |
| | N of Rows in Working Data File | 484 |

### Notes

| | |
|---|---|
| Syntax | GGRAPH<br>/GRAPHDATASET<br>NAME="graphdataset"<br>VARIABLES=EpidemicWa<br>ve<br>MEANCI<br>(COVID19_Hospitalised_A<br>dmitted_Age_09p, 95)<br>MEANCI<br>(COVID19_Hospitalised_A<br>dmitted_Age_1019p,<br>95) MEANCI<br>(COVID19_Hospitalised_A<br>dmitted_Age_2029p, 95)<br>MEANCI<br>(COVID19_Hospitalised_A<br>dmitted_Age_3039p, 95)<br>MEANCI<br>(COVID19_Hospitalised_A<br>dmitted_Age_4049p,<br>95) MEANCI<br>(COVID19_Hospitalised_A<br>dmitted_Age_5059p, 95)<br>MEANCI<br>(COVID19_Hospitalised_A<br>dmitted_Age_6069p, 95)<br>MEANCI<br>(COVID19_Hospitalised_A<br>dmitted_Age_7079p,<br>95) MEANCI<br>(COVID19_Hospitalised_A<br>dmitted_Age_8089p, 95)<br>MISSING=LISTWISE<br>REPORTMISSING=NO<br><br>TRANSFORM=VARSTOC<br>ASES(SUMMARY="<br>#SUMMARY" INDEX="<br>#INDEX" LOW="#LOW"<br>HIGH="#HIGH")<br>/GRAPHSPEC<br>SOURCE=INLINE.<br>BEGIN GPL<br>SOURCE: s=userSource<br>(id("graphdataset"))<br>DATA:<br>EpidemicWave=col(source<br>(s), name<br>("EpidemicWave"), unit.<br>category())<br>DATA: SUMMARY=col<br>(source(s), name<br>("#SUMMARY"))<br>DATA: INDEX=col<br>(source(s), name<br>("#INDEX"), unit.<br>category())<br>DATA: LOW=col(source<br>(s), name("#LOW"))<br>DATA: HIGH=col(source<br>(s), name("#HIGH"))<br>COORD: rect(dim(1,2) [illegible] |

### Notes

| | | |
|---|---|---|
| Resources | Processor Time | 00:00:01,23 |
| | Elapsed Time | 00:00:00,41 |

```
[DataSet1] C:\Users\ThaboMabuka\GOOGLE~1\ARIPRO~1\RESEAR~1\COVID--~1\Papers\ACMRG\THEIMP~1
\Data\ADMISS~2\P2_ANA~3.SAV
```

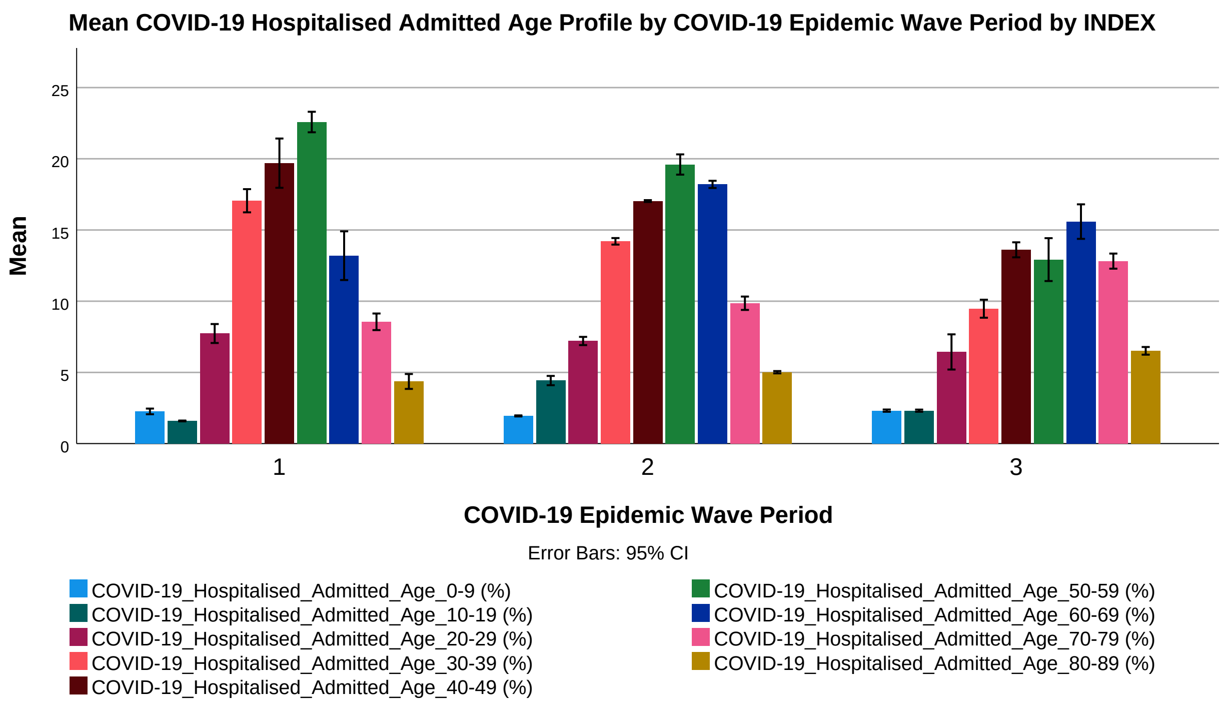
