## Supplementary material for "The Impact of SARS-CoV-2 Lineages (Variants) on the COVID-19 Epidemic in South Africa": SPSS_Program_(COVID-19 Hospitalised Cases Admission Status)

```
DESCRIPTIVES VARIABLES=COVID19_Hospital_Admitted_Cases COVID19_Hospitalised_General_Ward_
n
    COVID19_Hospitalised_High_Care_n COVID19_Hospitalised_Intensive_Care_Unit_n
    COVID19_Hospitalised_Isolation_Ward_n COVID19_Hospitalised_On_Oxygen_n
    COVID19_Hospitalised_On_Ventilator_n
  /STATISTICS=MEAN STDDEV MIN MAX.
```

### Descriptives

#### Notes

| | | |
|---|---|---|
| Output Created | | 24-SEP-2021 05:40:36 |
| Comments | | |
| Input | Data | C:\Users\ThaboMabuka\Google Drive\ARI Projects\Research Projects\COVID-19 in Africa\Papers\ACMRG\The Impact of SARS-CoV-2 Variants on the COVID-19 Epidemic in South Africa\Data\P2_Analysis_Dataset_(COVID-19 Hospitalised Cases Admission Status).sav |
| | Active Dataset | DataSet1 |
| | Filter | <none> |
| | Weight | <none> |
| | Split File | COVID-19_Epidemic_Wave |
| | N of Rows in Working Data File | 484 |
| Missing Value Handling | Definition of Missing | User defined missing values are treated as missing. |
| | Cases Used | All non-missing data are used. |

### Notes

| | | |
|---|---|---|
| Syntax | | DESCRIPTIVES VARIABLES=COVID19_Hospital_Admitted_Cases COVID19_Hospitalised_General_Ward_n<br><br>COVID19_Hospitalised_High_Care_n COVID19_Hospitalised_Intensive_Care_Unit_n<br><br>COVID19_Hospitalised_Isolation_Ward_n COVID19_Hospitalised_On_Oxygen_n<br><br>COVID19_Hospitalised_On_Ventilator_n<br>/STATISTICS=MEAN STDDEV MIN MAX. |
| Resources | Processor Time | 00:00:00,00 |
| | Elapsed Time | 00:00:00,01 |

### Descriptive Statistics

| COVID-19_Epidemic_Wave | | N | Minimum | Maximum | Mean | Std. Deviation |
|---|---|---|---|---|---|---|
| 1 | COVID-19_Hospital_Admitted_Cases | 126 | 991 | 8319 | 4823.11 | 1931.169 |
| | COVID-19_Hospitalised_General_Ward_(n) | 126 | 715 | 5745 | 3474.37 | 1315.685 |
| | COVID-19_Hospitalised_High_Care_(n) | 126 | 94 | 612 | 378.87 | 146.463 |
| | COVID-19_Hospitalised_Intensive_Care_Unit_(n) | 126 | 146 | 1520 | 833.82 | 392.593 |
| | COVID-19_Hospitalised_Isolation_Ward_(n) | 100 | 31 | 470 | 172.43 | 127.811 |
| | COVID-19_Hospitalised_On_Oxygen_(n) | 126 | 145 | 2100 | 777.34 | 326.450 |
| | COVID-19_Hospitalised_On_Ventilator_(n) | 125 | 66 | 799 | 404.66 | 221.513 |
| | Valid N (listwise) | 99 | | | | |

**Descriptive Statistics**

| COVID-19_Epidemic_Wave | | N | Minimum | Maximum | Mean | Std. Deviation |
|---|---|---|---|---|---|---|
| 2 | COVID-19_Hospital_Admitted_Cases | 187 | 3554 | 24891 | 7861.43 | 4367.278 |
| | COVID-19_Hospitalised_General_Ward_(n) | 187 | 2607 | 20501 | 6196.87 | 3460.209 |
| | COVID-19_Hospitalised_High_Care_(n) | 187 | 287 | 1359 | 558.93 | 292.349 |
| | COVID-19_Hospitalised_Intensive_Care_Unit_(n) | 187 | 506 | 3031 | 1105.63 | 642.521 |
| | COVID-19_Hospitalised_Isolation_Ward_(n) | 0 | | | | |
| | COVID-19_Hospitalised_On_Oxygen_(n) | 187 | 665 | 6700 | 2319.96 | 1614.645 |
| | COVID-19_Hospitalised_On_Ventilator_(n) | 187 | 237 | 1644 | 544.06 | 346.577 |
| | Valid N (listwise) | 0 | | | | |
| 3 | COVID-19_Hospital_Admitted_Cases | 146 | 3661 | 17560 | 10715.70 | 4561.508 |
| | COVID-19_Hospitalised_General_Ward_(n) | 146 | 2732 | 14012 | 8245.42 | 3686.501 |
| | COVID-19_Hospitalised_High_Care_(n) | 146 | 329 | 1142 | 774.85 | 258.617 |
| | COVID-19_Hospitalised_Intensive_Care_Unit_(n) | 146 | 585 | 2597 | 1695.43 | 635.211 |
| | COVID-19_Hospitalised_Isolation_Ward_(n) | 0 | | | | |
| | COVID-19_Hospitalised_On_Oxygen_(n) | 146 | 735 | 5728 | 2356.05 | 1195.342 |
| | COVID-19_Hospitalised_On_Ventilator_(n) | 146 | 307 | 1508 | 922.29 | 366.601 |
| | Valid N (listwise) | 0 | | | | |

```
DESCRIPTIVES VARIABLES=COVID19_Hospitalised_General_Ward COVID19_Hospitalised_High_Care
    COVID19_Hospitalised_Intensive_Care_Unit COVID19_Hospitalised_Isolation_Ward
    COVID19_Hospitalised_On_Oxygen COVID19_Hospitalised_On_Ventilator
  /STATISTICS=MEAN STDDEV MIN MAX.
```

### Descriptives

#### Notes

| | | |
|---|---|---|
| Output Created | | 24-SEP-2021 05:41:17 |
| Comments | | |
| Input | Data | C:\Users\ThaboMabuka\Google Drive\ARI Projects\Research Projects\COVID-19 in Africa\Papers\ACMRG\The Impact of SARS-CoV-2 Variants on the COVID-19 Epidemic in South Africa\Data\P2_Analysis_Dataset_(COVID-19 Hospitalised Cases Admission Status).sav |
| | Active Dataset | DataSet1 |
| | Filter | <none> |
| | Weight | <none> |
| | Split File | COVID-19_Epidemic_Wave |
| | N of Rows in Working Data File | 484 |
| Missing Value Handling | Definition of Missing | User defined missing values are treated as missing. |
| | Cases Used | All non-missing data are used. |

### Notes

| | | |
|---|---|---|
| Syntax | | DESCRIPTIVES VARIABLES=COVID19_Hospitalised_General_Ward COVID19_Hospitalised_High_Care<br><br>COVID19_Hospitalised_Intensive_Care_Unit COVID19_Hospitalised_Isolation_Ward<br><br>COVID19_Hospitalised_On_Oxygen COVID19_Hospitalised_On_Ventilator<br>/STATISTICS=MEAN STDDEV MIN MAX. |
| Resources | Processor Time | 00:00:00,00 |
| | Elapsed Time | 00:00:00,00 |

### Descriptive Statistics

| COVID-19_Epidemic_Wave | | N | Minimum | Maximum | Mean |
|---|---|---|---|---|---|
| 1 | COVID-19_Hospitalised_General_Ward_(%) | 125 | 68.69013559 | 78.09583074 | 72.78961470 |
| | COVID-19_Hospitalised_High_Care_(%) | 125 | 5.065856130 | 10.54107093 | 7.936821459 |
| | COVID-19_Hospitalised_Intensive_Care_Unit_(%) | 125 | 11.87683284 | 20.63121891 | 16.67675889 |
| | COVID-19_Hospitalised_Isolation_Ward_(%) | 125 | .0000000000 | 7.536882617 | 2.616858013 |
| | COVID-19_Hospitalised_On_Oxygen_(%) | 125 | 10.62044298 | 30.77108434 | 16.76696183 |
| | COVID-19_Hospitalised_On_Ventilator_(%) | 125 | 4.870335231 | 10.82089552 | 7.819128274 |
| | Valid N (listwise) | 125 | | | |

### Descriptive Statistics

| COVID-19_Epidemic_Wave | | Std. Deviation |
|---|---|---|
| 1 | COVID-19_Hospitalised_General_Ward_(%) | 2.611606832 |
| | COVID-19_Hospitalised_High_Care_(%) | 1.031071490 |
| | COVID-19_Hospitalised_Intensive_Care_Unit_(%) | 2.161813436 |
| | COVID-19_Hospitalised_Isolation_Ward_(%) | 1.812089351 |
| | COVID-19_Hospitalised_On_Oxygen_(%) | 5.781032954 |
| | COVID-19_Hospitalised_On_Ventilator_(%) | 1.790443905 |
| | Valid N (listwise) | |

### Descriptive Statistics

| COVID-19_Epidemic_Wave | | N | Minimum | Maximum | Mean |
|---|---|---|---|---|---|
| 2 | COVID-19_Hospitalised_General_Ward_(%) | 187 | 73.35396736 | 86.20907797 | 78.65586079 |
| | COVID-19_Hospitalised_High_Care_(%) | 187 | 5.329748537 | 9.431910845 | 7.274425074 |
| | COVID-19_Hospitalised_Intensive_Care_Unit_(%) | 187 | 8.461173494 | 17.64209342 | 14.06971414 |
| | COVID-19_Hospitalised_Isolation_Ward_(%) | 187 | .0000000000 | .0000000000 | .0000000000 |
| | COVID-19_Hospitalised_On_Oxygen_(%) | 187 | 18.68898187 | 38.63435864 | 27.57906718 |
| | COVID-19_Hospitalised_On_Ventilator_(%) | 187 | 4.324403029 | 9.368035657 | 6.933912439 |
| | Valid N (listwise) | 187 | | | |
| 3 | COVID-19_Hospitalised_General_Ward_(%) | 146 | 71.46054841 | 80.66104079 | 76.04201547 |
| | COVID-19_Hospitalised_High_Care_(%) | 146 | 6.130774535 | 10.14736057 | 7.649415570 |
| | COVID-19_Hospitalised_Intensive_Care_Unit_(%) | 146 | 12.86366060 | 19.49516649 | 16.30856896 |
| | COVID-19_Hospitalised_Isolation_Ward_(%) | 146 | .0000000000 | .0000000000 | .0000000000 |
| | COVID-19_Hospitalised_On_Oxygen_(%) | 146 | 17.15513442 | 34.22257440 | 21.86374389 |
| | COVID-19_Hospitalised_On_Ventilator_(%) | 146 | 7.205071991 | 10.44575725 | 8.740822626 |
| | Valid N (listwise) | 146 | | | |

### Descriptive Statistics

| COVID-19_Epidemic_Wave | | Std. Deviation |
|---|---|---|
| 2 | COVID-19_Hospitalised_General_Ward_(%) | 3.215741971 |
| | COVID-19_Hospitalised_High_Care_(%) | .9765781175 |
| | COVID-19_Hospitalised_Intensive_Care_Unit_(%) | 2.313063736 |
| | COVID-19_Hospitalised_Isolation_Ward_(%) | .0000000000 |
| | COVID-19_Hospitalised_On_Oxygen_(%) | 4.514258926 |
| | COVID-19_Hospitalised_On_Ventilator_(%) | 1.772518507 |
| | Valid N (listwise) | |
| 3 | COVID-19_Hospitalised_General_Ward_(%) | 2.477272864 |
| | COVID-19_Hospitalised_High_Care_(%) | 1.050822069 |
| | COVID-19_Hospitalised_Intensive_Care_Unit_(%) | 1.652759602 |
| | COVID-19_Hospitalised_Isolation_Ward_(%) | .0000000000 |
| | COVID-19_Hospitalised_On_Oxygen_(%) | 4.845728960 |
| | COVID-19_Hospitalised_On_Ventilator_(%) | .6241362644 |
| | Valid N (listwise) | |

```
 * Define Variable Properties.
*COVID19_Epidemic_Wave.
VARIABLE LEVEL  COVID19_Epidemic_Wave(NOMINAL).
EXECUTE.
```

```
DATASET ACTIVATE DataSet1.

SAVE OUTFILE='C:\Users\ThaboMabuka\Google Drive\ARI Projects\Research Projects\COVID-19 i
n '+
    'Africa\Papers\ACMRG\The Impact of SARS-CoV-2 Variants on the COVID-19 Epidemic in So
uth '+
    'Africa\Data\P2_Analysis_Dataset_(COVID-19 Hospitalised Cases Admission Status).sav'
  /COMPRESSED.
SPSSINC SPLIT DATASET SPLITVAR=COVID19_Epidemic_Wave
/OUTPUT DIRECTORY= "C:\Users\ThaboMabuka\Google Drive\ARI Projects\Research Projects\COVI
D-19 in "+
    "Africa\Papers\ACMRG\The Impact of SARS-CoV-2 Variants on the COVID-19 Epidemic in So
uth "+
    "Africa\Data" DELETECONTENTS=NO
/OPTIONS NAMES=VALUES.

>Warning # 10903.  Command name: AGGREGATE
>A SPLIT FILE command is in effect but will be ignored for AGGREGATE
>processing.
```

### SPSSINC SPLIT DATASET

#### Notes

| | | |
|---|---|---|
| Output Created | | 24-SEP-2021 05:42:39 |
| Comments | | |
| Input | Data | C:\Users\ThaboMabuka\Google Drive\ARI Projects\Research Projects\COVID-19 in Africa\Papers\ACMRG\The Impact of SARS-CoV-2 Variants on the COVID-19 Epidemic in South Africa\Data\P2_Analysis_Dataset_(COVID-19 Hospitalised Cases Admission Status).sav |
| | Active Dataset | DataSet1 |
| | Filter | <none> |
| | Weight | <none> |
| | Split File | COVID-19_Epidemic_Wave |
| Syntax | | BEGIN PROGRAM '# '. |
| Resources | Processor Time | 00:00:00,00 |
| | Elapsed Time | 00:00:00,02 |

```
[DataSet1] C:\Users\ThaboMabuka\Google Drive\ARI Projects\Research Projects\COVID-19 in A
frica\Papers\ACMRG\The Impact of SARS-CoV-2 Variants on the COVID-19 Epidemic in South Af
rica\Data\P2_Analysis_Dataset_(COVID-19 Hospitalised Cases Admission Status).sav
```

**Split File Information**

| | Settings and Statistics |
|---|---|
| Split Variable Names | COVID19_Epidemic_Wave |
| Output Directory | C:\Users\ThaboMabuka\Google Drive\ARI Projects\Research Projects\COVID-19 in Africa\Papers\ACMRG\The Impact of SARS-CoV-2 Variants on the COVID-19 Epidemic in South Africa\Data |
| Files Deleted | 0 |
| Files Written | 3 |
| File List | None |
| Directories Cleared | No |

#### Values and File Names for Split Files Written

| | Values or Labels | Directory | Data File |
|---|---|---|---|
| 1 | 1 | C:\Users\Thabo Mabuka\Google Drive\ARI Projects\Research Projects\COVID-19 in Africa\Papers\ACMRG\The Impact of SARS-CoV-2 Variants on the COVID-19 Epidemic in South Africa\Data | 1.sav |

### Values and File Names for Split Files Written

| | Values or Labels | Directory | Data File |
|---|---|---|---|
| 2 | 2 | C: \Users\Thabo Mabuka\Google Drive\ARI Projects\Research Projects\COVID-19 in Africa\Papers\ACMRG\The Impact of SARS-CoV-2 Variants on the COVID-19 Epidemic in South Africa\Data | 2.sav |
| 3 | 3 | C: \Users\Thabo Mabuka\Google Drive\ARI Projects\Research Projects\COVID-19 in Africa\Papers\ACMRG\The Impact of SARS-CoV-2 Variants on the COVID-19 Epidemic in South Africa\Data | 3.sav |

Based on Variables: COVID19_Epidemic_Wave

```
 DATASET ACTIVATE DataSet1.

SAVE OUTFILE='C:\Users\ThaboMabuka\Google Drive\ARI Projects\Research Projects\COVID-19 i
n '+
    'Africa\Papers\ACMRG\The Impact of SARS-CoV-2 Variants on the COVID-19 Epidemic in So
uth '+
    'Africa\Data\P2_Analysis_Dataset_(COVID-19 Hospitalised Cases Admission Status).sav'
  /COMPRESSED.
```

```
 GET
  FILE='C:\Users\ThaboMabuka\Google Drive\ARI Projects\Research Projects\COVID-19 in Afri
ca\Papers\ACMRG\The Impact of SARS-CoV-2 Variants on the COVID-19 Epidemic in South Afric
a\Data\P2_Analysis_Dataset_(COVID-19 Hospitalised Cases Admission Status)_1.sav'.
DATASET NAME DataSet1 WINDOW=FRONT.
* Define Variable Properties.
*Data_Point.
FORMATS  Data_Point(F8.0).
EXECUTE.
DATASET ACTIVATE DataSet1.

SAVE OUTFILE='C:\Users\ThaboMabuka\Google Drive\ARI Projects\Research Projects\COVID-19 i
n '+
    'Africa\Papers\ACMRG\The Impact of SARS-CoV-2 Variants on the COVID-19 Epidemic in So
uth '+
    'Africa\Data\P2_Analysis_Dataset_(COVID-19 Hospitalised Cases Admission Status)_1.sav
'
  /COMPRESSED.
DATASET ACTIVATE DataSet1.

SAVE OUTFILE='C:\Users\ThaboMabuka\Google Drive\ARI Projects\Research Projects\COVID-19 i
n '+
    'Africa\Papers\ACMRG\The Impact of SARS-CoV-2 Variants on the COVID-19 Epidemic in So
uth '+
    'Africa\Data\P2_Analysis_Dataset_(COVID-19 Hospitalised Cases Admission Status)_1.sav
'
  /COMPRESSED.
GET
  FILE='C:\Users\ThaboMabuka\Google Drive\ARI Projects\Research Projects\COVID-19 in Afri
ca\Papers\ACMRG\The Impact of SARS-CoV-2 Variants on the COVID-19 Epidemic in South Afric
a\Data\P2_Analysis_Dataset_(COVID-19 Hospitalised Cases Admission Status)_2.sav'.
DATASET NAME DataSet2 WINDOW=FRONT.
DATASET ACTIVATE DataSet2.

SAVE OUTFILE='C:\Users\ThaboMabuka\Google Drive\ARI Projects\Research Projects\COVID-19 i
n '+
    'Africa\Papers\ACMRG\The Impact of SARS-CoV-2 Variants on the COVID-19 Epidemic in So
uth '+
    'Africa\Data\P2_Analysis_Dataset_(COVID-19 Hospitalised Cases Admission Status)_2.sav
'
  /COMPRESSED.
GET
  FILE='C:\Users\ThaboMabuka\Google Drive\ARI Projects\Research Projects\COVID-19 in Afri
ca\Papers\ACMRG\The Impact of SARS-CoV-2 Variants on the COVID-19 Epidemic in South Afric
a\Data\P2_Analysis_Dataset_(COVID-19 Hospitalised Cases Admission Status)_3.sav'.
DATASET NAME DataSet3 WINDOW=FRONT.
DATASET ACTIVATE DataSet3.
```

```
SAVE OUTFILE='C:\Users\ThaboMabuka\Google Drive\ARI Projects\Research Projects\COVID-19 i
n '+
    'Africa\Papers\ACMRG\The Impact of SARS-CoV-2 Variants on the COVID-19 Epidemic in So
uth '+
    'Africa\Data\P2_Analysis_Dataset_(COVID-19 Hospitalised Cases Admission Status)_3.sav
'
  /COMPRESSED.
DATASET ACTIVATE DataSet1.
SORT CASES BY Data_Point.
DATASET ACTIVATE DataSet2.
SORT CASES BY Data_Point.
DATASET ACTIVATE DataSet1.
MATCH FILES /FILE=*
  /FILE='DataSet2'
  /BY Data_Point.
EXECUTE.
SORT CASES BY Data_Point.
DATASET ACTIVATE DataSet3.
SORT CASES BY Data_Point.
DATASET ACTIVATE DataSet1.
MATCH FILES /FILE=*
  /FILE='DataSet3'
  /BY Data_Point.
EXECUTE.

SAVE OUTFILE='C:\Users\ThaboMabuka\Google Drive\ARI Projects\Research Projects\COVID-19 i
n '+
    'Africa\Papers\ACMRG\The Impact of SARS-CoV-2 Variants on the COVID-19 Epidemic in So
uth '+
    'Africa\Data\P2_Analysis_Dataset_(COVID-19 Hospitalised Cases Admission Status)_M.sav
'
  /COMPRESSED.
DATASET ACTIVATE DataSet3.

SAVE OUTFILE='C:\Users\ThaboMabuka\Google Drive\ARI Projects\Research Projects\COVID-19 i
n '+
    'Africa\Papers\ACMRG\The Impact of SARS-CoV-2 Variants on the COVID-19 Epidemic in So
uth '+
    'Africa\Data\P2_Analysis_Dataset_(COVID-19 Hospitalised Cases Admission Status)_3.sav
'
  /COMPRESSED.
DATASET ACTIVATE DataSet1.
DATASET CLOSE DataSet3.
DATASET ACTIVATE DataSet2.
DATASET ACTIVATE DataSet2.

SAVE OUTFILE='C:\Users\ThaboMabuka\Google Drive\ARI Projects\Research Projects\COVID-19 i
n '+
```

```
    'Africa\Papers\ACMRG\The Impact of SARS-CoV-2 Variants on the COVID-19 Epidemic in So
uth '+
    'Africa\Data\P2_Analysis_Dataset_(COVID-19 Hospitalised Cases Admission Status)_2.sav
'
  /COMPRESSED.
DATASET ACTIVATE DataSet1.
DATASET CLOSE DataSet2.



 GET
  FILE='C:\Users\ThaboMabuka\Google Drive\ARI Projects\Research Projects\COVID-19 in Afri
ca\Papers\ACMRG\The Impact of SARS-CoV-2 Variants on the COVID-19 Epidemic in South Afric
a\Data\P2_Analysis_Dataset_(COVID-19 Hospitalised Cases Admission Status)_3.sav'.
DATASET NAME DataSet4 WINDOW=FRONT.
DATASET ACTIVATE DataSet4.

SAVE OUTFILE='C:\Users\ThaboMabuka\Google Drive\ARI Projects\Research Projects\COVID-19 i
n '+
    'Africa\Papers\ACMRG\The Impact of SARS-CoV-2 Variants on the COVID-19 Epidemic in So
uth '+
    'Africa\Data\P2_Analysis_Dataset_(COVID-19 Hospitalised Cases Admission Status)_3.sav
'
  /COMPRESSED.
DATASET ACTIVATE DataSet1.
SORT CASES BY Data_Point.
DATASET ACTIVATE DataSet4.
SORT CASES BY Data_Point.
DATASET ACTIVATE DataSet1.
MATCH FILES /FILE=*
  /FILE='DataSet4'
  /BY Data_Point.
EXECUTE.
DATASET ACTIVATE DataSet1.

SAVE OUTFILE='C:\Users\ThaboMabuka\Google Drive\ARI Projects\Research Projects\COVID-19 i
n '+
    'Africa\Papers\ACMRG\The Impact of SARS-CoV-2 Variants on the COVID-19 Epidemic in So
uth '+
    'Africa\Data\P2_Analysis_Dataset_(COVID-19 Hospitalised Cases Admission Status)_M.sav
'
  /COMPRESSED.
DATASET ACTIVATE DataSet4.
DATASET ACTIVATE DataSet4.

SAVE OUTFILE='C:\Users\ThaboMabuka\Google Drive\ARI Projects\Research Projects\COVID-19 i
n '+
    'Africa\Papers\ACMRG\The Impact of SARS-CoV-2 Variants on the COVID-19 Epidemic in So
uth '+
```

```
    'Africa\Data\P2_Analysis_Dataset_(COVID-19 Hospitalised Cases Admission Status)_3.sav
'
  /COMPRESSED.
DATASET ACTIVATE DataSet1.
DATASET CLOSE DataSet4.
T-TEST PAIRS=COVID19_Hospital_Admitted_Cases_1 COVID19_Hospital_Admitted_Cases_1
    COVID19_Hospital_Admitted_Cases_2 COVID19_Hospitalised_General_Ward_n_1
    COVID19_Hospitalised_General_Ward_n_1 COVID19_Hospitalised_General_Ward_n_2
    COVID19_Hospitalised_High_Care_n_1 COVID19_Hospitalised_High_Care_n_1
    COVID19_Hospitalised_High_Care_n_2 COVID19_Hospitalised_Intensive_Care_Unit_n_1
    COVID19_Hospitalised_Intensive_Care_Unit_n_1 COVID19_Hospitalised_Intensive_Care_Unit
_n_2
    COVID19_Hospitalised_On_Oxygen_n_1 COVID19_Hospitalised_On_Oxygen_n_1
    COVID19_Hospitalised_On_Oxygen_n_2 COVID19_Hospitalised_On_Ventilator_n_1
    COVID19_Hospitalised_On_Ventilator_n_1 COVID19_Hospitalised_On_Ventilator_n_2 WITH
    COVID19_Hospital_Admitted_Cases_2 COVID19_Hospital_Admitted_Cases_3
    COVID19_Hospital_Admitted_Cases_3 COVID19_Hospitalised_General_Ward_n_2
    COVID19_Hospitalised_General_Ward_n_3 COVID19_Hospitalised_General_Ward_n_3
    COVID19_Hospitalised_High_Care_n_2 COVID19_Hospitalised_High_Care_n_3
    COVID19_Hospitalised_High_Care_n_3 COVID19_Hospitalised_Intensive_Care_Unit_n_2
    COVID19_Hospitalised_Intensive_Care_Unit_n_3 COVID19_Hospitalised_Intensive_Care_Unit
_n_3
    COVID19_Hospitalised_On_Oxygen_n_2 COVID19_Hospitalised_On_Oxygen_n_3
    COVID19_Hospitalised_On_Oxygen_n_3 COVID19_Hospitalised_On_Ventilator_n_2
    COVID19_Hospitalised_On_Ventilator_n_3 COVID19_Hospitalised_On_Ventilator_n_3 (PAIRED
)
  /ES DISPLAY(TRUE) STANDARDIZER(SD)
  /CRITERIA=CI(.9500)
  /MISSING=ANALYSIS.
```

### T-Test

### Notes

| | | |
|---|---|---|
| Output Created | | 24-SEP-2021 06:02:41 |
| Comments | | |
| Input | Data | C:\Users\ThaboMabuka\Google Drive\ARI Projects\Research Projects\COVID-19 in Africa\Papers\ACMRG\The Impact of SARS-CoV-2 Variants on the COVID-19 Epidemic in South Africa\Data\P2_Analysis_Dataset_(COVID-19 Hospitalised Cases Admission Status)_M.sav |
| | Active Dataset | DataSet1 |
| | Filter | <none> |
| | Weight | <none> |
| | Split File | <none> |
| | N of Rows in Working Data File | 208 |
| Missing Value Handling | Definition of Missing | User defined missing values are treated as missing. |
| | Cases Used | Statistics for each analysis are based on the cases with no missing or out-of-range data for any variable in the analysis. |

### Notes

| | |
|---|---|
| Syntax | T-TEST PAIRS=COVID19_Hospital_Admitted_Cases_1 COVID19_Hospital_Admitted_Cases_1<br><br>COVID19_Hospital_Admitted_Cases_2 COVID19_Hospitalised_General_Ward_n_1<br><br>COVID19_Hospitalised_General_Ward_n_1 COVID19_Hospitalised_General_Ward_n_2<br><br>COVID19_Hospitalised_High_Care_n_1 COVID19_Hospitalised_High_Care_n_1<br><br>COVID19_Hospitalised_High_Care_n_2 COVID19_Hospitalised_Intensive_Care_Unit_n_1<br><br>COVID19_Hospitalised_Intensive_Care_Unit_n_1 COVID19_Hospitalised_Intensive_Care_Unit_n_2<br><br>COVID19_Hospitalised_On_Oxygen_n_1 COVID19_Hospitalised_On_Oxygen_n_1<br><br>COVID19_Hospitalised_On_Oxygen_n_2 COVID19_Hospitalised_On_Ventilator_n_1<br><br>COVID19_Hospitalised_On_Ventilator_n_1 COVID19_Hospitalised_On_Ventilator_n_2 WITH<br><br>COVID19_Hospital_Admitted_Cases_2 COVID19_Hospital_Admitted_Cases_3<br><br>COVID19_Hospital_Admitted_Cases_3 COVID19_Hospitalised_General_Ward_n_2<br><br>COVID19_Hospitalised_General_Ward_n_3 COVID19_Hospitalised_General_Ward_n_3<br><br>COVID19_Hospitalised_Hi |

### Notes

| | | |
|---|---|---|
| Resources | Processor Time | 00:00:00,00 |
| | Elapsed Time | 00:00:00,02 |

### Paired Samples Statistics

| | | Mean | N | Std. Deviation | Std. Error Mean |
|---|---|---|---|---|---|
| Pair 1 | COVID-19_Hospital_Admitted_Cases | 5262.40 | 106 | 1730.587 | 168.089 |
| | COVID-19_Hospital_Admitted_Cases | 9351.65 | 106 | 4780.117 | 464.286 |
| Pair 2 | COVID-19_Hospital_Admitted_Cases | 4823.11 | 126 | 1931.169 | 172.042 |
| | COVID-19_Hospital_Admitted_Cases | 10814.18 | 126 | 4831.872 | 430.457 |
| Pair 3 | COVID-19_Hospital_Admitted_Cases | 9453.39 | 125 | 4467.251 | 399.563 |
| | COVID-19_Hospital_Admitted_Cases | 11609.46 | 125 | 4112.418 | 367.826 |
| Pair 4 | COVID-19_Hospitalised_General_Ward_(n) | 3778.94 | 106 | 1169.316 | 113.574 |
| | COVID-19_Hospitalised_General_Ward_(n) | 7494.96 | 106 | 3715.849 | 360.915 |
| Pair 5 | COVID-19_Hospitalised_General_Ward_(n) | 3474.37 | 126 | 1315.685 | 117.211 |
| | COVID-19_Hospitalised_General_Ward_(n) | 8312.63 | 126 | 3905.594 | 347.938 |
| Pair 6 | COVID-19_Hospitalised_General_Ward_(n) | 7523.98 | 125 | 3462.869 | 309.728 |
| | COVID-19_Hospitalised_General_Ward_(n) | 8948.98 | 125 | 3344.598 | 299.150 |

### Paired Samples Statistics

| | | Mean | N | Std. Deviation | Std. Error Mean |
|---|---|---|---|---|---|
| Pair 7 | COVID-19_Hospitalised_High_Care_(n) | 413.96 | 106 | 126.465 | 12.283 |
| | COVID-19_Hospitalised_High_Care_(n) | 621.99 | 106 | 331.714 | 32.219 |
| Pair 8 | COVID-19_Hospitalised_High_Care_(n) | 378.87 | 126 | 146.463 | 13.048 |
| | COVID-19_Hospitalised_High_Care_(n) | 777.86 | 126 | 274.078 | 24.417 |
| Pair 9 | COVID-19_Hospitalised_High_Care_(n) | 646.54 | 125 | 317.108 | 28.363 |
| | COVID-19_Hospitalised_High_Care_(n) | 828.69 | 125 | 228.183 | 20.409 |
| Pair 10 | COVID-19_Hospitalised_Intensive_Care_Unit_(n) | 920.49 | 106 | 354.365 | 34.419 |
| | COVID-19_Hospitalised_Intensive_Care_Unit_(n) | 1234.70 | 106 | 749.702 | 72.818 |
| Pair 11 | COVID-19_Hospitalised_Intensive_Care_Unit_(n) | 833.82 | 126 | 392.593 | 34.975 |
| | COVID-19_Hospitalised_Intensive_Care_Unit_(n) | 1723.69 | 126 | 671.100 | 59.786 |
| Pair 12 | COVID-19_Hospitalised_Intensive_Care_Unit_(n) | 1282.87 | 125 | 712.573 | 63.734 |
| | COVID-19_Hospitalised_Intensive_Care_Unit_(n) | 1831.79 | 125 | 561.307 | 50.205 |
| Pair 13 | COVID-19_Hospitalised_On_Oxygen_(n) | 845.58 | 106 | 297.541 | 28.900 |
| | COVID-19_Hospitalised_On_Oxygen_(n) | 2851.35 | 106 | 1782.267 | 173.109 |

**Paired Samples Statistics**

| | | Mean | N | Std. Deviation | Std. Error Mean |
|---|---|---|---|---|---|
| Pair 14 | COVID-19_Hospitalised_On_Oxygen_(n) | 777.34 | 126 | 326.450 | 29.083 |
| | COVID-19_Hospitalised_On_Oxygen_(n) | 2441.07 | 126 | 1257.977 | 112.069 |
| Pair 15 | COVID-19_Hospitalised_On_Oxygen_(n) | 2894.20 | 125 | 1672.314 | 149.576 |
| | COVID-19_Hospitalised_On_Oxygen_(n) | 2549.38 | 125 | 1128.325 | 100.920 |
| Pair 16 | COVID-19_Hospitalised_On_Ventilator_(n) | 447.58 | 106 | 207.101 | 20.115 |
| | COVID-19_Hospitalised_On_Ventilator_(n) | 569.58 | 106 | 400.918 | 38.941 |
| Pair 17 | COVID-19_Hospitalised_On_Ventilator_(n) | 404.66 | 125 | 221.513 | 19.813 |
| | COVID-19_Hospitalised_On_Ventilator_(n) | 935.71 | 125 | 386.853 | 34.601 |
| Pair 18 | COVID-19_Hospitalised_On_Ventilator_(n) | 620.06 | 125 | 396.078 | 35.426 |
| | COVID-19_Hospitalised_On_Ventilator_(n) | 999.80 | 125 | 324.509 | 29.025 |

### Paired Samples Correlations

| | | N | Correlation | Sig. |
|---|---|---|---|---|
| Pair 1 | COVID-19_Hospital_Admitted_Cases & COVID-19_Hospital_Admitted_Cases | 106 | -.117 | .230 |
| Pair 2 | COVID-19_Hospital_Admitted_Cases & COVID-19_Hospital_Admitted_Cases | 126 | .726 | .000 |
| Pair 3 | COVID-19_Hospital_Admitted_Cases & COVID-19_Hospital_Admitted_Cases | 125 | .620 | .000 |
| Pair 4 | COVID-19_Hospitalised_General_Ward_(n) & COVID-19_Hospitalised_General_Ward_(n) | 106 | -.069 | .479 |
| Pair 5 | COVID-19_Hospitalised_General_Ward_(n) & COVID-19_Hospitalised_General_Ward_(n) | 126 | .724 | .000 |
| Pair 6 | COVID-19_Hospitalised_General_Ward_(n) & COVID-19_Hospitalised_General_Ward_(n) | 125 | .627 | .000 |
| Pair 7 | COVID-19_Hospitalised_High_Care_(n) & COVID-19_Hospitalised_High_Care_(n) | 106 | -.253 | .009 |
| Pair 8 | COVID-19_Hospitalised_High_Care_(n) & COVID-19_Hospitalised_High_Care_(n) | 126 | .631 | .000 |
| Pair 9 | COVID-19_Hospitalised_High_Care_(n) & COVID-19_Hospitalised_High_Care_(n) | 125 | .645 | .000 |

### Paired Samples Correlations

| | | N | Correlation | Sig. |
|---|---|---|---|---|
| Pair 10 | COVID-19_Hospitalised_Intensive_Care_Unit_(n) & COVID-19_Hospitalised_Intensive_Care_Unit_(n) | 106 | -.034 | .733 |
| Pair 11 | COVID-19_Hospitalised_Intensive_Care_Unit_(n) & COVID-19_Hospitalised_Intensive_Care_Unit_(n) | 126 | .863 | .000 |
| Pair 12 | COVID-19_Hospitalised_Intensive_Care_Unit_(n) & COVID-19_Hospitalised_Intensive_Care_Unit_(n) | 125 | .501 | .000 |
| Pair 13 | COVID-19_Hospitalised_On_Oxygen_(n) & COVID-19_Hospitalised_On_Oxygen_(n) | 106 | .211 | .030 |
| Pair 14 | COVID-19_Hospitalised_On_Oxygen_(n) & COVID-19_Hospitalised_On_Oxygen_(n) | 126 | .445 | .000 |
| Pair 15 | COVID-19_Hospitalised_On_Oxygen_(n) & COVID-19_Hospitalised_On_Oxygen_(n) | 125 | .234 | .009 |
| Pair 16 | COVID-19_Hospitalised_On_Ventilator_(n) & COVID-19_Hospitalised_On_Ventilator_(n) | 106 | -.185 | .058 |
| Pair 17 | COVID-19_Hospitalised_On_Ventilator_(n) & COVID-19_Hospitalised_On_Ventilator_(n) | 125 | .848 | .000 |
| Pair 18 | COVID-19_Hospitalised_On_Ventilator_(n) & COVID-19_Hospitalised_On_Ventilator_(n) | 125 | .327 | .000 |

### Paired Samples Test

| | | Paired Differences | | | |
|---|---|---|---|---|---|
| | | | | | 95% Confidence ... |
| | | Mean | Std. Deviation | Std. Error Mean | Lower |
| Pair 1 | COVID-19_Hospital_Admitted_Cases - COVID-19_Hospital_Admitted_Cases | -4089.255 | 5271.458 | 512.009 | -5104.474 |
| Pair 2 | COVID-19_Hospital_Admitted_Cases - COVID-19_Hospital_Admitted_Cases | -5991.071 | 3679.065 | 327.757 | -6639.743 |
| Pair 3 | COVID-19_Hospital_Admitted_Cases - COVID-19_Hospital_Admitted_Cases | -2156.072 | 3753.946 | 335.763 | -2820.641 |
| Pair 4 | COVID-19_Hospitalised_General_Ward_(n) - COVID-19_Hospitalised_General_Ward_(n) | -3716.019 | 3972.246 | 385.819 | -4481.026 |
| Pair 5 | COVID-19_Hospitalised_General_Ward_(n) - COVID-19_Hospitalised_General_Ward_(n) | -4838.262 | 3089.264 | 275.214 | -5382.944 |
| Pair 6 | COVID-19_Hospitalised_General_Ward_(n) - COVID-19_Hospitalised_General_Ward_(n) | -1425.008 | 2942.923 | 263.223 | -1946.000 |
| Pair 7 | COVID-19_Hospitalised_High_Care_(n) - COVID-19_Hospitalised_High_Care_(n) | -208.028 | 383.734 | 37.272 | -281.931 |
| Pair 8 | COVID-19_Hospitalised_High_Care_(n) - COVID-19_Hospitalised_High_Care_(n) | -398.992 | 214.231 | 19.085 | -436.764 |

### Paired Samples Test

| | | Paired ... 95% Confidence Interval of the ... | t | df | Sig. (2-tailed) |
|---|---|---|---|---|---|
| | | Upper | | | |
| Pair 1 | COVID-19_Hospital_Admitted_Cases - COVID-19_Hospital_Admitted_Cases | -3074.035 | -7.987 | 105 | .000 |
| Pair 2 | COVID-19_Hospital_Admitted_Cases - COVID-19_Hospital_Admitted_Cases | -5342.399 | -18.279 | 125 | .000 |
| Pair 3 | COVID-19_Hospital_Admitted_Cases - COVID-19_Hospital_Admitted_Cases | -1491.503 | -6.421 | 124 | .000 |
| Pair 4 | COVID-19_Hospitalised_General_Ward_(n) - COVID-19_Hospitalised_General_Ward_(n) | -2951.012 | -9.632 | 105 | .000 |
| Pair 5 | COVID-19_Hospitalised_General_Ward_(n) - COVID-19_Hospitalised_General_Ward_(n) | -4293.580 | -17.580 | 125 | .000 |
| Pair 6 | COVID-19_Hospitalised_General_Ward_(n) - COVID-19_Hospitalised_General_Ward_(n) | -904.016 | -5.414 | 124 | .000 |
| Pair 7 | COVID-19_Hospitalised_High_Care_(n) - COVID-19_Hospitalised_High_Care_(n) | -134.126 | -5.581 | 105 | .000 |
| Pair 8 | COVID-19_Hospitalised_High_Care_(n) - COVID-19_Hospitalised_High_Care_(n) | -361.220 | -20.906 | 125 | .000 |

**Paired Samples Test**

| | | Paired Differences | | | |
|---|---|---|---|---|---|
| | | | | | 95% Confidence ... |
| | | Mean | Std. Deviation | Std. Error Mean | Lower |
| Pair 9 | COVID-19_Hospitalised_High_Care_(n) - COVID-19_Hospitalised_High_Care_(n) | -182.144 | 243.475 | 21.777 | -225.247 |
| Pair 10 | COVID-19_Hospitalised_Intensive_Care_Unit_(n) - COVID-19_Hospitalised_Intensive_Care_Unit_(n) | -314.208 | 839.922 | 81.580 | -475.967 |
| Pair 11 | COVID-19_Hospitalised_Intensive_Care_Unit_(n) - COVID-19_Hospitalised_Intensive_Care_Unit_(n) | -889.873 | 386.864 | 34.465 | -958.083 |
| Pair 12 | COVID-19_Hospitalised_Intensive_Care_Unit_(n) - COVID-19_Hospitalised_Intensive_Care_Unit_(n) | -548.920 | 649.523 | 58.095 | -663.906 |
| Pair 13 | COVID-19_Hospitalised_On_Oxygen_(n) - COVID-19_Hospitalised_On_Oxygen_(n) | -2005.774 | 1743.791 | 169.372 | -2341.607 |
| Pair 14 | COVID-19_Hospitalised_On_Oxygen_(n) - COVID-19_Hospitalised_On_Oxygen_(n) | -1663.730 | 1150.631 | 102.506 | -1866.603 |
| Pair 15 | COVID-19_Hospitalised_On_Oxygen_(n) - COVID-19_Hospitalised_On_Oxygen_(n) | 344.824 | 1785.390 | 159.690 | 28.752 |
| Pair 16 | COVID-19_Hospitalised_On_Ventilator_(n) - COVID-19_Hospitalised_On_Ventilator_(n) | -122.000 | 484.039 | 47.014 | -215.220 |

**Paired Samples Test**

| | | Paired ... 95% Confidence Interval of the ... Upper | t | df | Sig. (2-tailed) |
|---|---|---|---|---|---|
| Pair 9 | COVID-19_Hospitalised_High_Care_(n) - COVID-19_Hospitalised_High_Care_(n) | -139.041 | -8.364 | 124 | .000 |
| Pair 10 | COVID-19_Hospitalised_Intensive_Care_Unit_(n) - COVID-19_Hospitalised_Intensive_Care_Unit_(n) | -152.449 | -3.852 | 105 | .000 |
| Pair 11 | COVID-19_Hospitalised_Intensive_Care_Unit_(n) - COVID-19_Hospitalised_Intensive_Care_Unit_(n) | -821.663 | -25.820 | 125 | .000 |
| Pair 12 | COVID-19_Hospitalised_Intensive_Care_Unit_(n) - COVID-19_Hospitalised_Intensive_Care_Unit_(n) | -433.934 | -9.449 | 124 | .000 |
| Pair 13 | COVID-19_Hospitalised_On_Oxygen_(n) - COVID-19_Hospitalised_On_Oxygen_(n) | -1669.940 | -11.842 | 105 | .000 |
| Pair 14 | COVID-19_Hospitalised_On_Oxygen_(n) - COVID-19_Hospitalised_On_Oxygen_(n) | -1460.857 | -16.231 | 125 | .000 |
| Pair 15 | COVID-19_Hospitalised_On_Oxygen_(n) - COVID-19_Hospitalised_On_Oxygen_(n) | 660.896 | 2.159 | 124 | .033 |
| Pair 16 | COVID-19_Hospitalised_On_Ventilator_(n) - COVID-19_Hospitalised_On_Ventilator_(n) | -28.780 | -2.595 | 105 | .011 |

### Paired Samples Test

| | | Paired Differences | | | |
|---|---|---|---|---|---|
| | | | | | 95% Confidence ... |
| | | Mean | Std. Deviation | Std. Error Mean | Lower |
| Pair 17 | COVID-19_Hospitalised_On_Ventilator_(n) - COVID-19_Hospitalised_On_Ventilator_(n) | -531.056 | 231.085 | 20.669 | -571.965 |
| Pair 18 | COVID-19_Hospitalised_On_Ventilator_(n) - COVID-19_Hospitalised_On_Ventilator_(n) | -379.736 | 421.898 | 37.736 | -454.426 |

### Paired Samples Test

| | | Paired ... | | | |
|---|---|---|---|---|---|
| | | 95% Confidence Interval of the ... | | | |
| | | Upper | t | df | Sig. (2-tailed) |
| Pair 17 | COVID-19_Hospitalised_On_Ventilator_(n) - COVID-19_Hospitalised_On_Ventilator_(n) | -490.147 | -25.694 | 124 | .000 |
| Pair 18 | COVID-19_Hospitalised_On_Ventilator_(n) - COVID-19_Hospitalised_On_Ventilator_(n) | -305.046 | -10.063 | 124 | .000 |

### Paired Samples Effect Sizes

| | | | Standardizer[a] | Point Estimate | 95% ... Lower |
|---|---|---|---|---|---|
| Pair 1 | COVID-19_Hospital_Admitted_Cases - COVID-19_Hospital_Admitted_Cases | Cohen's d | 5271.458 | -.776 | -.992 |
| | | Hedges' correction | 5290.378 | -.773 | -.988 |
| Pair 2 | COVID-19_Hospital_Admitted_Cases - COVID-19_Hospital_Admitted_Cases | Cohen's d | 3679.065 | -1.628 | -1.894 |
| | | Hedges' correction | 3690.148 | -1.624 | -1.888 |
| Pair 3 | COVID-19_Hospital_Admitted_Cases - COVID-19_Hospital_Admitted_Cases | Cohen's d | 3753.946 | -.574 | -.763 |
| | | Hedges' correction | 3765.346 | -.573 | -.760 |
| Pair 4 | COVID-19_Hospitalised_General_Ward_(n) - COVID-19_Hospitalised_General_Ward_(n) | Cohen's d | 3972.246 | -.935 | -1.162 |
| | | Hedges' correction | 3986.504 | -.932 | -1.158 |
| Pair 5 | COVID-19_Hospitalised_General_Ward_(n) - COVID-19_Hospitalised_General_Ward_(n) | Cohen's d | 3089.264 | -1.566 | -1.826 |
| | | Hedges' correction | 3098.571 | -1.561 | -1.820 |
| Pair 6 | COVID-19_Hospitalised_General_Ward_(n) - COVID-19_Hospitalised_General_Ward_(n) | Cohen's d | 2942.923 | -.484 | -.669 |
| | | Hedges' correction | 2951.861 | -.483 | -.667 |
| Pair 7 | COVID-19_Hospitalised_High_Care_(n) - COVID-19_Hospitalised_High_Care_(n) | Cohen's d | 383.734 | -.542 | -.745 |
| | | Hedges' correction | 385.112 | -.540 | -.742 |
| Pair 8 | COVID-19_Hospitalised_High_Care_(n) - COVID-19_Hospitalised_High_Care_(n) | Cohen's d | 214.231 | -1.862 | -2.150 |
| | | Hedges' correction | 214.876 | -1.857 | -2.144 |
| Pair 9 | COVID-19_Hospitalised_High_Care_(n) - COVID-19_Hospitalised_High_Care_(n) | Cohen's d | 243.475 | -.748 | -.945 |
| | | Hedges' correction | 244.214 | -.746 | -.942 |

### Paired Samples Effect Sizes

| | | | 95% ... |
|---|---|---|---|
| | | | Upper |
| Pair 1 | COVID-19_Hospital_Admitted_Cases - COVID-19_Hospital_Admitted_Cases | Cohen's d | -.557 |
| | | Hedges' correction | -.555 |
| Pair 2 | COVID-19_Hospital_Admitted_Cases - COVID-19_Hospital_Admitted_Cases | Cohen's d | -1.360 |
| | | Hedges' correction | -1.356 |
| Pair 3 | COVID-19_Hospital_Admitted_Cases - COVID-19_Hospital_Admitted_Cases | Cohen's d | -.384 |
| | | Hedges' correction | -.383 |
| Pair 4 | COVID-19_Hospitalised_General_Ward_(n) - COVID-19_Hospitalised_General_Ward_(n) | Cohen's d | -.705 |
| | | Hedges' correction | -.703 |
| Pair 5 | COVID-19_Hospitalised_General_Ward_(n) - COVID-19_Hospitalised_General_Ward_(n) | Cohen's d | -1.304 |
| | | Hedges' correction | -1.300 |
| Pair 6 | COVID-19_Hospitalised_General_Ward_(n) - COVID-19_Hospitalised_General_Ward_(n) | Cohen's d | -.298 |
| | | Hedges' correction | -.297 |
| Pair 7 | COVID-19_Hospitalised_High_Care_(n) - COVID-19_Hospitalised_High_Care_(n) | Cohen's d | -.337 |
| | | Hedges' correction | -.336 |
| Pair 8 | COVID-19_Hospitalised_High_Care_(n) - COVID-19_Hospitalised_High_Care_(n) | Cohen's d | -1.572 |
| | | Hedges' correction | -1.567 |
| Pair 9 | COVID-19_Hospitalised_High_Care_(n) - COVID-19_Hospitalised_High_Care_(n) | Cohen's d | -.548 |
| | | Hedges' correction | -.547 |

**Paired Samples Effect Sizes**

| | | | Standardizer[a] | Point Estimate | 95% ... |
|---|---|---|---|---|---|
| | | | | | Lower |
| Pair 10 | COVID-19_Hospitalised_Intensive_Care_Unit_(n) - COVID-19_Hospitalised_Intensive_Care_Unit_(n) | Cohen's d | 839.922 | -.374 | -.570 |
| | | Hedges' correction | 842.937 | -.373 | -.568 |
| Pair 11 | COVID-19_Hospitalised_Intensive_Care_Unit_(n) - COVID-19_Hospitalised_Intensive_Care_Unit_(n) | Cohen's d | 386.864 | -2.300 | -2.633 |
| | | Hedges' correction | 388.029 | -2.293 | -2.625 |
| Pair 12 | COVID-19_Hospitalised_Intensive_Care_Unit_(n) - COVID-19_Hospitalised_Intensive_Care_Unit_(n) | Cohen's d | 649.523 | -.845 | -1.048 |
| | | Hedges' correction | 651.495 | -.843 | -1.045 |
| Pair 13 | COVID-19_Hospitalised_On_Oxygen_(n) - COVID-19_Hospitalised_On_Oxygen_(n) | Cohen's d | 1743.791 | -1.150 | -1.394 |
| | | Hedges' correction | 1750.050 | -1.146 | -1.389 |
| Pair 14 | COVID-19_Hospitalised_On_Oxygen_(n) - COVID-19_Hospitalised_On_Oxygen_(n) | Cohen's d | 1150.631 | -1.446 | -1.695 |
| | | Hedges' correction | 1154.098 | -1.442 | -1.689 |
| Pair 15 | COVID-19_Hospitalised_On_Oxygen_(n) - COVID-19_Hospitalised_On_Oxygen_(n) | Cohen's d | 1785.390 | .193 | .016 |
| | | Hedges' correction | 1790.812 | .193 | .016 |
| Pair 16 | COVID-19_Hospitalised_On_Ventilator_(n) - COVID-19_Hospitalised_On_Ventilator_(n) | Cohen's d | 484.039 | -.252 | -.445 |
| | | Hedges' correction | 485.777 | -.251 | -.443 |
| Pair 17 | COVID-19_Hospitalised_On_Ventilator_(n) - COVID-19_Hospitalised_On_Ventilator_(n) | Cohen's d | 231.085 | -2.298 | -2.632 |
| | | Hedges' correction | 231.787 | -2.291 | -2.624 |

### Paired Samples Effect Sizes

| | | | 95% ... |
|---|---|---|---|
| | | | Upper |
| Pair 10 | COVID-19_Hospitalised_Intensive_Care_Unit_(n) - COVID-19_Hospitalised_Intensive_Care_Unit_(n) | Cohen's d | -.176 |
| | | Hedges' correction | -.176 |
| Pair 11 | COVID-19_Hospitalised_Intensive_Care_Unit_(n) - COVID-19_Hospitalised_Intensive_Care_Unit_(n) | Cohen's d | -1.965 |
| | | Hedges' correction | -1.959 |
| Pair 12 | COVID-19_Hospitalised_Intensive_Care_Unit_(n) - COVID-19_Hospitalised_Intensive_Care_Unit_(n) | Cohen's d | -.639 |
| | | Hedges' correction | -.638 |
| Pair 13 | COVID-19_Hospitalised_On_Oxygen_(n) - COVID-19_Hospitalised_On_Oxygen_(n) | Cohen's d | -.903 |
| | | Hedges' correction | -.900 |
| Pair 14 | COVID-19_Hospitalised_On_Oxygen_(n) - COVID-19_Hospitalised_On_Oxygen_(n) | Cohen's d | -1.194 |
| | | Hedges' correction | -1.191 |
| Pair 15 | COVID-19_Hospitalised_On_Oxygen_(n) - COVID-19_Hospitalised_On_Oxygen_(n) | Cohen's d | .370 |
| | | Hedges' correction | .369 |
| Pair 16 | COVID-19_Hospitalised_On_Ventilator_(n) - COVID-19_Hospitalised_On_Ventilator_(n) | Cohen's d | -.058 |
| | | Hedges' correction | -.058 |
| Pair 17 | COVID-19_Hospitalised_On_Ventilator_(n) - COVID-19_Hospitalised_On_Ventilator_(n) | Cohen's d | -1.961 |
| | | Hedges' correction | -1.956 |

### Paired Samples Effect Sizes

| | | | Standardizer[a] | Point Estimate | 95% ... Lower |
|---|---|---|---|---|---|
| Pair 18 | COVID-19_Hospitalised_On_Ventilator_(n) - COVID-19_Hospitalised_On_Ventilator_(n) | Cohen's d | 421.898 | -.900 | -1.107 |
| | | Hedges' correction | 423.179 | -.897 | -1.103 |

### Paired Samples Effect Sizes

| | | | 95% ... Upper |
|---|---|---|---|
| Pair 18 | COVID-19_Hospitalised_On_Ventilator_(n) - COVID-19_Hospitalised_On_Ventilator_(n) | Cohen's d | -.691 |
| | | Hedges' correction | -.689 |

a. The denominator used in estimating the effect sizes.
Cohen's d uses the sample standard deviation of the mean difference.
Hedges' correction uses the sample standard deviation of the mean difference, plus a correction factor.

```
T-TEST PAIRS=COVID19_Hospitalised_General_Ward_1_1 COVID19_Hospitalised_General_Ward_1_1
   COVID19_Hospitalised_General_Ward_2 COVID19_Hospitalised_High_Care_1
   COVID19_Hospitalised_High_Care_1 COVID19_Hospitalised_High_Care_2
   COVID19_Hospitalised_Intensive_Care_Unit_1 COVID19_Hospitalised_Intensive_Care_Unit_1
   COVID19_Hospitalised_Intensive_Care_Unit_2 COVID19_Hospitalised_On_Oxygen_1
   COVID19_Hospitalised_On_Oxygen_1 COVID19_Hospitalised_On_Oxygen_2
   COVID19_Hospitalised_On_Ventilator_1 COVID19_Hospitalised_On_Ventilator_1
   COVID19_Hospitalised_On_Ventilator_2 WITH COVID19_Hospitalised_General_Ward_2
   COVID19_Hospitalised_General_Ward_3 COVID19_Hospitalised_General_Ward_3
   COVID19_Hospitalised_High_Care_2 COVID19_Hospitalised_High_Care_3 COVID19_Hospitalise
d_High_Care_3
   COVID19_Hospitalised_Intensive_Care_Unit_2 COVID19_Hospitalised_Intensive_Care_Unit_3
   COVID19_Hospitalised_Intensive_Care_Unit_3 COVID19_Hospitalised_On_Oxygen_2
   COVID19_Hospitalised_On_Oxygen_3 COVID19_Hospitalised_On_Oxygen_3
   COVID19_Hospitalised_On_Ventilator_2 COVID19_Hospitalised_On_Ventilator_3
   COVID19_Hospitalised_On_Ventilator_3 (PAIRED)
  /ES DISPLAY(TRUE) STANDARDIZER(SD)
  /CRITERIA=CI(.9500)
  /MISSING=ANALYSIS.
```

#### T-Test

### Notes

| | | |
|---|---|---|
| Output Created | | 24-SEP-2021 06:08:22 |
| Comments | | |
| Input | Data | C:\Users\ThaboMabuka\Google Drive\ARI Projects\Research Projects\COVID-19 in Africa\Papers\ACMRG\The Impact of SARS-CoV-2 Variants on the COVID-19 Epidemic in South Africa\Data\P2_Analysis_Dataset_(COVID-19 Hospitalised Cases Admission Status)_M.sav |
| | Active Dataset | DataSet1 |
| | Filter | <none> |
| | Weight | <none> |
| | Split File | <none> |
| | N of Rows in Working Data File | 208 |
| Missing Value Handling | Definition of Missing | User defined missing values are treated as missing. |
| | Cases Used | Statistics for each analysis are based on the cases with no missing or out-of-range data for any variable in the analysis. |

### Notes

| | |
|---|---|
| Syntax | T-TEST PAIRS=COVID19_Hospitalised_General_Ward_1_1 COVID19_Hospitalised_General_Ward_1_1<br><br>COVID19_Hospitalised_General_Ward_2 COVID19_Hospitalised_High_Care_1<br><br>COVID19_Hospitalised_High_Care_1 COVID19_Hospitalised_High_Care_2<br><br>COVID19_Hospitalised_Intensive_Care_Unit_1 COVID19_Hospitalised_Intensive_Care_Unit_1<br><br>COVID19_Hospitalised_Intensive_Care_Unit_2 COVID19_Hospitalised_On_Oxygen_1<br><br>COVID19_Hospitalised_On_Oxygen_1 COVID19_Hospitalised_On_Oxygen_2<br><br>COVID19_Hospitalised_On_Ventilator_1 COVID19_Hospitalised_On_Ventilator_1<br><br>COVID19_Hospitalised_On_Ventilator_2 WITH COVID19_Hospitalised_General_Ward_2<br><br>COVID19_Hospitalised_General_Ward_3 COVID19_Hospitalised_General_Ward_3<br><br>COVID19_Hospitalised_High_Care_2 COVID19_Hospitalised_High_Care_3 COVID19_Hospitalised_High_Care_3<br><br>COVID19_Hospitalised_Intensive_Care_Unit_2 COVID19_Hospitalised_Intensive_Care_Unit_3<br><br>COVID19_Hospitalised_Intensive_Care_Unit_3 COVID19_Hospitalised_On_Oxygen_2 |

### Notes

| | | |
|---|---|---|
| Resources | Processor Time | 00:00:00,05 |
| | Elapsed Time | 00:00:00,02 |

### Paired Samples Statistics

| | | Mean | N | Std. Deviation | Std. Error Mean |
|---|---|---|---|---|---|
| Pair 1 | COVID-19_Hospitalised_General_Ward_(%) | 72.38300053 | 106 | 2.498224444 | .2426490084 |
| | COVID-19_Hospitalised_General_Ward_(%) | 80.89483037 | 106 | 2.249772988 | .2185172697 |
| Pair 2 | COVID-19_Hospitalised_General_Ward_(%) | 72.78961470 | 125 | 2.611606832 | .2335892163 |
| | COVID-19_Hospitalised_General_Ward_(%) | 75.87899040 | 125 | 2.551615795 | .2282234548 |
| Pair 3 | COVID-19_Hospitalised_General_Ward_(%) | 80.28816300 | 125 | 2.582231191 | .2309617791 |
| | COVID-19_Hospitalised_General_Ward_(%) | 76.36833848 | 125 | 2.403017401 | .2149324104 |
| Pair 4 | COVID-19_Hospitalised_High_Care_(%) | 7.994293139 | 106 | 1.019082256 | .0989820188 |
| | COVID-19_Hospitalised_High_Care_(%) | 6.569416777 | 106 | .5666795793 | .0550407864 |
| Pair 5 | COVID-19_Hospitalised_High_Care_(%) | 7.936821459 | 125 | 1.031071490 | .0922218376 |
| | COVID-19_Hospitalised_High_Care_(%) | 7.641151590 | 125 | 1.095833752 | .0980143505 |
| Pair 6 | COVID-19_Hospitalised_High_Care_(%) | 6.752847721 | 125 | .6947844159 | .0621434073 |
| | COVID-19_Hospitalised_High_Care_(%) | 7.447045287 | 125 | .9447656215 | .0845024061 |

### Paired Samples Statistics

| | | Mean | N | Std. Deviation | Std. Error Mean |
|---|---|---|---|---|---|
| Pair 7 | COVID-19_Hospitalised_Intensive_Care_Unit_(%) | 17.07467350 | 106 | 2.014260494 | .1956422741 |
| | COVID-19_Hospitalised_Intensive_Care_Unit_(%) | 12.53575285 | 106 | 1.788139213 | .1736794338 |
| Pair 8 | COVID-19_Hospitalised_Intensive_Care_Unit_(%) | 16.67675889 | 125 | 2.161813436 | .1933584719 |
| | COVID-19_Hospitalised_Intensive_Care_Unit_(%) | 16.47985801 | 125 | 1.679371053 | .1502075134 |
| Pair 9 | COVID-19_Hospitalised_Intensive_Care_Unit_(%) | 12.95898927 | 125 | 1.973907673 | .1765516695 |
| | COVID-19_Hospitalised_Intensive_Care_Unit_(%) | 16.18461623 | 125 | 1.682122263 | .1504535891 |
| Pair 10 | COVID-19_Hospitalised_On_Oxygen_(%) | 16.81655348 | 106 | 6.141809635 | .5965452868 |
| | COVID-19_Hospitalised_On_Oxygen_(%) | 28.87142697 | 106 | 4.209337659 | .4088470158 |
| Pair 11 | COVID-19_Hospitalised_On_Oxygen_(%) | 16.76696183 | 125 | 5.781032954 | .5170713066 |
| | COVID-19_Hospitalised_On_Oxygen_(%) | 22.47077214 | 125 | 4.968953604 | .4444367214 |
| Pair 12 | COVID-19_Hospitalised_On_Oxygen_(%) | 29.17949359 | 125 | 4.053927114 | .3625942642 |
| | COVID-19_Hospitalised_On_Oxygen_(%) | 21.87019881 | 125 | 5.084207977 | .4547453859 |
| Pair 13 | COVID-19_Hospitalised_On_Ventilator_(%) | 8.116632834 | 106 | 1.721676320 | .1672239869 |
| | COVID-19_Hospitalised_On_Ventilator_(%) | 5.694133926 | 106 | 1.322056436 | .1284094725 |

### Paired Samples Statistics

| | | Mean | N | Std. Deviation | Std. Error Mean |
|---|---|---|---|---|---|
| Pair 14 | COVID-19_Hospitalised_On_Ventilator_(%) | 7.819128274 | 125 | 1.790443905 | .1601421712 |
| | COVID-19_Hospitalised_On_Ventilator_(%) | 8.756973849 | 125 | .6586055370 | .0589074700 |
| Pair 15 | COVID-19_Hospitalised_On_Ventilator_(%) | 6.179555424 | 125 | 1.707527365 | .1527258905 |
| | COVID-19_Hospitalised_On_Ventilator_(%) | 8.743013947 | 125 | .6563689175 | .0587074207 |

### Paired Samples Correlations

| | | N | Correlation | Sig. |
|---|---|---|---|---|
| Pair 1 | COVID-19_Hospitalised_General_Ward_(%) & COVID-19_Hospitalised_General_Ward_(%) | 106 | -.394 | .000 |
| Pair 2 | COVID-19_Hospitalised_General_Ward_(%) & COVID-19_Hospitalised_General_Ward_(%) | 125 | -.084 | .354 |
| Pair 3 | COVID-19_Hospitalised_General_Ward_(%) & COVID-19_Hospitalised_General_Ward_(%) | 125 | -.697 | .000 |
| Pair 4 | COVID-19_Hospitalised_High_Care_(%) & COVID-19_Hospitalised_High_Care_(%) | 106 | -.084 | .393 |
| Pair 5 | COVID-19_Hospitalised_High_Care_(%) & COVID-19_Hospitalised_High_Care_(%) | 125 | .508 | .000 |
| Pair 6 | COVID-19_Hospitalised_High_Care_(%) & COVID-19_Hospitalised_High_Care_(%) | 125 | -.207 | .020 |

### Paired Samples Correlations

| | | N | Correlation | Sig. |
|---|---|---|---|---|
| Pair 7 | COVID-19_Hospitalised_Intensive_Care_Unit_(%) & COVID-19_Hospitalised_Intensive_Care_Unit_(%) | 106 | .336 | .000 |
| Pair 8 | COVID-19_Hospitalised_Intensive_Care_Unit_(%) & COVID-19_Hospitalised_Intensive_Care_Unit_(%) | 125 | -.526 | .000 |
| Pair 9 | COVID-19_Hospitalised_Intensive_Care_Unit_(%) & COVID-19_Hospitalised_Intensive_Care_Unit_(%) | 125 | -.796 | .000 |
| Pair 10 | COVID-19_Hospitalised_On_Oxygen_(%) & COVID-19_Hospitalised_On_Oxygen_(%) | 106 | .370 | .000 |
| Pair 11 | COVID-19_Hospitalised_On_Oxygen_(%) & COVID-19_Hospitalised_On_Oxygen_(%) | 125 | -.366 | .000 |
| Pair 12 | COVID-19_Hospitalised_On_Oxygen_(%) & COVID-19_Hospitalised_On_Oxygen_(%) | 125 | -.395 | .000 |
| Pair 13 | COVID-19_Hospitalised_On_Ventilator_(%) & COVID-19_Hospitalised_On_Ventilator_(%) | 106 | -.325 | .001 |
| Pair 14 | COVID-19_Hospitalised_On_Ventilator_(%) & COVID-19_Hospitalised_On_Ventilator_(%) | 125 | -.300 | .001 |
| Pair 15 | COVID-19_Hospitalised_On_Ventilator_(%) & COVID-19_Hospitalised_On_Ventilator_(%) | 125 | -.393 | .000 |

### Paired Samples Test

| | | Paired Differences | | | |
|---|---|---|---|---|---|
| | | | | | 95% Confidence ... |
| | | Mean | Std. Deviation | Std. Error Mean | Lower |
| Pair 1 | COVID-19_Hospitalised_General_Ward_(%) - COVID-19_Hospitalised_General_Ward_(%) | -8.51182984 | 3.966823545 | .3852919628 | -9.27579263 |
| Pair 2 | COVID-19_Hospitalised_General_Ward_(%) - COVID-19_Hospitalised_General_Ward_(%) | -3.08937570 | 3.800741213 | .3399486287 | -3.76222927 |
| Pair 3 | COVID-19_Hospitalised_General_Ward_(%) - COVID-19_Hospitalised_General_Ward_(%) | 3.919824526 | 4.593289894 | .4108363377 | 3.106664331 |
| Pair 4 | COVID-19_Hospitalised_High_Care_(%) - COVID-19_Hospitalised_High_Care_(%) | 1.424876361 | 1.206812135 | .1172159565 | 1.192458772 |
| Pair 5 | COVID-19_Hospitalised_High_Care_(%) - COVID-19_Hospitalised_High_Care_(%) | .2956698690 | 1.056529136 | .0944988387 | .1086301965 |
| Pair 6 | COVID-19_Hospitalised_High_Care_(%) - COVID-19_Hospitalised_High_Care_(%) | -.694197566 | 1.283533088 | .1148026894 | -.921424246 |
| Pair 7 | COVID-19_Hospitalised_Intensive_Care_Unit_(%) - COVID-19_Hospitalised_Intensive_Care_Unit_(%) | 4.538920648 | 2.199471275 | .2136315354 | 4.115328776 |
| Pair 8 | COVID-19_Hospitalised_Intensive_Care_Unit_(%) - COVID-19_Hospitalised_Intensive_Care_Unit_(%) | .1969008848 | 3.363843160 | .3008712789 | -.398607655 |

### Paired Samples Test

| | | Paired ... 95% Confidence Interval of the ... | t | df | Sig. (2-tailed) |
|---|---|---|---|---|---|
| | | Upper | | | |
| Pair 1 | COVID-19_Hospitalised_General_Ward_(%) - COVID-19_Hospitalised_General_Ward_(%) | -7.74786705 | -22.092 | 105 | .000 |
| Pair 2 | COVID-19_Hospitalised_General_Ward_(%) - COVID-19_Hospitalised_General_Ward_(%) | -2.41652214 | -9.088 | 124 | .000 |
| Pair 3 | COVID-19_Hospitalised_General_Ward_(%) - COVID-19_Hospitalised_General_Ward_(%) | 4.732984720 | 9.541 | 124 | .000 |
| Pair 4 | COVID-19_Hospitalised_High_Care_(%) - COVID-19_Hospitalised_High_Care_(%) | 1.657293950 | 12.156 | 105 | .000 |
| Pair 5 | COVID-19_Hospitalised_High_Care_(%) - COVID-19_Hospitalised_High_Care_(%) | .4827095415 | 3.129 | 124 | .002 |
| Pair 6 | COVID-19_Hospitalised_High_Care_(%) - COVID-19_Hospitalised_High_Care_(%) | -.466970885 | -6.047 | 124 | .000 |
| Pair 7 | COVID-19_Hospitalised_Intensive_Care_Unit_(%) - COVID-19_Hospitalised_Intensive_Care_Unit_(%) | 4.962512519 | 21.246 | 105 | .000 |
| Pair 8 | COVID-19_Hospitalised_Intensive_Care_Unit_(%) - COVID-19_Hospitalised_Intensive_Care_Unit_(%) | .7924094248 | .654 | 124 | .514 |

### Paired Samples Test

| | | Paired Differences | | | |
|---|---|---|---|---|---|
| | | | | | 95% Confidence ... |
| | | Mean | Std. Deviation | Std. Error Mean | Lower |
| Pair 9 | COVID-19_Hospitalised_Intensive_Care_Unit_(%) - COVID-19_Hospitalised_Intensive_Care_Unit_(%) | -3.22562696 | 3.465941824 | .3100032610 | -3.83921025 |
| Pair 10 | COVID-19_Hospitalised_On_Oxygen_(%) - COVID-19_Hospitalised_On_Oxygen_(%) | -12.0548735 | 6.024770609 | .5851774516 | -13.2151723 |
| Pair 11 | COVID-19_Hospitalised_On_Oxygen_(%) - COVID-19_Hospitalised_On_Oxygen_(%) | -5.70381031 | 8.897303508 | .7957990184 | -7.27891949 |
| Pair 12 | COVID-19_Hospitalised_On_Oxygen_(%) - COVID-19_Hospitalised_On_Oxygen_(%) | 7.309294780 | 7.653347554 | .6845362155 | 5.954405859 |
| Pair 13 | COVID-19_Hospitalised_On_Ventilator_(%) - COVID-19_Hospitalised_On_Ventilator_(%) | 2.422498907 | 2.488458832 | .2417004882 | 1.943251488 |
| Pair 14 | COVID-19_Hospitalised_On_Ventilator_(%) - COVID-19_Hospitalised_On_Ventilator_(%) | -.937845575 | 2.084799930 | .1864701745 | -1.30692228 |
| Pair 15 | COVID-19_Hospitalised_On_Ventilator_(%) - COVID-19_Hospitalised_On_Ventilator_(%) | -2.56345852 | 2.056170670 | .1839094957 | -2.92746693 |

**Paired Samples Test**

| | | Paired ... 95% Confidence Interval of the ... Upper | t | df | Sig. (2-tailed) |
|---|---|---|---|---|---|
| Pair 9 | COVID-19_Hospitalised_Intensive_Care_Unit_(%) - COVID-19_Hospitalised_Intensive_Care_Unit_(%) | -2.61204367 | -10.405 | 124 | .000 |
| Pair 10 | COVID-19_Hospitalised_On_Oxygen_(%) - COVID-19_Hospitalised_On_Oxygen_(%) | -10.8945747 | -20.600 | 105 | .000 |
| Pair 11 | COVID-19_Hospitalised_On_Oxygen_(%) - COVID-19_Hospitalised_On_Oxygen_(%) | -4.12870114 | -7.167 | 124 | .000 |
| Pair 12 | COVID-19_Hospitalised_On_Oxygen_(%) - COVID-19_Hospitalised_On_Oxygen_(%) | 8.664183700 | 10.678 | 124 | .000 |
| Pair 13 | COVID-19_Hospitalised_On_Ventilator_(%) - COVID-19_Hospitalised_On_Ventilator_(%) | 2.901746327 | 10.023 | 105 | .000 |
| Pair 14 | COVID-19_Hospitalised_On_Ventilator_(%) - COVID-19_Hospitalised_On_Ventilator_(%) | -.568768866 | -5.029 | 124 | .000 |
| Pair 15 | COVID-19_Hospitalised_On_Ventilator_(%) - COVID-19_Hospitalised_On_Ventilator_(%) | -2.19945011 | -13.939 | 124 | .000 |

**Paired Samples Effect Sizes**

| | | | Standardizer[a] | Point Estimate | 95% ... Lower |
|---|---|---|---|---|---|
| Pair 1 | COVID-19_Hospitalised_General_Ward_(%) - COVID-19_Hospitalised_General_Ward_(%) | Cohen's d | 3.966823545 | -2.146 | -2.491 |
| | | Hedges' correction | 3.981061399 | -2.138 | -2.482 |
| Pair 2 | COVID-19_Hospitalised_General_Ward_(%) - COVID-19_Hospitalised_General_Ward_(%) | Cohen's d | 3.800741213 | -.813 | -1.014 |
| | | Hedges' correction | 3.812283874 | -.810 | -1.011 |
| Pair 3 | COVID-19_Hospitalised_General_Ward_(%) - COVID-19_Hospitalised_General_Ward_(%) | Cohen's d | 4.593289894 | .853 | .647 |
| | | Hedges' correction | 4.607239485 | .851 | .645 |
| Pair 4 | COVID-19_Hospitalised_High_Care_(%) - COVID-19_Hospitalised_High_Care_(%) | Cohen's d | 1.206812135 | 1.181 | .931 |
| | | Hedges' correction | 1.211143665 | 1.176 | .927 |
| Pair 5 | COVID-19_Hospitalised_High_Care_(%) - COVID-19_Hospitalised_High_Care_(%) | Cohen's d | 1.056529136 | .280 | .101 |
| | | Hedges' correction | 1.059737762 | .279 | .100 |
| Pair 6 | COVID-19_Hospitalised_High_Care_(%) - COVID-19_Hospitalised_High_Care_(%) | Cohen's d | 1.283533088 | -.541 | -.728 |
| | | Hedges' correction | 1.287431113 | -.539 | -.725 |
| Pair 7 | COVID-19_Hospitalised_Intensive_Care_Unit_(%) - COVID-19_Hospitalised_Intensive_Care_Unit_(%) | Cohen's d | 2.199471275 | 2.064 | 1.724 |
| | | Hedges' correction | 2.207365690 | 2.056 | 1.718 |
| Pair 8 | COVID-19_Hospitalised_Intensive_Care_Unit_(%) - COVID-19_Hospitalised_Intensive_Care_Unit_(%) | Cohen's d | 3.363843160 | .059 | -.117 |
| | | Hedges' correction | 3.374058984 | .058 | -.117 |

### Paired Samples Effect Sizes

| | | | 95% ... Upper |
|---|---|---|---|
| Pair 1 | COVID-19_Hospitalised_General_Ward_(%) - COVID-19_Hospitalised_General_Ward_(%) | Cohen's d | -1.797 |
| | | Hedges' correction | -1.791 |
| Pair 2 | COVID-19_Hospitalised_General_Ward_(%) - COVID-19_Hospitalised_General_Ward_(%) | Cohen's d | -.609 |
| | | Hedges' correction | -.607 |
| Pair 3 | COVID-19_Hospitalised_General_Ward_(%) - COVID-19_Hospitalised_General_Ward_(%) | Cohen's d | 1.057 |
| | | Hedges' correction | 1.054 |
| Pair 4 | COVID-19_Hospitalised_High_Care_(%) - COVID-19_Hospitalised_High_Care_(%) | Cohen's d | 1.427 |
| | | Hedges' correction | 1.422 |
| Pair 5 | COVID-19_Hospitalised_High_Care_(%) - COVID-19_Hospitalised_High_Care_(%) | Cohen's d | .458 |
| | | Hedges' correction | .457 |
| Pair 6 | COVID-19_Hospitalised_High_Care_(%) - COVID-19_Hospitalised_High_Care_(%) | Cohen's d | -.352 |
| | | Hedges' correction | -.351 |
| Pair 7 | COVID-19_Hospitalised_Intensive_Care_Unit_(%) - COVID-19_Hospitalised_Intensive_Care_Unit_(%) | Cohen's d | 2.399 |
| | | Hedges' correction | 2.391 |
| Pair 8 | COVID-19_Hospitalised_Intensive_Care_Unit_(%) - COVID-19_Hospitalised_Intensive_Care_Unit_(%) | Cohen's d | .234 |
| | | Hedges' correction | .233 |

### Paired Samples Effect Sizes

| | | | Standardizer[a] | Point Estimate | 95% ... |
|---|---|---|---|---|---|
| | | | | | Lower |
| Pair 9 | COVID-19_Hospitalised_Intensive_Care_Unit_(%) - COVID-19_Hospitalised_Intensive_Care_Unit_(%) | Cohen's d | 3.465941824 | -.931 | -1.139 |
| | | Hedges' correction | 3.476467716 | -.928 | -1.136 |
| Pair 10 | COVID-19_Hospitalised_On_Oxygen_(%) - COVID-19_Hospitalised_On_Oxygen_(%) | Cohen's d | 6.024770609 | -2.001 | -2.330 |
| | | Hedges' correction | 6.046394914 | -1.994 | -2.321 |
| Pair 11 | COVID-19_Hospitalised_On_Oxygen_(%) - COVID-19_Hospitalised_On_Oxygen_(%) | Cohen's d | 8.897303508 | -.641 | -.833 |
| | | Hedges' correction | 8.924324174 | -.639 | -.830 |
| Pair 12 | COVID-19_Hospitalised_On_Oxygen_(%) - COVID-19_Hospitalised_On_Oxygen_(%) | Cohen's d | 7.653347554 | .955 | .742 |
| | | Hedges' correction | 7.676590388 | .952 | .740 |
| Pair 13 | COVID-19_Hospitalised_On_Ventilator_(%) - COVID-19_Hospitalised_On_Ventilator_(%) | Cohen's d | 2.488458832 | .973 | .741 |
| | | Hedges' correction | 2.497390490 | .970 | .738 |
| Pair 14 | COVID-19_Hospitalised_On_Ventilator_(%) - COVID-19_Hospitalised_On_Ventilator_(%) | Cohen's d | 2.084799930 | -.450 | -.633 |
| | | Hedges' correction | 2.091131363 | -.448 | -.631 |
| Pair 15 | COVID-19_Hospitalised_On_Ventilator_(%) - COVID-19_Hospitalised_On_Ventilator_(%) | Cohen's d | 2.056170670 | -1.247 | -1.479 |
| | | Hedges' correction | 2.062415158 | -1.243 | -1.475 |

### Paired Samples Effect Sizes

| | | | 95% ... |
|---|---|---|---|
| | | | Upper |
| Pair 9 | COVID-19_Hospitalised_Intensive_Care_Unit_(%) - COVID-19_Hospitalised_Intensive_Care_Unit_(%) | Cohen's d | -.719 |
| | | Hedges' correction | -.717 |
| Pair 10 | COVID-19_Hospitalised_On_Oxygen_(%) - COVID-19_Hospitalised_On_Oxygen_(%) | Cohen's d | -1.669 |
| | | Hedges' correction | -1.663 |
| Pair 11 | COVID-19_Hospitalised_On_Oxygen_(%) - COVID-19_Hospitalised_On_Oxygen_(%) | Cohen's d | -.447 |
| | | Hedges' correction | -.446 |
| Pair 12 | COVID-19_Hospitalised_On_Oxygen_(%) - COVID-19_Hospitalised_On_Oxygen_(%) | Cohen's d | 1.165 |
| | | Hedges' correction | 1.162 |
| Pair 13 | COVID-19_Hospitalised_On_Ventilator_(%) - COVID-19_Hospitalised_On_Ventilator_(%) | Cohen's d | 1.203 |
| | | Hedges' correction | 1.199 |
| Pair 14 | COVID-19_Hospitalised_On_Ventilator_(%) - COVID-19_Hospitalised_On_Ventilator_(%) | Cohen's d | -.265 |
| | | Hedges' correction | -.264 |
| Pair 15 | COVID-19_Hospitalised_On_Ventilator_(%) - COVID-19_Hospitalised_On_Ventilator_(%) | Cohen's d | -1.011 |
| | | Hedges' correction | -1.008 |

a. The denominator used in estimating the effect sizes.
Cohen's d uses the sample standard deviation of the mean difference.
Hedges' correction uses the sample standard deviation of the mean difference, plus a correction factor.

```
 * Chart Builder.
GGRAPH
```

```
  /GRAPHDATASET NAME="graphdataset" VARIABLES=COVID19_Epidemic_Wave
    MEAN(COVID19_Hospitalised_Isolation_Ward) MEAN(COVID19_Hospitalised_On_Ventilator) MI
SSING=LISTWISE
    REPORTMISSING=NO
    TRANSFORM=VARSTOCASES(SUMMARY="#SUMMARY" INDEX="#INDEX")
  /GRAPHSPEC SOURCE=INLINE.
BEGIN GPL
  SOURCE: s=userSource(id("graphdataset"))
  DATA: COVID19_Epidemic_Wave=col(source(s), name("COVID19_Epidemic_Wave"), unit.category
())
  DATA: SUMMARY=col(source(s), name("#SUMMARY"))
  DATA: INDEX=col(source(s), name("#INDEX"), unit.category())
  COORD: rect(dim(1,2), cluster(3,0))
  GUIDE: axis(dim(3), label("COVID-19_Epidemic_Wave"))
  GUIDE: axis(dim(2), label("Mean"))
  GUIDE: legend(aesthetic(aesthetic.color.interior), label(""))
  GUIDE: text.title(label("Clustered Bar Mean of COVID-19_Hospitalised_Isolation_Ward_(%)
, Mean ",
    "of COVID-19_Hospitalised_On_Ventilator_(%) by COVID-19_Epidemic_Wave..."))
  SCALE: linear(dim(2), include(0))
  SCALE: cat(aesthetic(aesthetic.color.interior), include("0", "1"))
  SCALE: cat(dim(1), include("0", "1"))
  ELEMENT: interval(position(INDEX*SUMMARY*COVID19_Epidemic_Wave), color.interior(INDEX),
    shape.interior(shape.square))
END GPL.
```

### GGraph

### Notes

| | | |
|---|---|---|
| Output Created | | 10-OCT-2021 20:43:32 |
| Comments | | |
| Input | Data | C: \Users\ThaboMabuka\GOOGLE~1\ARIPRO~1\RESEAR~1\COVID-~1\Papers\ACMRG\THEIMP~1\Data\ADMISS~1\P2B9E7~1.SAV |
| | Active Dataset | DataSet1 |
| | Filter | <none> |
| | Weight | <none> |
| | Split File | <none> |
| | N of Rows in Working Data File | 484 |

### Notes

| | |
|---|---|
| Syntax | GGRAPH<br>/GRAPHDATASET<br>NAME="graphdataset"<br>VARIABLES=COVID19_E<br>pidemic_Wave<br>MEAN<br>(COVID19_Hospitalised_I<br>solation_Ward) MEAN<br>(COVID19_Hospitalised_<br>On_Ventilator)<br>MISSING=LISTWISE<br>REPORTMISSING=NO<br><br>TRANSFORM=VARSTOC<br>ASES(SUMMARY="<br>#SUMMARY" INDEX="<br>#INDEX")<br>/GRAPHSPEC<br>SOURCE=INLINE.<br>BEGIN GPL<br>SOURCE: s=userSource<br>(id("graphdataset"))<br>DATA:<br>COVID19_Epidemic_Wav<br>e=col(source(s), name<br>("COVID19_Epidemic_Wa<br>ve"), unit.category())<br>DATA: SUMMARY=col<br>(source(s), name<br>("#SUMMARY"))<br>DATA: INDEX=col<br>(source(s), name<br>("#INDEX"), unit.<br>category())<br>COORD: rect(dim(1,2),<br>cluster(3,0))<br>GUIDE: axis(dim(3), label<br>("COVID-<br>19_Epidemic_Wave"))<br>GUIDE: axis(dim(2), label<br>("Mean"))<br>GUIDE: legend(aesthetic<br>(aesthetic.color.interior),<br>label(""))<br>GUIDE: text.title(label<br>("Clustered Bar Mean of<br>COVID-<br>19_Hospitalised_Isolation<br>_Ward_(%), Mean ",<br>"of COVID-<br>19_Hospitalised_On_Venti<br>lator_(%) by COVID-<br>19_Epidemic_Wave..."))<br>SCALE: linear(dim(2),<br>include(0))<br>SCALE: cat(aesthetic<br>(aesthetic.color.interior),<br>include("0", "1"))<br>SCALE: cat(dim(1),<br>include("0", "1"))<br>ELEMENT: interval<br>(position |

### Notes

| | | |
|---|---|---|
| Resources | Processor Time | 00:00:01,06 |
| | Elapsed Time | 00:00:00,36 |

```
[DataSet1] C:\Users\ThaboMabuka\GOOGLE~1\ARIPRO~1\RESEAR~1\COVID-~1\Papers\ACMRG\THEIMP~1
\Data\ADMISS~1\P2B9E7~1.SAV
```

```
 * Chart Builder.
GGRAPH
  /GRAPHDATASET NAME="graphdataset" VARIABLES=COVID19_Epidemic_Wave
    MEANCI(COVID19_Hospitalised_General_Ward, 95) MEANCI(COVID19_Hospitalised_High_Care,
95)
    MEANCI(COVID19_Hospitalised_Intensive_Care_Unit, 95) MEANCI(COVID19_Hospitalised_On_O
xygen, 95)
    MEANCI(COVID19_Hospitalised_On_Ventilator, 95) MISSING=LISTWISE REPORTMISSING=NO
    TRANSFORM=VARSTOCASES(SUMMARY="#SUMMARY" INDEX="#INDEX" LOW="#LOW" HIGH="#HIGH")
  /GRAPHSPEC SOURCE=INLINE.
BEGIN GPL
  SOURCE: s=userSource(id("graphdataset"))
  DATA: COVID19_Epidemic_Wave=col(source(s), name("COVID19_Epidemic_Wave"), unit.category
())
  DATA: SUMMARY=col(source(s), name("#SUMMARY"))
  DATA: INDEX=col(source(s), name("#INDEX"), unit.category())
  DATA: LOW=col(source(s), name("#LOW"))
  DATA: HIGH=col(source(s), name("#HIGH"))
  COORD: rect(dim(1,2), cluster(3,0))
  GUIDE: axis(dim(3), label("COVID-19_Epidemic_Wave"))
  GUIDE: axis(dim(2), label("Mean"))
  GUIDE: legend(aesthetic(aesthetic.color.interior), label(""))
  GUIDE: text.title(label("Simple Bar Mean of COVID-19_Hospitalised_General_Ward_(%), Mea
n of ",
    "COVID-19_Hospitalised_High_Care_(%), Mean of COVID-19_Hospitalised_Intensive_Care_Un
it_(%), ",
    "Mean of COVID-19_Hospitalised_On_Oxygen_(%), Mean of COVID-19_Hospitalised_On_Ventil
ator_(%) ",
    "by COVID-19_Epidemic_Wave by INDEX"))
  GUIDE: text.footnote(label("Error Bars: 95% CI"))
  SCALE: linear(dim(2), include(0))
  SCALE: cat(aesthetic(aesthetic.color.interior), include("0", "1", "2", "3", "4"))
  SCALE: cat(dim(1), include("0", "1", "2", "3", "4"))
  ELEMENT: interval(position(INDEX*SUMMARY*COVID19_Epidemic_Wave), color.interior(INDEX),
    shape.interior(shape.square))
  ELEMENT: interval(position(region.spread.range(INDEX*(LOW+HIGH)*COVID19_Epidemic_Wave))
,
    shape.interior(shape.ibeam))
END GPL.
```

### GGraph

#### Notes

| | | |
|---|---|---|
| Output Created | | 10-OCT-2021 20:52:06 |
| Comments | | |
| Input | Data | C: \Users\ThaboMabuka\GOOGLE~1\ARIPRO~1\RESEAR~1\COVID-~1\Papers\ACMRG\THEIMP~1\Data\ADMISS~1\P2B9E7~1.SAV |
| | Active Dataset | DataSet1 |
| | Filter | <none> |
| | Weight | <none> |
| | Split File | <none> |
| | N of Rows in Working Data File | 484 |

### Notes

| | |
|---|---|
| Syntax | GGRAPH<br>/GRAPHDATASET NAME="graphdataset" VARIABLES=COVID19_Epidemic_Wave<br>MEANCI(COVID19_Hospitalised_General_Ward, 95) MEANCI(COVID19_Hospitalised_High_Care, 95)<br>MEANCI(COVID19_Hospitalised_Intensive_Care_Unit, 95) MEANCI(COVID19_Hospitalised_On_Oxygen, 95)<br>MEANCI(COVID19_Hospitalised_On_Ventilator, 95) MISSING=LISTWISE REPORTMISSING=NO<br><br>TRANSFORM=VARSTOCASES(SUMMARY="#SUMMARY" INDEX="#INDEX" LOW="#LOW" HIGH="#HIGH")<br>/GRAPHSPEC SOURCE=INLINE.<br>BEGIN GPL<br>SOURCE: s=userSource(id("graphdataset"))<br>DATA: COVID19_Epidemic_Wave=col(source(s), name("COVID19_Epidemic_Wave"), unit.category())<br>DATA: SUMMARY=col(source(s), name("#SUMMARY"))<br>DATA: INDEX=col(source(s), name("#INDEX"), unit.category())<br>DATA: LOW=col(source(s), name("#LOW"))<br>DATA: HIGH=col(source(s), name("#HIGH"))<br>COORD: rect(dim(1,2), cluster(3,0))<br>GUIDE: axis(dim(3), label("COVID-19_Epidemic_Wave"))<br>GUIDE: axis(dim(2), label("Mean"))<br>GUIDE: legend(aesthetic(aesthetic.color.interior), label(""))<br>GUIDE: text.title(label("Simple Bar Mean of [illegible] |

### Notes

| | | |
|---|---|---|
| Resources | Processor Time | 00:00:00,25 |
| | Elapsed Time | 00:00:00,23 |

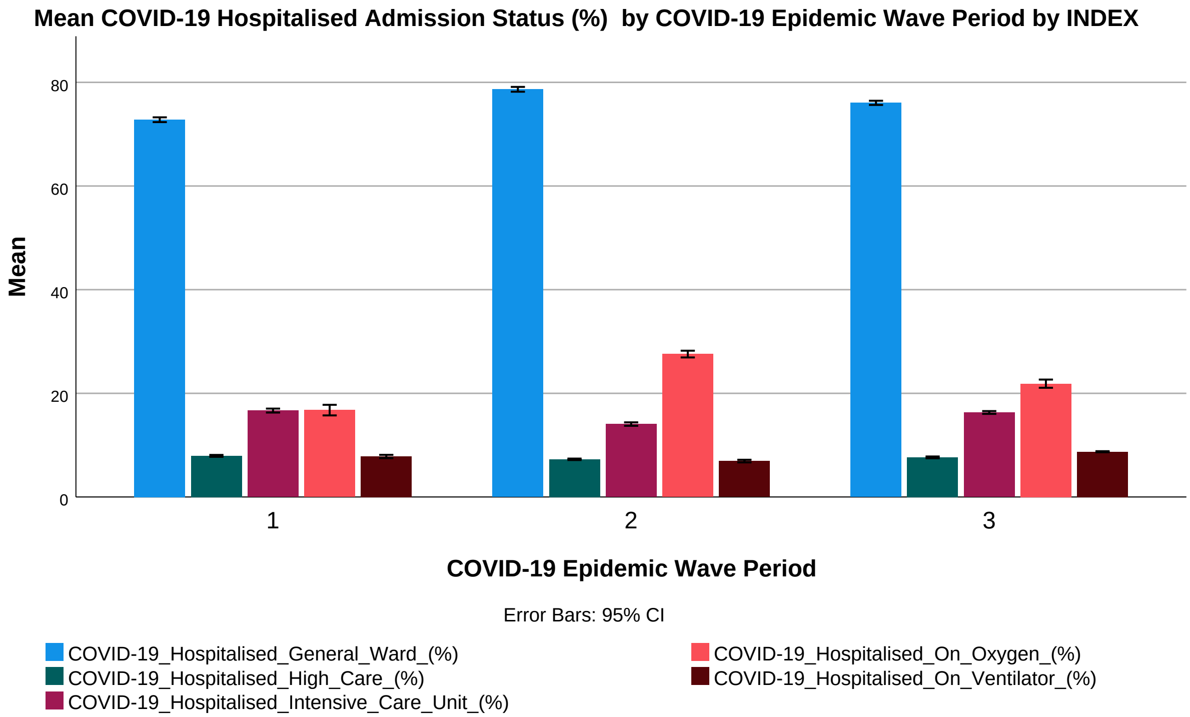


```
 SORT CASES  BY COVID19_Epidemic_Wave.
SPLIT FILE LAYERED BY COVID19_Epidemic_Wave.
DESCRIPTIVES VARIABLES=COVID19_Hospital_To_Active_Cases
  /STATISTICS=MEAN STDDEV MIN MAX.
```

### Descriptives

#### Notes

| | | |
|---|---|---|
| Output Created | | 14-OCT-2021 19:43:39 |
| Comments | | |
| Input | Data | C: \Users\ThaboMabuka\GOOGLE~1\ARIPRO~1\RESEAR~1\COVID-~1\Papers\ACMRG\THEIMP~1\Data\ADMISS~1\P2B9E7~1.SAV |
| | Active Dataset | DataSet1 |
| | Filter | <none> |
| | Weight | <none> |
| | Split File | COVID-19_Epidemic_Wave |
| | N of Rows in Working Data File | 484 |
| Missing Value Handling | Definition of Missing | User defined missing values are treated as missing. |
| | Cases Used | All non-missing data are used. |
| Syntax | | DESCRIPTIVES VARIABLES=COVID19_Hospital_To_Active_Cases /STATISTICS=MEAN STDDEV MIN MAX. |
| Resources | Processor Time | 00:00:00,00 |
| | Elapsed Time | 00:00:00,00 |

```
[DataSet1] C:\Users\ThaboMabuka\GOOGLE~1\ARIPRO~1\RESEAR~1\COVID-~1\Papers\ACMRG\THEIMP~1
\Data\ADMISS~1\P2B9E7~1.SAV
```

### Descriptive Statistics

| COVID-19_Epidemic_Wave | | N | Minimum | Maximum | Mean | Std. Deviation |
|---|---|---|---|---|---|---|
| 1 | COVID-19_Hospital_To_Active_Cases (%) | 126 | 4.02 | 12.69 | 6.8042 | 1.82080 |
| | Valid N (listwise) | 126 | | | | |
| 2 | COVID-19_Hospital_To_Active_Cases (%) | 187 | 6.46 | 23.51 | 14.4623 | 4.68271 |
| | Valid N (listwise) | 187 | | | | |
| 3 | COVID-19_Hospital_To_Active_Cases (%) | 145 | 7.18 | 18.33 | 10.6121 | 2.81241 |
| | Valid N (listwise) | 145 | | | | |

```
 SPLIT FILE OFF.
* Chart Builder.
GGRAPH
  /GRAPHDATASET NAME="graphdataset" VARIABLES=COVID19_Epidemic_Wave
    MEAN(COVID19_Hospital_To_Active_Cases)[name="MEAN_COVID19_Hospital_To_Active_Cases"]
    MISSING=LISTWISE REPORTMISSING=NO
  /GRAPHSPEC SOURCE=INLINE.
BEGIN GPL
  SOURCE: s=userSource(id("graphdataset"))
  DATA: COVID19_Epidemic_Wave=col(source(s), name("COVID19_Epidemic_Wave"), unit.category
())
  DATA: MEAN_COVID19_Hospital_To_Active_Cases=col(source(s),
    name("MEAN_COVID19_Hospital_To_Active_Cases"))
  GUIDE: axis(dim(1), label("COVID-19_Epidemic_Wave"))
  GUIDE: axis(dim(2), label("Mean COVID-19_Hospital_To_Active_Cases (%)"))
  GUIDE: text.title(label("Simple Bar Mean of COVID-19_Hospital_To_Active_Cases (%) by ",
    "COVID-19_Epidemic_Wave"))
  SCALE: linear(dim(2), include(0))
  ELEMENT: interval(position(COVID19_Epidemic_Wave*MEAN_COVID19_Hospital_To_Active_Cases)
,
    shape.interior(shape.square))
END GPL.
```

```
 * Chart Builder.
GGRAPH
  /GRAPHDATASET NAME="graphdataset" VARIABLES=COVID19_Epidemic_Wave
    MEANCI(COVID19_Hospital_To_Active_Cases, 95)[name="MEAN_COVID19_Hospital_To_Active_Ca
ses"
```

```
    LOW="MEAN_COVID19_Hospital_To_Active_Cases_LOW" HIGH="MEAN_COVID19_Hospital_To_Active
_Cases_HIGH"]
    MISSING=LISTWISE REPORTMISSING=NO
  /GRAPHSPEC SOURCE=INLINE.
BEGIN GPL
  SOURCE: s=userSource(id("graphdataset"))
  DATA: COVID19_Epidemic_Wave=col(source(s), name("COVID19_Epidemic_Wave"), unit.category
())
  DATA: MEAN_COVID19_Hospital_To_Active_Cases=col(source(s),
    name("MEAN_COVID19_Hospital_To_Active_Cases"))
  DATA: LOW=col(source(s), name("MEAN_COVID19_Hospital_To_Active_Cases_LOW"))
  DATA: HIGH=col(source(s), name("MEAN_COVID19_Hospital_To_Active_Cases_HIGH"))
  GUIDE: axis(dim(1), label("COVID-19_Epidemic_Wave"))
  GUIDE: axis(dim(2), label("Mean COVID-19_Hospital_To_Active_Cases (%)"))
  GUIDE: text.title(label("Simple Bar Mean of COVID-19_Hospital_To_Active_Cases (%) by ",
    "COVID-19_Epidemic_Wave"))
  GUIDE: text.footnote(label("Error Bars: 95% CI"))
  SCALE: linear(dim(2), include(0))
  ELEMENT: interval(position(COVID19_Epidemic_Wave*MEAN_COVID19_Hospital_To_Active_Cases)
,
    shape.interior(shape.square))
  ELEMENT: interval(position(region.spread.range(COVID19_Epidemic_Wave*(LOW+HIGH))),
    shape.interior(shape.ibeam))
END GPL.
```

### GGraph

### Notes

| | | |
|---|---|---|
| Output Created | | 14-OCT-2021 19:45:35 |
| Comments | | |
| Input | Data | C: \Users\ThaboMabuka\GOOGLE~1\ARIPRO~1\RESEAR~1\COVID-~1\Papers\ACMRG\THEIMP~1\Data\ADMISS~1\P2B9E7~1.SAV |
| | Active Dataset | DataSet1 |
| | Filter | <none> |
| | Weight | <none> |
| | Split File | <none> |
| | N of Rows in Working Data File | 484 |

### Notes

| | |
|---|---|
| Syntax | GGRAPH<br> /GRAPHDATASET<br>NAME="graphdataset"<br>VARIABLES=COVID19_E<br>pidemic_Wave<br> MEANCI<br>(COVID19_Hospital_To_A<br>ctive_Cases, 95)[name="<br>MEAN_COVID19_Hospital<br>_To_Active_Cases"<br> LOW="<br>MEAN_COVID19_Hospital<br>_To_Active_Cases_LOW"<br>HIGH="<br>MEAN_COVID19_Hospital<br>_To_Active_Cases_HIGH"<br>]<br> MISSING=LISTWISE<br>REPORTMISSING=NO<br> /GRAPHSPEC<br>SOURCE=INLINE.<br>BEGIN GPL<br> SOURCE: s=userSource<br>(id("graphdataset"))<br> DATA:<br>COVID19_Epidemic_Wav<br>e=col(source(s), name<br>("COVID19_Epidemic_Wa<br>ve"), unit.category())<br> DATA:<br>MEAN_COVID19_Hospital<br>_To_Active_Cases=col<br>(source(s),<br> name<br>("MEAN_COVID19_Hospit<br>al_To_Active_Cases"))<br> DATA: LOW=col(source<br>(s), name<br>("MEAN_COVID19_Hospit<br>al_To_Active_Cases_LO<br>W"))<br> DATA: HIGH=col(source<br>(s), name<br>("MEAN_COVID19_Hospit<br>al_To_Active_Cases_HIG<br>H"))<br> GUIDE: axis(dim(1), label<br>("COVID-<br>19_Epidemic_Wave"))<br> GUIDE: axis(dim(2), label<br>("Mean COVID-<br>19_Hospital_To_Active_C<br>ases (%)"))<br> GUIDE: text.title(label<br>("Simple Bar Mean of<br>COVID-<br>19_Hospital_To_Active_C<br>ases (%) by ",<br> "COVID-<br>19_Epidemic_Wave"))<br> GUIDE: text.footnote<br>(label("Error Bars: 95% |

### Notes

| | | |
|---|---|---|
| Resources | Processor Time | 00:00:00,45 |
| | Elapsed Time | 00:00:00,22 |

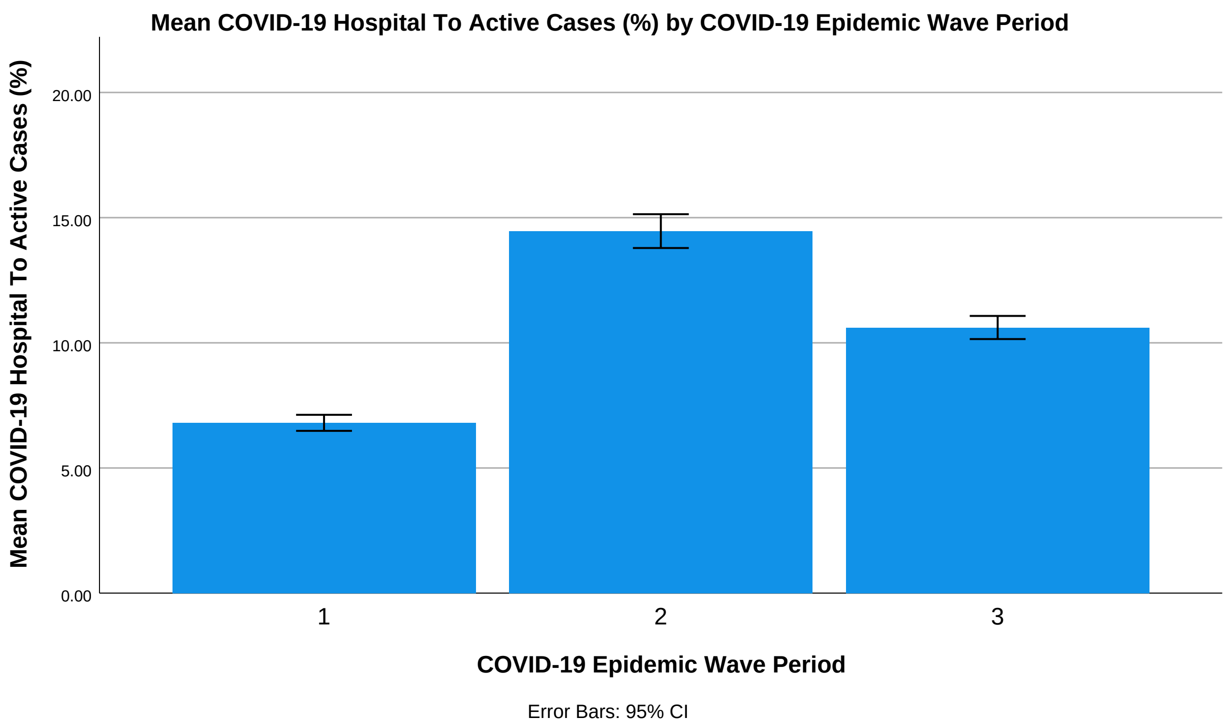


```
 DATASET ACTIVATE DataSet1.

SAVE OUTFILE=
    'C:\Users\ThaboMabuka\GOOGLE~1\ARIPRO~1\RESEAR~1\COVID-~1\Papers\ACMRG\THEIMP~1\Data\
ADMISS~1\P2B'+
    '9E7~1.SAV'
  /COMPRESSED.



 DATASET ACTIVATE DataSet1.

SAVE OUTFILE=
    'C:\Users\ThaboMabuka\GOOGLE~1\ARIPRO~1\RESEAR~1\COVID-~1\Papers\ACMRG\THEIMP~1\Data\
ADMISS~1\P2_'+
    'ANA~3.SAV'
  /COMPRESSED.
T-TEST PAIRS=COVID19_Hospital_To_Active_Cases_1 COVID19_Hospital_To_Active_Cases_1
    COVID19_Hospital_To_Active_Cases_2 WITH COVID19_Hospital_To_Active_Cases_2
```

```
  COVID19_Hospital_To_Active_Cases_3 COVID19_Hospital_To_Active_Cases_3 (PAIRED)
/ES DISPLAY(TRUE) STANDARDIZER(SD)
/CRITERIA=CI(.9500)
/MISSING=ANALYSIS.
```

### T-Test

#### Notes

| | | |
|---|---|---|
| Output Created | | 14-OCT-2021 20:06:24 |
| Comments | | |
| Input | Data | C:\Users\ThaboMabuka\GOOGLE~1\ARIPRO~1\RESEAR~1\COVID-~1\Papers\ACMRG\THEIMP~1\Data\ADMISS~1\P2_ANA~3.SAV |
| | Active Dataset | DataSet1 |
| | Filter | <none> |
| | Weight | <none> |
| | Split File | <none> |
| | N of Rows in Working Data File | 208 |
| Missing Value Handling | Definition of Missing | User defined missing values are treated as missing. |
| | Cases Used | Statistics for each analysis are based on the cases with no missing or out-of-range data for any variable in the analysis. |

### Notes

| | | |
|---|---|---|
| Syntax | | T-TEST PAIRS=COVID19_Hospital_To_Active_Cases_1 COVID19_Hospital_To_Active_Cases_1<br><br>COVID19_Hospital_To_Active_Cases_2 WITH COVID19_Hospital_To_Active_Cases_2<br><br>COVID19_Hospital_To_Active_Cases_3 COVID19_Hospital_To_Active_Cases_3 (PAIRED)<br>/ES DISPLAY(TRUE) STANDARDIZER(SD)<br>/CRITERIA=CI(.9500)<br>/MISSING=ANALYSIS. |
| Resources | Processor Time | 00:00:00,02 |
| | Elapsed Time | 00:00:00,01 |

```
[DataSet1] C:\Users\ThaboMabuka\GOOGLE~1\ARIPRO~1\RESEAR~1\COVID-~1\Papers\ACMRG\THEIMP~1
\Data\ADMISS~1\P2_ANA~3.SAV
```

### Paired Samples Statistics

| | | Mean | N | Std. Deviation | Std. Error Mean |
|---|---|---|---|---|---|
| Pair 1 | COVID-19_Hospital_To_Active_Cases_1 (%) | 6.4990 | 106 | 1.78381 | .17326 |
| | COVID-19_Hospital_To_Active_Cases_2 (% | 11.1072 | 106 | 2.91998 | .28361 |
| Pair 2 | COVID-19_Hospital_To_Active_Cases_1 (%) | 6.7968 | 125 | 1.82622 | .16334 |
| | COVID-19_Hospital_To_Active_Cases_3 (%) | 10.6499 | 125 | 2.90750 | .26005 |
| Pair 3 | COVID-19_Hospital_To_Active_Cases_2 (% | 12.2297 | 124 | 3.90958 | .35109 |
| | COVID-19_Hospital_To_Active_Cases_3 (%) | 10.0568 | 124 | 2.50533 | .22499 |

### Paired Samples Correlations

| | | N | Correlation | Sig. |
|---|---|---|---|---|
| Pair 1 | COVID-19_Hospital_To_Active_Cases_1 (%) & COVID-19_Hospital_To_Active_Cases_2 (% | 106 | -.412 | .000 |
| Pair 2 | COVID-19_Hospital_To_Active_Cases_1 (%) & COVID-19_Hospital_To_Active_Cases_3 (%) | 125 | .734 | .000 |
| Pair 3 | COVID-19_Hospital_To_Active_Cases_2 (% & COVID-19_Hospital_To_Active_Cases_3 (%) | 124 | -.178 | .048 |

### Paired Samples Test

| | | Paired Differences | | | |
|---|---|---|---|---|---|
| | | | | | 95% Confidence ... |
| | | Mean | Std. Deviation | Std. Error Mean | Lower |
| Pair 1 | COVID-19_Hospital_To_Active_Cases_1 (%) - COVID-19_Hospital_To_Active_Cases_2 (% | -4.60818 | 4.00043 | .38856 | -5.37862 |
| Pair 2 | COVID-19_Hospital_To_Active_Cases_1 (%) - COVID-19_Hospital_To_Active_Cases_3 (%) | -3.85313 | 1.99820 | .17872 | -4.20688 |
| Pair 3 | COVID-19_Hospital_To_Active_Cases_2 (% - COVID-19_Hospital_To_Active_Cases_3 (%) | 2.17297 | 5.00475 | .44944 | 1.28333 |

### Paired Samples Test

| | | Paired ... 95% Confidence Interval of the ... Upper | t | df | Sig. (2-tailed) |
|---|---|---|---|---|---|
| Pair 1 | COVID-19_Hospital_To_Active_Cases_1 (%) - COVID-19_Hospital_To_Active_Cases_2 (% | -3.83775 | -11.860 | 105 | .000 |
| Pair 2 | COVID-19_Hospital_To_Active_Cases_1 (%) - COVID-19_Hospital_To_Active_Cases_3 (%) | -3.49939 | -21.559 | 124 | .000 |
| Pair 3 | COVID-19_Hospital_To_Active_Cases_2 (% - COVID-19_Hospital_To_Active_Cases_3 (%) | 3.06261 | 4.835 | 123 | .000 |

### Paired Samples Effect Sizes

| | | | Standardizer[a] | Point Estimate | 95% ... Lower |
|---|---|---|---|---|---|
| Pair 1 | COVID-19_Hospital_To_Active_Cases_1 (%) - COVID-19_Hospital_To_Active_Cases_2 (% | Cohen's d | 4.00043 | -1.152 | -1.396 |
| | | Hedges' correction | 4.01479 | -1.148 | -1.391 |
| Pair 2 | COVID-19_Hospital_To_Active_Cases_1 (%) - COVID-19_Hospital_To_Active_Cases_3 (%) | Cohen's d | 1.99820 | -1.928 | -2.224 |
| | | Hedges' correction | 2.00427 | -1.922 | -2.217 |
| Pair 3 | COVID-19_Hospital_To_Active_Cases_2 (% - COVID-19_Hospital_To_Active_Cases_3 (%) | Cohen's d | 5.00475 | .434 | .249 |
| | | Hedges' correction | 5.02007 | .433 | .248 |

### Paired Samples Effect Sizes

| | | | 95% ... |
|---|---|---|---|
| | | | Upper |
| Pair 1 | COVID-19_Hospital_To_Active_Cases_1 (%) - COVID-19_Hospital_To_Active_Cases_2 (% | Cohen's d | -.904 |
| | | Hedges' correction | -.901 |
| Pair 2 | COVID-19_Hospital_To_Active_Cases_1 (%) - COVID-19_Hospital_To_Active_Cases_3 (%) | Cohen's d | -1.630 |
| | | Hedges' correction | -1.625 |
| Pair 3 | COVID-19_Hospital_To_Active_Cases_2 (% - COVID-19_Hospital_To_Active_Cases_3 (%) | Cohen's d | .618 |
| | | Hedges' correction | .616 |

a. The denominator used in estimating the effect sizes.
Cohen's d uses the sample standard deviation of the mean difference.
Hedges' correction uses the sample standard deviation of the mean difference, plus a correction factor.
