## Supplementary material for "The Impact of SARS-CoV-2 Lineages (Variants) on the COVID-19 Epidemic in South Africa": SPSS_Program_(COVID-19 Hospitalised Cases Discharge, Deaths)

```
 GET DATA
  /TYPE=XLSX
  /FILE='C:\Users\ThaboMabuka\Google Drive\ARI Projects\Research Projects\COVID-19 in Afr
ica\Papers\ACMRG\The Impact of SARS-CoV-2 Variants on the COVID-19 Epidemic in South Afri
ca\Data\P2_Analysis_Dataset_(COVID-19 Hospitalised Cases Admission,Discharge,Death).xlsx'
  /SHEET=name 'Hospitalised_Case_Data_Admiss'
  /CELLRANGE=FULL
  /READNAMES=ON
  /LEADINGSPACES IGNORE=YES
  /TRAILINGSPACES IGNORE=YES
  /DATATYPEMIN PERCENTAGE=95.0
  /HIDDEN IGNORE=YES.
EXECUTE.
DATASET NAME DataSet1 WINDOW=FRONT.

SAVE OUTFILE='C:\Users\ThaboMabuka\Google Drive\ARI Projects\Research Projects\COVID-19 i
n '+
    'Africa\Papers\ACMRG\The Impact of SARS-CoV-2 Variants on the COVID-19 Epidemic in So
uth '+
    'Africa\Data\P2_Analysis_Dataset_(COVID-19 Hospitalised Cases Admission,Discharge,Dea
th).sav'
  /COMPRESSED.

>Error # 61 in column 14.  Text: C:\Users\ThaboMabuka\Google Drive\ARI Projects\Research
Projects\COVID-19 in Africa\Papers\ACMRG\The Impact of SARS-CoV-2 Variants on the COVID-1
9 Epidemic in South Africa\Data\P2_Analysis_Dataset_(COVID-19 Hospitalised Cases Admissio
n,Discharge,Death).sav
>The filename is not valid.
>Execution of this command stops.

SAVE OUTFILE='C:\Users\ThaboMabuka\Google Drive\ARI Projects\Research Projects\COVID-19 i
n '+
    'Africa\Papers\ACMRG\The Impact of SARS-CoV-2 Variants on the COVID-19 Epidemic in So
uth '+
    'Africa\Data\P2_Analysis_Dataset_(COVID-19 Hospitalised Cases Admission,Discharge,Dea
th).sav'
  /COMPRESSED.

>Error # 61 in column 14.  Text: C:\Users\ThaboMabuka\Google Drive\ARI Projects\Research
Projects\COVID-19 in Africa\Papers\ACMRG\The Impact of SARS-CoV-2 Variants on the COVID-1
9 Epidemic in South Africa\Data\P2_Analysis_Dataset_(COVID-19 Hospitalised Cases Admissio
n,Discharge,Death).sav
>The filename is not valid.
>Execution of this command stops.

SAVE OUTFILE='C:\Users\ThaboMabuka\Google Drive\ARI Projects\Research Projects\COVID-19 i
```

```
n '+
    'Africa\Papers\ACMRG\The Impact of SARS-CoV-2 Variants on the COVID-19 Epidemic in So
uth '+
    'Africa\Data\P2_Analysis_Dataset_(COVID-19 Hospitalised Cases Admission,Discharge,Dea
th).sav'
  /COMPRESSED.

>Error # 61 in column 14.  Text: C:\Users\ThaboMabuka\Google Drive\ARI Projects\Research
Projects\COVID-19 in Africa\Papers\ACMRG\The Impact of SARS-CoV-2 Variants on the COVID-1
9 Epidemic in South Africa\Data\P2_Analysis_Dataset_(COVID-19 Hospitalised Cases Admissio
n,Discharge,Death).sav
>The filename is not valid.
>Execution of this command stops.

SAVE OUTFILE='C:\Users\ThaboMabuka\Google Drive\ARI Projects\Research Projects\COVID-19 i
n '+
    'Africa\Papers\ACMRG\The Impact of SARS-CoV-2 Variants on the COVID-19 Epidemic in So
uth '+
    'Africa\Data\P2_Analysis_(COVID-19 Hospitalised Cases Admission,Discharge,Death).sav'
  /COMPRESSED.
DATASET ACTIVATE DataSet1.

SAVE OUTFILE='C:\Users\ThaboMabuka\Google Drive\ARI Projects\Research Projects\COVID-19 i
n '+
    'Africa\Papers\ACMRG\The Impact of SARS-CoV-2 Variants on the COVID-19 Epidemic in So
uth '+
    'Africa\Data\P2_Analysis_(COVID-19 Hospitalised Cases Admission,Discharge,Death).sav'
  /COMPRESSED.
* Define Variable Properties.
*EpidemicWave.
VARIABLE LEVEL  EpidemicWave(SCALE).
EXECUTE.
DESCRIPTIVES VARIABLES=Cumulative_COVID19_Hospitalised_Casesn
    Cumulative_COVID19_DischargedAliveAdjn Cumulative_COVID19_Hospital_DeathsAdjn
    Daily_COVID19_Hospital_Deathsn Daily_COVID19_Hospital_Dischargen Cumulative_COVID19_D
ischargedAlive
    Cumulative_COVID19_Hospital_Deaths COVID19_CFR DischargeRate
  /STATISTICS=MEAN STDDEV MIN MAX.



 SORT CASES  BY EpidemicWave.
SPLIT FILE LAYERED BY EpidemicWave.
DATASET ACTIVATE DataSet1.

SAVE OUTFILE='C:\Users\ThaboMabuka\Google Drive\ARI Projects\Research Projects\COVID-19 i
n '+
    'Africa\Papers\ACMRG\The Impact of SARS-CoV-2 Variants on the COVID-19 Epidemic in So
uth '+
```

```
    'Africa\Data\P2_Analysis_(COVID-19 Hospitalised Cases Admission,Discharge,Death).sav'
  /COMPRESSED.
DESCRIPTIVES VARIABLES=Cumulative_COVID19_Hospitalised_Casesn
    Cumulative_COVID19_DischargedAliveAdjn Cumulative_COVID19_Hospital_DeathsAdjn
    Daily_COVID19_Hospital_Deathsn Daily_COVID19_Hospital_Dischargen Cumulative_COVID19_D
ischargedAlive
    Cumulative_COVID19_Hospital_Deaths COVID19_CFR DischargeRate
  /STATISTICS=MEAN STDDEV MIN MAX.
```

### Descriptives

#### Notes

| | | |
|---|---|---|
| Output Created | | 24-SEP-2021 15:31:06 |
| Comments | | |
| Input | Data | C:\Users\ThaboMabuka\Google Drive\ARI Projects\Research Projects\COVID-19 in Africa\Papers\ACMRG\The Impact of SARS-CoV-2 Variants on the COVID-19 Epidemic in South Africa\Data\P2_Analysis_(COVID-19 Hospitalised Cases Admission, Discharge,Death).sav |
| | Active Dataset | DataSet1 |
| | Filter | <none> |
| | Weight | <none> |
| | Split File | Epidemic Wave |
| | N of Rows in Working Data File | 484 |
| Missing Value Handling | Definition of Missing | User defined missing values are treated as missing. |
| | Cases Used | All non-missing data are used. |

**Notes**

| | | |
|---|---|---|
| Syntax | | DESCRIPTIVES VARIABLES=Cumulative_COVID19_Hospitalised_Casesn<br><br>Cumulative_COVID19_DischargedAliveAdjn Cumulative_COVID19_Hospital_DeathsAdjn<br><br>Daily_COVID19_Hospital_Deathsn Daily_COVID19_Hospital_Dischargen Cumulative_COVID19_DischargedAlive<br><br>Cumulative_COVID19_Hospital_Deaths COVID19_CFR DischargeRate<br>/STATISTICS=MEAN STDDEV MIN MAX. |
| Resources | Processor Time | 00:00:00,02 |
| | Elapsed Time | 00:00:00,01 |

### Descriptive Statistics

| Epidemic Wave | | N | Minimum | Maximum | Mean |
|---|---|---|---|---|---|
| 1 | Cumulative_COVID-19_Hospitalised_Cases (n) | 126 | 3046 | 74167 | 36755.98 |
| | Cumulative_COVID-19_Discharged Alive Adj (n) | 126 | 1615 | 55631 | 25499.67 |
| | Cumulative_COVID-19_Hospital_Deaths Adj (n) | 130 | 0 | 13261 | 5783.65 |
| | Daily_COVID-19_Hospital_Deaths (n) | 122 | 0 | 1966 | 117.88 |
| | Daily_COVID-19_Hospital_Discharge (n) | 121 | 0 | 3537 | 430.37 |
| | Cumulative_COVID-19_Discharged Alive (%) | 126 | 48.51628468 | 75.02356129 | 63.81771338 |
| | Cumulative_COVID-19_Hospital_Deaths (%) | 126 | 11.99034982 | 17.87991964 | 15.20918523 |
| | COVID-19_CFR | 121 | .0000000000 | .0686567164 | .0205939570 |
| | Discharge Rate | 121 | .0000000000 | .4251712946 | .0840101227 |
| | Valid N (listwise) | 121 | | | |
| 2 | Cumulative_COVID-19_Hospitalised_Cases (n) | 187 | 74736 | 251329 | 173948.20 |
| | Cumulative_COVID-19_Discharged Alive Adj (n) | 187 | 425 | 137231 | 75034.23 |
| | Cumulative_COVID-19_Hospital_Deaths Adj (n) | 187 | 80 | 40644 | 21389.16 |
| | Daily_COVID-19_Hospital_Deaths (n) | 180 | 0 | 1702 | 218.38 |
| | Daily_COVID-19_Hospital_Discharge (n) | 183 | -2819 | 7418 | 680.13 |
| | Cumulative_COVID-19_Discharged Alive (%) | 187 | 64.55310891 | 87.95099849 | 76.31095074 |
| | Cumulative_COVID-19_Hospital_Deaths (%) | 187 | 8.091080911 | 22.94701968 | 19.55184446 |
| | COVID-19_CFR | 183 | .0000000000 | .1288905718 | .0229211199 |
| | Discharge Rate | 183 | -.256226141 | .7762138226 | .0772757880 |
| | Valid N (listwise) | 180 | | | |

### Descriptive Statistics

| Epidemic Wave | | Std. Deviation |
|---|---|---|
| 1 | Cumulative_COVID-19_Hospitalised_Cases (n) | 24140.634 |
| | Cumulative_COVID-19_Discharged Alive Adj (n) | 18668.781 |
| | Cumulative_COVID-19_Hospital_Deaths Adj (n) | 4395.590 |
| | Daily_COVID-19_Hospital_Deaths (n) | 187.139 |
| | Daily_COVID-19_Hospital_Discharge (n) | 390.552 |
| | Cumulative_COVID-19_Discharged Alive (%) | 8.596085224 |
| | Cumulative_COVID-19_Hospital_Deaths (%) | 1.668336167 |
| | COVID-19_CFR | .0110042873 |
| | Discharge Rate | .0489421167 |
| | Valid N (listwise) | |
| 2 | Cumulative_COVID-19_Hospitalised_Cases (n) | 61666.441 |
| | Cumulative_COVID-19_Discharged Alive Adj (n) | 46932.008 |
| | Cumulative_COVID-19_Hospital_Deaths Adj (n) | 14698.148 |
| | Daily_COVID-19_Hospital_Deaths (n) | 225.579 |
| | Daily_COVID-19_Hospital_Discharge (n) | 916.917 |
| | Cumulative_COVID-19_Discharged Alive (%) | 5.810340774 |
| | Cumulative_COVID-19_Hospital_Deaths (%) | 3.234161419 |
| | COVID-19_CFR | .0160110544 |
| | Discharge Rate | .0897568589 |
| | Valid N (listwise) | |

### Descriptive Statistics

| Epidemic Wave | | N | Minimum | Maximum | Mean |
|---|---|---|---|---|---|
| 3 | Cumulative_COVID-19_Hospitalised_Cases (n) | 146 | 251360 | 411640 | 321061.07 |
| | Cumulative_COVID-19_Discharged Alive Adj (n) | 146 | 504 | 137283 | 48547.23 |
| | Cumulative_COVID-19_Hospital_Deaths Adj (n) | 146 | 115 | 40648 | 15665.08 |
| | Daily_COVID-19_Hospital_Deaths (n) | 146 | 4 | 1058 | 256.79 |
| | Daily_COVID-19_Hospital_Discharge (n) | 146 | -199 | 1960 | 808.75 |
| | Cumulative_COVID-19_Discharged Alive (%) | 146 | 60.84279766 | 77.47653688 | 68.41836986 |
| | Cumulative_COVID-19_Hospital_Deaths (%) | 146 | 15.87209302 | 24.32432432 | 20.86873814 |
| | COVID-19_CFR | 146 | .0008816399 | .1011278914 | .0207770790 |
| | Discharge Rate | 146 | -.039096267 | .1595721458 | .0683603838 |
| | Valid N (listwise) | 146 | | | |

### Descriptive Statistics

| Epidemic Wave | | Std. Deviation |
|---|---|---|
| 3 | Cumulative_COVID-19_Hospitalised_Cases (n) | 55396.756 |
| | Cumulative_COVID-19_Discharged Alive Adj (n) | 39853.828 |
| | Cumulative_COVID-19_Hospital_Deaths Adj (n) | 13245.904 |
| | Daily_COVID-19_Hospital_Deaths (n) | 195.467 |
| | Daily_COVID-19_Hospital_Discharge (n) | 498.046 |
| | Cumulative_COVID-19_Discharged Alive (%) | 4.464377614 |
| | Cumulative_COVID-19_Hospital_Deaths (%) | 2.334849542 |
| | COVID-19_CFR | .0115727781 |
| | Discharge Rate | .0272483911 |
| | Valid N (listwise) | |

```
 * Define Variable Properties.
*EpidemicWave.
VARIABLE LEVEL  EpidemicWave(NOMINAL).
EXECUTE.
SPSSINC SPLIT DATASET SPLITVAR=EpidemicWave
/OUTPUT DIRECTORY= "C:\Users\ThaboMabuka\Google Drive\ARI Projects\Research Projects\COVI
D-19 in "+
    "Africa\Papers\ACMRG\The Impact of SARS-CoV-2 Variants on the COVID-19 Epidemic in So
uth "+
    "Africa\Data" DELETECONTENTS=NO
/OPTIONS NAMES=VALUES.

>Warning # 10903.  Command name: AGGREGATE
>A SPLIT FILE command is in effect but will be ignored for AGGREGATE
>processing.
```

#### SPSSINC SPLIT DATASET

**Notes**

| | | |
|---|---|---|
| Output Created | | 24-SEP-2021 15:32:54 |
| Comments | | |
| Input | Data | C:\Users\ThaboMabuka\Google Drive\ARI Projects\Research Projects\COVID-19 in Africa\Papers\ACMRG\The Impact of SARS-CoV-2 Variants on the COVID-19 Epidemic in South Africa\Data\P2_Analysis_(COVID-19 Hospitalised Cases Admission, Discharge,Death).sav |
| | Active Dataset | DataSet1 |
| | Filter | <none> |
| | Weight | <none> |
| | Split File | Epidemic Wave |
| Syntax | | BEGIN PROGRAM '# '. |
| Resources | Processor Time | 00:00:00,00 |
| | Elapsed Time | 00:00:00,05 |

```
[DataSet1] C:\Users\ThaboMabuka\Google Drive\ARI Projects\Research Projects\COVID-19 in A
frica\Papers\ACMRG\The Impact of SARS-CoV-2 Variants on the COVID-19 Epidemic in South Af
rica\Data\P2_Analysis_(COVID-19 Hospitalised Cases Admission,Discharge,Death).sav
```

### Split File Information

| | Settings and Statistics |
|---|---|
| Split Variable Names | EpidemicWave |
| Output Directory | C:\Users\Thabo Mabuka\Google Drive\ARI Projects\Research Projects\COVID-19 in Africa\Papers\ACMRG\The Impact of SARS-CoV-2 Variants on the COVID-19 Epidemic in South Africa\Data |
| Files Deleted | 0 |
| Files Written | 3 |
| File List | None |
| Directories Cleared | No |

### Values and File Names for Split Files Written

| | Values or Labels | Directory | Data File |
|---|---|---|---|
| 1 | 1 | C:\Users\Thabo Mabuka\Google Drive\ARI Projects\Research Projects\COVID-19 in Africa\Papers\ACMRG\The Impact of SARS-CoV-2 Variants on the COVID-19 Epidemic in South Africa\Data | 1.sav |

**Values and File Names for Split Files Written**

| | Values or Labels | Directory | Data File |
|---|---|---|---|
| 2 | 2 | C: \Users\Thabo Mabuka\Google Drive\ARI Projects\Research Projects\COVID-19 in Africa\Papers\ACMRG\The Impact of SARS-CoV-2 Variants on the COVID-19 Epidemic in South Africa\Data | 2.sav |
| 3 | 3 | C: \Users\Thabo Mabuka\Google Drive\ARI Projects\Research Projects\COVID-19 in Africa\Papers\ACMRG\The Impact of SARS-CoV-2 Variants on the COVID-19 Epidemic in South Africa\Data | 3.sav |

Based on Variables: EpidemicWave

```
DATASET ACTIVATE DataSet1.

SAVE OUTFILE='C:\Users\ThaboMabuka\Google Drive\ARI Projects\Research Projects\COVID-19 i
n '+
    'Africa\Papers\ACMRG\The Impact of SARS-CoV-2 Variants on the COVID-19 Epidemic in So
uth '+
    'Africa\Data\P2_Analysis_(COVID-19 Hospitalised Cases Admission,Discharge,Death).sav'
  /COMPRESSED.
```

```
 GET
  FILE='C:\Users\ThaboMabuka\Google Drive\ARI Projects\Research Projects\COVID-19 in Afri
ca\Papers\ACMRG\The Impact of SARS-CoV-2 Variants on the COVID-19 Epidemic in South Afric
a\Data\P2_Analysis_(COVID-19 Hospitalised Cases Admission,Discharge,Death)_1.sav'.
DATASET NAME DataSet1 WINDOW=FRONT.
DATASET ACTIVATE DataSet1.

SAVE OUTFILE='C:\Users\ThaboMabuka\Google Drive\ARI Projects\Research Projects\COVID-19 i
n '+
    'Africa\Papers\ACMRG\The Impact of SARS-CoV-2 Variants on the COVID-19 Epidemic in So
uth '+
    'Africa\Data\P2_Analysis_(COVID-19 Hospitalised Cases Admission,Discharge,Death)_1.sa
v'
  /COMPRESSED.
DATASET ACTIVATE DataSet1.

SAVE OUTFILE='C:\Users\ThaboMabuka\Google Drive\ARI Projects\Research Projects\COVID-19 i
n '+
    'Africa\Papers\ACMRG\The Impact of SARS-CoV-2 Variants on the COVID-19 Epidemic in So
uth '+
    'Africa\Data\P2_Analysis_(COVID-19 Hospitalised Cases Admission,Discharge,Death)_1.sa
v'
  /COMPRESSED.
GET
  FILE='C:\Users\ThaboMabuka\Google Drive\ARI Projects\Research Projects\COVID-19 in Afri
ca\Papers\ACMRG\The Impact of SARS-CoV-2 Variants on the COVID-19 Epidemic in South Afric
a\Data\P2_Analysis_(COVID-19 Hospitalised Cases Admission,Discharge,Death)_2.sav'.
DATASET NAME DataSet2 WINDOW=FRONT.
DATASET ACTIVATE DataSet2.

SAVE OUTFILE='C:\Users\ThaboMabuka\Google Drive\ARI Projects\Research Projects\COVID-19 i
n '+
    'Africa\Papers\ACMRG\The Impact of SARS-CoV-2 Variants on the COVID-19 Epidemic in So
uth '+
    'Africa\Data\P2_Analysis_(COVID-19 Hospitalised Cases Admission,Discharge,Death)_2.sa
v'
  /COMPRESSED.
DATASET ACTIVATE DataSet2.

SAVE OUTFILE='C:\Users\ThaboMabuka\Google Drive\ARI Projects\Research Projects\COVID-19 i
n '+
    'Africa\Papers\ACMRG\The Impact of SARS-CoV-2 Variants on the COVID-19 Epidemic in So
uth '+
    'Africa\Data\P2_Analysis_(COVID-19 Hospitalised Cases Admission,Discharge,Death)_2.sa
v'
  /COMPRESSED.
GET
```

```
  FILE='C:\Users\ThaboMabuka\Google Drive\ARI Projects\Research Projects\COVID-19 in Afri
ca\Papers\ACMRG\The Impact of SARS-CoV-2 Variants on the COVID-19 Epidemic in South Afric
a\Data\P2_Analysis_(COVID-19 Hospitalised Cases Admission,Discharge,Death)_3.sav'.
DATASET NAME DataSet3 WINDOW=FRONT.
DATASET ACTIVATE DataSet3.

SAVE OUTFILE='C:\Users\ThaboMabuka\Google Drive\ARI Projects\Research Projects\COVID-19 i
n '+
    'Africa\Papers\ACMRG\The Impact of SARS-CoV-2 Variants on the COVID-19 Epidemic in So
uth '+
    'Africa\Data\P2_Analysis_(COVID-19 Hospitalised Cases Admission,Discharge,Death)_3.sa
v'
  /COMPRESSED.
DATASET ACTIVATE DataSet3.

SAVE OUTFILE='C:\Users\ThaboMabuka\Google Drive\ARI Projects\Research Projects\COVID-19 i
n '+
    'Africa\Papers\ACMRG\The Impact of SARS-CoV-2 Variants on the COVID-19 Epidemic in So
uth '+
    'Africa\Data\P2_Analysis_(COVID-19 Hospitalised Cases Admission,Discharge,Death)_3.sa
v'
  /COMPRESSED.
DATASET ACTIVATE DataSet1.
SORT CASES BY Data_Point.
DATASET ACTIVATE DataSet2.
SORT CASES BY Data_Point.
DATASET ACTIVATE DataSet1.
MATCH FILES /FILE=*
  /FILE='DataSet2'
  /BY Data_Point.
EXECUTE.
SORT CASES BY Data_Point.
DATASET ACTIVATE DataSet3.
SORT CASES BY Data_Point.
DATASET ACTIVATE DataSet1.
MATCH FILES /FILE=*
  /FILE='DataSet3'
  /BY Data_Point.
EXECUTE.

SAVE OUTFILE='C:\Users\ThaboMabuka\Google Drive\ARI Projects\Research Projects\COVID-19 i
n '+
    'Africa\Papers\ACMRG\The Impact of SARS-CoV-2 Variants on the COVID-19 Epidemic in So
uth '+
    'Africa\Data\P2_Analysis_(COVID-19 Hospitalised Cases Admission,Discharge,Death)_M.sa
v'
  /COMPRESSED.
```

```
 DATASET ACTIVATE DataSet3.
DATASET ACTIVATE DataSet3.

SAVE OUTFILE='C:\Users\ThaboMabuka\Google Drive\ARI Projects\Research Projects\COVID-19 i
n '+
    'Africa\Papers\ACMRG\The Impact of SARS-CoV-2 Variants on the COVID-19 Epidemic in So
uth '+
    'Africa\Data\P2_Analysis_(COVID-19 Hospitalised Cases Admission,Discharge,Death)_3.sa
v'
  /COMPRESSED.
DATASET ACTIVATE DataSet1.
DATASET CLOSE DataSet3.
DATASET ACTIVATE DataSet2.
DATASET ACTIVATE DataSet2.

SAVE OUTFILE='C:\Users\ThaboMabuka\Google Drive\ARI Projects\Research Projects\COVID-19 i
n '+
    'Africa\Papers\ACMRG\The Impact of SARS-CoV-2 Variants on the COVID-19 Epidemic in So
uth '+
    'Africa\Data\P2_Analysis_(COVID-19 Hospitalised Cases Admission,Discharge,Death)_2.sa
v'
  /COMPRESSED.
DATASET ACTIVATE DataSet1.
DATASET CLOSE DataSet2.
T-TEST PAIRS=Cumulative_COVID19_DischargedAliveAdjn_1 Cumulative_COVID19_DischargedAliveA
djn_1
    Cumulative_COVID19_DischargedAliveAdjn_2 Cumulative_COVID19_Hospital_DeathsAdjn_1
    Cumulative_COVID19_Hospital_DeathsAdjn_1 Cumulative_COVID19_Hospital_DeathsAdjn_2
    Cumulative_COVID19_DischargedAlive_1 Cumulative_COVID19_DischargedAlive_1
    Cumulative_COVID19_DischargedAlive_2 Cumulative_COVID19_Hospital_Deaths_1
    Cumulative_COVID19_Hospital_Deaths_1 Cumulative_COVID19_Hospital_Deaths_2 WITH
    Cumulative_COVID19_DischargedAliveAdjn_2 Cumulative_COVID19_DischargedAliveAdjn_3
    Cumulative_COVID19_DischargedAliveAdjn_3 Cumulative_COVID19_Hospital_DeathsAdjn_2
    Cumulative_COVID19_Hospital_DeathsAdjn_3 Cumulative_COVID19_Hospital_DeathsAdjn_3
    Cumulative_COVID19_DischargedAlive_2 Cumulative_COVID19_DischargedAlive_3
    Cumulative_COVID19_DischargedAlive_3 Cumulative_COVID19_Hospital_Deaths_2
    Cumulative_COVID19_Hospital_Deaths_3 Cumulative_COVID19_Hospital_Deaths_3 (PAIRED)
  /ES DISPLAY(TRUE) STANDARDIZER(SD)
  /CRITERIA=CI(.9500)
  /MISSING=ANALYSIS.
```

### T-Test

### Notes

| | | |
|---|---|---|
| Output Created | | 24-SEP-2021 16:04:15 |
| Comments | | |
| Input | Data | C:\Users\ThaboMabuka\Google Drive\ARI Projects\Research Projects\COVID-19 in Africa\Papers\ACMRG\The Impact of SARS-CoV-2 Variants on the COVID-19 Epidemic in South Africa\Data\P2_Analysis_(COVID-19 Hospitalised Cases Admission, Discharge,Death)_M.sav |
| | Active Dataset | DataSet1 |
| | Filter | <none> |
| | Weight | <none> |
| | Split File | <none> |
| | N of Rows in Working Data File | 208 |
| Missing Value Handling | Definition of Missing | User defined missing values are treated as missing. |
| | Cases Used | Statistics for each analysis are based on the cases with no missing or out-of-range data for any variable in the analysis. |

### Notes

| | |
|---|---|
| Syntax | T-TEST<br>PAIRS=Cumulative_COVID19_DischargedAliveAdjn_1<br>Cumulative_COVID19_DischargedAliveAdjn_1<br><br>Cumulative_COVID19_DischargedAliveAdjn_2<br>Cumulative_COVID19_Hospital_DeathsAdjn_1<br><br>Cumulative_COVID19_Hospital_DeathsAdjn_1<br>Cumulative_COVID19_Hospital_DeathsAdjn_2<br><br>Cumulative_COVID19_DischargedAlive_1<br>Cumulative_COVID19_DischargedAlive_1<br><br>Cumulative_COVID19_DischargedAlive_2<br>Cumulative_COVID19_Hospital_Deaths_1<br><br>Cumulative_COVID19_Hospital_Deaths_1<br>Cumulative_COVID19_Hospital_Deaths_2 WITH<br><br>Cumulative_COVID19_DischargedAliveAdjn_2<br>Cumulative_COVID19_DischargedAliveAdjn_3<br><br>Cumulative_COVID19_DischargedAliveAdjn_3<br>Cumulative_COVID19_Hospital_DeathsAdjn_2<br><br>Cumulative_COVID19_Hospital_DeathsAdjn_3<br>Cumulative_COVID19_Hospital_DeathsAdjn_3<br><br>Cumulative_COVID19_DischargedAlive_2<br>Cumulative_COVID19_DischargedAlive_3<br><br>Cumulative_COVID19_DischargedAlive_3<br>Cumulative_COVID19_Hospital_Deaths_2<br><br>Cumulative_COVID19_Hospital_Deaths_3<br>Cumulative_COVID19_Hospital_Deaths_3 (PAIRED)<br>/ES DISPLAY(TRUE) |

**Notes**

| | | |
|---|---|---|
| Resources | Processor Time | 00:00:00,02 |
| | Elapsed Time | 00:00:00,01 |

**Paired Samples Statistics**

| | | Mean | N | Std. Deviation | Std. Error Mean |
|---|---|---|---|---|---|
| Pair 1 | Cumulative_COVID-19_Discharged Alive Adj (n) _1 | 28780.08 | 106 | 17742.759 | 1723.329 |
| | Cumulative_COVID-19_Discharged Alive Adj (n) _2 | 38597.01 | 106 | 24489.171 | 2378.599 |
| Pair 2 | Cumulative_COVID-19_Discharged Alive Adj (n) _1 | 25499.67 | 126 | 18668.781 | 1663.147 |
| | Cumulative_COVID-19_Discharged Alive Adj (n) _3 | 40228.07 | 126 | 34191.258 | 3045.999 |
| Pair 3 | Cumulative_COVID-19_Discharged Alive Adj (n) _2 | 48174.29 | 125 | 33083.845 | 2959.109 |
| | Cumulative_COVID-19_Discharged Alive Adj (n) _3 | 54781.62 | 125 | 38721.686 | 3463.373 |
| Pair 4 | Cumulative_COVID-19_Hospital_Deaths Adj (n) _1 | 6538.17 | 109 | 4219.384 | 404.144 |
| | Cumulative_COVID-19_Hospital_Deaths Adj (n) _2 | 10400.85 | 109 | 8625.568 | 826.180 |
| Pair 5 | Cumulative_COVID-19_Hospital_Deaths Adj (n) _1 | 5783.65 | 130 | 4395.590 | 385.519 |
| | Cumulative_COVID-19_Hospital_Deaths Adj (n) _3 | 13130.37 | 130 | 11749.060 | 1030.461 |
| Pair 6 | Cumulative_COVID-19_Hospital_Deaths Adj (n) _2 | 13209.94 | 125 | 10917.905 | 976.527 |
| | Cumulative_COVID-19_Hospital_Deaths Adj (n) _3 | 17688.93 | 125 | 12923.152 | 1155.882 |

### Paired Samples Statistics

| | | Mean | N | Std. Deviation | Std. Error Mean |
|---|---|---|---|---|---|
| Pair 7 | Cumulative_COVID-19_Discharged Alive (%)_1 | 65.24825587 | 106 | 8.298294475 | .8060016105 |
| | Cumulative_COVID-19_Discharged Alive (%)_2 | 76.18726976 | 106 | 7.400603651 | .7188101699 |
| Pair 8 | Cumulative_COVID-19_Discharged Alive (%)_1 | 63.81771338 | 126 | 8.596085224 | .7658001375 |
| | Cumulative_COVID-19_Discharged Alive (%)_3 | 67.50184876 | 126 | 4.045514349 | .3604030630 |
| Pair 9 | Cumulative_COVID-19_Discharged Alive (%)_2 | 75.80392065 | 125 | 7.045051816 | .6301285906 |
| | Cumulative_COVID-19_Discharged Alive (%)_3 | 67.87510275 | 125 | 4.397068488 | .3932857616 |
| Pair 10 | Cumulative_COVID-19_Hospital_Deaths (%)_1 | 15.44156691 | 106 | 1.652760255 | .1605302669 |
| | Cumulative_COVID-19_Hospital_Deaths (%)_2 | 17.36718621 | 106 | 2.600114955 | .2525454896 |
| Pair 11 | Cumulative_COVID-19_Hospital_Deaths (%)_1 | 15.20918523 | 126 | 1.668336167 | .1486271987 |
| | Cumulative_COVID-19_Hospital_Deaths (%)_3 | 20.53527493 | 126 | 2.307523902 | .2055705679 |
| Pair 12 | Cumulative_COVID-19_Hospital_Deaths (%)_2 | 18.00322113 | 125 | 2.893723593 | .2588225065 |
| | Cumulative_COVID-19_Hospital_Deaths (%)_3 | 20.80158262 | 125 | 2.497709869 | .2234019622 |

### Paired Samples Correlations

| | | N | Correlation | Sig. |
|---|---|---|---|---|
| Pair 1 | Cumulative_COVID-19_Discharged Alive Adj (n)_1 & Cumulative_COVID-19_Discharged Alive Adj (n)_2 | 106 | .937 | .000 |
| Pair 2 | Cumulative_COVID-19_Discharged Alive Adj (n)_1 & Cumulative_COVID-19_Discharged Alive Adj (n)_3 | 126 | .916 | .000 |
| Pair 3 | Cumulative_COVID-19_Discharged Alive Adj (n)_2 & Cumulative_COVID-19_Discharged Alive Adj (n)_3 | 125 | .927 | .000 |
| Pair 4 | Cumulative_COVID-19_Hospital_Deaths Adj (n)_1 & Cumulative_COVID-19_Hospital_Deaths Adj (n)_2 | 109 | .804 | .000 |
| Pair 5 | Cumulative_COVID-19_Hospital_Deaths Adj (n)_1 & Cumulative_COVID-19_Hospital_Deaths Adj (n)_3 | 130 | .864 | .000 |
| Pair 6 | Cumulative_COVID-19_Hospital_Deaths Adj (n)_2 & Cumulative_COVID-19_Hospital_Deaths Adj (n)_3 | 125 | .933 | .000 |
| Pair 7 | Cumulative_COVID-19_Discharged Alive (%)_1 & Cumulative_COVID-19_Discharged Alive (%)_2 | 106 | -.741 | .000 |
| Pair 8 | Cumulative_COVID-19_Discharged Alive (%)_1 & Cumulative_COVID-19_Discharged Alive (%)_3 | 126 | -.298 | .001 |

### Paired Samples Correlations

| | | N | Correlation | Sig. |
|---|---|---|---|---|
| Pair 9 | Cumulative_COVID-19_Discharged Alive (%)_2 & Cumulative_COVID-19_Discharged Alive (%)_3 | 125 | -.209 | .019 |
| Pair 10 | Cumulative_COVID-19_Hospital_Deaths (%)_1 & Cumulative_COVID-19_Hospital_Deaths (%)_2 | 106 | .875 | .000 |
| Pair 11 | Cumulative_COVID-19_Hospital_Deaths (%)_1 & Cumulative_COVID-19_Hospital_Deaths (%)_3 | 126 | .555 | .000 |
| Pair 12 | Cumulative_COVID-19_Hospital_Deaths (%)_2 & Cumulative_COVID-19_Hospital_Deaths (%)_3 | 125 | .570 | .000 |

### Paired Samples Test

| | | Paired Differences | | | |
|---|---|---|---|---|---|
| | | | | | 95% Confidence ... |
| | | Mean | Std. Deviation | Std. Error Mean | Lower |
| Pair 1 | Cumulative_COVID-19_Discharged Alive Adj (n)_1 - Cumulative_COVID-19_Discharged Alive Adj (n)_2 | -9816.934 | 10013.785 | 972.625 | -11745.469 |
| Pair 2 | Cumulative_COVID-19_Discharged Alive Adj (n)_1 - Cumulative_COVID-19_Discharged Alive Adj (n)_3 | -14728.397 | 18664.726 | 1662.786 | -18019.257 |
| Pair 3 | Cumulative_COVID-19_Discharged Alive Adj (n)_2 - Cumulative_COVID-19_Discharged Alive Adj (n)_3 | -6607.328 | 14796.446 | 1323.434 | -9226.775 |

### Paired Samples Test

| | | Paired ... 95% Confidence Interval of the ... | | | |
|---|---|---|---|---|---|
| | | Upper | t | df | Sig. (2-tailed) |
| Pair 1 | Cumulative_COVID-19_Discharged Alive Adj (n) _1 - Cumulative_COVID-19_Discharged Alive Adj (n) _2 | -7888.399 | -10.093 | 105 | .000 |
| Pair 2 | Cumulative_COVID-19_Discharged Alive Adj (n) _1 - Cumulative_COVID-19_Discharged Alive Adj (n) _3 | -11437.537 | -8.858 | 125 | .000 |
| Pair 3 | Cumulative_COVID-19_Discharged Alive Adj (n) _2 - Cumulative_COVID-19_Discharged Alive Adj (n) _3 | -3987.881 | -4.993 | 124 | .000 |

### Paired Samples Test

| | | Paired Differences | | | |
|---|---|---|---|---|---|
| | | | | | 95% Confidence ... |
| | | Mean | Std. Deviation | Std. Error Mean | Lower |
| Pair 4 | Cumulative_COVID-19_Hospital_Deaths Adj (n)_1 - Cumulative_COVID-19_Hospital_Deaths Adj (n)_2 | -3862.688 | 5805.706 | 556.086 | -4964.947 |
| Pair 5 | Cumulative_COVID-19_Hospital_Deaths Adj (n)_1 - Cumulative_COVID-19_Hospital_Deaths Adj (n)_3 | -7346.723 | 8254.537 | 723.971 | -8779.117 |
| Pair 6 | Cumulative_COVID-19_Hospital_Deaths Adj (n)_2 - Cumulative_COVID-19_Hospital_Deaths Adj (n)_3 | -4478.992 | 4790.013 | 428.432 | -5326.978 |
| Pair 7 | Cumulative_COVID-19_Discharged Alive (%)_1 - Cumulative_COVID-19_Discharged Alive (%)_2 | -10.9390139 | 14.65234539 | 1.423161593 | -13.7608804 |
| Pair 8 | Cumulative_COVID-19_Discharged Alive (%)_1 - Cumulative_COVID-19_Discharged Alive (%)_3 | -3.68413538 | 10.53443059 | .9384816677 | -5.54150707 |
| Pair 9 | Cumulative_COVID-19_Discharged Alive (%)_2 - Cumulative_COVID-19_Discharged Alive (%)_3 | 7.928817899 | 9.051132559 | .8095579070 | 6.326476030 |
| Pair 10 | Cumulative_COVID-19_Hospital_Deaths (%)_1 - Cumulative_COVID-19_Hospital_Deaths (%)_2 | -1.92561931 | 1.402960588 | .1362675784 | -2.19581274 |
| Pair 11 | Cumulative_COVID-19_Hospital_Deaths (%)_1 - Cumulative_COVID-19_Hospital_Deaths (%)_3 | -5.32608970 | 1.958943497 | .1745165573 | -5.67147963 |

### Paired Samples Test

| | | Paired ... 95% Confidence Interval of the ... Upper | t | df | Sig. (2-tailed) |
|---|---|---|---|---|---|
| Pair 4 | Cumulative_COVID-19_Hospital_Deaths Adj (n)_1 - Cumulative_COVID-19_Hospital_Deaths Adj (n)_2 | -2760.430 | -6.946 | 108 | .000 |
| Pair 5 | Cumulative_COVID-19_Hospital_Deaths Adj (n)_1 - Cumulative_COVID-19_Hospital_Deaths Adj (n)_3 | -5914.329 | -10.148 | 129 | .000 |
| Pair 6 | Cumulative_COVID-19_Hospital_Deaths Adj (n)_2 - Cumulative_COVID-19_Hospital_Deaths Adj (n)_3 | -3631.006 | -10.454 | 124 | .000 |
| Pair 7 | Cumulative_COVID-19_Discharged Alive (%)_1 - Cumulative_COVID-19_Discharged Alive (%)_2 | -8.11714734 | -7.686 | 105 | .000 |
| Pair 8 | Cumulative_COVID-19_Discharged Alive (%)_1 - Cumulative_COVID-19_Discharged Alive (%)_3 | -1.82676368 | -3.926 | 125 | .000 |
| Pair 9 | Cumulative_COVID-19_Discharged Alive (%)_2 - Cumulative_COVID-19_Discharged Alive (%)_3 | 9.531159768 | 9.794 | 124 | .000 |
| Pair 10 | Cumulative_COVID-19_Hospital_Deaths (%)_1 - Cumulative_COVID-19_Hospital_Deaths (%)_2 | -1.65542587 | -14.131 | 105 | .000 |
| Pair 11 | Cumulative_COVID-19_Hospital_Deaths (%)_1 - Cumulative_COVID-19_Hospital_Deaths (%)_3 | -4.98069978 | -30.519 | 125 | .000 |

### Paired Samples Test

| | | Paired Differences | | | |
|---|---|---|---|---|---|
| | | | | | 95% Confidence ... |
| | | Mean | Std. Deviation | Std. Error Mean | Lower |
| Pair 12 | Cumulative_COVID-19_Hospital_Deaths (%)_2 - Cumulative_COVID-19_Hospital_Deaths (%)_3 | -2.79836149 | 2.524615971 | .2258085171 | -3.24529979 |

### Paired Samples Test

| | | Paired ... | | | |
|---|---|---|---|---|---|
| | | 95% Confidence Interval of the ... | | | |
| | | Upper | t | df | Sig. (2-tailed) |
| Pair 12 | Cumulative_COVID-19_Hospital_Deaths (%)_2 - Cumulative_COVID-19_Hospital_Deaths (%)_3 | -2.35142318 | -12.393 | 124 | .000 |

```
 T-TEST PAIRS=COVID19_CFR_1 COVID19_CFR_1 COVID19_CFR_2 DischargeRate_1 DischargeRate_1
    DischargeRate_2 WITH COVID19_CFR_2 COVID19_CFR_3 COVID19_CFR_3 DischargeRate_2 Discha
rgeRate_3
    DischargeRate_3 (PAIRED)
  /ES DISPLAY(TRUE) STANDARDIZER(SD)
  /CRITERIA=CI(.9500)
  /MISSING=ANALYSIS.
```

### T-Test

### Notes

| | | |
|---|---|---|
| Output Created | | 24-SEP-2021 16:06:00 |
| Comments | | |
| Input | Data | C: \Users\ThaboMabuka\Google Drive\ARI Projects\Research Projects\COVID-19 in Africa\Papers\ACMRG\The Impact of SARS-CoV-2 Variants on the COVID-19 Epidemic in South Africa\Data\P2_Analysis_(COVID-19 Hospitalised Cases Admission, Discharge,Death)_M.sav |
| | Active Dataset | DataSet1 |
| | Filter | <none> |
| | Weight | <none> |
| | Split File | <none> |
| | N of Rows in Working Data File | 208 |
| Missing Value Handling | Definition of Missing | User defined missing values are treated as missing. |
| | Cases Used | Statistics for each analysis are based on the cases with no missing or out-of-range data for any variable in the analysis. |
| Syntax | | T-TEST PAIRS=COVID19_CFR_1 COVID19_CFR_1 COVID19_CFR_2 DischargeRate_1 DischargeRate_1 DischargeRate_2 WITH COVID19_CFR_2 COVID19_CFR_3 COVID19_CFR_3 DischargeRate_2 DischargeRate_3 DischargeRate_3 (PAIRED) /ES DISPLAY(TRUE) STANDARDIZER(SD) /CRITERIA=CI(.9500) /MISSING=ANALYSIS. |

### Notes

| | | |
|---|---|---|
| Resources | Processor Time | 00:00:00,02 |
| | Elapsed Time | 00:00:00,01 |

### Paired Samples Statistics

| | | Mean | N | Std. Deviation | Std. Error Mean |
|---|---|---|---|---|---|
| Pair 1 | COVID-19_CFR_1 | .0203564476 | 99 | .0106671499 | .0010720889 |
| | COVID-19_CFR_2 | .0248669910 | 99 | .0184052056 | .0018497928 |
| Pair 2 | COVID-19_CFR_1 | .0205939570 | 121 | .0110042873 | .0010003898 |
| | COVID-19_CFR_3 | .0217627209 | 121 | .0119543711 | .0010867610 |
| Pair 3 | COVID-19_CFR_2 | .0244344685 | 121 | .0170616632 | .0015510603 |
| | COVID-19_CFR_3 | .0217336502 | 121 | .0118692338 | .0010790213 |
| Pair 4 | Discharge Rate_1 | .0851480861 | 99 | .0501163305 | .0050368807 |
| | Discharge Rate_2 | .0808847250 | 99 | .0992689734 | .0099769072 |
| Pair 5 | Discharge Rate_1 | .0840101227 | 121 | .0489421167 | .0044492833 |
| | Discharge Rate_3 | .0694106188 | 121 | .0266981047 | .0024271004 |
| Pair 6 | Discharge Rate_2 | .0842181235 | 121 | .1066080753 | .0096916432 |
| | Discharge Rate_3 | .0704163477 | 121 | .0246138176 | .0022376198 |

### Paired Samples Correlations

| | | N | Correlation | Sig. |
|---|---|---|---|---|
| Pair 1 | COVID-19_CFR_1 & COVID-19_CFR_2 | 99 | .118 | .245 |
| Pair 2 | COVID-19_CFR_1 & COVID-19_CFR_3 | 121 | -.003 | .970 |
| Pair 3 | COVID-19_CFR_2 & COVID-19_CFR_3 | 121 | .048 | .603 |
| Pair 4 | Discharge Rate_1 & Discharge Rate_2 | 99 | .008 | .940 |
| Pair 5 | Discharge Rate_1 & Discharge Rate_3 | 121 | -.039 | .668 |
| Pair 6 | Discharge Rate_2 & Discharge Rate_3 | 121 | -.127 | .164 |

### Paired Samples Test

| | | Paired Differences | | | |
|---|---|---|---|---|---|
| | | | | | 95% Confidence ... |
| | | Mean | Std. Deviation | Std. Error Mean | Lower |
| Pair 1 | COVID-19_CFR_1 - COVID-19_CFR_2 | -.004510543 | .0201549649 | .0020256502 | -.008530380 |
| Pair 2 | COVID-19_CFR_1 - COVID-19_CFR_3 | -.001168764 | .0162756187 | .0014796017 | -.004098272 |
| Pair 3 | COVID-19_CFR_2 - COVID-19_CFR_3 | .0027008183 | .0203131128 | .0018466466 | -.000955413 |
| Pair 4 | Discharge Rate_1 - Discharge Rate_2 | .0042633611 | .1108597012 | .0111418192 | -.017847216 |
| Pair 5 | Discharge Rate_1 - Discharge Rate_3 | .0145995039 | .0566648001 | .0051513455 | .0044001984 |
| Pair 6 | Discharge Rate_2 - Discharge Rate_3 | .0138017758 | .1124257861 | .0102205260 | -.006434154 |

### Paired Samples Test

| | | Paired ... | | | |
|---|---|---|---|---|---|
| | | 95% Confidence Interval of the ... | | | |
| | | Upper | t | df | Sig. (2-tailed) |
| Pair 1 | COVID-19_CFR_1 - COVID-19_CFR_2 | -.000490707 | -2.227 | 98 | .028 |
| Pair 2 | COVID-19_CFR_1 - COVID-19_CFR_3 | .0017607445 | -.790 | 120 | .431 |
| Pair 3 | COVID-19_CFR_2 - COVID-19_CFR_3 | .0063570501 | 1.463 | 120 | .146 |
| Pair 4 | Discharge Rate_1 - Discharge Rate_2 | .0263739386 | .383 | 98 | .703 |
| Pair 5 | Discharge Rate_1 - Discharge Rate_3 | .0247988094 | 2.834 | 120 | .005 |
| Pair 6 | Discharge Rate_2 - Discharge Rate_3 | .0340377060 | 1.350 | 120 | .179 |

**Paired Samples Effect Sizes**

| | | | Standardizer[a] | Point Estimate | 95% ... Lower |
|---|---|---|---|---|---|
| Pair 1 | COVID-19_CFR_1 - COVID-19_CFR_2 | Cohen's d | .0201549649 | -.224 | -.423 |
| | | Hedges' correction | .0202325006 | -.223 | -.421 |
| Pair 2 | COVID-19_CFR_1 - COVID-19_CFR_3 | Cohen's d | .0162756187 | -.072 | -.250 |
| | | Hedges' correction | .0163267017 | -.072 | -.249 |
| Pair 3 | COVID-19_CFR_2 - COVID-19_CFR_3 | Cohen's d | .0203131128 | .133 | -.046 |
| | | Hedges' correction | .0203768680 | .133 | -.046 |
| Pair 4 | Discharge Rate_1 - Discharge Rate_2 | Cohen's d | .1108597012 | .038 | -.159 |
| | | Hedges' correction | .1112861758 | .038 | -.158 |
| Pair 5 | Discharge Rate_1 - Discharge Rate_3 | Cohen's d | .0566648001 | .258 | .076 |
| | | Hedges' correction | .0568426495 | .257 | .076 |
| Pair 6 | Discharge Rate_2 - Discharge Rate_3 | Cohen's d | .1124257861 | .123 | -.056 |
| | | Hedges' correction | .1127786483 | .122 | -.056 |

**Paired Samples Effect Sizes**

| | | | 95% ... Upper |
|---|---|---|---|
| Pair 1 | COVID-19_CFR_1 - COVID-19_CFR_2 | Cohen's d | -.024 |
| | | Hedges' correction | -.024 |
| Pair 2 | COVID-19_CFR_1 - COVID-19_CFR_3 | Cohen's d | .107 |
| | | Hedges' correction | .106 |
| Pair 3 | COVID-19_CFR_2 - COVID-19_CFR_3 | Cohen's d | .312 |
| | | Hedges' correction | .311 |
| Pair 4 | Discharge Rate_1 - Discharge Rate_2 | Cohen's d | .235 |
| | | Hedges' correction | .235 |
| Pair 5 | Discharge Rate_1 - Discharge Rate_3 | Cohen's d | .438 |
| | | Hedges' correction | .437 |
| Pair 6 | Discharge Rate_2 - Discharge Rate_3 | Cohen's d | .301 |
| | | Hedges' correction | .300 |

a. The denominator used in estimating the effect sizes.
Cohen's d uses the sample standard deviation of the mean difference.
Hedges' correction uses the sample standard deviation of the mean difference, plus a correction factor.
