## Supplementary material for "The Impact of SARS-CoV-2 Lineages (Variants) on the COVID-19 Epidemic in South Africa": SPSS_Program_(COVID-19 Hospitalised Deaths Age Profile)

```
 DATASET ACTIVATE DataSet1.

SAVE OUTFILE='C:\Users\ThaboMabuka\Google Drive\ARI Projects\Research Projects\COVID-19 i
n '+
    'Africa\Papers\ACMRG\The Impact of SARS-CoV-2 Variants on the COVID-19 Epidemic in So
uth '+
    'Africa\Data\Hospital Deaths\P2_Analysis_(COVID-19 Hospitalised Deaths Age Profile).s
av'
  /COMPRESSED.
SORT CASES  BY EpidemicWave.
SPLIT FILE LAYERED BY EpidemicWave.
DESCRIPTIVES VARIABLES=COVID19_Hospital_Deaths_Age_09 COVID19_Hospital_Deaths_Age_1019
    COVID19_Hospital_Deaths_Age_2029 COVID19_Hospital_Deaths_Age_3039 COVID19_Hospital_De
aths_Age_4049
    COVID19_Hospital_Deaths_Age_5059 COVID19_Hospital_Deaths_Age_6069 COVID19_Hospital_De
aths_Age_7079
    COVID19_Hospital_Deaths_Age_8089
  /STATISTICS=MEAN STDDEV MIN MAX.
```

### Descriptives

#### Notes

| | | |
|---|---|---|
| Output Created | | 14-OCT-2021 18:42:10 |
| Comments | | |
| Input | Data | C:\Users\ThaboMabuka\Google Drive\ARI Projects\Research Projects\COVID-19 in Africa\Papers\ACMRG\The Impact of SARS-CoV-2 Variants on the COVID-19 Epidemic in South Africa\Data\Hospital Deaths\P2_Analysis_(COVID-19 Hospitalised Deaths Age Profile).sav |
| | Active Dataset | DataSet1 |
| | Filter | <none> |
| | Weight | <none> |
| | Split File | Epidemic Wave |
| | N of Rows in Working Data File | 484 |

### Notes

| | | |
|---|---|---|
| Missing Value Handling | Definition of Missing | User defined missing values are treated as missing. |
| | Cases Used | All non-missing data are used. |
| Syntax | | DESCRIPTIVES VARIABLES=COVID19_Hospital_Deaths_Age_09 COVID19_Hospital_Deaths_Age_1019 COVID19_Hospital_Deaths_Age_2029 COVID19_Hospital_Deaths_Age_3039 COVID19_Hospital_Deaths_Age_4049 COVID19_Hospital_Deaths_Age_5059 COVID19_Hospital_Deaths_Age_6069 COVID19_Hospital_Deaths_Age_7079 COVID19_Hospital_Deaths_Age_8089 /STATISTICS=MEAN STDDEV MIN MAX. |
| Resources | Processor Time | 00:00:00,00 |
| | Elapsed Time | 00:00:00,00 |

### Descriptive Statistics

| Epidemic Wave | | N | Minimum | Maximum | Mean | Std. Deviation |
|---|---|---|---|---|---|---|
| 1 | COVID-19_Hospital_Deaths_Age_0-9 (%) | 126 | .00 | .64 | .1942 | .11128 |
| | COVID-19_Hospital_Deaths_Age_10-19 (%) | 126 | .00 | .34 | .2544 | .08934 |
| | COVID-19_Hospital_Deaths_Age_20-29 (%) | 126 | .00 | 2.26 | 1.3410 | .31224 |
| | COVID-19_Hospital_Deaths_Age_30-39 (%) | 126 | .73 | 6.33 | 5.2010 | .61878 |
| | COVID-19_Hospital_Deaths_Age_40-49 (%) | 126 | 5.13 | 13.32 | 11.7260 | .89310 |
| | COVID-19_Hospital_Deaths_Age_50-59 (%) | 125 | .00 | 27.91 | 14.7614 | 12.00757 |
| | COVID-19_Hospital_Deaths_Age_60-69 (%) | 126 | .00 | 28.86 | 25.8210 | 2.74159 |
| | COVID-19_Hospital_Deaths_Age_70-79 (%) | 126 | .00 | 19.83 | 18.1087 | 2.18075 |
| | COVID-19_Hospital_Deaths_Age_80-89(%) | 126 | .00 | 13.96 | 11.7430 | 2.28145 |
| | Valid N (listwise) | 125 | | | | |
| 2 | COVID-19_Hospital_Deaths_Age_0-9 (%) | 187 | .00 | .44 | .2502 | .09471 |
| | COVID-19_Hospital_Deaths_Age_10-19 (%) | 187 | .00 | 1.25 | .3979 | .13191 |
| | COVID-19_Hospital_Deaths_Age_20-29 (%) | 187 | .00 | 3.81 | 1.9649 | .49953 |
| | COVID-19_Hospital_Deaths_Age_30-39 (%) | 187 | 5.10 | 7.27 | 5.6677 | .53078 |
| | COVID-19_Hospital_Deaths_Age_40-49 (%) | 187 | 9.81 | 11.63 | 10.8030 | .31590 |

### Descriptive Statistics

| Epidemic Wave | | N | Minimum | Maximum | Mean | Std. Deviation |
|---|---|---|---|---|---|---|
| | COVID-19_Hospital_Deaths_Age_50-59 (%) | 162 | .00 | 98.20 | 41.2426 | 20.65819 |
| | COVID-19_Hospital_Deaths_Age_60-69 (%) | 187 | .00 | 31.19 | 28.5435 | 4.27460 |
| | COVID-19_Hospital_Deaths_Age_70-79 (%) | 187 | .00 | 21.31 | 20.0140 | 3.11582 |
| | COVID-19_Hospital_Deaths_Age_80-89(%) | 187 | .00 | 23.75 | 10.6127 | 1.94063 |
| | Valid N (listwise) | 162 | | | | |
| 3 | COVID-19_Hospital_Deaths_Age_0-9 (%) | 146 | .00 | 75.00 | .9437 | 6.17647 |
| | COVID-19_Hospital_Deaths_Age_10-19 (%) | 146 | .00 | .31 | .2167 | .07356 |
| | COVID-19_Hospital_Deaths_Age_20-29 (%) | 146 | .00 | 1.61 | .9903 | .29675 |
| | COVID-19_Hospital_Deaths_Age_30-39 (%) | 146 | 1.96 | 25.00 | 4.6036 | 2.01891 |
| | COVID-19_Hospital_Deaths_Age_40-49 (%) | 146 | .00 | 10.38 | 7.8210 | 1.97488 |
| | COVID-19_Hospital_Deaths_Age_50-59 (%) | 146 | 8.39 | 25.00 | 18.0521 | 3.97176 |
| | COVID-19_Hospital_Deaths_Age_60-69 (%) | 146 | .00 | 36.97 | 25.5792 | 6.93017 |
| | COVID-19_Hospital_Deaths_Age_70-79 (%) | 146 | 8.54 | 50.00 | 21.2839 | 5.43116 |
| | COVID-19_Hospital_Deaths_Age_80-89(%) | 146 | 5.41 | 25.00 | 13.4049 | 3.24290 |
| | Valid N (listwise) | 146 | | | | |

```
SPLIT FILE OFF.
```

```
* Chart Builder.
GGRAPH
  /GRAPHDATASET NAME="graphdataset" VARIABLES=EpidemicWave MEANCI(COVID19_Hospital_Deaths
_Age_09,
    95) MEANCI(COVID19_Hospital_Deaths_Age_1019, 95) MEANCI(COVID19_Hospital_Deaths_Age_2
029, 95)
    MEANCI(COVID19_Hospital_Deaths_Age_3039, 95) MEANCI(COVID19_Hospital_Deaths_Age_4049,
 95)
    MEANCI(COVID19_Hospital_Deaths_Age_5059, 95) MEANCI(COVID19_Hospital_Deaths_Age_6069,
 95)
    MEANCI(COVID19_Hospital_Deaths_Age_7079, 95) MEANCI(COVID19_Hospital_Deaths_Age_8089,
 95)
    MEANCI(COVID19_Hospital_Deaths_Age_9099, 95) MISSING=LISTWISE REPORTMISSING=NO
    TRANSFORM=VARSTOCASES(SUMMARY="#SUMMARY" INDEX="#INDEX" LOW="#LOW" HIGH="#HIGH")
  /GRAPHSPEC SOURCE=INLINE.
BEGIN GPL
  SOURCE: s=userSource(id("graphdataset"))
  DATA: EpidemicWave=col(source(s), name("EpidemicWave"), unit.category())
  DATA: SUMMARY=col(source(s), name("#SUMMARY"))
  DATA: INDEX=col(source(s), name("#INDEX"), unit.category())
  DATA: LOW=col(source(s), name("#LOW"))
  DATA: HIGH=col(source(s), name("#HIGH"))
  COORD: rect(dim(1,2), cluster(3,0))
  GUIDE: axis(dim(3), label("Epidemic Wave"))
  GUIDE: axis(dim(2), label("Mean"))
  GUIDE: legend(aesthetic(aesthetic.color.interior), label(""))
  GUIDE: text.title(label("Simple Bar Mean of COVID-19_Hospital_Deaths_Age_0-9 (%), Mean
of ",
    "COVID-19_Hospital_Deaths_Age_10-19 (%), Mean of COVID-19_Hospital_Deaths_Age_20-29 (
%), Mean ",
    "of COVID-19_Hospital_Deaths_Age_30-39 (%), Mean of COVID-19_Hospital_Deaths_Age_40-4
9 (%), ",
    "Mean of COVID-19_Hospital_Deaths_Age_50-59 (%), Mean of COVID-19_Hospital_Deaths_Age
_60-69 ",
    "(%), Mean of COVID-19_Hospital_Deaths_Age_70-79 (%), Mean of ",
    "COVID-19_Hospital_Deaths_Age_80-89(%), Mean of COVID-19_Hospital_Deaths_Age_90-99 (%
) by ",
    "Epidemic Wave by INDEX"))
  GUIDE: text.footnote(label("Error Bars: 95% CI"))
  SCALE: linear(dim(2), include(0))
  SCALE: cat(aesthetic(aesthetic.color.interior), include("0", "1", "2", "3", "4", "5", "
6",
    "7", "8", "9"))
  SCALE: cat(dim(1), include("0", "1", "2", "3", "4", "5", "6", "7", "8", "9"))
  ELEMENT: interval(position(INDEX*SUMMARY*EpidemicWave), color.interior(INDEX),
    shape.interior(shape.square))
  ELEMENT: interval(position(region.spread.range(INDEX*(LOW+HIGH)*EpidemicWave)),
    shape.interior(shape.ibeam))
END GPL.
```

### GGraph

#### Notes

| | | |
|---|---|---|
| Output Created | | 14-OCT-2021 18:43:40 |
| Comments | | |
| Input | Data | C:\Users\ThaboMabuka\Google Drive\ARI Projects\Research Projects\COVID-19 in Africa\Papers\ACMRG\The Impact of SARS-CoV-2 Variants on the COVID-19 Epidemic in South Africa\Data\Hospital Deaths\P2_Analysis_(COVID-19 Hospitalised Deaths Age Profile).sav |
| | Active Dataset | DataSet1 |
| | Filter | <none> |
| | Weight | <none> |
| | Split File | <none> |
| | N of Rows in Working Data File | 484 |

### Notes

| | |
|---|---|
| Syntax | GGRAPH<br> /GRAPHDATASET<br>NAME="graphdataset"<br>VARIABLES=EpidemicWa<br>ve MEANCI<br>(COVID19_Hospital_Deat<br>hs_Age_09,<br> 95) MEANCI<br>(COVID19_Hospital_Deat<br>hs_Age_1019, 95)<br>MEANCI<br>(COVID19_Hospital_Deat<br>hs_Age_2029, 95)<br> MEANCI<br>(COVID19_Hospital_Deat<br>hs_Age_3039, 95)<br>MEANCI<br>(COVID19_Hospital_Deat<br>hs_Age_4049, 95)<br> MEANCI<br>(COVID19_Hospital_Deat<br>hs_Age_5059, 95)<br>MEANCI<br>(COVID19_Hospital_Deat<br>hs_Age_6069, 95)<br> MEANCI<br>(COVID19_Hospital_Deat<br>hs_Age_7079, 95)<br>MEANCI<br>(COVID19_Hospital_Deat<br>hs_Age_8089, 95)<br> MEANCI<br>(COVID19_Hospital_Deat<br>hs_Age_9099, 95)<br>MISSING=LISTWISE<br>REPORTMISSING=NO<br><br>TRANSFORM=VARSTOC<br>ASES(SUMMARY="<br>#SUMMARY" INDEX="<br>#INDEX" LOW="#LOW"<br>HIGH="#HIGH")<br> /GRAPHSPEC<br>SOURCE=INLINE.<br>BEGIN GPL<br> SOURCE: s=userSource<br>(id("graphdataset"))<br> DATA:<br>EpidemicWave=col(source<br>(s), name<br>("EpidemicWave"), unit.<br>category())<br> DATA: SUMMARY=col<br>(source(s), name<br>("#SUMMARY"))<br> DATA: INDEX=col<br>(source(s), name<br>("#INDEX"), unit.<br>category())<br> DATA: LOW=col(source<br>(s), name("#LOW"))<br> DATA: HIGH=col(source |

### Notes

| | | |
|---|---|---|
| Resources | Processor Time | 00:00:00,25 |
| | Elapsed Time | 00:00:00,20 |

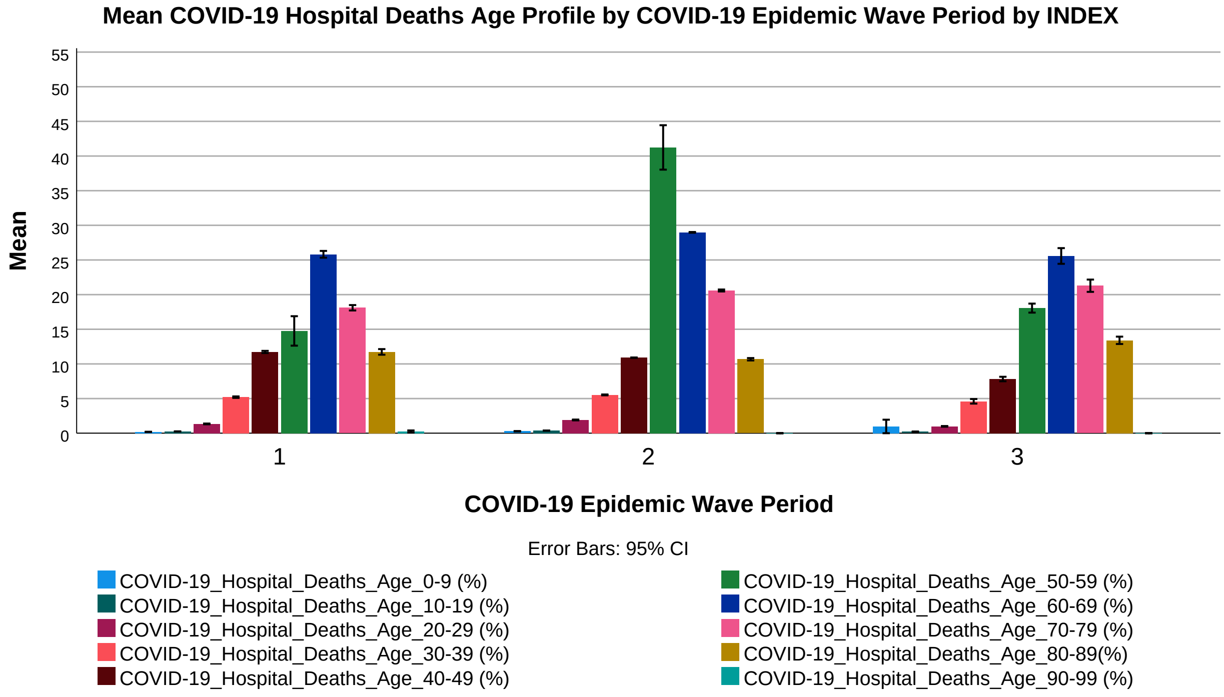


```
 GET
  FILE='C:\Users\ThaboMabuka\Google Drive\ARI Projects\Research Projects\COVID-19 in Afri
ca\Papers\ACMRG\The Impact of SARS-CoV-2 Variants on the COVID-19 Epidemic in South Afric
a\Data\Hospital Deaths\P2_Analysis_(COVID-19 Hospitalised Deaths Age Profile)_1.sav'.
DATASET NAME DataSet2 WINDOW=FRONT.
DATASET ACTIVATE DataSet1.
DATASET CLOSE DataSet2.
GET
  FILE='C:\Users\ThaboMabuka\Google Drive\ARI Projects\Research Projects\COVID-19 in Afri
ca\Papers\ACMRG\The Impact of SARS-CoV-2 Variants on the COVID-19 Epidemic in South Afric
a\Data\Hospital Deaths\P2_Analysis_(COVID-19 Hospitalised Deaths Age Profile)_1.sav'.
DATASET NAME DataSet3 WINDOW=FRONT.
DATASET ACTIVATE DataSet3.

SAVE OUTFILE='C:\Users\ThaboMabuka\Google Drive\ARI Projects\Research Projects\COVID-19 i
n '+
    'Africa\Papers\ACMRG\The Impact of SARS-CoV-2 Variants on the COVID-19 Epidemic in So
uth '+
    'Africa\Data\Hospital Deaths\P2_Analysis_(COVID-19 Hospitalised Deaths Age Profile)_1
```

```
.sav'
  /COMPRESSED.
DATASET ACTIVATE DataSet1.
DATASET CLOSE DataSet3.
GET
  FILE='C:\Users\ThaboMabuka\Google Drive\ARI Projects\Research Projects\COVID-19 in Afri
ca\Papers\ACMRG\The Impact of SARS-CoV-2 Variants on the COVID-19 Epidemic in South Afric
a\Data\Hospital Deaths\P2_Analysis_(COVID-19 Hospitalised Deaths Age Profile)_2.sav'.
DATASET NAME DataSet4 WINDOW=FRONT.
DATASET ACTIVATE DataSet4.

SAVE OUTFILE='C:\Users\ThaboMabuka\Google Drive\ARI Projects\Research Projects\COVID-19 i
n '+
    'Africa\Papers\ACMRG\The Impact of SARS-CoV-2 Variants on the COVID-19 Epidemic in So
uth '+
    'Africa\Data\Hospital Deaths\P2_Analysis_(COVID-19 Hospitalised Deaths Age Profile)_2
.sav'
  /COMPRESSED.
DATASET ACTIVATE DataSet1.
DATASET CLOSE DataSet4.
GET
  FILE='C:\Users\ThaboMabuka\Google Drive\ARI Projects\Research Projects\COVID-19 in Afri
ca\Papers\ACMRG\The Impact of SARS-CoV-2 Variants on the COVID-19 Epidemic in South Afric
a\Data\Hospital Deaths\P2_Analysis_(COVID-19 Hospitalised Deaths Age Profile)_3.sav'.
DATASET NAME DataSet5 WINDOW=FRONT.
DATASET ACTIVATE DataSet5.
DATASET ACTIVATE DataSet5.

SAVE OUTFILE='C:\Users\ThaboMabuka\Google Drive\ARI Projects\Research Projects\COVID-19 i
n '+
    'Africa\Papers\ACMRG\The Impact of SARS-CoV-2 Variants on the COVID-19 Epidemic in So
uth '+
    'Africa\Data\Hospital Deaths\P2_Analysis_(COVID-19 Hospitalised Deaths Age Profile)_3
.sav'
  /COMPRESSED.
DATASET ACTIVATE DataSet1.
DATASET CLOSE DataSet5.



 T-TEST PAIRS=COVID19_Hospital_Deaths_Age_09_1 COVID19_Hospital_Deaths_Age_09_1
    COVID19_Hospital_Deaths_Age_09_2 COVID19_Hospital_Deaths_Age_1019_1
    COVID19_Hospital_Deaths_Age_1019_1 COVID19_Hospital_Deaths_Age_1019_2
    COVID19_Hospital_Deaths_Age_2029_1 COVID19_Hospital_Deaths_Age_2029_1
    COVID19_Hospital_Deaths_Age_2029_2 COVID19_Hospital_Deaths_Age_3039_1
    COVID19_Hospital_Deaths_Age_3039_1 COVID19_Hospital_Deaths_Age_3039_2
    COVID19_Hospital_Deaths_Age_4049_1 COVID19_Hospital_Deaths_Age_4049_1
    COVID19_Hospital_Deaths_Age_4049_2 COVID19_Hospital_Deaths_Age_5059_1
    COVID19_Hospital_Deaths_Age_5059_1 COVID19_Hospital_Deaths_Age_5059_2
```

```
  COVID19_Hospital_Deaths_Age_6069_1 COVID19_Hospital_Deaths_Age_6069_1
  COVID19_Hospital_Deaths_Age_6069_2 COVID19_Hospital_Deaths_Age_7079_1
  COVID19_Hospital_Deaths_Age_7079_1 COVID19_Hospital_Deaths_Age_7079_2
  COVID19_Hospital_Deaths_Age_8089_1 COVID19_Hospital_Deaths_Age_8089_1
  COVID19_Hospital_Deaths_Age_8089_2 WITH COVID19_Hospital_Deaths_Age_09_2
  COVID19_Hospital_Deaths_Age_09_3 COVID19_Hospital_Deaths_Age_09_3
  COVID19_Hospital_Deaths_Age_1019_2 COVID19_Hospital_Deaths_Age_1019_3
  COVID19_Hospital_Deaths_Age_1019_3 COVID19_Hospital_Deaths_Age_2029_2
  COVID19_Hospital_Deaths_Age_2029_3 COVID19_Hospital_Deaths_Age_2029_3
  COVID19_Hospital_Deaths_Age_3039_2 COVID19_Hospital_Deaths_Age_3039_3
  COVID19_Hospital_Deaths_Age_3039_3 COVID19_Hospital_Deaths_Age_4049_2
  COVID19_Hospital_Deaths_Age_4049_3 COVID19_Hospital_Deaths_Age_4049_3
  COVID19_Hospital_Deaths_Age_5059_2 COVID19_Hospital_Deaths_Age_5059_3
  COVID19_Hospital_Deaths_Age_5059_3 COVID19_Hospital_Deaths_Age_6069_2
  COVID19_Hospital_Deaths_Age_6069_3 COVID19_Hospital_Deaths_Age_6069_3
  COVID19_Hospital_Deaths_Age_7079_2 COVID19_Hospital_Deaths_Age_7079_3
  COVID19_Hospital_Deaths_Age_7079_3 COVID19_Hospital_Deaths_Age_8089_2
  COVID19_Hospital_Deaths_Age_8089_3 COVID19_Hospital_Deaths_Age_8089_3 (PAIRED)
/ES DISPLAY(TRUE) STANDARDIZER(SD)
/CRITERIA=CI(.9500)
/MISSING=ANALYSIS.
```

### T-Test

#### Notes

| | | |
|---|---|---|
| Output Created | | 14-OCT-2021 19:30:20 |
| Comments | | |
| Input | Data | C:\Users\ThaboMabuka\Google Drive\ARI Projects\Research Projects\COVID-19 in Africa\Papers\ACMRG\The Impact of SARS-CoV-2 Variants on the COVID-19 Epidemic in South Africa\Data\Hospital Deaths\P2_Analysis_(COVID-19 Hospitalised Deaths Age Profile)_M.sav |
| | Active Dataset | DataSet1 |
| | Filter | <none> |
| | Weight | <none> |
| | Split File | <none> |
| | N of Rows in Working Data File | 208 |

**Notes**

| | | |
|---|---|---|
| Missing Value Handling | Definition of Missing | User defined missing values are treated as missing. |
| | Cases Used | Statistics for each analysis are based on the cases with no missing or out-of-range data for any variable in the analysis. |

### Notes

| | |
|---|---|
| Syntax | T-TEST<br>PAIRS=COVID19_Hospita<br>l_Deaths_Age_09_1<br>COVID19_Hospital_Death<br>s_Age_09_1<br><br>COVID19_Hospital_Death<br>s_Age_09_2<br>COVID19_Hospital_Death<br>s_Age_1019_1<br><br>COVID19_Hospital_Death<br>s_Age_1019_1<br>COVID19_Hospital_Death<br>s_Age_1019_2<br><br>COVID19_Hospital_Death<br>s_Age_2029_1<br>COVID19_Hospital_Death<br>s_Age_2029_1<br><br>COVID19_Hospital_Death<br>s_Age_2029_2<br>COVID19_Hospital_Death<br>s_Age_3039_1<br><br>COVID19_Hospital_Death<br>s_Age_3039_1<br>COVID19_Hospital_Death<br>s_Age_3039_2<br><br>COVID19_Hospital_Death<br>s_Age_4049_1<br>COVID19_Hospital_Death<br>s_Age_4049_1<br><br>COVID19_Hospital_Death<br>s_Age_4049_2<br>COVID19_Hospital_Death<br>s_Age_5059_1<br><br>COVID19_Hospital_Death<br>s_Age_5059_1<br>COVID19_Hospital_Death<br>s_Age_5059_2<br><br>COVID19_Hospital_Death<br>s_Age_6069_1<br>COVID19_Hospital_Death<br>s_Age_6069_1<br><br>COVID19_Hospital_Death<br>s_Age_6069_2<br>COVID19_Hospital_Death<br>s_Age_7079_1<br><br>COVID19_Hospital_Death<br>s_Age_7079_1<br>COVID19_Hospital_Death<br>s_Age_7079_2<br><br>COVID19_Hospital_Death |

**Notes**

| | | |
|---|---|---|
| Resources | Processor Time | 00:00:00,05 |
| | Elapsed Time | 00:00:00,03 |

[DataSet1] C:\Users\ThaboMabuka\Google Drive\ARI Projects\Research Projects\COVID-19 in Africa\Papers\ACMRG\The Impact of SARS-CoV-2 Variants on the COVID-19 Epidemic in South Africa\Data\Hospital Deaths\P2_Analysis_(COVID-19 Hospitalised Deaths Age Profile)_M.sav

**Paired Samples Statistics**

| | | Mean | N | Std. Deviation | Std. Error Mean |
|---|---|---|---|---|---|
| Pair 1 | COVID-19_Hospital_Deaths_Age_0-9 (%)_1 | .2016 | 106 | .11412 | .01108 |
| | COVID-19_Hospital_Deaths_Age_0-9 (%)_2 | .2240 | 106 | .11500 | .01117 |
| Pair 2 | COVID-19_Hospital_Deaths_Age_0-9 (%)_1 | .1942 | 126 | .11128 | .00991 |
| | COVID-19_Hospital_Deaths_Age_0-9 (%)_3 | 1.0616 | 126 | 6.64438 | .59193 |
| Pair 3 | COVID-19_Hospital_Deaths_Age_0-9 (%)_2 | .2337 | 125 | .11172 | .00999 |
| | COVID-19_Hospital_Deaths_Age_0-9 (%)_3 | 1.0466 | 125 | 6.67329 | .59688 |
| Pair 4 | COVID-19_Hospital_Deaths_Age_10-19 (%)_1 | .2558 | 106 | .09506 | .00923 |
| | COVID-19_Hospital_Deaths_Age_10-19 (%)_2 | .4260 | 106 | .16241 | .01577 |
| Pair 5 | COVID-19_Hospital_Deaths_Age_10-19 (%)_1 | .2544 | 126 | .08934 | .00796 |
| | COVID-19_Hospital_Deaths_Age_10-19 (%)_3 | .2275 | 126 | .07066 | .00630 |

### Paired Samples Statistics

| | | Mean | N | Std. Deviation | Std. Error Mean |
|---|---|---|---|---|---|
| Pair 6 | COVID-19_Hospital_Deaths_Age_10-19 (%)_2 | .4091 | 125 | .15618 | .01397 |
| | COVID-19_Hospital_Deaths_Age_10-19 (%)_3 | .2229 | 125 | .07387 | .00661 |
| Pair 7 | COVID-19_Hospital_Deaths_Age_20-29 (%)_1 | 1.3625 | 106 | .31953 | .03104 |
| | COVID-19_Hospital_Deaths_Age_20-29 (%)_2 | 2.1169 | 106 | .61127 | .05937 |
| Pair 8 | COVID-19_Hospital_Deaths_Age_20-29 (%)_1 | 1.3410 | 126 | .31224 | .02782 |
| | COVID-19_Hospital_Deaths_Age_20-29 (%)_3 | 1.0356 | 126 | .26866 | .02393 |
| Pair 9 | COVID-19_Hospital_Deaths_Age_20-29 (%)_2 | 2.0478 | 125 | .59282 | .05302 |
| | COVID-19_Hospital_Deaths_Age_20-29 (%)_3 | .9625 | 125 | .28987 | .02593 |
| Pair 10 | COVID-19_Hospital_Deaths_Age_30-39 (%)_1 | 5.1402 | 106 | .64107 | .06227 |
| | COVID-19_Hospital_Deaths_Age_30-39 (%)_2 | 5.9239 | 106 | .56684 | .05506 |
| Pair 11 | COVID-19_Hospital_Deaths_Age_30-39 (%)_1 | 5.2010 | 126 | .61878 | .05513 |
| | COVID-19_Hospital_Deaths_Age_30-39 (%)_3 | 4.9066 | 126 | 1.94482 | .17326 |
| Pair 12 | COVID-19_Hospital_Deaths_Age_30-39 (%)_2 | 5.8211 | 125 | .59096 | .05286 |
| | COVID-19_Hospital_Deaths_Age_30-39 (%)_3 | 4.4468 | 125 | 2.13216 | .19071 |

**Paired Samples Statistics**

| | | Mean | N | Std. Deviation | Std. Error Mean |
|---|---|---|---|---|---|
| Pair 13 | COVID-19_Hospital_Deaths_Age_40-49 (%)_1 | 11.6042 | 106 | .87555 | .08504 |
| | COVID-19_Hospital_Deaths_Age_40-49 (%)_2 | 10.7569 | 106 | .40533 | .03937 |
| Pair 14 | COVID-19_Hospital_Deaths_Age_40-49 (%)_1 | 11.7260 | 126 | .89310 | .07956 |
| | COVID-19_Hospital_Deaths_Age_40-49 (%)_3 | 8.2463 | 126 | 1.60215 | .14273 |
| Pair 15 | COVID-19_Hospital_Deaths_Age_40-49 (%)_2 | 10.7668 | 125 | .38069 | .03405 |
| | COVID-19_Hospital_Deaths_Age_40-49 (%)_3 | 7.8911 | 125 | 2.09913 | .18775 |
| Pair 16 | COVID-19_Hospital_Deaths_Age_50-59 (%)_1 | 9.7222 | 81 | 11.84023 | 1.31558 |
| | COVID-19_Hospital_Deaths_Age_50-59 (%)_2 | 53.1409 | 81 | 23.67716 | 2.63080 |
| Pair 17 | COVID-19_Hospital_Deaths_Age_50-59 (%)_1 | 14.7614 | 125 | 12.00757 | 1.07399 |
| | COVID-19_Hospital_Deaths_Age_50-59 (%)_3 | 19.1764 | 125 | 2.37774 | .21267 |
| Pair 18 | COVID-19_Hospital_Deaths_Age_50-59 (%)_2 | 49.0835 | 100 | 23.05747 | 2.30575 |
| | COVID-19_Hospital_Deaths_Age_50-59 (%)_3 | 18.0041 | 100 | 4.73654 | .47365 |
| Pair 19 | COVID-19_Hospital_Deaths_Age_60-69 (%)_1 | 25.5944 | 106 | 2.92808 | .28440 |
| | COVID-19_Hospital_Deaths_Age_60-69 (%)_2 | 28.3219 | 106 | 5.67557 | .55126 |

### Paired Samples Statistics

| | | Mean | N | Std. Deviation | Std. Error Mean |
|---|---|---|---|---|---|
| Pair 20 | COVID-19_Hospital_Deaths_Age_60-69 (%)_1 | 25.8210 | 126 | 2.74159 | .24424 |
| | COVID-19_Hospital_Deaths_Age_60-69 (%)_3 | 27.4823 | 126 | 4.53710 | .40420 |
| Pair 21 | COVID-19_Hospital_Deaths_Age_60-69 (%)_2 | 28.4272 | 125 | 5.23116 | .46789 |
| | COVID-19_Hospital_Deaths_Age_60-69 (%)_3 | 24.7546 | 125 | 7.13520 | .63819 |
| Pair 22 | COVID-19_Hospital_Deaths_Age_70-79 (%)_1 | 18.2120 | 106 | 2.34886 | .22814 |
| | COVID-19_Hospital_Deaths_Age_70-79 (%)_2 | 19.2431 | 106 | 3.97199 | .38579 |
| Pair 23 | COVID-19_Hospital_Deaths_Age_70-79 (%)_1 | 18.1087 | 126 | 2.18075 | .19428 |
| | COVID-19_Hospital_Deaths_Age_70-79 (%)_3 | 22.8384 | 126 | 3.43194 | .30574 |
| Pair 24 | COVID-19_Hospital_Deaths_Age_70-79 (%)_2 | 19.5230 | 125 | 3.71853 | .33260 |
| | COVID-19_Hospital_Deaths_Age_70-79 (%)_3 | 21.0478 | 125 | 5.83344 | .52176 |
| Pair 25 | COVID-19_Hospital_Deaths_Age_80-89(%)_1 | 12.0930 | 106 | 2.22568 | .21618 |
| | COVID-19_Hospital_Deaths_Age_80-89(%)_2 | 10.4360 | 106 | 2.56499 | .24913 |
| Pair 26 | COVID-19_Hospital_Deaths_Age_80-89(%)_1 | 11.7430 | 126 | 2.28145 | .20325 |
| | COVID-19_Hospital_Deaths_Age_80-89(%)_3 | 14.3756 | 126 | 1.85370 | .16514 |

### Paired Samples Statistics

| | | Mean | N | Std. Deviation | Std. Error Mean |
|---|---|---|---|---|---|
| Pair 27 | COVID-19_Hospital_Deaths_Age_80-89(%)_2 | 10.4987 | 125 | 2.36835 | .21183 |
| | COVID-19_Hospital_Deaths_Age_80-89(%)_3 | 13.3440 | 125 | 3.49355 | .31247 |

### Paired Samples Correlations

| | | N | Correlation | Sig. |
|---|---|---|---|---|
| Pair 1 | COVID-19_Hospital_Deaths_Age_0-9 (%)_1 & COVID-19_Hospital_Deaths_Age_0-9 (%)_2 | 106 | .014 | .885 |
| Pair 2 | COVID-19_Hospital_Deaths_Age_0-9 (%)_1 & COVID-19_Hospital_Deaths_Age_0-9 (%)_3 | 126 | .260 | .003 |
| Pair 3 | COVID-19_Hospital_Deaths_Age_0-9 (%)_2 & COVID-19_Hospital_Deaths_Age_0-9 (%)_3 | 125 | -.209 | .019 |
| Pair 4 | COVID-19_Hospital_Deaths_Age_10-19 (%)_1 & COVID-19_Hospital_Deaths_Age_10-19 (%)_2 | 106 | -.487 | .000 |
| Pair 5 | COVID-19_Hospital_Deaths_Age_10-19 (%)_1 & COVID-19_Hospital_Deaths_Age_10-19 (%)_3 | 126 | .366 | .000 |
| Pair 6 | COVID-19_Hospital_Deaths_Age_10-19 (%)_2 & COVID-19_Hospital_Deaths_Age_10-19 (%)_3 | 125 | -.447 | .000 |
| Pair 7 | COVID-19_Hospital_Deaths_Age_20-29 (%)_1 & COVID-19_Hospital_Deaths_Age_20-29 (%)_2 | 106 | .526 | .000 |

**Paired Samples Correlations**

| | | N | Correlation | Sig. |
|---|---|---|---|---|
| Pair 8 | COVID-19_Hospital_Deaths_Age_20-29 (%)_1 & COVID-19_Hospital_Deaths_Age_20-29 (%)_3 | 126 | .241 | .006 |
| Pair 9 | COVID-19_Hospital_Deaths_Age_20-29 (%)_2 & COVID-19_Hospital_Deaths_Age_20-29 (%)_3 | 125 | .165 | .067 |
| Pair 10 | COVID-19_Hospital_Deaths_Age_30-39 (%)_1 & COVID-19_Hospital_Deaths_Age_30-39 (%)_2 | 106 | .223 | .022 |
| Pair 11 | COVID-19_Hospital_Deaths_Age_30-39 (%)_1 & COVID-19_Hospital_Deaths_Age_30-39 (%)_3 | 126 | .055 | .538 |
| Pair 12 | COVID-19_Hospital_Deaths_Age_30-39 (%)_2 & COVID-19_Hospital_Deaths_Age_30-39 (%)_3 | 125 | .310 | .000 |
| Pair 13 | COVID-19_Hospital_Deaths_Age_40-49 (%)_1 & COVID-19_Hospital_Deaths_Age_40-49 (%)_2 | 106 | -.003 | .979 |
| Pair 14 | COVID-19_Hospital_Deaths_Age_40-49 (%)_1 & COVID-19_Hospital_Deaths_Age_40-49 (%)_3 | 126 | .052 | .562 |
| Pair 15 | COVID-19_Hospital_Deaths_Age_40-49 (%)_2 & COVID-19_Hospital_Deaths_Age_40-49 (%)_3 | 125 | .230 | .010 |
| Pair 16 | COVID-19_Hospital_Deaths_Age_50-59 (%)_1 & COVID-19_Hospital_Deaths_Age_50-59 (%)_2 | 81 | -.082 | .469 |

### Paired Samples Correlations

| | | N | Correlation | Sig. |
|---|---|---|---|---|
| Pair 17 | COVID-19_Hospital_Deaths_Age_50-59 (%)_1 & COVID-19_Hospital_Deaths_Age_50-59 (%)_3 | 125 | -.149 | .096 |
| Pair 18 | COVID-19_Hospital_Deaths_Age_50-59 (%)_2 & COVID-19_Hospital_Deaths_Age_50-59 (%)_3 | 100 | .196 | .051 |
| Pair 19 | COVID-19_Hospital_Deaths_Age_60-69 (%)_1 & COVID-19_Hospital_Deaths_Age_60-69 (%)_2 | 106 | -.183 | .061 |
| Pair 20 | COVID-19_Hospital_Deaths_Age_60-69 (%)_1 & COVID-19_Hospital_Deaths_Age_60-69 (%)_3 | 126 | -.094 | .293 |
| Pair 21 | COVID-19_Hospital_Deaths_Age_60-69 (%)_2 & COVID-19_Hospital_Deaths_Age_60-69 (%)_3 | 125 | -.150 | .096 |
| Pair 22 | COVID-19_Hospital_Deaths_Age_70-79 (%)_1 & COVID-19_Hospital_Deaths_Age_70-79 (%)_2 | 106 | .158 | .106 |
| Pair 23 | COVID-19_Hospital_Deaths_Age_70-79 (%)_1 & COVID-19_Hospital_Deaths_Age_70-79 (%)_3 | 126 | -.100 | .263 |
| Pair 24 | COVID-19_Hospital_Deaths_Age_70-79 (%)_2 & COVID-19_Hospital_Deaths_Age_70-79 (%)_3 | 125 | -.176 | .050 |
| Pair 25 | COVID-19_Hospital_Deaths_Age_80-89(%)_1 & COVID-19_Hospital_Deaths_Age_80-89(%)_2 | 106 | .075 | .444 |

### Paired Samples Correlations

| | | N | Correlation | Sig. |
|---|---|---|---|---|
| Pair 26 | COVID-19_Hospital_Deaths_Age_80-89(%)_1 & COVID-19_Hospital_Deaths_Age_80-89(%)_3 | 126 | .097 | .279 |
| Pair 27 | COVID-19_Hospital_Deaths_Age_80-89(%)_2 & COVID-19_Hospital_Deaths_Age_80-89(%)_3 | 125 | .094 | .298 |

### Paired Samples Test

| | | Paired Differences | | | |
|---|---|---|---|---|---|
| | | | | | 95% Confidence ... |
| | | Mean | Std. Deviation | Std. Error Mean | Lower |
| Pair 1 | COVID-19_Hospital_Deaths_Age_0-9 (%)_1 - COVID-19_Hospital_Deaths_Age_0-9 (%)_2 | -.02236 | .16085 | .01562 | -.05334 |
| Pair 2 | COVID-19_Hospital_Deaths_Age_0-9 (%)_1 - COVID-19_Hospital_Deaths_Age_0-9 (%)_3 | -.86738 | 6.61632 | .58943 | -2.03393 |
| Pair 3 | COVID-19_Hospital_Deaths_Age_0-9 (%)_2 - COVID-19_Hospital_Deaths_Age_0-9 (%)_3 | -.81288 | 6.69754 | .59905 | -1.99856 |
| Pair 4 | COVID-19_Hospital_Deaths_Age_10-19 (%)_1 - COVID-19_Hospital_Deaths_Age_10-19 (%)_2 | -.17028 | .22463 | .02182 | -.21354 |
| Pair 5 | COVID-19_Hospital_Deaths_Age_10-19 (%)_1 - COVID-19_Hospital_Deaths_Age_10-19 (%)_3 | .02690 | .09143 | .00815 | .01078 |
| Pair 6 | COVID-19_Hospital_Deaths_Age_10-19 (%)_2 - COVID-19_Hospital_Deaths_Age_10-19 (%)_3 | .18624 | .20038 | .01792 | .15077 |

### Paired Samples Test

| | | Paired ... 95% Confidence Interval of the ... Upper | t | df | Sig. (2-tailed) |
|---|---|---|---|---|---|
| Pair 1 | COVID-19_Hospital_Deaths_Age_0-9 (%)_1 - COVID-19_Hospital_Deaths_Age_0-9 (%)_2 | .00862 | -1.431 | 105 | .155 |
| Pair 2 | COVID-19_Hospital_Deaths_Age_0-9 (%)_1 - COVID-19_Hospital_Deaths_Age_0-9 (%)_3 | .29917 | -1.472 | 125 | .144 |
| Pair 3 | COVID-19_Hospital_Deaths_Age_0-9 (%)_2 - COVID-19_Hospital_Deaths_Age_0-9 (%)_3 | .37280 | -1.357 | 124 | .177 |
| Pair 4 | COVID-19_Hospital_Deaths_Age_10-19 (%)_1 - COVID-19_Hospital_Deaths_Age_10-19 (%)_2 | -.12702 | -7.805 | 105 | .000 |
| Pair 5 | COVID-19_Hospital_Deaths_Age_10-19 (%)_1 - COVID-19_Hospital_Deaths_Age_10-19 (%)_3 | .04302 | 3.303 | 125 | .001 |
| Pair 6 | COVID-19_Hospital_Deaths_Age_10-19 (%)_2 - COVID-19_Hospital_Deaths_Age_10-19 (%)_3 | .22171 | 10.392 | 124 | .000 |

### Paired Samples Test

| | | Paired Differences | | | |
|---|---|---|---|---|---|
| | | | | | 95% Confidence ... |
| | | Mean | Std. Deviation | Std. Error Mean | Lower |
| Pair 7 | COVID-19_Hospital_Deaths_Age_20-29 (%)_1 - COVID-19_Hospital_Deaths_Age_20-29 (%)_2 | -.75434 | .51974 | .05048 | -.85443 |
| Pair 8 | COVID-19_Hospital_Deaths_Age_20-29 (%)_1 - COVID-19_Hospital_Deaths_Age_20-29 (%)_3 | .30540 | .35939 | .03202 | .24203 |
| Pair 9 | COVID-19_Hospital_Deaths_Age_20-29 (%)_2 - COVID-19_Hospital_Deaths_Age_20-29 (%)_3 | 1.08528 | .61557 | .05506 | .97630 |
| Pair 10 | COVID-19_Hospital_Deaths_Age_30-39 (%)_1 - COVID-19_Hospital_Deaths_Age_30-39 (%)_2 | -.78368 | .75519 | .07335 | -.92912 |
| Pair 11 | COVID-19_Hospital_Deaths_Age_30-39 (%)_1 - COVID-19_Hospital_Deaths_Age_30-39 (%)_3 | .29437 | 2.00800 | .17889 | -.05967 |
| Pair 12 | COVID-19_Hospital_Deaths_Age_30-39 (%)_2 - COVID-19_Hospital_Deaths_Age_30-39 (%)_3 | 1.37432 | 2.02848 | .18143 | 1.01521 |
| Pair 13 | COVID-19_Hospital_Deaths_Age_40-49 (%)_1 - COVID-19_Hospital_Deaths_Age_40-49 (%)_2 | .84726 | .96578 | .09380 | .66127 |
| Pair 14 | COVID-19_Hospital_Deaths_Age_40-49 (%)_1 - COVID-19_Hospital_Deaths_Age_40-49 (%)_3 | 3.47968 | 1.79316 | .15975 | 3.16352 |

### Paired Samples Test

| | | Paired ... 95% Confidence Interval of the ... Upper | t | df | Sig. (2-tailed) |
|---|---|---|---|---|---|
| Pair 7 | COVID-19_Hospital_Deaths_Age_20-29 (%)_1 - COVID-19_Hospital_Deaths_Age_20-29 (%)_2 | -.65424 | -14.943 | 105 | .000 |
| Pair 8 | COVID-19_Hospital_Deaths_Age_20-29 (%)_1 - COVID-19_Hospital_Deaths_Age_20-29 (%)_3 | .36876 | 9.539 | 125 | .000 |
| Pair 9 | COVID-19_Hospital_Deaths_Age_20-29 (%)_2 - COVID-19_Hospital_Deaths_Age_20-29 (%)_3 | 1.19426 | 19.711 | 124 | .000 |
| Pair 10 | COVID-19_Hospital_Deaths_Age_30-39 (%)_1 - COVID-19_Hospital_Deaths_Age_30-39 (%)_2 | -.63824 | -10.684 | 105 | .000 |
| Pair 11 | COVID-19_Hospital_Deaths_Age_30-39 (%)_1 - COVID-19_Hospital_Deaths_Age_30-39 (%)_3 | .64840 | 1.646 | 125 | .102 |
| Pair 12 | COVID-19_Hospital_Deaths_Age_30-39 (%)_2 - COVID-19_Hospital_Deaths_Age_30-39 (%)_3 | 1.73343 | 7.575 | 124 | .000 |
| Pair 13 | COVID-19_Hospital_Deaths_Age_40-49 (%)_1 - COVID-19_Hospital_Deaths_Age_40-49 (%)_2 | 1.03326 | 9.032 | 105 | .000 |
| Pair 14 | COVID-19_Hospital_Deaths_Age_40-49 (%)_1 - COVID-19_Hospital_Deaths_Age_40-49 (%)_3 | 3.79584 | 21.782 | 125 | .000 |

### Paired Samples Test

| | | Paired Differences | | | |
|---|---|---|---|---|---|
| | | | | | 95% Confidence ... |
| | | Mean | Std. Deviation | Std. Error Mean | Lower |
| Pair 15 | COVID-19_Hospital_Deaths_Age_40-49 (%)_2 - COVID-19_Hospital_Deaths_Age_40-49 (%)_3 | 2.87568 | 2.04555 | .18296 | 2.51355 |
| Pair 16 | COVID-19_Hospital_Deaths_Age_50-59 (%)_1 - COVID-19_Hospital_Deaths_Age_50-59 (%)_2 | -43.41864 | 27.32211 | 3.03579 | -49.46006 |
| Pair 17 | COVID-19_Hospital_Deaths_Age_50-59 (%)_1 - COVID-19_Hospital_Deaths_Age_50-59 (%)_3 | -4.41504 | 12.58429 | 1.12557 | -6.64286 |
| Pair 18 | COVID-19_Hospital_Deaths_Age_50-59 (%)_2 - COVID-19_Hospital_Deaths_Age_50-59 (%)_3 | 31.07940 | 22.61123 | 2.26112 | 26.59284 |
| Pair 19 | COVID-19_Hospital_Deaths_Age_60-69 (%)_1 - COVID-19_Hospital_Deaths_Age_60-69 (%)_2 | -2.72745 | 6.84518 | .66486 | -4.04575 |
| Pair 20 | COVID-19_Hospital_Deaths_Age_60-69 (%)_1 - COVID-19_Hospital_Deaths_Age_60-69 (%)_3 | -1.66127 | 5.51823 | .49160 | -2.63421 |
| Pair 21 | COVID-19_Hospital_Deaths_Age_60-69 (%)_2 - COVID-19_Hospital_Deaths_Age_60-69 (%)_3 | 3.67256 | 9.45731 | .84589 | 1.99831 |
| Pair 22 | COVID-19_Hospital_Deaths_Age_70-79 (%)_1 - COVID-19_Hospital_Deaths_Age_70-79 (%)_2 | -1.03113 | 4.28348 | .41605 | -1.85608 |

### Paired Samples Test

| | | Paired ... 95% Confidence Interval of the ... Upper | t | df | Sig. (2-tailed) |
|---|---|---|---|---|---|
| Pair 15 | COVID-19_Hospital_Deaths_Age_40-49 (%)_2 - COVID-19_Hospital_Deaths_Age_40-49 (%)_3 | 3.23781 | 15.718 | 124 | .000 |
| Pair 16 | COVID-19_Hospital_Deaths_Age_50-59 (%)_1 - COVID-19_Hospital_Deaths_Age_50-59 (%)_2 | -37.37723 | -14.302 | 80 | .000 |
| Pair 17 | COVID-19_Hospital_Deaths_Age_50-59 (%)_1 - COVID-19_Hospital_Deaths_Age_50-59 (%)_3 | -2.18722 | -3.922 | 124 | .000 |
| Pair 18 | COVID-19_Hospital_Deaths_Age_50-59 (%)_2 - COVID-19_Hospital_Deaths_Age_50-59 (%)_3 | 35.56596 | 13.745 | 99 | .000 |
| Pair 19 | COVID-19_Hospital_Deaths_Age_60-69 (%)_1 - COVID-19_Hospital_Deaths_Age_60-69 (%)_2 | -1.40915 | -4.102 | 105 | .000 |
| Pair 20 | COVID-19_Hospital_Deaths_Age_60-69 (%)_1 - COVID-19_Hospital_Deaths_Age_60-69 (%)_3 | -.68833 | -3.379 | 125 | .001 |
| Pair 21 | COVID-19_Hospital_Deaths_Age_60-69 (%)_2 - COVID-19_Hospital_Deaths_Age_60-69 (%)_3 | 5.34681 | 4.342 | 124 | .000 |
| Pair 22 | COVID-19_Hospital_Deaths_Age_70-79 (%)_1 - COVID-19_Hospital_Deaths_Age_70-79 (%)_2 | -.20619 | -2.478 | 105 | .015 |

### Paired Samples Test

| | | Paired Differences | | | |
|---|---|---|---|---|---|
| | | | | | 95% Confidence ... |
| | | Mean | Std. Deviation | Std. Error Mean | Lower |
| Pair 23 | COVID-19_Hospital_Deaths_Age_70-79 (%)_1 - COVID-19_Hospital_Deaths_Age_70-79 (%)_3 | -4.72968 | 4.24704 | .37836 | -5.47850 |
| Pair 24 | COVID-19_Hospital_Deaths_Age_70-79 (%)_2 - COVID-19_Hospital_Deaths_Age_70-79 (%)_3 | -1.52480 | 7.44908 | .66627 | -2.84353 |
| Pair 25 | COVID-19_Hospital_Deaths_Age_80-89(%)_1 - COVID-19_Hospital_Deaths_Age_80-89(%)_2 | 1.65698 | 3.26720 | .31734 | 1.02776 |
| Pair 26 | COVID-19_Hospital_Deaths_Age_80-89(%)_1 - COVID-19_Hospital_Deaths_Age_80-89(%)_3 | -2.63262 | 2.79616 | .24910 | -3.12562 |
| Pair 27 | COVID-19_Hospital_Deaths_Age_80-89(%)_2 - COVID-19_Hospital_Deaths_Age_80-89(%)_3 | -2.84528 | 4.03237 | .36067 | -3.55914 |

### Paired Samples Test

| | | Paired ... | t | df | Sig. (2-tailed) |
|---|---|---|---|---|---|
| | | 95% Confidence Interval of the ... | | | |
| | | Upper | | | |
| Pair 23 | COVID-19_Hospital_Deaths_Age_70-79 (%)_1 - COVID-19_Hospital_Deaths_Age_70-79 (%)_3 | -3.98087 | -12.501 | 125 | .000 |
| Pair 24 | COVID-19_Hospital_Deaths_Age_70-79 (%)_2 - COVID-19_Hospital_Deaths_Age_70-79 (%)_3 | -.20607 | -2.289 | 124 | .024 |
| Pair 25 | COVID-19_Hospital_Deaths_Age_80-89(%)_1 - COVID-19_Hospital_Deaths_Age_80-89(%)_2 | 2.28621 | 5.221 | 105 | .000 |
| Pair 26 | COVID-19_Hospital_Deaths_Age_80-89(%)_1 - COVID-19_Hospital_Deaths_Age_80-89(%)_3 | -2.13962 | -10.568 | 125 | .000 |
| Pair 27 | COVID-19_Hospital_Deaths_Age_80-89(%)_2 - COVID-19_Hospital_Deaths_Age_80-89(%)_3 | -2.13142 | -7.889 | 124 | .000 |

### Paired Samples Effect Sizes

| | | | Standardizer[a] | Point Estimate | 95% ... Lower |
|---|---|---|---|---|---|
| Pair 1 | COVID-19_Hospital_Deaths_Age_0-9 (%)_1 - COVID-19_Hospital_Deaths_Age_0-9 (%)_2 | Cohen's d | .16085 | -.139 | -.330 |
| | | Hedges' correction | .16143 | -.139 | -.329 |
| Pair 2 | COVID-19_Hospital_Deaths_Age_0-9 (%)_1 - COVID-19_Hospital_Deaths_Age_0-9 (%)_3 | Cohen's d | 6.61632 | -.131 | -.306 |
| | | Hedges' correction | 6.63625 | -.131 | -.305 |
| Pair 3 | COVID-19_Hospital_Deaths_Age_0-9 (%)_2 - COVID-19_Hospital_Deaths_Age_0-9 (%)_3 | Cohen's d | 6.69754 | -.121 | -.297 |
| | | Hedges' correction | 6.71788 | -.121 | -.296 |
| Pair 4 | COVID-19_Hospital_Deaths_Age_10-19 (%)_1 - COVID-19_Hospital_Deaths_Age_10-19 (%)_2 | Cohen's d | .22463 | -.758 | -.973 |
| | | Hedges' correction | .22543 | -.755 | -.969 |
| Pair 5 | COVID-19_Hospital_Deaths_Age_10-19 (%)_1 - COVID-19_Hospital_Deaths_Age_10-19 (%)_3 | Cohen's d | .09143 | .294 | .115 |
| | | Hedges' correction | .09170 | .293 | .115 |
| Pair 6 | COVID-19_Hospital_Deaths_Age_10-19 (%)_2 - COVID-19_Hospital_Deaths_Age_10-19 (%)_3 | Cohen's d | .20038 | .929 | .718 |
| | | Hedges' correction | .20099 | .927 | .716 |
| Pair 7 | COVID-19_Hospital_Deaths_Age_20-29 (%)_1 - COVID-19_Hospital_Deaths_Age_20-29 (%)_2 | Cohen's d | .51974 | -1.451 | -1.723 |
| | | Hedges' correction | .52160 | -1.446 | -1.717 |
| Pair 8 | COVID-19_Hospital_Deaths_Age_20-29 (%)_1 - COVID-19_Hospital_Deaths_Age_20-29 (%)_3 | Cohen's d | .35939 | .850 | .645 |
| | | Hedges' correction | .36047 | .847 | .643 |
| Pair 9 | COVID-19_Hospital_Deaths_Age_20-29 (%)_2 - COVID-19_Hospital_Deaths_Age_20-29 (%)_3 | Cohen's d | .61557 | 1.763 | 1.481 |
| | | Hedges' correction | .61744 | 1.758 | 1.476 |

### Paired Samples Effect Sizes

| | | | 95% ... |
|---|---|---|---|
| | | | Upper |
| Pair 1 | COVID-19_Hospital_Deaths_Age_0-9 (%)_1 - COVID-19_Hospital_Deaths_Age_0-9 (%)_2 | Cohen's d | .053 |
| | | Hedges' correction | .052 |
| Pair 2 | COVID-19_Hospital_Deaths_Age_0-9 (%)_1 - COVID-19_Hospital_Deaths_Age_0-9 (%)_3 | Cohen's d | .045 |
| | | Hedges' correction | .044 |
| Pair 3 | COVID-19_Hospital_Deaths_Age_0-9 (%)_2 - COVID-19_Hospital_Deaths_Age_0-9 (%)_3 | Cohen's d | .055 |
| | | Hedges' correction | .055 |
| Pair 4 | COVID-19_Hospital_Deaths_Age_10-19 (%)_1 - COVID-19_Hospital_Deaths_Age_10-19 (%)_2 | Cohen's d | -.540 |
| | | Hedges' correction | -.539 |
| Pair 5 | COVID-19_Hospital_Deaths_Age_10-19 (%)_1 - COVID-19_Hospital_Deaths_Age_10-19 (%)_3 | Cohen's d | .472 |
| | | Hedges' correction | .471 |
| Pair 6 | COVID-19_Hospital_Deaths_Age_10-19 (%)_2 - COVID-19_Hospital_Deaths_Age_10-19 (%)_3 | Cohen's d | 1.138 |
| | | Hedges' correction | 1.135 |
| Pair 7 | COVID-19_Hospital_Deaths_Age_20-29 (%)_1 - COVID-19_Hospital_Deaths_Age_20-29 (%)_2 | Cohen's d | -1.176 |
| | | Hedges' correction | -1.172 |
| Pair 8 | COVID-19_Hospital_Deaths_Age_20-29 (%)_1 - COVID-19_Hospital_Deaths_Age_20-29 (%)_3 | Cohen's d | 1.052 |
| | | Hedges' correction | 1.049 |
| Pair 9 | COVID-19_Hospital_Deaths_Age_20-29 (%)_2 - COVID-19_Hospital_Deaths_Age_20-29 (%)_3 | Cohen's d | 2.042 |
| | | Hedges' correction | 2.036 |

### Paired Samples Effect Sizes

| | | | Standardizer[a] | Point Estimate | 95% ... |
|---|---|---|---|---|---|
| | | | | | Lower |
| Pair 10 | COVID-19_Hospital_Deaths_Age_30-39 (%)_1 - COVID-19_Hospital_Deaths_Age_30-39 (%)_2 | Cohen's d | .75519 | -1.038 | -1.272 |
| | | Hedges' correction | .75790 | -1.034 | -1.268 |
| Pair 11 | COVID-19_Hospital_Deaths_Age_30-39 (%)_1 - COVID-19_Hospital_Deaths_Age_30-39 (%)_3 | Cohen's d | 2.00800 | .147 | -.029 |
| | | Hedges' correction | 2.01405 | .146 | -.029 |
| Pair 12 | COVID-19_Hospital_Deaths_Age_30-39 (%)_2 - COVID-19_Hospital_Deaths_Age_30-39 (%)_3 | Cohen's d | 2.02848 | .678 | .482 |
| | | Hedges' correction | 2.03464 | .675 | .480 |
| Pair 13 | COVID-19_Hospital_Deaths_Age_40-49 (%)_1 - COVID-19_Hospital_Deaths_Age_40-49 (%)_2 | Cohen's d | .96578 | .877 | .652 |
| | | Hedges' correction | .96924 | .874 | .649 |
| Pair 14 | COVID-19_Hospital_Deaths_Age_40-49 (%)_1 - COVID-19_Hospital_Deaths_Age_40-49 (%)_3 | Cohen's d | 1.79316 | 1.941 | 1.642 |
| | | Hedges' correction | 1.79856 | 1.935 | 1.637 |
| Pair 15 | COVID-19_Hospital_Deaths_Age_40-49 (%)_2 - COVID-19_Hospital_Deaths_Age_40-49 (%)_3 | Cohen's d | 2.04555 | 1.406 | 1.157 |
| | | Hedges' correction | 2.05176 | 1.402 | 1.153 |
| Pair 16 | COVID-19_Hospital_Deaths_Age_50-59 (%)_1 - COVID-19_Hospital_Deaths_Age_50-59 (%)_2 | Cohen's d | 27.32211 | -1.589 | -1.915 |
| | | Hedges' correction | 27.45102 | -1.582 | -1.906 |
| Pair 17 | COVID-19_Hospital_Deaths_Age_50-59 (%)_1 - COVID-19_Hospital_Deaths_Age_50-59 (%)_3 | Cohen's d | 12.58429 | -.351 | -.531 |
| | | Hedges' correction | 12.62251 | -.350 | -.529 |
| Pair 18 | COVID-19_Hospital_Deaths_Age_50-59 (%)_2 - COVID-19_Hospital_Deaths_Age_50-59 (%)_3 | Cohen's d | 22.61123 | 1.375 | 1.099 |
| | | Hedges' correction | 22.69733 | 1.369 | 1.095 |

### Paired Samples Effect Sizes

| | | | 95% ... |
|---|---|---|---|
| | | | Upper |
| Pair 10 | COVID-19_Hospital_Deaths_Age_30-39 (%)_1 - COVID-19_Hospital_Deaths_Age_30-39 (%)_2 | Cohen's d | -.800 |
| | | Hedges' correction | -.797 |
| Pair 11 | COVID-19_Hospital_Deaths_Age_30-39 (%)_1 - COVID-19_Hospital_Deaths_Age_30-39 (%)_3 | Cohen's d | .322 |
| | | Hedges' correction | .321 |
| Pair 12 | COVID-19_Hospital_Deaths_Age_30-39 (%)_2 - COVID-19_Hospital_Deaths_Age_30-39 (%)_3 | Cohen's d | .871 |
| | | Hedges' correction | .868 |
| Pair 13 | COVID-19_Hospital_Deaths_Age_40-49 (%)_1 - COVID-19_Hospital_Deaths_Age_40-49 (%)_2 | Cohen's d | 1.100 |
| | | Hedges' correction | 1.096 |
| Pair 14 | COVID-19_Hospital_Deaths_Age_40-49 (%)_1 - COVID-19_Hospital_Deaths_Age_40-49 (%)_3 | Cohen's d | 2.236 |
| | | Hedges' correction | 2.229 |
| Pair 15 | COVID-19_Hospital_Deaths_Age_40-49 (%)_2 - COVID-19_Hospital_Deaths_Age_40-49 (%)_3 | Cohen's d | 1.652 |
| | | Hedges' correction | 1.647 |
| Pair 16 | COVID-19_Hospital_Deaths_Age_50-59 (%)_1 - COVID-19_Hospital_Deaths_Age_50-59 (%)_2 | Cohen's d | -1.258 |
| | | Hedges' correction | -1.253 |
| Pair 17 | COVID-19_Hospital_Deaths_Age_50-59 (%)_1 - COVID-19_Hospital_Deaths_Age_50-59 (%)_3 | Cohen's d | -.170 |
| | | Hedges' correction | -.169 |
| Pair 18 | COVID-19_Hospital_Deaths_Age_50-59 (%)_2 - COVID-19_Hospital_Deaths_Age_50-59 (%)_3 | Cohen's d | 1.646 |
| | | Hedges' correction | 1.640 |

### Paired Samples Effect Sizes

| | | | Standardizer[a] | Point Estimate | 95% ... |
|---|---|---|---|---|---|
| | | | | | Lower |
| Pair 19 | COVID-19_Hospital_Deaths_Age_60-69 (%)_1 - COVID-19_Hospital_Deaths_Age_60-69 (%)_2 | Cohen's d | 6.84518 | -.398 | -.595 |
| | | Hedges' correction | 6.86975 | -.397 | -.593 |
| Pair 20 | COVID-19_Hospital_Deaths_Age_60-69 (%)_1 - COVID-19_Hospital_Deaths_Age_60-69 (%)_3 | Cohen's d | 5.51823 | -.301 | -.479 |
| | | Hedges' correction | 5.53486 | -.300 | -.478 |
| Pair 21 | COVID-19_Hospital_Deaths_Age_60-69 (%)_2 - COVID-19_Hospital_Deaths_Age_60-69 (%)_3 | Cohen's d | 9.45731 | .388 | .206 |
| | | Hedges' correction | 9.48603 | .387 | .205 |
| Pair 22 | COVID-19_Hospital_Deaths_Age_70-79 (%)_1 - COVID-19_Hospital_Deaths_Age_70-79 (%)_2 | Cohen's d | 4.28348 | -.241 | -.433 |
| | | Hedges' correction | 4.29885 | -.240 | -.432 |
| Pair 23 | COVID-19_Hospital_Deaths_Age_70-79 (%)_1 - COVID-19_Hospital_Deaths_Age_70-79 (%)_3 | Cohen's d | 4.24704 | -1.114 | -1.335 |
| | | Hedges' correction | 4.25984 | -1.110 | -1.331 |
| Pair 24 | COVID-19_Hospital_Deaths_Age_70-79 (%)_2 - COVID-19_Hospital_Deaths_Age_70-79 (%)_3 | Cohen's d | 7.44908 | -.205 | -.381 |
| | | Hedges' correction | 7.47171 | -.204 | -.380 |
| Pair 25 | COVID-19_Hospital_Deaths_Age_80-89(%)_1 - COVID-19_Hospital_Deaths_Age_80-89(%)_2 | Cohen's d | 3.26720 | .507 | .304 |
| | | Hedges' correction | 3.27893 | .505 | .303 |
| Pair 26 | COVID-19_Hospital_Deaths_Age_80-89(%)_1 - COVID-19_Hospital_Deaths_Age_80-89(%)_3 | Cohen's d | 2.79616 | -.942 | -1.150 |
| | | Hedges' correction | 2.80458 | -.939 | -1.147 |
| Pair 27 | COVID-19_Hospital_Deaths_Age_80-89(%)_2 - COVID-19_Hospital_Deaths_Age_80-89(%)_3 | Cohen's d | 4.03237 | -.706 | -.900 |
| | | Hedges' correction | 4.04462 | -.703 | -.898 |

### Paired Samples Effect Sizes

| | | | 95% ... |
|---|---|---|---|
| | | | Upper |
| Pair 19 | COVID-19_Hospital_Deaths_Age_60-69 (%)_1 - COVID-19_Hospital_Deaths_Age_60-69 (%)_2 | Cohen's d | -.200 |
| | | Hedges' correction | -.199 |
| Pair 20 | COVID-19_Hospital_Deaths_Age_60-69 (%)_1 - COVID-19_Hospital_Deaths_Age_60-69 (%)_3 | Cohen's d | -.122 |
| | | Hedges' correction | -.122 |
| Pair 21 | COVID-19_Hospital_Deaths_Age_60-69 (%)_2 - COVID-19_Hospital_Deaths_Age_60-69 (%)_3 | Cohen's d | .569 |
| | | Hedges' correction | .568 |
| Pair 22 | COVID-19_Hospital_Deaths_Age_70-79 (%)_1 - COVID-19_Hospital_Deaths_Age_70-79 (%)_2 | Cohen's d | -.047 |
| | | Hedges' correction | -.047 |
| Pair 23 | COVID-19_Hospital_Deaths_Age_70-79 (%)_1 - COVID-19_Hospital_Deaths_Age_70-79 (%)_3 | Cohen's d | -.890 |
| | | Hedges' correction | -.887 |
| Pair 24 | COVID-19_Hospital_Deaths_Age_70-79 (%)_2 - COVID-19_Hospital_Deaths_Age_70-79 (%)_3 | Cohen's d | -.027 |
| | | Hedges' correction | -.027 |
| Pair 25 | COVID-19_Hospital_Deaths_Age_80-89(%)_1 - COVID-19_Hospital_Deaths_Age_80-89(%)_2 | Cohen's d | .708 |
| | | Hedges' correction | .706 |
| Pair 26 | COVID-19_Hospital_Deaths_Age_80-89(%)_1 - COVID-19_Hospital_Deaths_Age_80-89(%)_3 | Cohen's d | -.730 |
| | | Hedges' correction | -.728 |
| Pair 27 | COVID-19_Hospital_Deaths_Age_80-89(%)_2 - COVID-19_Hospital_Deaths_Age_80-89(%)_3 | Cohen's d | -.508 |
| | | Hedges' correction | -.507 |

a. The denominator used in estimating the effect sizes.
Cohen's d uses the sample standard deviation of the mean difference.
Hedges' correction uses the sample standard deviation of the mean difference, plus a correction factor.
