## Supplementary material for "The Impact of SARS-CoV-2 Lineages (Variants) on the COVID-19 Epidemic in South Africa": SPSS_Program_(COVID-19 Reported Cases)

```
 GET
  FILE='C:\Users\ThaboMabuka\GOOGLE~1\ARIPRO~1\RESEAR~1\COVID-~1\Papers\ACMRG\THEIMP~1\Da
ta\ADMISS~1\P2B9E7~1.SAV'.
DATASET NAME DataSet1 WINDOW=FRONT.
GET
  FILE='C:\Users\ThaboMabuka\Google Drive\ARI Projects\Research Projects\COVID-19 in Afri
ca\Papers\ACMRG\The Impact of SARS-CoV-2 Variants on the COVID-19 Epidemic in South Afric
a\Data\Reported Cases\P2_Analysis_Dataset_(COVID-19 Reported Cases).sav'.
DATASET NAME DataSet2 WINDOW=FRONT.
DATASET ACTIVATE DataSet1.
DATASET ACTIVATE DataSet1.

SAVE OUTFILE=
    'C:\Users\ThaboMabuka\GOOGLE~1\ARIPRO~1\RESEAR~1\COVID-~1\Papers\ACMRG\THEIMP~1\Data\
ADMISS~1\P2B'+
    '9E7~1.SAV'
  /COMPRESSED.
COMPUTE COVID19_Hospital_To_Active_Cases=COVID19_Hospital_Admitted_Cases/COVID19_Active_C
ases*100.
EXECUTE.
DATASET ACTIVATE DataSet1.

SAVE OUTFILE=
    'C:\Users\ThaboMabuka\GOOGLE~1\ARIPRO~1\RESEAR~1\COVID-~1\Papers\ACMRG\THEIMP~1\Data\
ADMISS~1\P2B'+
    '9E7~1.SAV'
  /COMPRESSED.



 DATASET ACTIVATE DataSet1.

SAVE OUTFILE='C:\Users\ThaboMabuka\Google Drive\ARI Projects\Research Projects\COVID-19 i
n '+
    'Africa\Papers\ACMRG\The Impact of SARS-CoV-2 Variants on the COVID-19 Epidemic in So
uth '+
    'Africa\Data\P2_Analysis_Dataset_(COVID-19 Reported Cases).sav'
  /COMPRESSED.
DESCRIPTIVES VARIABLES=COVID19_Active_Cases COVID19_Daily_Deaths
  /STATISTICS=MEAN STDDEV MIN MAX.
```

#### Descriptives

### Notes

| | | |
|---|---|---|
| Output Created | | 24-SEP-2021 03:06:33 |
| Comments | | |
| Input | Data | C:\Users\ThaboMabuka\Google Drive\ARI Projects\Research Projects\COVID-19 in Africa\Papers\ACMRG\The Impact of SARS-CoV-2 Variants on the COVID-19 Epidemic in South Africa\Data\P2_Analysis_Dataset_(COVID-19 Reported Cases).sav |
| | Active Dataset | DataSet1 |
| | Filter | <none> |
| | Weight | <none> |
| | Split File | COVID-19_Epidemic_Wave |
| | N of Rows in Working Data File | 564 |
| Missing Value Handling | Definition of Missing | User defined missing values are treated as missing. |
| | Cases Used | All non-missing data are used. |
| Syntax | | DESCRIPTIVES VARIABLES=COVID19_Active_Cases COVID19_Daily_Deaths /STATISTICS=MEAN STDDEV MIN MAX. |
| Resources | Processor Time | 00:00:00,00 |
| | Elapsed Time | 00:00:00,01 |

```
[DataSet1] C:\Users\ThaboMabuka\Google Drive\ARI Projects\Research Projects\COVID-19 in A
frica\Papers\ACMRG\The Impact of SARS-CoV-2 Variants on the COVID-19 Epidemic in South Af
rica\Data\P2_Analysis_Dataset_(COVID-19 Reported Cases).sav
```

### Descriptive Statistics

| COVID-19_Epidemic_Wave | | N | Minimum | Maximum | Mean | Std. Deviation |
|---|---|---|---|---|---|---|
| 1 | COVID-19_Active_Cases | 210 | 1 | 173590 | 51746.60 | 54617.959 |
| | COVID-19_Daily_Deaths | 210 | 0 | 572 | 79.69 | 91.878 |
| | Valid N (listwise) | 210 | | | | |
| 2 | COVID-19_Active_Cases | 208 | 19809 | 239799 | 66178.25 | 53878.453 |
| | COVID-19_Daily_Deaths | 208 | 8 | 844 | 180.06 | 179.252 |
| | Valid N (listwise) | 208 | | | | |
| 3 | COVID-19_Active_Cases | 146 | -76869 | 211052 | 112235.60 | 60814.000 |
| | COVID-19_Daily_Deaths | 146 | 0 | 633 | 219.10 | 152.939 |
| | Valid N (listwise) | 146 | | | | |

```
 * Define Variable Properties.
*COVID19_Epidemic_Wave.
VARIABLE LEVEL  COVID19_Epidemic_Wave(NOMINAL).
EXECUTE.
SPSSINC SPLIT DATASET SPLITVAR=COVID19_Epidemic_Wave
/OUTPUT DIRECTORY= DELETECONTENTS=NO
/OPTIONS NAMES=VALUES.
```

```
 SPSSINC SPLIT DATASET SPLITVAR=COVID19_Epidemic_Wave
/OUTPUT DIRECTORY= "C:\Users\ThaboMabuka\Google Drive\ARI Projects\Research Projects\COVI
D-19 in "+
    "Africa\Papers\ACMRG\The Impact of SARS-CoV-2 Variants on the COVID-19 Epidemic in So
uth "+
    "Africa\Data" DELETECONTENTS=NO
/OPTIONS NAMES=VALUES.
```

#### SPSSINC SPLIT DATASET

### Notes

| | | |
|---|---|---|
| Output Created | | 24-SEP-2021 03:23:47 |
| Comments | | |
| Input | Data | C:<br>\Users\ThaboMabuka\Google Drive\ARI Projects\Research Projects\COVID-19 in Africa\Papers\ACMRG\The Impact of SARS-CoV-2 Variants on the COVID-19 Epidemic in South Africa\Data\P2_Analysis_Dataset_(COVID-19 Reported Cases).sav |
| | Active Dataset | DataSet1 |
| | Filter | <none> |
| | Weight | <none> |
| | Split File | <none> |
| Syntax | | BEGIN PROGRAM '#<br>'. |
| Resources | Processor Time | 00:00:00,00 |
| | Elapsed Time | 00:00:00,02 |

```
[DataSet1] C:\Users\ThaboMabuka\Google Drive\ARI Projects\Research Projects\COVID-19 in A
frica\Papers\ACMRG\The Impact of SARS-CoV-2 Variants on the COVID-19 Epidemic in South Af
rica\Data\P2_Analysis_Dataset_(COVID-19 Reported Cases).sav
```

### Split File Information

| | Settings and Statistics |
|---|---|
| Split Variable Names | COVID19_Epidemic_Wave |
| Output Directory | C:\Users\Thabo Mabuka\Google Drive\ARI Projects\Research Projects\COVID-19 in Africa\Papers\ACMRG\The Impact of SARS-CoV-2 Variants on the COVID-19 Epidemic in South Africa\Data |
| Files Deleted | 0 |
| Files Written | 3 |
| File List | None |
| Directories Cleared | No |

**Values and File Names for Split Files Written**

| | Values or Labels | Directory | Data File |
|---|---|---|---|
| 1 | 1 | C: \Users\Thabo Mabuka\Google Drive\ARI Projects\Research Projects\COVID-19 in Africa\Papers\ACMRG\The Impact of SARS-CoV-2 Variants on the COVID-19 Epidemic in South Africa\Data | 1.sav |

**Values and File Names for Split Files Written**

| | Values or Labels | Directory | Data File |
|---|---|---|---|
| 2 | 2 | C: \Users\Thabo Mabuka\Google Drive\ARI Projects\Research Projects\COVID-19 in Africa\Papers\ACMRG\The Impact of SARS-CoV-2 Variants on the COVID-19 Epidemic in South Africa\Data | 2.sav |
| 3 | 3 | C: \Users\Thabo Mabuka\Google Drive\ARI Projects\Research Projects\COVID-19 in Africa\Papers\ACMRG\The Impact of SARS-CoV-2 Variants on the COVID-19 Epidemic in South Africa\Data | 3.sav |

Based on Variables: COVID19_Epidemic_Wave

```
GET
  FILE='C:\Users\ThaboMabuka\Google Drive\ARI Projects\Research Projects\COVID-19 in Afri
ca\Papers\ACMRG\The Impact of SARS-CoV-2 Variants on the COVID-19 Epidemic in South Afric
a\Data\P2_Analysis_Dataset_(COVID-19 Reported Cases)_1.sav'.
DATASET NAME DataSet2 WINDOW=FRONT.
DATASET ACTIVATE DataSet2.

SAVE OUTFILE='C:\Users\ThaboMabuka\Google Drive\ARI Projects\Research Projects\COVID-19 i
```

```
n '+
    'Africa\Papers\ACMRG\The Impact of SARS-CoV-2 Variants on the COVID-19 Epidemic in So
uth '+
    'Africa\Data\P2_Analysis_Dataset_(COVID-19 Reported Cases)_1.sav'
  /COMPRESSED.
DATASET ACTIVATE DataSet1.
DATASET CLOSE DataSet2.
GET
  FILE='C:\Users\ThaboMabuka\Google Drive\ARI Projects\Research Projects\COVID-19 in Afri
ca\Papers\ACMRG\The Impact of SARS-CoV-2 Variants on the COVID-19 Epidemic in South Afric
a\Data\P2_Analysis_Dataset_(COVID-19 Reported Cases)_2.sav'.
DATASET NAME DataSet3 WINDOW=FRONT.
DATASET ACTIVATE DataSet3.

SAVE OUTFILE='C:\Users\ThaboMabuka\Google Drive\ARI Projects\Research Projects\COVID-19 i
n '+
    'Africa\Papers\ACMRG\The Impact of SARS-CoV-2 Variants on the COVID-19 Epidemic in So
uth '+
    'Africa\Data\P2_Analysis_Dataset_(COVID-19 Reported Cases)_2.sav'
  /COMPRESSED.
DATASET ACTIVATE DataSet1.
DATASET CLOSE DataSet3.
GET
  FILE='C:\Users\ThaboMabuka\Google Drive\ARI Projects\Research Projects\COVID-19 in Afri
ca\Papers\ACMRG\The Impact of SARS-CoV-2 Variants on the COVID-19 Epidemic in South Afric
a\Data\P2_Analysis_Dataset_(COVID-19 Reported Cases)_3.sav'.
DATASET NAME DataSet4 WINDOW=FRONT.
DATASET ACTIVATE DataSet4.

SAVE OUTFILE='C:\Users\ThaboMabuka\Google Drive\ARI Projects\Research Projects\COVID-19 i
n '+
    'Africa\Papers\ACMRG\The Impact of SARS-CoV-2 Variants on the COVID-19 Epidemic in So
uth '+
    'Africa\Data\P2_Analysis_Dataset_(COVID-19 Reported Cases)_3.sav'
  /COMPRESSED.
DATASET ACTIVATE DataSet1.
DATASET CLOSE DataSet4.
DATASET ACTIVATE DataSet1.

SAVE OUTFILE='C:\Users\ThaboMabuka\Google Drive\ARI Projects\Research Projects\COVID-19 i
n '+
    'Africa\Papers\ACMRG\The Impact of SARS-CoV-2 Variants on the COVID-19 Epidemic in So
uth '+
    'Africa\Data\P2_Analysis_Dataset_(COVID-19 Reported Cases).sav'
  /COMPRESSED.



 GET
```

```
  FILE='C:\Users\ThaboMabuka\Google Drive\ARI Projects\Research Projects\COVID-19 in Afri
ca\Papers\ACMRG\The Impact of SARS-CoV-2 Variants on the COVID-19 Epidemic in South Afric
a\Data\P2_Analysis_Dataset_(COVID-19 Reported Cases)_1.sav'.
DATASET NAME DataSet1 WINDOW=FRONT.
DATASET ACTIVATE DataSet1.

SAVE OUTFILE='C:\Users\ThaboMabuka\Google Drive\ARI Projects\Research Projects\COVID-19 i
n '+
    'Africa\Papers\ACMRG\The Impact of SARS-CoV-2 Variants on the COVID-19 Epidemic in So
uth '+
    'Africa\Data\P2_Analysis_Dataset_(COVID-19 Reported Cases)_1.sav'
  /COMPRESSED.



 GET
  FILE='C:\Users\ThaboMabuka\Google Drive\ARI Projects\Research Projects\COVID-19 in Afri
ca\Papers\ACMRG\The Impact of SARS-CoV-2 Variants on the COVID-19 Epidemic in South Afric
a\Data\P2_Analysis_Dataset_(COVID-19 Reported Cases)_1.sav'.
DATASET NAME DataSet1 WINDOW=FRONT.
GET
  FILE='C:\Users\ThaboMabuka\Google Drive\ARI Projects\Research Projects\COVID-19 in Afri
ca\Papers\ACMRG\The Impact of SARS-CoV-2 Variants on the COVID-19 Epidemic in South Afric
a\Data\P2_Analysis_Dataset_(COVID-19 Reported Cases)_2.sav'.
DATASET NAME DataSet2 WINDOW=FRONT.
GET
  FILE='C:\Users\ThaboMabuka\Google Drive\ARI Projects\Research Projects\COVID-19 in Afri
ca\Papers\ACMRG\The Impact of SARS-CoV-2 Variants on the COVID-19 Epidemic in South Afric
a\Data\P2_Analysis_Dataset_(COVID-19 Reported Cases)_3.sav'.
DATASET NAME DataSet3 WINDOW=FRONT.
DATASET ACTIVATE DataSet1.
SORT CASES BY Epidemic_Day.
DATASET ACTIVATE DataSet2.
SORT CASES BY Epidemic_Day.
DATASET ACTIVATE DataSet1.
MATCH FILES /FILE=*
  /FILE='DataSet2'
  /BY Epidemic_Day.
EXECUTE.
SORT CASES BY Epidemic_Day.
DATASET ACTIVATE DataSet3.
SORT CASES BY Epidemic_Day.
DATASET ACTIVATE DataSet1.
MATCH FILES /FILE=*
  /FILE='DataSet3'
  /BY Epidemic_Day.
EXECUTE.

SAVE OUTFILE='C:\Users\ThaboMabuka\Google Drive\ARI Projects\Research Projects\COVID-19 i
```

```
n '+
    'Africa\Papers\ACMRG\The Impact of SARS-CoV-2 Variants on the COVID-19 Epidemic in So
uth '+
    'Africa\Data\P2_Analysis_Dataset_(COVID-19 Reported Cases)_M.sav'
  /COMPRESSED.



 DATASET CLOSE DataSet2.
DATASET ACTIVATE DataSet1.
DATASET CLOSE DataSet3.
T-TEST PAIRS=COVID19_Daily_Deaths_1 COVID19_Daily_Deaths_1 COVID19_Daily_Deaths_2
    COVID19_Active_Cases_1 COVID19_Active_Cases_1 COVID19_Active_Cases_2 WITH COVID19_Dai
ly_Deaths_2
    COVID19_Daily_Deaths_3 COVID19_Daily_Deaths_3 COVID19_Active_Cases_2 COVID19_Active_C
ases_3
    COVID19_Active_Cases_3 (PAIRED)
  /ES DISPLAY(TRUE) STANDARDIZER(SD)
  /CRITERIA=CI(.9500)
  /MISSING=ANALYSIS.
```

### T-Test

#### Notes

| | | |
|---|---|---|
| Output Created | | 24-SEP-2021 04:27:25 |
| Comments | | |
| Input | Data | C:\Users\ThaboMabuka\Google Drive\ARI Projects\Research Projects\COVID-19 in Africa\Papers\ACMRG\The Impact of SARS-CoV-2 Variants on the COVID-19 Epidemic in South Africa\Data\P2_Analysis_Dataset_(COVID-19 Reported Cases)_M.sav |
| | Active Dataset | DataSet1 |
| | Filter | <none> |
| | Weight | <none> |
| | Split File | <none> |
| | N of Rows in Working Data File | 210 |

### Notes

| | | |
|---|---|---|
| Missing Value Handling | Definition of Missing | User defined missing values are treated as missing. |
| | Cases Used | Statistics for each analysis are based on the cases with no missing or out-of-range data for any variable in the analysis. |
| Syntax | | T-TEST PAIRS=COVID19_Daily_Deaths_1 COVID19_Daily_Deaths_1 COVID19_Daily_Deaths_2<br><br>COVID19_Active_Cases_1 COVID19_Active_Cases_1 COVID19_Active_Cases_2 WITH COVID19_Daily_Deaths_2<br><br>COVID19_Daily_Deaths_3 COVID19_Daily_Deaths_3 COVID19_Active_Cases_2 COVID19_Active_Cases_3<br><br>COVID19_Active_Cases_3 (PAIRED)<br>/ES DISPLAY(TRUE) STANDARDIZER(SD)<br>/CRITERIA=CI(.9500)<br>/MISSING=ANALYSIS. |
| Resources | Processor Time | 00:00:00,02 |
| | Elapsed Time | 00:00:00,01 |

### Paired Samples Statistics

| | | Mean | N | Std. Deviation |
|---|---|---|---|---|
| Pair 1 | COVID-19_Daily_Deaths | 79.74 | 208 | 92.316 |
| | COVID-19_Daily_Deaths | 180.06 | 208 | 179.252 |
| Pair 2 | COVID-19_Daily_Deaths | 49.71 | 146 | 75.751 |
| | COVID-19_Daily_Deaths | 219.10 | 146 | 152.939 |
| Pair 3 | COVID-19_Daily_Deaths | 223.83 | 146 | 195.874 |
| | COVID-19_Daily_Deaths | 219.10 | 146 | 152.939 |
| Pair 4 | COVID-19_Active_Cases | 51768.79 | 208 | 54880.706 |
| | COVID-19_Active_Cases | 66178.25 | 208 | 53878.453 |
| Pair 5 | COVID-19_Active_Cases | 36407.75 | 146 | 54020.872 |
| | COVID-19_Active_Cases | 112235.60 | 146 | 60814.000 |
| Pair 6 | COVID-19_Active_Cases | 84054.80 | 146 | 55280.275 |
| | COVID-19_Active_Cases | 112235.60 | 146 | 60814.000 |

### Paired Samples Statistics

| | | Std. Error Mean |
|---|---|---|
| Pair 1 | COVID-19_Daily_Deaths | 6.401 |
| | COVID-19_Daily_Deaths | 12.429 |
| Pair 2 | COVID-19_Daily_Deaths | 6.269 |
| | COVID-19_Daily_Deaths | 12.657 |
| Pair 3 | COVID-19_Daily_Deaths | 16.211 |
| | COVID-19_Daily_Deaths | 12.657 |
| Pair 4 | COVID-19_Active_Cases | 3805.292 |
| | COVID-19_Active_Cases | 3735.799 |
| Pair 5 | COVID-19_Active_Cases | 4470.799 |
| | COVID-19_Active_Cases | 5033.002 |
| Pair 6 | COVID-19_Active_Cases | 4575.028 |
| | COVID-19_Active_Cases | 5033.002 |

### Paired Samples Correlations

| | | N | Correlation | Sig. |
|---|---|---|---|---|
| Pair 1 | COVID-19_Daily_Deaths & COVID-19_Daily_Deaths | 208 | .043 | .536 |
| Pair 2 | COVID-19_Daily_Deaths & COVID-19_Daily_Deaths | 146 | .181 | .029 |
| Pair 3 | COVID-19_Daily_Deaths & COVID-19_Daily_Deaths | 146 | .465 | .000 |
| Pair 4 | COVID-19_Active_Cases & COVID-19_Active_Cases | 208 | -.152 | .029 |
| Pair 5 | COVID-19_Active_Cases & COVID-19_Active_Cases | 146 | .188 | .023 |
| Pair 6 | COVID-19_Active_Cases & COVID-19_Active_Cases | 146 | .452 | .000 |

### Paired Samples Test

| | | Paired Differences | | | |
|---|---|---|---|---|---|
| | | | | | 95% Confidence ... |
| | | Mean | Std. Deviation | Std. Error Mean | Lower |
| Pair 1 | COVID-19_Daily_Deaths - COVID-19_Daily_Deaths | -100.317 | 198.058 | 13.733 | -127.392 |
| Pair 2 | COVID-19_Daily_Deaths - COVID-19_Daily_Deaths | -169.390 | 157.890 | 13.067 | -195.217 |
| Pair 3 | COVID-19_Daily_Deaths - COVID-19_Daily_Deaths | 4.733 | 184.056 | 15.233 | -25.374 |
| Pair 4 | COVID-19_Active_Cases - COVID-19_Active_Cases | -14409.457 | 82536.397 | 5722.869 | -25692.039 |
| Pair 5 | COVID-19_Active_Cases - COVID-19_Active_Cases | -75827.842 | 73360.877 | 6071.389 | -87827.698 |
| Pair 6 | COVID-19_Active_Cases - COVID-19_Active_Cases | -28180.795 | 60967.183 | 5045.680 | -38153.377 |

### Paired Samples Test

| | | Paired ... | | | |
|---|---|---|---|---|---|
| | | 95% Confidence Interval of the ... | | | |
| | | Upper | t | df | Sig. (2-tailed) |
| Pair 1 | COVID-19_Daily_Deaths - COVID-19_Daily_Deaths | -73.243 | -7.305 | 207 | .000 |
| Pair 2 | COVID-19_Daily_Deaths - COVID-19_Daily_Deaths | -143.564 | -12.963 | 145 | .000 |
| Pair 3 | COVID-19_Daily_Deaths - COVID-19_Daily_Deaths | 34.839 | .311 | 145 | .756 |
| Pair 4 | COVID-19_Active_Cases - COVID-19_Active_Cases | -3126.875 | -2.518 | 207 | .013 |
| Pair 5 | COVID-19_Active_Cases - COVID-19_Active_Cases | -63827.987 | -12.489 | 145 | .000 |
| Pair 6 | COVID-19_Active_Cases - COVID-19_Active_Cases | -18208.212 | -5.585 | 145 | .000 |

### Paired Samples Effect Sizes

| | | | Standardizer[a] | Point Estimate | 95% ... Lower |
|---|---|---|---|---|---|
| Pair 1 | COVID-19_Daily_Deaths - COVID-19_Daily_Deaths | Cohen's d | 198.058 | -.507 | -.650 |
| | | Hedges' correction | 198.418 | -.506 | -.649 |
| Pair 2 | COVID-19_Daily_Deaths - COVID-19_Daily_Deaths | Cohen's d | 157.890 | -1.073 | -1.275 |
| | | Hedges' correction | 158.300 | -1.070 | -1.272 |
| Pair 3 | COVID-19_Daily_Deaths - COVID-19_Daily_Deaths | Cohen's d | 184.056 | .026 | -.137 |
| | | Hedges' correction | 184.534 | .026 | -.136 |
| Pair 4 | COVID-19_Active_Cases - COVID-19_Active_Cases | Cohen's d | 82536.397 | -.175 | -.311 |
| | | Hedges' correction | 82686.296 | -.174 | -.311 |
| Pair 5 | COVID-19_Active_Cases - COVID-19_Active_Cases | Cohen's d | 73360.877 | -1.034 | -1.234 |
| | | Hedges' correction | 73551.287 | -1.031 | -1.230 |
| Pair 6 | COVID-19_Active_Cases - COVID-19_Active_Cases | Cohen's d | 60967.183 | -.462 | -.632 |
| | | Hedges' correction | 61125.425 | -.461 | -.631 |

### Paired Samples Effect Sizes

| | | | 95% ... Upper |
|---|---|---|---|
| Pair 1 | COVID-19_Daily_Deaths - COVID-19_Daily_Deaths | Cohen's d | -.362 |
| | | Hedges' correction | -.361 |
| Pair 2 | COVID-19_Daily_Deaths - COVID-19_Daily_Deaths | Cohen's d | -.868 |
| | | Hedges' correction | -.866 |
| Pair 3 | COVID-19_Daily_Deaths - COVID-19_Daily_Deaths | Cohen's d | .188 |
| | | Hedges' correction | .187 |
| Pair 4 | COVID-19_Active_Cases - COVID-19_Active_Cases | Cohen's d | -.037 |
| | | Hedges' correction | -.037 |
| Pair 5 | COVID-19_Active_Cases - COVID-19_Active_Cases | Cohen's d | -.831 |
| | | Hedges' correction | -.829 |
| Pair 6 | COVID-19_Active_Cases - COVID-19_Active_Cases | Cohen's d | -.291 |
| | | Hedges' correction | -.290 |

a. The denominator used in estimating the effect sizes.
Cohen's d uses the sample standard deviation of the mean difference.
Hedges' correction uses the sample standard deviation of the mean difference, plus a correction factor.

```
 DATASET ACTIVATE DataSet1.

SAVE OUTFILE='C:\Users\ThaboMabuka\Google Drive\ARI Projects\Research Projects\COVID-19 i
n '+
    'Africa\Papers\ACMRG\The Impact of SARS-CoV-2 Variants on the COVID-19 Epidemic in So
uth '+
    'Africa\Data\Reported Cases\P2_Analysis_Dataset_(COVID-19 Reported Cases).sav'
  /COMPRESSED.
SORT CASES  BY COVID19_Epidemic_Wave.
SPLIT FILE LAYERED BY COVID19_Epidemic_Wave.
DESCRIPTIVES VARIABLES=COVID19_Daily_Tests COVID19_Daily_Positive_Tests
  /STATISTICS=MEAN STDDEV MIN MAX.
```

### Descriptives

#### Notes

| | | |
|---|---|---|
| Output Created | | 29-SEP-2021 00:37:23 |
| Comments | | |
| Input | Data | C:\Users\ThaboMabuka\Google Drive\ARI Projects\Research Projects\COVID-19 in Africa\Papers\ACMRG\The Impact of SARS-CoV-2 Variants on the COVID-19 Epidemic in South Africa\Data\Reported Cases\P2_Analysis_Dataset_(COVID-19 Reported Cases).sav |
| | Active Dataset | DataSet1 |
| | Filter | <none> |
| | Weight | <none> |
| | Split File | COVID-19_Epidemic_Wave |
| | N of Rows in Working Data File | 564 |

**Notes**

| | | |
|---|---|---|
| Missing Value Handling | Definition of Missing | User defined missing values are treated as missing. |
| | Cases Used | All non-missing data are used. |
| Syntax | | DESCRIPTIVES VARIABLES=COVID19_Daily_Tests COVID19_Daily_Positive_Tests /STATISTICS=MEAN STDDEV MIN MAX. |
| Resources | Processor Time | 00:00:00,00 |
| | Elapsed Time | 00:00:00,01 |

```
[DataSet1] C:\Users\ThaboMabuka\Google Drive\ARI Projects\Research Projects\COVID-19 in A
frica\Papers\ACMRG\The Impact of SARS-CoV-2 Variants on the COVID-19 Epidemic in South Af
rica\Data\Reported Cases\P2_Analysis_Dataset_(COVID-19 Reported Cases).sav
```

**Descriptive Statistics**

| COVID-19_Epidemic_Wave | | N | Minimum | Maximum | Mean | Std. Deviation |
|---|---|---|---|---|---|---|
| 1 | COVID-19_Daily_Tests | 200 | 41 | 56663 | 20883.46 | 13937.266 |
| | COVID-19_Daily_Positive_Tests | 200 | .00 | 33.96 | 11.6486 | 8.52239 |
| | Valid N (listwise) | 200 | | | | |
| 2 | COVID-19_Daily_Tests | 201 | 10402 | 77167 | 31046.31 | 14114.793 |
| | COVID-19_Daily_Positive_Tests | 201 | 2.71 | 33.68 | 11.4671 | 8.45221 |
| | Valid N (listwise) | 201 | | | | |
| 3 | COVID-19_Daily_Tests | 138 | 16194 | 132360 | 46811.72 | 17542.503 |
| | COVID-19_Daily_Positive_Tests | 138 | .00 | 63.78 | 17.7520 | 8.91266 |
| | Valid N (listwise) | 138 | | | | |

```
DATASET CLOSE DataSet2.
```

```
T-TEST PAIRS=COVID19_Daily_Tests_1 COVID19_Daily_Tests_1 COVID19_Daily_Tests_2
    COVID19_Daily_Positive_Tests_1 COVID19_Daily_Positive_Tests_1 COVID19_Daily_Positive_
Tests_2 WITH COVID19_Daily_Tests_2
    COVID19_Daily_Tests_3 COVID19_Daily_Tests_3 COVID19_Daily_Positive_Tests_2 COVID19_Da
```

```
ily_Positive_Tests_3
    COVID19_Daily_Positive_Tests_3 (PAIRED)
  /ES DISPLAY(TRUE) STANDARDIZER(SD)
  /CRITERIA=CI(.9500)
  /MISSING=ANALYSIS.
```

### T-Test

#### Notes

| | | |
|---|---|---|
| Output Created | | 29-SEP-2021 01:35:55 |
| Comments | | |
| Input | Data | C:\Users\ThaboMabuka\Google Drive\ARI Projects\Research Projects\COVID-19 in Africa\Papers\ACMRG\The Impact of SARS-CoV-2 Variants on the COVID-19 Epidemic in South Africa\Data\Reported Cases\P2_Analysis_Dataset_(COVID-19 Reported Cases)_M.sav |
| | Active Dataset | DataSet1 |
| | Filter | <none> |
| | Weight | <none> |
| | Split File | <none> |
| | N of Rows in Working Data File | 210 |
| Missing Value Handling | Definition of Missing | User defined missing values are treated as missing. |
| | Cases Used | Statistics for each analysis are based on the cases with no missing or out-of-range data for any variable in the analysis. |

### Notes

| | | |
|---|---|---|
| Syntax | | T-TEST PAIRS=COVID19_Daily_Tests_1 COVID19_Daily_Tests_1 COVID19_Daily_Tests_2<br><br>COVID19_Daily_Positive_Tests_1 COVID19_Daily_Positive_Tests_1 COVID19_Daily_Positive_Tests_2 WITH COVID19_Daily_Tests_2<br><br>COVID19_Daily_Tests_3 COVID19_Daily_Tests_3 COVID19_Daily_Positive_Tests_2 COVID19_Daily_Positive_Tests_3<br><br>COVID19_Daily_Positive_Tests_3 (PAIRED)<br>/ES DISPLAY(TRUE) STANDARDIZER(SD)<br>/CRITERIA=CI(.9500)<br>/MISSING=ANALYSIS. |
| Resources | Processor Time | 00:00:00,02 |
| | Elapsed Time | 00:00:00,01 |

### Paired Samples Statistics

| | | Mean | N | Std. Deviation | Std. Error Mean |
|---|---|---|---|---|---|
| Pair 1 | COVID-19_Daily_Tests_1 | 21238.92 | 191 | 14139.627 | 1023.108 |
| | COVID-19_Daily_Tests_2 | 31614.80 | 191 | 14207.302 | 1028.004 |
| Pair 2 | COVID-19_Daily_Tests_1 | 20013.23 | 128 | 15799.449 | 1396.487 |
| | COVID-19_Daily_Tests_3 | 48569.46 | 128 | 16926.335 | 1496.091 |
| Pair 3 | COVID-19_Daily_Tests_2 | 33403.42 | 131 | 15981.638 | 1396.322 |
| | COVID-19_Daily_Tests_3 | 46033.73 | 131 | 17497.892 | 1528.798 |
| Pair 4 | COVID-19_Daily_Positive_Tests_1 | 12.0065 | 191 | 8.54615 | .61838 |
| | COVID-19_Daily_Positive_Tests_2 | 11.6484 | 191 | 8.63228 | .62461 |
| Pair 5 | COVID-19_Daily_Positive_Tests_1 | 9.8635 | 128 | 9.07552 | .80217 |
| | COVID-19_Daily_Positive_Tests_3 | 18.5391 | 128 | 8.57600 | .75802 |
| Pair 6 | COVID-19_Daily_Positive_Tests_2 | 14.7729 | 131 | 8.48144 | .74103 |
| | COVID-19_Daily_Positive_Tests_3 | 17.3118 | 131 | 8.93236 | .78042 |

#### Paired Samples Correlations

| | | N | Correlation | Sig. |
|---|---|---|---|---|
| Pair 1 | COVID-19_Daily_Tests_1 & COVID-19_Daily_Tests_2 | 191 | .447 | .000 |
| Pair 2 | COVID-19_Daily_Tests_1 & COVID-19_Daily_Tests_3 | 128 | .266 | .002 |
| Pair 3 | COVID-19_Daily_Tests_2 & COVID-19_Daily_Tests_3 | 131 | .244 | .005 |
| Pair 4 | COVID-19_Daily_Positive_Tests_1 & COVID-19_Daily_Positive_Tests_2 | 191 | -.190 | .009 |
| Pair 5 | COVID-19_Daily_Positive_Tests_1 & COVID-19_Daily_Positive_Tests_3 | 128 | -.135 | .130 |
| Pair 6 | COVID-19_Daily_Positive_Tests_2 & COVID-19_Daily_Positive_Tests_3 | 131 | .678 | .000 |

### Paired Samples Test

| | | Paired Differences | | | |
|---|---|---|---|---|---|
| | | | | | 95% Confidence ... |
| | | Mean | Std. Deviation | Std. Error Mean | Lower |
| Pair 1 | COVID-19_Daily_Tests_1 - COVID-19_Daily_Tests_2 | -10375.880 | 14909.088 | 1078.784 | -12503.811 |
| Pair 2 | COVID-19_Daily_Tests_1 - COVID-19_Daily_Tests_3 | -28556.234 | 19847.683 | 1754.304 | -32027.685 |
| Pair 3 | COVID-19_Daily_Tests_2 - COVID-19_Daily_Tests_3 | -12630.305 | 20612.469 | 1800.920 | -16193.210 |
| Pair 4 | COVID-19_Daily_Positive_Tests_1 - COVID-19_Daily_Positive_Tests_2 | .35817 | 13.24970 | .95871 | -1.53292 |
| Pair 5 | COVID-19_Daily_Positive_Tests_1 - COVID-19_Daily_Positive_Tests_3 | -8.67563 | 13.29920 | 1.17549 | -11.00172 |
| Pair 6 | COVID-19_Daily_Positive_Tests_2 - COVID-19_Daily_Positive_Tests_3 | -2.53893 | 6.99456 | .61112 | -3.74795 |

### Paired Samples Test

| | | Paired ... 95% Confidence Interval of the ... Upper | t | df | Sig. (2-tailed) |
|---|---|---|---|---|---|
| Pair 1 | COVID-19_Daily_Tests_1 - COVID-19_Daily_Tests_2 | -8247.948 | -9.618 | 190 | .000 |
| Pair 2 | COVID-19_Daily_Tests_1 - COVID-19_Daily_Tests_3 | -25084.784 | -16.278 | 127 | .000 |
| Pair 3 | COVID-19_Daily_Tests_2 - COVID-19_Daily_Tests_3 | -9067.401 | -7.013 | 130 | .000 |
| Pair 4 | COVID-19_Daily_Positive_Tests_1 - COVID-19_Daily_Positive_Tests_2 | 2.24926 | .374 | 190 | .709 |
| Pair 5 | COVID-19_Daily_Positive_Tests_1 - COVID-19_Daily_Positive_Tests_3 | -6.34953 | -7.380 | 127 | .000 |
| Pair 6 | COVID-19_Daily_Positive_Tests_2 - COVID-19_Daily_Positive_Tests_3 | -1.32991 | -4.155 | 130 | .000 |

### Paired Samples Effect Sizes

| | | | Standardizer[a] | Point Estimate | 95% ... |
|---|---|---|---|---|---|
| | | | | | Lower |
| Pair 1 | COVID-19_Daily_Tests_1 - COVID-19_Daily_Tests_2 | Cohen's d | 14909.088 | -.696 | -.853 |
| | | Hedges' correction | 14938.594 | -.695 | -.852 |
| Pair 2 | COVID-19_Daily_Tests_1 - COVID-19_Daily_Tests_3 | Cohen's d | 19847.683 | -1.439 | -1.685 |
| | | Hedges' correction | 19906.530 | -1.435 | -1.680 |
| Pair 3 | COVID-19_Daily_Tests_2 - COVID-19_Daily_Tests_3 | Cohen's d | 20612.469 | -.613 | -.798 |
| | | Hedges' correction | 20672.168 | -.611 | -.796 |
| Pair 4 | COVID-19_Daily_Positive_Tests_1 - COVID-19_Daily_Positive_Tests_2 | Cohen's d | 13.24970 | .027 | -.115 |
| | | Hedges' correction | 13.27593 | .027 | -.115 |
| Pair 5 | COVID-19_Daily_Positive_Tests_1 - COVID-19_Daily_Positive_Tests_3 | Cohen's d | 13.29920 | -.652 | -.842 |
| | | Hedges' correction | 13.33863 | -.650 | -.840 |
| Pair 6 | COVID-19_Daily_Positive_Tests_2 - COVID-19_Daily_Positive_Tests_3 | Cohen's d | 6.99456 | -.363 | -.539 |
| | | Hedges' correction | 7.01481 | -.362 | -.538 |

#### Paired Samples Effect Sizes

| | | | 95% ... |
|---|---|---|---|
| | | | Upper |
| Pair 1 | COVID-19_Daily_Tests_1 - COVID-19_Daily_Tests_2 | Cohen's d | -.537 |
| | | Hedges' correction | -.536 |
| Pair 2 | COVID-19_Daily_Tests_1 - COVID-19_Daily_Tests_3 | Cohen's d | -1.190 |
| | | Hedges' correction | -1.186 |
| Pair 3 | COVID-19_Daily_Tests_2 - COVID-19_Daily_Tests_3 | Cohen's d | -.425 |
| | | Hedges' correction | -.424 |
| Pair 4 | COVID-19_Daily_Positive_Tests_1 - COVID-19_Daily_Positive_Tests_2 | Cohen's d | .169 |
| | | Hedges' correction | .169 |
| Pair 5 | COVID-19_Daily_Positive_Tests_1 - COVID-19_Daily_Positive_Tests_3 | Cohen's d | -.460 |
| | | Hedges' correction | -.459 |
| Pair 6 | COVID-19_Daily_Positive_Tests_2 - COVID-19_Daily_Positive_Tests_3 | Cohen's d | -.186 |
| | | Hedges' correction | -.185 |

a. The denominator used in estimating the effect sizes.
Cohen's d uses the sample standard deviation of the mean difference.
Hedges' correction uses the sample standard deviation of the mean difference, plus a correction factor.

```
 SORT CASES  BY COVID19_Epidemic_Wave.
SPLIT FILE LAYERED BY COVID19_Epidemic_Wave.
DATASET CLOSE DataSet2.
DESCRIPTIVES VARIABLES=COVID19_Hospital_To_Active_Cases
  /STATISTICS=MEAN STDDEV MIN MAX.
```

#### Descriptives

### Notes

| | | |
|---|---|---|
| Output Created | | 09-OCT-2021 13:10:31 |
| Comments | | |
| Input | Data | C:\Users\ThaboMabuka\GOOGLE~1\ARIPRO~1\RESEAR~1\COVID-~1\Papers\ACMRG\THEIMP~1\Data\ADMISS~1\P2B9E7~1.SAV |
| | Active Dataset | DataSet1 |
| | Filter | <none> |
| | Weight | <none> |
| | Split File | COVID-19_Epidemic_Wave |
| | N of Rows in Working Data File | 484 |
| Missing Value Handling | Definition of Missing | User defined missing values are treated as missing. |
| | Cases Used | All non-missing data are used. |
| Syntax | | DESCRIPTIVES VARIABLES=COVID19_Hospital_To_Active_Cases /STATISTICS=MEAN STDDEV MIN MAX. |
| Resources | Processor Time | 00:00:00,00 |
| | Elapsed Time | 00:00:00,02 |

### Descriptive Statistics

| COVID-19_Epidemic_Wave | | N | Minimum | Maximum | Mean | Std. Deviation |
|---|---|---|---|---|---|---|
| 1 | COVID-19_Hospital_To_Active_Cases (%) | 126 | 4.02 | 12.69 | 6.8042 | 1.82080 |
| | Valid N (listwise) | 126 | | | | |
| 2 | COVID-19_Hospital_To_Active_Cases (%) | 187 | 6.46 | 23.51 | 14.4623 | 4.68271 |
| | Valid N (listwise) | 187 | | | | |
| 3 | COVID-19_Hospital_To_Active_Cases (%) | 145 | 7.18 | 18.33 | 10.6121 | 2.81241 |
| | Valid N (listwise) | 145 | | | | |

```
 DATASET ACTIVATE DataSet1.

SAVE OUTFILE=
    'C:\Users\ThaboMabuka\GOOGLE~1\ARIPRO~1\RESEAR~1\COVID-~1\Papers\ACMRG\THEIMP~1\Data\
ADMISS~1\P2B'+
    '9E7~1.SAV'
  /COMPRESSED.



 GET
  FILE='C:\Users\ThaboMabuka\Google Drive\ARI Projects\Research Projects\COVID-19 in Afri
ca\Papers\ACMRG\The Impact of SARS-CoV-2 Variants on the COVID-19 Epidemic in South Afric
a\Data\Reported Cases\P2_Analysis_Dataset_(COVID-19 Reported Cases).sav'.
DATASET NAME DataSet1 WINDOW=FRONT.
DESCRIPTIVES VARIABLES=COVID19_Active_Cases COVID19_Daily_New_Cases COVID19_Daily_Positiv
e_Tests
  /STATISTICS=MEAN STDDEV MIN MAX.
```

### Descriptives

#### Notes

| | | |
|---|---|---|
| Output Created | | 10-OCT-2021 19:01:06 |
| Comments | | |
| Input | Data | C:\Users\ThaboMabuka\Google Drive\ARI Projects\Research Projects\COVID-19 in Africa\Papers\ACMRG\The Impact of SARS-CoV-2 Variants on the COVID-19 Epidemic in South Africa\Data\Reported Cases\P2_Analysis_Dataset_(COVID-19 Reported Cases).sav |
| | Active Dataset | DataSet1 |
| | Filter | <none> |
| | Weight | <none> |
| | Split File | <none> |
| | N of Rows in Working Data File | 564 |

### Notes

| | | |
|---|---|---|
| Missing Value Handling | Definition of Missing | User defined missing values are treated as missing. |
| | Cases Used | All non-missing data are used. |
| Syntax | | DESCRIPTIVES VARIABLES=COVID19_Active_Cases COVID19_Daily_New_Cases COVID19_Daily_Positive_Tests /STATISTICS=MEAN STDDEV MIN MAX. |
| Resources | Processor Time | 00:00:00,00 |
| | Elapsed Time | 00:00:00,02 |

```
[DataSet1] C:\Users\ThaboMabuka\Google Drive\ARI Projects\Research Projects\COVID-19 in A
frica\Papers\ACMRG\The Impact of SARS-CoV-2 Variants on the COVID-19 Epidemic in South Af
rica\Data\Reported Cases\P2_Analysis_Dataset_(COVID-19 Reported Cases).sav
```

### Descriptive Statistics

| | N | Minimum | Maximum | Mean | Std. Deviation |
|---|---|---|---|---|---|
| COVID-19_Active_Cases | 564 | -76869 | 239799 | 72727.41 | 60926.578 |
| COVID-19_Daily_New_Cases | 564 | 0 | 26485 | 5111.05 | 5394.089 |
| COVID-19_Daily_Positive_Tests | 539 | .00 | 63.78 | 13.1435 | 8.99886 |
| Valid N (listwise) | 539 | | | | |

```
 SORT CASES  BY COVID19_Epidemic_Wave.
SPLIT FILE LAYERED BY COVID19_Epidemic_Wave.
DESCRIPTIVES VARIABLES=COVID19_Active_Cases COVID19_Daily_New_Cases COVID19_Daily_Positiv
e_Tests
  /STATISTICS=MEAN STDDEV MIN MAX.
```

### Descriptives

### Notes

| | | |
|---|---|---|
| Output Created | | 10-OCT-2021 19:02:20 |
| Comments | | |
| Input | Data | C:\Users\ThaboMabuka\Google Drive\ARI Projects\Research Projects\COVID-19 in Africa\Papers\ACMRG\The Impact of SARS-CoV-2 Variants on the COVID-19 Epidemic in South Africa\Data\Reported Cases\P2_Analysis_Dataset_(COVID-19 Reported Cases).sav |
| | Active Dataset | DataSet1 |
| | Filter | <none> |
| | Weight | <none> |
| | Split File | COVID-19_Epidemic_Wave |
| | N of Rows in Working Data File | 564 |
| Missing Value Handling | Definition of Missing | User defined missing values are treated as missing. |
| | Cases Used | All non-missing data are used. |
| Syntax | | DESCRIPTIVES VARIABLES=COVID19_Active_Cases COVID19_Daily_New_Cases COVID19_Daily_Positive_Tests /STATISTICS=MEAN STDDEV MIN MAX. |
| Resources | Processor Time | 00:00:00,00 |
| | Elapsed Time | 00:00:00,00 |

### Descriptive Statistics

| COVID-19_Epidemic_Wave | | N | Minimum | Maximum | Mean |
|---|---|---|---|---|---|
| 1 | COVID-19_Active_Cases | 210 | 1 | 173590 | 51746.60 |
| | COVID-19_Daily_New_Cases | 210 | 0 | 13944 | 3211.14 |
| | COVID-19_Daily_Positive_Tests | 200 | .00 | 33.96 | 11.6486 |
| | Valid N (listwise) | 200 | | | |
| 2 | COVID-19_Active_Cases | 208 | 19809 | 239799 | 66178.25 |
| | COVID-19_Daily_New_Cases | 208 | 437 | 21980 | 4336.45 |
| | COVID-19_Daily_Positive_Tests | 201 | 2.71 | 33.68 | 11.4671 |
| | Valid N (listwise) | 201 | | | |
| 3 | COVID-19_Active_Cases | 146 | -76869 | 211052 | 112235.60 |
| | COVID-19_Daily_New_Cases | 146 | 0 | 26485 | 8947.33 |
| | COVID-19_Daily_Positive_Tests | 138 | .00 | 63.78 | 17.7520 |
| | Valid N (listwise) | 138 | | | |

### Descriptive Statistics

| COVID-19_Epidemic_Wave | | Std. Deviation |
|---|---|---|
| 1 | COVID-19_Active_Cases | 54617.959 |
| | COVID-19_Daily_New_Cases | 3882.002 |
| | COVID-19_Daily_Positive_Tests | 8.52239 |
| | Valid N (listwise) | |
| 2 | COVID-19_Active_Cases | 53878.453 |
| | COVID-19_Daily_New_Cases | 5033.929 |
| | COVID-19_Daily_Positive_Tests | 8.45221 |
| | Valid N (listwise) | |
| 3 | COVID-19_Active_Cases | 60814.000 |
| | COVID-19_Daily_New_Cases | 5845.452 |
| | COVID-19_Daily_Positive_Tests | 8.91266 |
| | Valid N (listwise) | |

```
 * Chart Builder.
GGRAPH
  /GRAPHDATASET NAME="graphdataset" VARIABLES=COVID19_Epidemic_Wave
    MEANCI(COVID19_Daily_Positive_Tests, 95)[name="MEAN_COVID19_Daily_Positive_Tests"
    LOW="MEAN_COVID19_Daily_Positive_Tests_LOW" HIGH="MEAN_COVID19_Daily_Positive_Tests_H
IGH"]
    MISSING=LISTWISE REPORTMISSING=NO
  /GRAPHSPEC SOURCE=INLINE.
BEGIN GPL
  SOURCE: s=userSource(id("graphdataset"))
  DATA: COVID19_Epidemic_Wave=col(source(s), name("COVID19_Epidemic_Wave"), unit.category
())
  DATA: MEAN_COVID19_Daily_Positive_Tests=col(source(s), name("MEAN_COVID19_Daily_Positiv
e_Tests"))
  DATA: LOW=col(source(s), name("MEAN_COVID19_Daily_Positive_Tests_LOW"))
  DATA: HIGH=col(source(s), name("MEAN_COVID19_Daily_Positive_Tests_HIGH"))
  GUIDE: axis(dim(1), label("COVID-19_Epidemic_Wave"))
  GUIDE: axis(dim(2), label("Mean COVID-19_Daily_Positive_Tests"))
  GUIDE: text.title(label("Simple Bar Mean of COVID-19_Daily_Positive_Tests by ",
    "COVID-19_Epidemic_Wave"))
  GUIDE: text.footnote(label("Error Bars: 95% CI"))
  SCALE: linear(dim(2), include(0))
  ELEMENT: interval(position(COVID19_Epidemic_Wave*MEAN_COVID19_Daily_Positive_Tests),
    shape.interior(shape.square))
  ELEMENT: interval(position(region.spread.range(COVID19_Epidemic_Wave*(LOW+HIGH))),
    shape.interior(shape.ibeam))
END GPL.
```

### GGraph

### Notes

| | | |
|---|---|---|
| Output Created | | 10-OCT-2021 19:26:12 |
| Comments | | |
| Input | Data | C:\Users\ThaboMabuka\Google Drive\ARI Projects\Research Projects\COVID-19 in Africa\Papers\ACMRG\The Impact of SARS-CoV-2 Variants on the COVID-19 Epidemic in South Africa\Data\Reported Cases\P2_Analysis_Dataset_(COVID-19 Reported Cases).sav |
| | Active Dataset | DataSet1 |
| | Filter | <none> |
| | Weight | <none> |
| | Split File | COVID-19_Epidemic_Wave |
| | N of Rows in Working Data File | 564 |

### Notes

| | |
|---|---|
| Syntax | GGRAPH<br> /GRAPHDATASET<br>NAME="graphdataset"<br>VARIABLES=COVID19_E<br>pidemic_Wave<br> MEANCI<br>(COVID19_Daily_Positive<br>_Tests, 95)[name="<br>MEAN_COVID19_Daily_P<br>ositive_Tests"<br> LOW="<br>MEAN_COVID19_Daily_P<br>ositive_Tests_LOW"<br>HIGH="<br>MEAN_COVID19_Daily_P<br>ositive_Tests_HIGH"]<br> MISSING=LISTWISE<br>REPORTMISSING=NO<br> /GRAPHSPEC<br>SOURCE=INLINE.<br>BEGIN GPL<br> SOURCE: s=userSource<br>(id("graphdataset"))<br> DATA:<br>COVID19_Epidemic_Wav<br>e=col(source(s), name<br>("COVID19_Epidemic_Wa<br>ve"), unit.category())<br> DATA:<br>MEAN_COVID19_Daily_P<br>ositive_Tests=col(source<br>(s), name<br>("MEAN_COVID19_Daily_<br>Positive_Tests"))<br> DATA: LOW=col(source<br>(s), name<br>("MEAN_COVID19_Daily_<br>Positive_Tests_LOW"))<br> DATA: HIGH=col(source<br>(s), name<br>("MEAN_COVID19_Daily_<br>Positive_Tests_HIGH"))<br> GUIDE: axis(dim(1), label<br>("COVID-<br>19_Epidemic_Wave"))<br> GUIDE: axis(dim(2), label<br>("Mean COVID-<br>19_Daily_Positive_Tests")<br>)<br> GUIDE: text.title(label<br>("Simple Bar Mean of<br>COVID-<br>19_Daily_Positive_Tests<br>by ",<br> "COVID-<br>19_Epidemic_Wave"))<br> GUIDE: text.footnote<br>(label("Error Bars: 95%<br>CI"))<br> SCALE: linear(dim(2),<br>include(0))<br> ELEMENT: interval |

### Notes

| | | |
|---|---|---|
| Resources | Processor Time | 00:00:00,81 |
| | Elapsed Time | 00:00:00,35 |

```
 SORT CASES  BY COVID19_Epidemic_Wave.
SPLIT FILE LAYERED BY COVID19_Epidemic_Wave.
* Chart Builder.
GGRAPH
  /GRAPHDATASET NAME="graphdataset" VARIABLES=COVID19_Epidemic_Wave
    MEANCI(COVID19_Daily_Positive_Tests, 95)[name="MEAN_COVID19_Daily_Positive_Tests"
    LOW="MEAN_COVID19_Daily_Positive_Tests_LOW" HIGH="MEAN_COVID19_Daily_Positive_Tests_H
IGH"]
    MISSING=LISTWISE REPORTMISSING=NO
  /GRAPHSPEC SOURCE=INLINE.
BEGIN GPL
  SOURCE: s=userSource(id("graphdataset"))
  DATA: COVID19_Epidemic_Wave=col(source(s), name("COVID19_Epidemic_Wave"))
  DATA: MEAN_COVID19_Daily_Positive_Tests=col(source(s), name("MEAN_COVID19_Daily_Positiv
e_Tests"))
  DATA: LOW=col(source(s), name("MEAN_COVID19_Daily_Positive_Tests_LOW"))
  DATA: HIGH=col(source(s), name("MEAN_COVID19_Daily_Positive_Tests_HIGH"))
  GUIDE: axis(dim(1), label("COVID-19_Epidemic_Wave"))
  GUIDE: axis(dim(2), label("Mean COVID-19_Daily_Positive_Tests"))
  GUIDE: text.title(label("Simple Bar Mean of COVID-19_Daily_Positive_Tests by ",
    "COVID-19_Epidemic_Wave"))
  GUIDE: text.footnote(label("Error Bars: 95% CI"))
  ELEMENT: interval(position(COVID19_Epidemic_Wave*MEAN_COVID19_Daily_Positive_Tests),
    shape.interior(shape.square))
  ELEMENT: interval(position(region.spread.range(COVID19_Epidemic_Wave*(LOW+HIGH))),
    shape.interior(shape.ibeam))
END GPL.
```

#### GGraph

### Notes

| | | |
|---|---|---|
| Output Created | | 10-OCT-2021 19:28:19 |
| Comments | | |
| Input | Data | C:<br>\Users\ThaboMabuka\Google Drive\ARI Projects\Research Projects\COVID-19 in Africa\Papers\ACMRG\The Impact of SARS-CoV-2 Variants on the COVID-19 Epidemic in South Africa\Data\Reported Cases\P2_Analysis_Dataset_(COVID-19 Reported Cases).sav |
| | Active Dataset | DataSet1 |
| | Filter | <none> |
| | Weight | <none> |
| | Split File | COVID-19_Epidemic_Wave |
| | N of Rows in Working Data File | 564 |

### Notes

| | |
|---|---|
| Syntax | GGRAPH<br> /GRAPHDATASET NAME="graphdataset" VARIABLES=COVID19_Epidemic_Wave<br> MEANCI(COVID19_Daily_Positive_Tests, 95)[name="MEAN_COVID19_Daily_Positive_Tests"<br> LOW="MEAN_COVID19_Daily_Positive_Tests_LOW" HIGH="MEAN_COVID19_Daily_Positive_Tests_HIGH"]<br> MISSING=LISTWISE REPORTMISSING=NO<br> /GRAPHSPEC SOURCE=INLINE.<br>BEGIN GPL<br> SOURCE: s=userSource(id("graphdataset"))<br> DATA: COVID19_Epidemic_Wave=col(source(s), name("COVID19_Epidemic_Wave"))<br> DATA: MEAN_COVID19_Daily_Positive_Tests=col(source(s), name("MEAN_COVID19_Daily_Positive_Tests"))<br> DATA: LOW=col(source(s), name("MEAN_COVID19_Daily_Positive_Tests_LOW"))<br> DATA: HIGH=col(source(s), name("MEAN_COVID19_Daily_Positive_Tests_HIGH"))<br> GUIDE: axis(dim(1), label("COVID-19_Epidemic_Wave"))<br> GUIDE: axis(dim(2), label("Mean COVID-19_Daily_Positive_Tests"))<br> GUIDE: text.title(label("Simple Bar Mean of COVID-19_Daily_Positive_Tests by ",<br> "COVID-19_Epidemic_Wave"))<br> GUIDE: text.footnote(label("Error Bars: 95% CI"))<br> ELEMENT: interval(position(COVID19_Epidemic_Wav |

#### Notes

| | | |
|---|---|---|
| Resources | Processor Time | 00:00:00,27 |
| | Elapsed Time | 00:00:00,16 |

```
 SPLIT FILE OFF.
* Chart Builder.
GGRAPH
  /GRAPHDATASET NAME="graphdataset" VARIABLES=COVID19_Epidemic_Wave
    MEANCI(COVID19_Daily_Positive_Tests, 95)[name="MEAN_COVID19_Daily_Positive_Tests"
    LOW="MEAN_COVID19_Daily_Positive_Tests_LOW" HIGH="MEAN_COVID19_Daily_Positive_Tests_H
IGH"]
    MISSING=LISTWISE REPORTMISSING=NO
  /GRAPHSPEC SOURCE=INLINE.
BEGIN GPL
  SOURCE: s=userSource(id("graphdataset"))
  DATA: COVID19_Epidemic_Wave=col(source(s), name("COVID19_Epidemic_Wave"), unit.category
())
  DATA: MEAN_COVID19_Daily_Positive_Tests=col(source(s), name("MEAN_COVID19_Daily_Positiv
e_Tests"))
  DATA: LOW=col(source(s), name("MEAN_COVID19_Daily_Positive_Tests_LOW"))
  DATA: HIGH=col(source(s), name("MEAN_COVID19_Daily_Positive_Tests_HIGH"))
  GUIDE: axis(dim(1), label("COVID-19_Epidemic_Wave"))
  GUIDE: axis(dim(2), label("Mean COVID-19_Daily_Positive_Tests"))
  GUIDE: text.title(label("Simple Bar Mean of COVID-19_Daily_Positive_Tests by ",
    "COVID-19_Epidemic_Wave"))
  GUIDE: text.footnote(label("Error Bars: 95% CI"))
  SCALE: linear(dim(2), include(0))
  ELEMENT: interval(position(COVID19_Epidemic_Wave*MEAN_COVID19_Daily_Positive_Tests),
    shape.interior(shape.square))
  ELEMENT: interval(position(region.spread.range(COVID19_Epidemic_Wave*(LOW+HIGH))),
    shape.interior(shape.ibeam))
END GPL.
```

#### GGraph

### Notes

| | | |
|---|---|---|
| Output Created | | 10-OCT-2021 19:30:32 |
| Comments | | |
| Input | Data | C: \Users\ThaboMabuka\Google Drive\ARI Projects\Research Projects\COVID-19 in Africa\Papers\ACMRG\The Impact of SARS-CoV-2 Variants on the COVID-19 Epidemic in South Africa\Data\Reported Cases\P2_Analysis_Dataset_(COVID-19 Reported Cases).sav |
| | Active Dataset | DataSet1 |
| | Filter | <none> |
| | Weight | <none> |
| | Split File | <none> |
| | N of Rows in Working Data File | 564 |

### Notes

| | |
|---|---|
| Syntax | GGRAPH<br>/GRAPHDATASET<br>NAME="graphdataset"<br>VARIABLES=COVID19_E<br>pidemic_Wave<br>MEANCI<br>(COVID19_Daily_Positive<br>_Tests, 95)[name="<br>MEAN_COVID19_Daily_P<br>ositive_Tests"<br>LOW="<br>MEAN_COVID19_Daily_P<br>ositive_Tests_LOW"<br>HIGH="<br>MEAN_COVID19_Daily_P<br>ositive_Tests_HIGH"]<br>MISSING=LISTWISE<br>REPORTMISSING=NO<br>/GRAPHSPEC<br>SOURCE=INLINE.<br>BEGIN GPL<br>SOURCE: s=userSource<br>(id("graphdataset"))<br>DATA:<br>COVID19_Epidemic_Wav<br>e=col(source(s), name<br>("COVID19_Epidemic_Wa<br>ve"), unit.category())<br>DATA:<br>MEAN_COVID19_Daily_P<br>ositive_Tests=col(source<br>(s), name<br>("MEAN_COVID19_Daily_<br>Positive_Tests"))<br>DATA: LOW=col(source<br>(s), name<br>("MEAN_COVID19_Daily_<br>Positive_Tests_LOW"))<br>DATA: HIGH=col(source<br>(s), name<br>("MEAN_COVID19_Daily_<br>Positive_Tests_HIGH"))<br>GUIDE: axis(dim(1), label<br>("COVID-<br>19_Epidemic_Wave"))<br>GUIDE: axis(dim(2), label<br>("Mean COVID-<br>19_Daily_Positive_Tests")<br>)<br>GUIDE: text.title(label<br>("Simple Bar Mean of<br>COVID-<br>19_Daily_Positive_Tests<br>by ",<br>"COVID-<br>19_Epidemic_Wave"))<br>GUIDE: text.footnote<br>(label("Error Bars: 95%<br>CI"))<br>SCALE: linear(dim(2),<br>include(0))<br>ELEMENT: interval |

### Notes

| | | |
|---|---|---|
| Resources | Processor Time | 00:00:00,25 |
| | Elapsed Time | 00:00:00,16 |

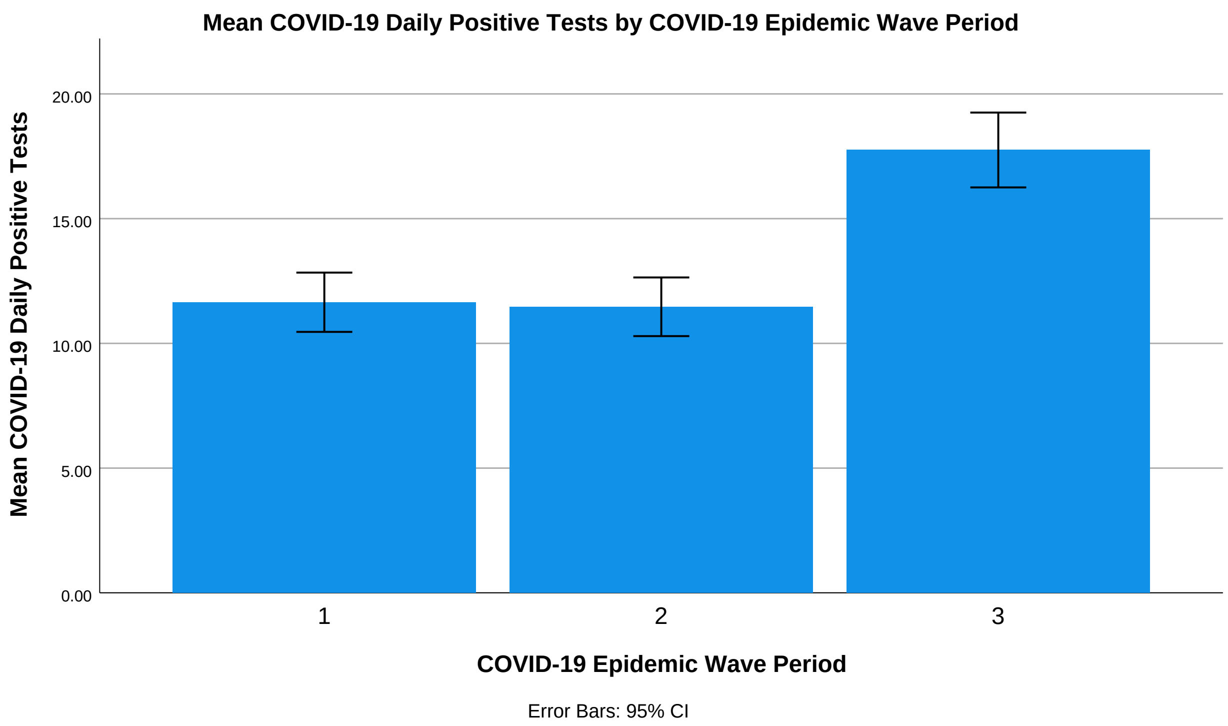


```
 DATASET ACTIVATE DataSet1.

SAVE OUTFILE='C:\Users\ThaboMabuka\Google Drive\ARI Projects\Research Projects\COVID-19 i
n '+
    'Africa\Papers\ACMRG\The Impact of SARS-CoV-2 Variants on the COVID-19 Epidemic in So
uth '+
    'Africa\Data\Reported Cases\P2_Analysis_Dataset_(COVID-19 Reported Cases).sav'
  /COMPRESSED.
```
