## Supplementary material for "The Impact of SARS-CoV-2 Lineages (Variants) on the COVID-19 Epidemic in South Africa": SPSS_Program_(Weekly COVID-19-Excess Deaths)

```
 GET DATA
  /TYPE=XLSX
  /FILE='C:\Users\ThaboMabuka\Google Drive\ARI Projects\Research Projects\COVID-19 in Afr
ica\Papers\ACMRG\The Impact of SARS-CoV-2 Variants on the COVID-19 Epidemic in South Afri
ca\Data\P2_Analysis_Dataset_(Weekly COVID-19-Excess Deaths).xlsx'
  /SHEET=name 'Excess Deaths'
  /CELLRANGE=FULL
  /READNAMES=ON
  /LEADINGSPACES IGNORE=YES
  /TRAILINGSPACES IGNORE=YES
  /DATATYPEMIN PERCENTAGE=95.0
  /HIDDEN IGNORE=YES.
EXECUTE.
DATASET NAME DataSet1 WINDOW=FRONT.

SAVE OUTFILE='C:\Users\ThaboMabuka\Google Drive\ARI Projects\Research Projects\COVID-19 i
n '+
    'Africa\Papers\ACMRG\The Impact of SARS-CoV-2 Variants on the COVID-19 Epidemic in So
uth '+
    'Africa\Data\P2_Analysis_Dataset_(Weekly COVID-19-Excess Deaths).sav'
  /COMPRESSED.
* Define Variable Properties.
*Epidemic_Wave.
VARIABLE LEVEL  Epidemic_Wave(SCALE).
EXECUTE.
SORT CASES  BY Epidemic_Wave.
SPLIT FILE LAYERED BY Epidemic_Wave.
DESCRIPTIVES VARIABLES=Deaths_Natural Excess_Deaths_Natural Weekly_Reported_COVID19_Death
s
  /STATISTICS=MEAN STDDEV MIN MAX.
```

### Descriptives

### Notes

| | | |
|---|---|---|
| Output Created | | 24-SEP-2021 16:15:30 |
| Comments | | |
| Input | Data | C:\Users\ThaboMabuka\Google Drive\ARI Projects\Research Projects\COVID-19 in Africa\Papers\ACMRG\The Impact of SARS-CoV-2 Variants on the COVID-19 Epidemic in South Africa\Data\P2_Analysis_Dataset_(Weekly COVID-19-Excess Deaths).sav |
| | Active Dataset | DataSet1 |
| | Filter | <none> |
| | Weight | <none> |
| | Split File | Epidemic_Wave |
| | N of Rows in Working Data File | 78 |
| Missing Value Handling | Definition of Missing | User defined missing values are treated as missing. |
| | Cases Used | All non-missing data are used. |
| Syntax | | DESCRIPTIVES VARIABLES=Deaths_Natural Excess_Deaths_Natural Weekly_Reported_COVID19_Deaths /STATISTICS=MEAN STDDEV MIN MAX. |
| Resources | Processor Time | 00:00:00,00 |
| | Elapsed Time | 00:00:00,00 |

```
[DataSet1] C:\Users\ThaboMabuka\Google Drive\ARI Projects\Research Projects\COVID-19 in Africa\Papers\ACMRG\The Impact of SARS-CoV-2 Variants on the COVID-19 Epidemic in South Africa\Data\P2_Analysis_Dataset_(Weekly COVID-19-Excess Deaths).sav
```

### Descriptive Statistics

| Epidemic_Wave | | N | Minimum | Maximum | Mean |
|---|---|---|---|---|---|
| 1 | Deaths_Natural | 30 | 7932.000000 | 15864.00000 | 10397.76667 |
| | Excess_Deaths_(Natural) | 29 | -871.000000 | 6673.000000 | 1619.034483 |
| | Weekly_Reported_COVID-19_Deaths | 30 | 1 | 2057 | 613.60 |
| | Valid N (listwise) | 29 | | | |
| 2 | Deaths_Natural | 30 | 9027.000000 | 24198.00000 | 12005.50000 |
| | Excess_Deaths_(Natural) | 30 | 831.0000000 | 16106.00000 | 3864.200000 |
| | Weekly_Reported_COVID-19_Deaths | 30 | 281 | 4027 | 1225.83 |
| | Valid N (listwise) | 30 | | | |
| 3 | Deaths_Natural | 18 | 10690.00000 | 19873.00000 | 14505.96183 |
| | Excess_Deaths_(Natural) | 18 | 1899.000000 | 10253.00000 | 5271.315632 |
| | Weekly_Reported_COVID-19_Deaths | 18 | 566 | 2812 | 1718.50 |
| | Valid N (listwise) | 18 | | | |

### Descriptive Statistics

| Epidemic_Wave | | Std. Deviation |
|---|---|---|
| 1 | Deaths_Natural | 2420.266666 |
| | Excess_Deaths_(Natural) | 2110.342888 |
| | Weekly_Reported_COVID-19_Deaths | 556.488 |
| | Valid N (listwise) | |
| 2 | Deaths_Natural | 4300.891630 |
| | Excess_Deaths_(Natural) | 4242.700258 |
| | Weekly_Reported_COVID-19_Deaths | 1153.173 |
| | Valid N (listwise) | |
| 3 | Deaths_Natural | 2826.034903 |
| | Excess_Deaths_(Natural) | 2635.824249 |
| | Weekly_Reported_COVID-19_Deaths | 807.793 |
| | Valid N (listwise) | |

```
DESCRIPTIVES VARIABLES=Unreported_Excess_Deaths_Natural_to_COVID19_Death_Ratio
  /STATISTICS=MEAN STDDEV MIN MAX.
```

### Descriptives

#### Notes

| | | |
|---|---|---|
| Output Created | | 24-SEP-2021 16:16:25 |
| Comments | | |
| Input | Data | C:\Users\ThaboMabuka\Google Drive\ARI Projects\Research Projects\COVID-19 in Africa\Papers\ACMRG\The Impact of SARS-CoV-2 Variants on the COVID-19 Epidemic in South Africa\Data\P2_Analysis_Dataset_(Weekly COVID-19-Excess Deaths).sav |
| | Active Dataset | DataSet1 |
| | Filter | <none> |
| | Weight | <none> |
| | Split File | Epidemic_Wave |
| | N of Rows in Working Data File | 78 |
| Missing Value Handling | Definition of Missing | User defined missing values are treated as missing. |
| | Cases Used | All non-missing data are used. |
| Syntax | | DESCRIPTIVES VARIABLES=Unreported_Excess_Deaths_Natural_to_COVID19_Death_Ratio /STATISTICS=MEAN STDDEV MIN MAX. |
| Resources | Processor Time | 00:00:00,00 |
| | Elapsed Time | 00:00:00,00 |

### Descriptive Statistics

| Epidemic_Wave | | N | Minimum | Maximum | Mean |
|---|---|---|---|---|---|
| 1 | Unreported_Excess_Deaths_(Natural)_to_COVID-19_Death_Ratio | 30 | -.488635161 | 5.692765648 | 1.057503441 |
| | Valid N (listwise) | 30 | | | |
| 2 | Unreported_Excess_Deaths_(Natural)_to_COVID-19_Death_Ratio | 30 | .4101190641 | 4.672628546 | 2.173347039 |
| | Valid N (listwise) | 30 | | | |
| 3 | Unreported_Excess_Deaths_(Natural)_to_COVID-19_Death_Ratio | 18 | .9340890381 | 4.644122338 | 2.271512782 |
| | Valid N (listwise) | 18 | | | |

### Descriptive Statistics

| Epidemic_Wave | | Std. Deviation |
|---|---|---|
| 1 | Unreported_Excess_Deaths_(Natural)_to_COVID-19_Death_Ratio | 1.684895223 |
| | Valid N (listwise) | |
| 2 | Unreported_Excess_Deaths_(Natural)_to_COVID-19_Death_Ratio | 1.065533109 |
| | Valid N (listwise) | |
| 3 | Unreported_Excess_Deaths_(Natural)_to_COVID-19_Death_Ratio | .9935464046 |
| | Valid N (listwise) | |

```
 DATASET ACTIVATE DataSet1.

SAVE OUTFILE='C:\Users\ThaboMabuka\Google Drive\ARI Projects\Research Projects\COVID-19 i
n '+
    'Africa\Papers\ACMRG\The Impact of SARS-CoV-2 Variants on the COVID-19 Epidemic in So
uth '+
    'Africa\Data\P2_Analysis_Dataset_(Weekly COVID-19-Excess Deaths).sav'
  /COMPRESSED.
* Define Variable Properties.
*Epidemic_Wave.
VARIABLE LEVEL  Epidemic_Wave(NOMINAL).
EXECUTE.
DATASET ACTIVATE DataSet1.
```

```
SAVE OUTFILE='C:\Users\ThaboMabuka\Google Drive\ARI Projects\Research Projects\COVID-19 i
n '+
    'Africa\Papers\ACMRG\The Impact of SARS-CoV-2 Variants on the COVID-19 Epidemic in So
uth '+
    'Africa\Data\P2_Analysis_Dataset_(Weekly COVID-19-Excess Deaths)_2.sav'
  /COMPRESSED.
DATASET ACTIVATE DataSet1.
DATASET CLOSE DataSet2.
DATASET ACTIVATE DataSet3.
DATASET ACTIVATE DataSet3.

SAVE OUTFILE='C:\Users\ThaboMabuka\Google Drive\ARI Projects\Research Projects\COVID-19 i
n '+
    'Africa\Papers\ACMRG\The Impact of SARS-CoV-2 Variants on the COVID-19 Epidemic in So
uth '+
    'Africa\Data\P2_Analysis_Dataset_(Weekly COVID-19-Excess Deaths)_3.sav'
  /COMPRESSED.
DATASET ACTIVATE DataSet1.
DATASET CLOSE DataSet3.
GET
  FILE='C:\Users\ThaboMabuka\Google Drive\ARI Projects\Research Projects\COVID-19 in Afri
ca\Papers\ACMRG\The Impact of SARS-CoV-2 Variants on the COVID-19 Epidemic in South Afric
a\Data\P2_Analysis_Dataset_(Weekly COVID-19-Excess Deaths)_1.sav'.
DATASET NAME DataSet4 WINDOW=FRONT.
DATASET ACTIVATE DataSet4.

SAVE OUTFILE='C:\Users\ThaboMabuka\Google Drive\ARI Projects\Research Projects\COVID-19 i
n '+
    'Africa\Papers\ACMRG\The Impact of SARS-CoV-2 Variants on the COVID-19 Epidemic in So
uth '+
    'Africa\Data\P2_Analysis_Dataset_(Weekly COVID-19-Excess Deaths)_1.sav'
  /COMPRESSED.
DATASET ACTIVATE DataSet1.
DATASET CLOSE DataSet4.
T-TEST PAIRS=Excess_Deaths_Natural_1 Excess_Deaths_Natural_1 Excess_Deaths_Natural_2
    Weekly_Reported_COVID19_Deaths_1 Weekly_Reported_COVID19_Deaths_1 Weekly_Reported_COV
ID19_Deaths_2
    Excess_Natural_to_Natural_Deaths_1 Excess_Natural_to_Natural_Deaths_1
    Excess_Natural_to_Natural_Deaths_2 WITH Excess_Deaths_Natural_2 Excess_Deaths_Natural
_3
    Excess_Deaths_Natural_3 Weekly_Reported_COVID19_Deaths_2 Weekly_Reported_COVID19_Deat
hs_3
    Weekly_Reported_COVID19_Deaths_3 Excess_Natural_to_Natural_Deaths_2
    Excess_Natural_to_Natural_Deaths_3 Excess_Natural_to_Natural_Deaths_3 (PAIRED)
  /ES DISPLAY(TRUE) STANDARDIZER(SD)
  /CRITERIA=CI(.9500)
  /MISSING=ANALYSIS.
```

### T-Test

### Notes

| | | |
|---|---|---|
| Output Created | | 24-SEP-2021 16:51:44 |
| Comments | | |
| Input | Data | C:\Users\ThaboMabuka\Google Drive\ARI Projects\Research Projects\COVID-19 in Africa\Papers\ACMRG\The Impact of SARS-CoV-2 Variants on the COVID-19 Epidemic in South Africa\Data\P2_Analysis_Dataset_(Weekly COVID-19-Excess Deaths)_M.sav |
| | Active Dataset | DataSet1 |
| | Filter | <none> |
| | Weight | <none> |
| | Split File | <none> |
| | N of Rows in Working Data File | 30 |
| Missing Value Handling | Definition of Missing | User defined missing values are treated as missing. |
| | Cases Used | Statistics for each analysis are based on the cases with no missing or out-of-range data for any variable in the analysis. |

### Notes

| | | |
|---|---|---|
| Syntax | | T-TEST PAIRS=Excess_Deaths_Natural_1 Excess_Deaths_Natural_1 Excess_Deaths_Natural_2<br><br>Weekly_Reported_COVID19_Deaths_1 Weekly_Reported_COVID19_Deaths_1 Weekly_Reported_COVID19_Deaths_2<br><br>Excess_Natural_to_Natural_Deaths_1 Excess_Natural_to_Natural_Deaths_1<br><br>Excess_Natural_to_Natural_Deaths_2 WITH Excess_Deaths_Natural_2 Excess_Deaths_Natural_3<br><br>Excess_Deaths_Natural_3 Weekly_Reported_COVID19_Deaths_2 Weekly_Reported_COVID19_Deaths_3<br><br>Weekly_Reported_COVID19_Deaths_3 Excess_Natural_to_Natural_Deaths_2<br><br>Excess_Natural_to_Natural_Deaths_3 Excess_Natural_to_Natural_Deaths_3 (PAIRED)<br>/ES DISPLAY(TRUE) STANDARDIZER(SD)<br>/CRITERIA=CI(.9500)<br>/MISSING=ANALYSIS. |
| Resources | Processor Time | 00:00:00,02 |
| | Elapsed Time | 00:00:00,01 |

### Paired Samples Statistics

| | | Mean | N | Std. Deviation | Std. Error Mean |
|---|---|---|---|---|---|
| Pair 1 | Excess_Deaths_(Natural)_1 | 1619.034483 | 29 | 2110.342888 | 391.8808363 |
| | Excess_Deaths_(Natural)_2 | 3959.896552 | 29 | 4284.720972 | 795.6527099 |
| Pair 2 | Excess_Deaths_(Natural)_1 | 1525.058824 | 17 | 2439.302602 | 591.6177812 |
| | Excess_Deaths_(Natural)_3 | 5469.687140 | 17 | 2574.723250 | 624.4621126 |
| Pair 3 | Excess_Deaths_(Natural)_2 | 5397.833333 | 18 | 4943.978670 | 1165.306948 |
| | Excess_Deaths_(Natural)_3 | 5271.315632 | 18 | 2635.824249 | 621.2697336 |
| Pair 4 | Weekly_Reported_COVID-19_Deaths_1 | 613.60 | 30 | 556.488 | 101.600 |
| | Weekly_Reported_COVID-19_Deaths_2 | 1225.83 | 30 | 1153.173 | 210.540 |
| Pair 5 | Weekly_Reported_COVID-19_Deaths_1 | 369.72 | 18 | 458.744 | 108.127 |
| | Weekly_Reported_COVID-19_Deaths_3 | 1718.50 | 18 | 807.793 | 190.399 |
| Pair 6 | Weekly_Reported_COVID-19_Deaths_2 | 1696.22 | 18 | 1283.753 | 302.584 |
| | Weekly_Reported_COVID-19_Deaths_3 | 1718.50 | 18 | 807.793 | 190.399 |
| Pair 7 | Excess_(Natural)_to_Natural_Deaths (%)_1 | 11.85792536 | 30 | 14.44699427 | 2.637648216 |
| | Excess_(Natural)_to_Natural_Deaths (%)_2 | 26.42834263 | 30 | 17.20898661 | 3.141916720 |
| Pair 8 | Excess_(Natural)_to_Natural_Deaths (%)_1 | 9.895146998 | 18 | 16.43175932 | 3.873002815 |
| | Excess_(Natural)_to_Natural_Deaths (%)_3 | 34.42624374 | 18 | 10.72102577 | 2.526970008 |
| Pair 9 | Excess_(Natural)_to_Natural_Deaths (%)_2 | 33.29668241 | 18 | 19.42766810 | 4.579145286 |
| | Excess_(Natural)_to_Natural_Deaths (%)_3 | 34.42624374 | 18 | 10.72102577 | 2.526970008 |

#### Paired Samples Correlations

| | | N | Correlation | Sig. |
|---|---|---|---|---|
| Pair 1 | Excess_Deaths_(Natural)_1 & Excess_Deaths_(Natural)_2 | 29 | .061 | .754 |
| Pair 2 | Excess_Deaths_(Natural)_1 & Excess_Deaths_(Natural)_3 | 17 | -.329 | .197 |
| Pair 3 | Excess_Deaths_(Natural)_2 & Excess_Deaths_(Natural)_3 | 18 | .409 | .092 |
| Pair 4 | Weekly_Reported_COVID-19_Deaths_1 & Weekly_Reported_COVID-19_Deaths_2 | 30 | -.031 | .870 |
| Pair 5 | Weekly_Reported_COVID-19_Deaths_1 & Weekly_Reported_COVID-19_Deaths_3 | 18 | .184 | .465 |
| Pair 6 | Weekly_Reported_COVID-19_Deaths_2 & Weekly_Reported_COVID-19_Deaths_3 | 18 | .714 | .001 |
| Pair 7 | Excess_(Natural)_to_Natural_Deaths (%)_1 & Excess_(Natural)_to_Natural_Deaths (%)_2 | 30 | .206 | .275 |
| Pair 8 | Excess_(Natural)_to_Natural_Deaths (%)_1 & Excess_(Natural)_to_Natural_Deaths (%)_3 | 18 | -.104 | .683 |
| Pair 9 | Excess_(Natural)_to_Natural_Deaths (%)_2 & Excess_(Natural)_to_Natural_Deaths (%)_3 | 18 | .610 | .007 |

### Paired Samples Test

| | | Paired Differences | | | |
|---|---|---|---|---|---|
| | | | | | 95% Confidence ... |
| | | Mean | Std. Deviation | Std. Error Mean | Lower |
| Pair 1 | Excess_Deaths_(Natural)_1 - Excess_Deaths_(Natural)_2 | -2340.86207 | 4659.849520 | 865.3123325 | -4113.37403 |
| Pair 2 | Excess_Deaths_(Natural)_1 - Excess_Deaths_(Natural)_3 | -3944.62832 | 4088.097960 | 991.5093940 | -6046.53434 |
| Pair 3 | Excess_Deaths_(Natural)_2 - Excess_Deaths_(Natural)_3 | 126.5177011 | 4552.551207 | 1073.046610 | -2137.41275 |
| Pair 4 | Weekly_Reported_COVID-19_Deaths_1 - Weekly_Reported_COVID-19_Deaths_2 | -612.233 | 1295.909 | 236.600 | -1096.134 |
| Pair 5 | Weekly_Reported_COVID-19_Deaths_1 - Weekly_Reported_COVID-19_Deaths_3 | -1348.778 | 852.488 | 200.933 | -1772.710 |
| Pair 6 | Weekly_Reported_COVID-19_Deaths_2 - Weekly_Reported_COVID-19_Deaths_3 | -22.278 | 905.094 | 213.333 | -472.371 |
| Pair 7 | Excess_(Natural)_to_Natural_Deaths (%)_1 - Excess_(Natural)_to_Natural_Deaths (%)_2 | -14.5704173 | 20.06372136 | 3.663117593 | -22.0623340 |
| Pair 8 | Excess_(Natural)_to_Natural_Deaths (%)_1 - Excess_(Natural)_to_Natural_Deaths (%)_3 | -24.5310967 | 20.52835101 | 4.838578735 | -34.7396055 |
| Pair 9 | Excess_(Natural)_to_Natural_Deaths (%)_2 - Excess_(Natural)_to_Natural_Deaths (%)_3 | -1.12956134 | 15.44187010 | 3.639683688 | -8.80862268 |

### Paired Samples Test

| | | Paired ... 95% Confidence Interval of the ... Upper | t | df | Sig. (2-tailed) |
|---|---|---|---|---|---|
| Pair 1 | Excess_Deaths_(Natural)_1 - Excess_Deaths_(Natural)_2 | -568.350107 | -2.705 | 28 | .011 |
| Pair 2 | Excess_Deaths_(Natural)_1 - Excess_Deaths_(Natural)_3 | -1842.72230 | -3.978 | 16 | .001 |
| Pair 3 | Excess_Deaths_(Natural)_2 - Excess_Deaths_(Natural)_3 | 2390.448155 | .118 | 17 | .908 |
| Pair 4 | Weekly_Reported_COVID-19_Deaths_1 - Weekly_Reported_COVID-19_Deaths_2 | -128.333 | -2.588 | 29 | .015 |
| Pair 5 | Weekly_Reported_COVID-19_Deaths_1 - Weekly_Reported_COVID-19_Deaths_3 | -924.846 | -6.713 | 17 | .000 |
| Pair 6 | Weekly_Reported_COVID-19_Deaths_2 - Weekly_Reported_COVID-19_Deaths_3 | 427.815 | -.104 | 17 | .918 |
| Pair 7 | Excess_(Natural)_to_Natural_Deaths (%)_1 - Excess_(Natural)_to_Natural_Deaths (%)_2 | -7.07850059 | -3.978 | 29 | .000 |
| Pair 8 | Excess_(Natural)_to_Natural_Deaths (%)_1 - Excess_(Natural)_to_Natural_Deaths (%)_3 | -14.3225880 | -5.070 | 17 | .000 |
| Pair 9 | Excess_(Natural)_to_Natural_Deaths (%)_2 - Excess_(Natural)_to_Natural_Deaths (%)_3 | 6.549500008 | -.310 | 17 | .760 |

### Paired Samples Effect Sizes

| | | | Standardizer[a] | Point Estimate | 95% ... Lower |
|---|---|---|---|---|---|
| Pair 1 | Excess_Deaths_(Natural)_1 - Excess_Deaths_(Natural)_2 | Cohen's d | 4659.849520 | -.502 | -.885 |
| | | Hedges' correction | 4723.441252 | -.496 | -.873 |
| Pair 2 | Excess_Deaths_(Natural)_1 - Excess_Deaths_(Natural)_3 | Cohen's d | 4088.097960 | -.965 | -1.534 |
| | | Hedges' correction | 4187.137323 | -.942 | -1.498 |
| Pair 3 | Excess_Deaths_(Natural)_2 - Excess_Deaths_(Natural)_3 | Cohen's d | 4552.551207 | .028 | -.435 |
| | | Hedges' correction | 4656.149698 | .027 | -.425 |
| Pair 4 | Weekly_Reported_COVID-19_Deaths_1 - Weekly_Reported_COVID-19_Deaths_2 | Cohen's d | 1295.909 | -.472 | -.847 |
| | | Hedges' correction | 1312.973 | -.466 | -.836 |
| Pair 5 | Weekly_Reported_COVID-19_Deaths_1 - Weekly_Reported_COVID-19_Deaths_3 | Cohen's d | 852.488 | -1.582 | -2.273 |
| | | Hedges' correction | 871.887 | -1.547 | -2.223 |
| Pair 6 | Weekly_Reported_COVID-19_Deaths_2 - Weekly_Reported_COVID-19_Deaths_3 | Cohen's d | 905.094 | -.025 | -.486 |
| | | Hedges' correction | 925.691 | -.024 | -.475 |
| Pair 7 | Excess_(Natural)_to_Natural_Deaths (%)_1 - Excess_(Natural)_to_Natural_Deaths (%)_2 | Cohen's d | 20.06372136 | -.726 | -1.125 |
| | | Hedges' correction | 20.32791142 | -.717 | -1.110 |
| Pair 8 | Excess_(Natural)_to_Natural_Deaths (%)_1 - Excess_(Natural)_to_Natural_Deaths (%)_3 | Cohen's d | 20.52835101 | -1.195 | -1.795 |
| | | Hedges' correction | 20.99549703 | -1.168 | -1.755 |
| Pair 9 | Excess_(Natural)_to_Natural_Deaths (%)_2 - Excess_(Natural)_to_Natural_Deaths (%)_3 | Cohen's d | 15.44187010 | -.073 | -.535 |
| | | Hedges' correction | 15.79326746 | -.072 | -.523 |

### Paired Samples Effect Sizes

| | | | 95% ... |
|---|---|---|---|
| | | | Upper |
| Pair 1 | Excess_Deaths_(Natural)_1 - Excess_Deaths_(Natural)_2 | Cohen's d | -.112 |
| | | Hedges' correction | -.110 |
| Pair 2 | Excess_Deaths_(Natural)_1 - Excess_Deaths_(Natural)_3 | Cohen's d | -.375 |
| | | Hedges' correction | -.366 |
| Pair 3 | Excess_Deaths_(Natural)_2 - Excess_Deaths_(Natural)_3 | Cohen's d | .489 |
| | | Hedges' correction | .479 |
| Pair 4 | Weekly_Reported_COVID-19_Deaths_1 - Weekly_Reported_COVID-19_Deaths_2 | Cohen's d | -.091 |
| | | Hedges' correction | -.090 |
| Pair 5 | Weekly_Reported_COVID-19_Deaths_1 - Weekly_Reported_COVID-19_Deaths_3 | Cohen's d | -.871 |
| | | Hedges' correction | -.851 |
| Pair 6 | Weekly_Reported_COVID-19_Deaths_2 - Weekly_Reported_COVID-19_Deaths_3 | Cohen's d | .438 |
| | | Hedges' correction | .428 |
| Pair 7 | Excess_(Natural)_to_Natural_Deaths (%)_1 - Excess_(Natural)_to_Natural_Deaths (%)_2 | Cohen's d | -.318 |
| | | Hedges' correction | -.314 |
| Pair 8 | Excess_(Natural)_to_Natural_Deaths (%)_1 - Excess_(Natural)_to_Natural_Deaths (%)_3 | Cohen's d | -.575 |
| | | Hedges' correction | -.562 |
| Pair 9 | Excess_(Natural)_to_Natural_Deaths (%)_2 - Excess_(Natural)_to_Natural_Deaths (%)_3 | Cohen's d | .391 |
| | | Hedges' correction | .382 |

a. The denominator used in estimating the effect sizes.
Cohen's d uses the sample standard deviation of the mean difference.
Hedges' correction uses the sample standard deviation of the mean difference, plus a correction factor.
